# A shared cardiovascular and cerebrovascular risk marker phenotype: physiological context and prognosis

**DOI:** 10.64898/2026.11.20.26366994

**Authors:** Junhua Wu, Qiuxia Chen, Xi Chen, Min Wu, Shuai Mi, Hyo Jung Shin, Hee-Jung Song, Dong Woon Kim, Zhenhua Zhou

## Abstract

Abnormalities in routine cardiovascular and cerebrovascular risk markers often cluster. We characterised their common pattern, the shared adverse physiological axis (SAPA), using systolic and diastolic blood pressure, low- and high-density lipoprotein cholesterol, triglycerides, glycated haemoglobin, body mass index and C-reactive protein. The pattern recurred across clinical cohorts and genetic data. The genetic direction retained an estimated 71.8–91.1% of the linear genetic information these eight markers carried for myocardial infarction and stroke. At similar total fat, participants with higher SAPA had more visceral and less leg fat. Differences in the variability and regional distribution of body fat contributed to weaker marker covariation at older ages, where anatomical associations persisted. The fixed clinical score also distinguished stroke prognosis among Chinese participants initially classified as low risk. Net stroke probabilities over approximately seven years were 2.560% (95% interval, 1.742–3.431%) and 9.351% (5.906–13.037%) in the low and high SAPA groups.

## Introduction

Cardiovascular and cerebrovascular risk assessment draws on blood pressure, lipid levels, glycaemic control and adiposity.^1,2,3^ Common clinical measurements include systolic and diastolic blood pressure (SBP and DBP), low- and high-density lipoprotein cholesterol (LDL and HDL), triglycerides (TG), glycated haemoglobin (HbA1c) and body mass index (BMI). In a prospective study of women, C-reactive protein (CRP) measured with a high-sensitivity assay provided information on long-term cardiovascular risk alongside lipid measurements.^4^ Abnormalities in these eight markers often occur together. Their co-occurrence may reflect aspects of physiology that are difficult to recognise from individual results.

Earlier studies have examined this clustering for several purposes. Metabolic syndrome describes a combination of abnormalities associated with insulin resistance. Continuous scores quantify its severity using waist circumference, blood pressure, TG, HDL and fasting glucose.^5,6^ Risk equations combine measurements to estimate a person’s probability of disease,^7^ and genetic and biomarker studies have identified influences shared across diseases and physiological systems.^8,9,10^ Less is known about which bodily characteristics this recurring pattern reflects and whether it distinguishes prognosis within a conventional risk category. The relationships among markers also vary with age,^11^ raising the possibility that the same pattern has different implications in younger and older adults.

We characterised the shared adverse physiological axis (SAPA). In the clinical reference, higher scores correspond to higher blood pressure, LDL, TG, HbA1c, BMI and CRP together with lower HDL. We scored individuals using fixed weights and compared the pattern across clinical populations and genetic data. We examined how much of the markers’ genetic information for myocardial infarction (MI) and stroke it captured and whether it distinguished prognosis within established risk categories.^12,13^ To give the pattern a physiological interpretation, we related it to fat distribution, changes in waist circumference, fasting insulin and circulating interleukin-1 receptor antagonist (IL-1RA).

## Results

### SAPA recurred across populations and data sources

The clinical and genetic comparisons outlined in Fig. 1a identified a similar eight-marker pattern across sources (Fig. 1b,c, Supplementary Tables S1–S2). Across all three clinical cohorts, blood pressure, LDL, TG, HbA1c, BMI and CRP had positive loadings, and HDL had a negative loading (Fig. 1b). Among adults aged 40–54, the relative contributions closely resembled those estimated from Pan-UK Biobank (Pan-UKB) genetic data. Similarity coefficients ranged from 0.976 to 0.994 in the National Health and Nutrition Examination Survey (NHANES; n = 1,104), China Health and Retirement Longitudinal Study (CHARLS; n = 3,127) and English Longitudinal Study of Ageing (ELSA; n = 413; Fig. 1c).

**Figure 1.**
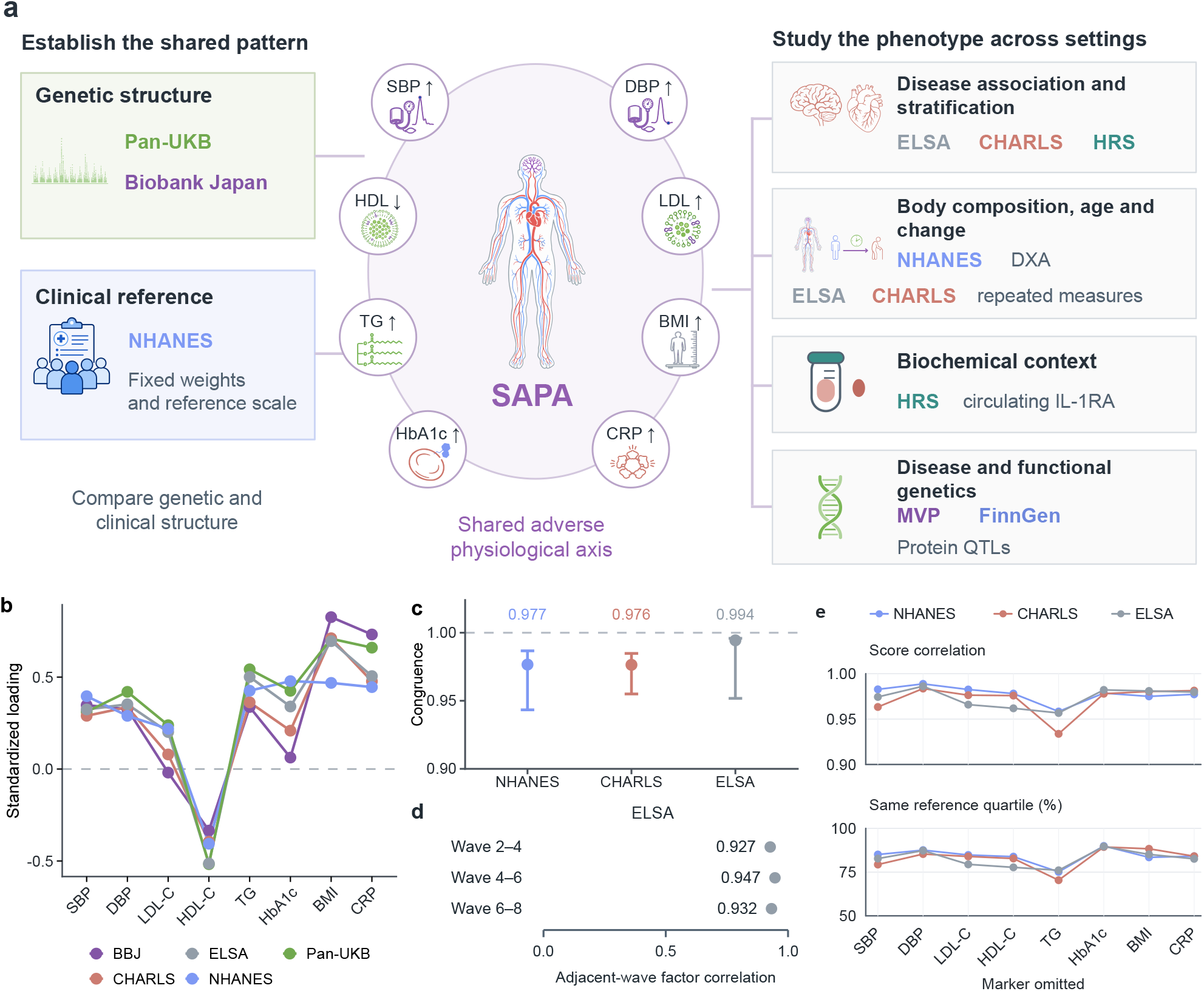
SAPA across clinical and genetic sources **a**, Study design and the shared adverse physiological axis (SAPA). Arrows show the clinical reference directions for blood pressure, lipids, glycated haemoglobin (HbA1c), body mass index (BMI) and C-reactive protein (CRP). Genetic sources inform structural comparisons; NHANES provides fixed clinical scoring weights. Clinical cohorts supply disease, anatomical, longitudinal and biochemical evaluations. **b**, Standardised factor loadings in genetic sources and clinical samples aged 40–54: NHANES (n = 1,104), CHARLS (n = 3,127) and ELSA (n = 413). Colours identify sources; lines join markers. Models include the three specified local relationships. **c**, Tucker congruence with Pan-UKB loadings; points are estimates and bars are 95% intervals from 500 participant resamples per cohort. The dashed line at 1 indicates identical relative loading profiles. **d**, Standardised latent SAPA-factor correlations between adjacent ELSA waves; descriptive estimates without interval estimates. **e**, Weighted Pearson correlations with complete SAPA scores (top) and weighted percentages in the same within-cohort quartile under complete and reduced scoring (bottom) after separately omitting each marker. Remaining terms retain their original weights without rescaling. Points show descriptive estimates without confidence intervals; lines connect omission results within each cohort. NHANES, n = 4,265; CHARLS, n = 8,335; ELSA, n = 4,260. Quartiles use each score version’s weighted within-cohort distribution. Colours identify cohorts as in b; TG and CRP remain log-transformed. Supplementary Table S3A gives all estimates. <u>Source data</u>. NHANES, National Health and Nutrition Examination Survey; CHARLS, China Health and Retirement Longitudinal Study; ELSA, English Longitudinal Study of Ageing; Pan-UKB, Pan-UK Biobank; BBJ, Biobank Japan. SBP/DBP, systolic/diastolic blood pressure; LDL/HDL, low-/high-density lipoprotein cholesterol; TG, triglycerides; IL-1RA, interleukin-1 receptor antagonist; DXA, dual-energy X-ray absorptiometry; GWAS, genome-wide association study. Extended Data Figures 1–2 and Supplementary Tables S1–S2 specify scales, populations, local relationships and scoring coefficients.

Model fit improved when the SBP–DBP, HDL–TG and BMI–CRP pairs could retain additional correlations beyond their shared SAPA component (Extended Data Figures 1a and 2d). Pan-UKB and Biobank Japan genetic profiles also aligned closely (cosine similarity, 0.922), although individual loadings differed (*P* < 0.001 for the joint comparison). The LDL loading in Biobank Japan was close to zero (−0.019, 95% interval, −0.150 to 0.092; Extended Data Fig. 2c). The latent SAPA factor remained strongly correlated across adjacent ELSA waves (0.927–0.947; Fig. 1d).

**Figure 2.**
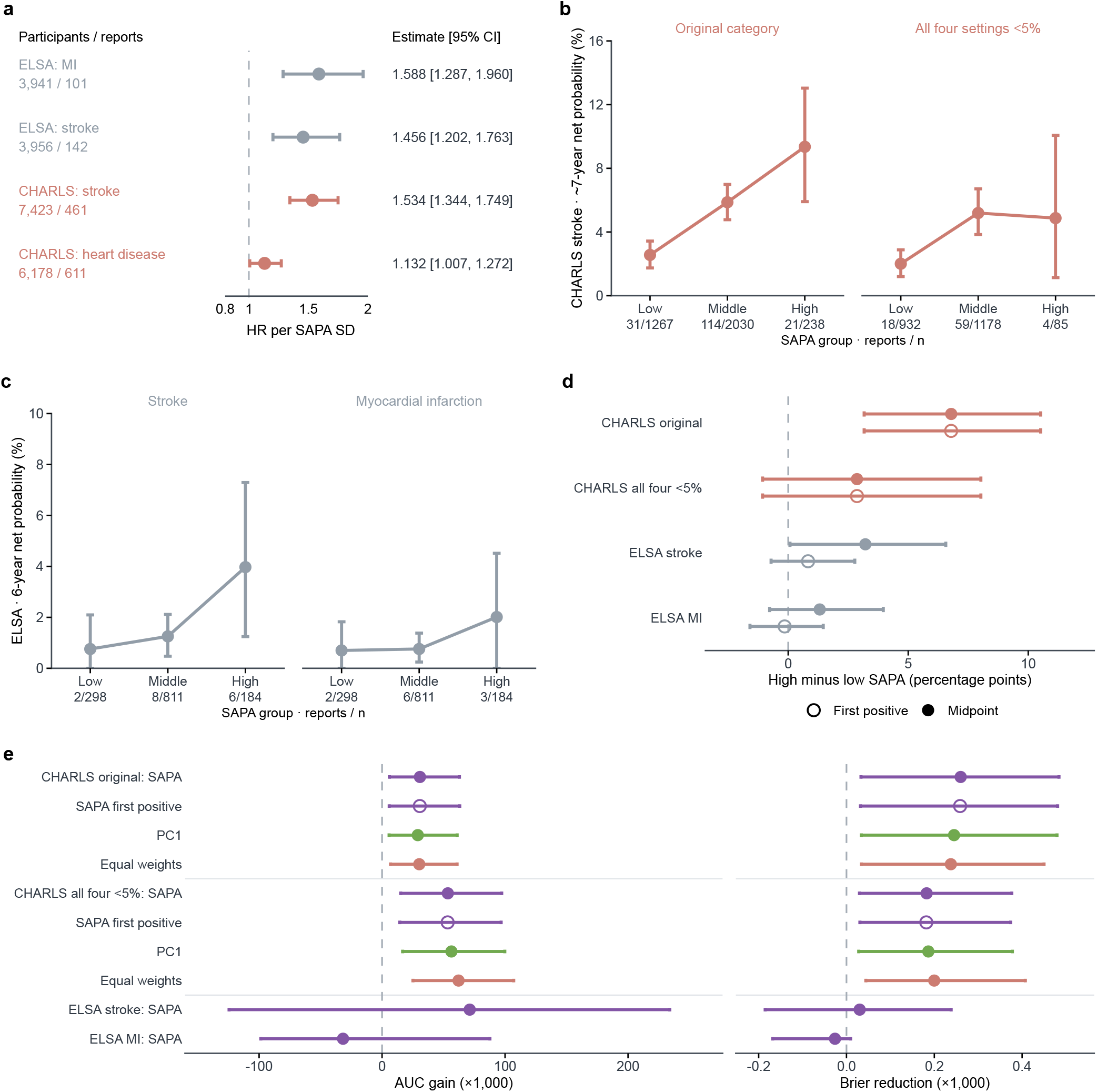
Prognosis after blood measurement **a**, Hazard ratios per analysis-specific SAPA standard deviation (SD) after blood measurement, with 95% confidence intervals and participant/report counts. SDs were 1.427 for ELSA MI, 1.426 for ELSA stroke and 1.499 for each CHARLS endpoint. Colours identify cohorts as in Fig. 1. MI denotes myocardial infarction; CHARLS heart disease is a broad questionnaire endpoint. **b,c**, Net probabilities by fixed NHANES SAPA group, starting at a disease-free, diabetes-negative visit after blood measurement. **b**, Approximately seven-year CHARLS stroke: original China-PAR <5% category (left; n = 3,535; 166 reports) and participants classified consistently under four input settings (right; n = 2,195; 81 reports). **c**, Six-year ELSA stroke/MI (n = 1,293 each; 16/11 midpoint-assigned reports) within lower SCORE2 categories. Bars are pointwise 95% percentile intervals from 1,000 fold-stratified cluster resamples. Net probabilities differ from competing-risk incidence; connecting lines impose no monotonic model. **d**, High-minus-low probability differences for the same populations. Filled/open points use midpoint/first-positive event timing; bars use the same paired resamples. **e**, Held-out area under the receiver operating characteristic curve (AUC) gains and Brier reductions, both multiplied by 1,000, after adding SAPA (purple), the first principal component (PC1; green) or equal weights (salmon). Positive values favour the addition; open points denote first-positive timing. Bars are pointwise 95% percentile intervals conditional on finite, warning-free fits from 200 attempted complete refits. Usable counts were 197/194 for CHARLS original midpoint/first-positive, 199/199 for the consistently classified subset, and 195/192 for ELSA midpoint stroke/MI. ELSA first-positive intervals are omitted because only 77/130 refits qualified. China-PAR, Prediction for Atherosclerotic Cardiovascular Disease Risk in China; SCORE2, Systematic Coronary Risk Evaluation 2. Supplementary Table S5A–E gives definitions, model terms, timing checks and fitting qualification; Supplementary Table S4 gives association models. <u>Source data</u>.

After removing one marker at a time, correlations with complete SAPA scores were 0.934–0.989, and 70.4–89.9% of participants occupied the same quartile when the complete and reduced scores were ranked separately within each cohort (Fig. 1e; Supplementary Table S3A). When one observed test was withheld and estimated from the other seven, the reconstructed SAPA score agreed more closely with the complete-input score than the estimated test agreed with its measured value (Extended Data Figure 3).

### SAPA distinguished prognosis within conventional risk categories

Higher SAPA was associated with subsequent MI and stroke reports (Fig. 2a). In ELSA, hazard ratios per analysis-specific standard deviation (SD) were 1.588 (95% confidence interval (CI), 1.287 to 1.960) for MI and 1.456 (1.202 to 1.763) for stroke. CHARLS estimates were 1.534 (1.344 to 1.749) for stroke and 1.132 (1.007 to 1.272) for broad questionnaire heart disease, including coronary disease, heart failure and other heart problems. The NHANES stroke-history odds ratio was 1.440 (1.091 to 1.899; Supplementary Figure 1a); the Health and Retirement Study (HRS) stroke estimate was less precise (Supplementary Table S4).

We compared prognosis within lower categories of the Prediction for Atherosclerotic Cardiovascular Disease Risk in China (China-PAR) and Systematic Coronary Risk Evaluation 2 (SCORE2) equations.^12,13^ Follow-up began at a disease-free, diabetes-negative visit after blood measurement. Low, middle and high SAPA groups used common NHANES cutoffs (Fig. 2b–e; Supplementary Table S5).

In CHARLS, 3,535 participants originally classified as China-PAR <5% contributed 166 stroke reports over approximately seven years. The high-SAPA group included 238 participants and 21 reports. Net stroke probabilities were 2.560%, 5.871% and 9.351% across low, middle and high SAPA (Fig. 2b). The high-minus-low difference was 6.791 percentage points (95% interval, 3.164 to 10.517, Fig. 2d). Among the 2,195 participants classified as low risk under all four settings for unavailable China-PAR inputs, 81 reported stroke. The corresponding probabilities were 2.007%, 5.192% and 4.872%. With 85 participants and four reports in the high-SAPA group, the high-minus-low difference was imprecise (2.865 percentage points, −1.068 to 8.026).

Continuous SAPA associations persisted in both CHARLS sets, with hazard ratios per raw score unit of 1.440 (95% CI, 1.270 to 1.634) and 1.484 (1.255 to 1.755). In held-out participants, adding SAPA to the risk equation and background model increased area under the receiver operating characteristic curve (AUC) from 0.614 to 0.645 in the original category. In the consistently classified subset, AUC increased from 0.589 to 0.642. Paired gains were 0.031 (0.006 to 0.063) and 0.054 (0.015 to 0.097; Fig. 2e). Prediction error and event-timing comparisons supported similar CHARLS findings (Supplementary Table S5).

In ELSA, 1,293 participants were initially in lower SCORE2 categories. Dating events at the midpoint between interviews placed 16 stroke and 11 MI reports within six years (Fig. 2c). The high-minus-low stroke difference was 3.214 percentage points (95% interval, 0.074 to 6.565) with midpoint dating and 0.827 (−0.714 to 2.773) with dating at the first positive report. Neither timing rule gave a precise MI contrast (Fig. 2d) or clear evidence of improved prediction in ELSA (Supplementary Table S5A–E).

In the original CHARLS category, AUCs were 0.645 for SAPA, 0.643 for the first principal component (PC1) and 0.644 for equal weights. Gains were also similar in the consistently classified subset (Fig. 2e, Supplementary Table S5).

### Fat distribution provided a bodily context for SAPA

We examined 1,876 NHANES adults aged 20–59 with observed dual-energy X-ray absorptiometry (DXA) measurements in 2015–2018. At similar total fat, higher SAPA corresponded to more visceral and less whole-leg fat. Partial correlations were 0.335 (95% CI, 0.267 to 0.399) for visceral adipose tissue (VAT) and −0.371 (−0.430 to −0.309) for whole-leg fat (Fig. 3a). The VAT correlation exceeded that for abdominal subcutaneous adipose tissue (SAT) by 0.269 (0.181 to 0.357). Omitting BMI from SAPA and conditioning on BMI retained a VAT correlation of 0.311 (0.246 to 0.373; Supplementary Table S6).

**Figure 3.**
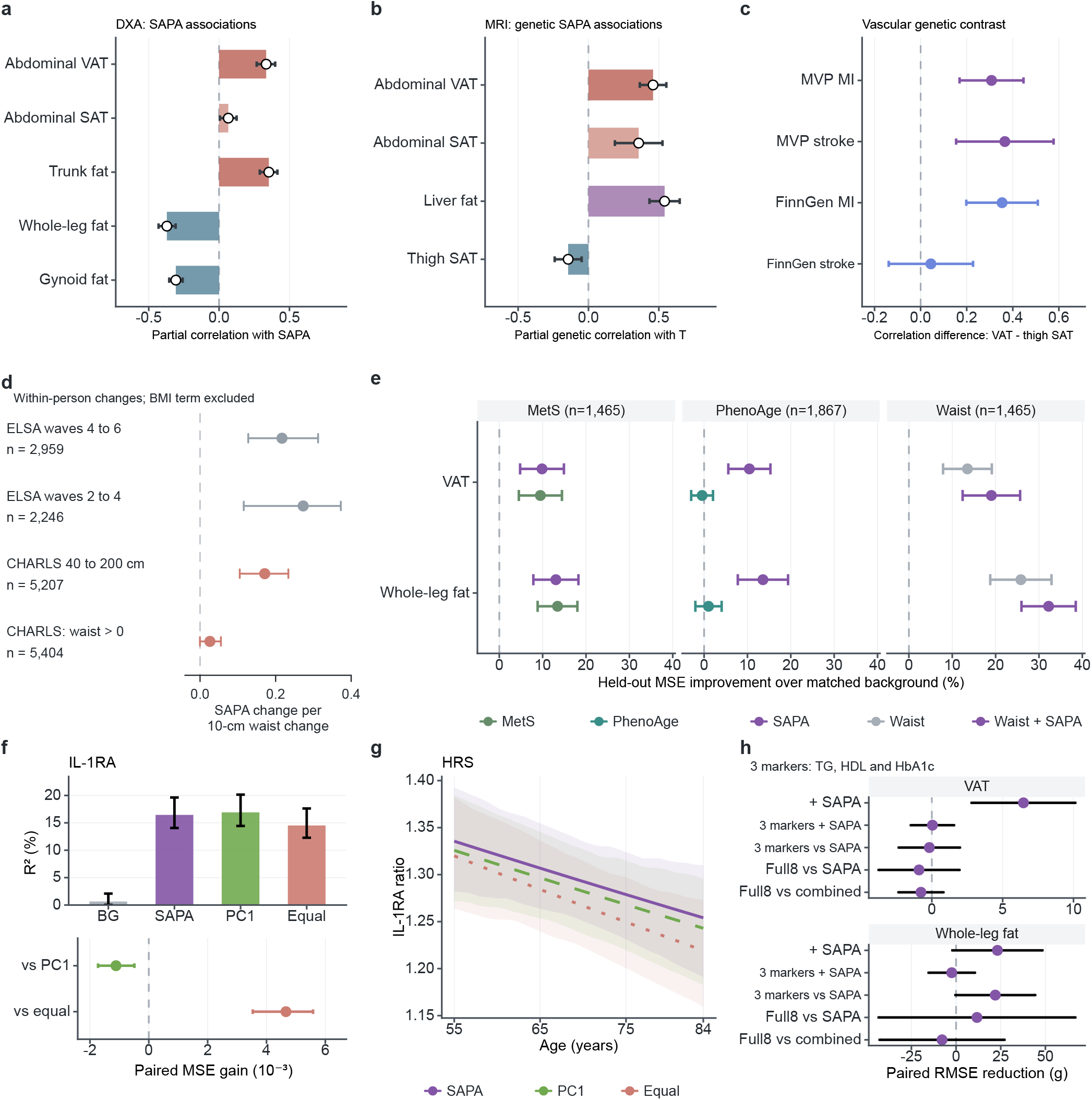
Physiological associations of SAPA across individuals and over time **a**, Partial SAPA correlations with dual-energy X-ray absorptiometry (DXA) fat measures in NHANES 2015–2018 (n = 1,876; ages 20–59), adjusted for total fat and background characteristics; survey-design 95% intervals. **b**, Genetic SAPA projection T correlations with magnetic resonance imaging (MRI) traits, conditional on whole-body fat; delta-method 95% intervals. Sample sizes: abdominal subcutaneous adipose tissue (SAT)/visceral adipose tissue (VAT), 37,589; thigh SAT, 37,584; liver fat, 30,085; whole-body fat, 412,524. Sources overlap; thigh SAT and whole-leg fat are distinct. **c**, Differences between the fat –disease partial genetic correlations: VAT minus thigh SAT for each vascular endpoint. Positive values indicate a stronger correlation for VAT. Bars are joint-error 95% intervals; Holm adjustment covered 20 comparisons. **d**, Associations between each participant’s waist change and SAPA change over the same pair of visits. SAPA excludes its BMI term; models adjust for BMI change. Estimates are score units per 10-cm waist change. Overlapping ELSA samples contain 2,959/2,246 people; CHARLS contains 5,207 with waists 40–200 cm and 5,404 with positive waists. Intervals use 999 household resamples. **e**, Held-out mean squared error (MSE) improvements over matched background: comparisons with continuous metabolic syndrome (MetS; n = 1,465), clinical PhenoAge (n = 1,867), and addition to waist (n = 1,465). Models share participants and background within tasks. **f**, Held-out R² and paired MSE improvements for log interleukin-1 receptor antagonist (IL-1RA; HRS, n = 3,464; 2,680 families). **g**, Geometric-mean ratios per reference SD by age. Intervals/bands use 200 family resamples; performance and age contrasts use separate Holm families. IL-1RA selection was not repeated within folds. **h**, Paired root MSE reductions without imaging inputs (n = 1,869). “3 markers” denotes TG, HDL and HbA1c plus background; combined adds SAPA; Full8 enters all eight markers. Positive values favour added/first-named information. Panels e,h use 60 survey jackknife replicates, reciprocal-cycle retraining and t intervals (30 degrees of freedom). All intervals are pointwise 95%. Models and eligibility: Supplementary Tables S6–S8. BG, background; PC1, first principal component; Equal, equal weights; SD, standard deviation; BMI, body mass index; TG, triglycerides; HDL, high-density lipoprotein cholesterol; HbA1c, glycated haemoglobin; HRS, Health and Retirement Study. <u>Source data</u>.

After adjustment for whole-body fat, genetic SAPA correlations with magnetic resonance imaging (MRI) traits were positive for VAT (0.459, 95% CI, 0.365 to 0.553) and negative for thigh SAT (−0.144, −0.240 to −0.049). MRI thigh SAT and DXA whole-leg fat represent different compartments. Abdominal SAT and liver fat showed positive correlations (Fig. 3b). VAT had a stronger genetic correlation than thigh SAT with MI and survey-reported stroke in the Million Veteran Program (MVP), and with MI in FinnGen. The FinnGen stroke contrast was uncertain (Fig. 3c). Liver fat also shared genetic covariance with all four endpoints along the SAPA direction (Supplementary Table S6).

Changes in waist circumference between two visits tracked changes in SAPA over the same interval within ELSA participants (Fig. 3d). Among 2,959 participants, a 10-cm waist increase accompanied a 0.351-unit increase (95% CI, 0.264 to 0.437) in SAPA with BMI omitted. Accounting for BMI change reduced this to 0.217 (0.128 to 0.312), below the between-person estimate of 0.341 (0.263 to 0.415; difference, −0.124; −0.223 to −0.019). A second, overlapping ELSA window supported the association. CHARLS estimates depended on waist values included: 0.171 (0.105 to 0.234) within 40–200 cm, versus 0.0261 (−0.0004 to 0.0552) with all positive values (Supplementary Figure 2; Supplementary Table S7).

After accounting for total fat and background characteristics, SAPA retained 84.9% (95% CI, 73.8 to 96.0) of the linear VAT information available from all eight markers. SAPA and the Gurka continuous metabolic syndrome severity score gave similar VAT prediction.^14^ Neither score clearly improved prediction beyond the other; clinical PhenoAge gave smaller gains (Fig. 3e; Supplementary Table S6). Adding SAPA to body shape measurements reduced root mean square prediction error by 6.466 g (2.805 to 10.127). With TG, HDL and HbA1c already included, the reduction was 0.041 g (−1.480 to 1.563; Fig. 3h; Extended Data Figure 4).

### SAPA was associated with fasting insulin and IL-1RA

In 2,475 NHANES participants without known diabetes, doubling fasting insulin corresponded to a 0.408-unit higher six-marker SAPA score after excluding BMI and CRP and adjusting for body composition and CRP (95% CI, 0.310 to 0.506). A 10% higher trunk-to-leg fat ratio corresponded to a 0.103-unit higher score (0.085 to 0.121) after accounting for insulin, CRP and the other body measures. The insulin–score association was positive in both cycles. The adjusted insulin–CRP association was uncertain (10.664% higher CRP per insulin doubling, −0.267% to 22.793%). The insulin-associated marker pattern resembled SAPA, with differences in relative contributions (Supplementary Table S8F–J).

Circulating IL-1RA was selected from six additional biomarkers evaluated in HRS (complete screen, Supplementary Table S8A). Among 3,464 participants, predictions evaluated outside the training samples explained 0.643% of IL-1RA variation using background characteristics and 16.452% after adding SAPA (Fig. 3f). PC1 explained 16.914% and equal weights 14.521%. Paired comparisons favoured PC1 over SAPA and SAPA over equal weights (Holm-adjusted *P* = 0.004 and *P* < 0.001). Per reference SD of SAPA, IL-1RA geometric means were 1.321-fold higher at age 60 and 1.265-fold higher at age 80 (95% CIs, 1.275 to 1.371 and 1.209 to 1.314). Both age-specific associations were positive, with no clear age contrast or predictive gain from updating the relationship in older adults (Fig. 3g; Supplementary Figure 3; Supplementary Table S8B–D).

The IL-1RA association extended beyond the BMI and CRP components of SAPA. After accounting for BMI, CRP and background characteristics, SAPA with these two terms omitted improved explained variation by 3.000 percentage points (95% interval, 1.955 to 4.190). Entering the six remaining markers separately added a further 3.408 points (2.054 to 5.588; Supplementary Table S8E).

### Shared disease-related genetic information coexisted with distinct functional profiles

Genetic SAPA (gSAPA) correlated with MI and stroke (Fig. 4a). Correlations were 0.523 (95% CI, 0.476 to 0.569) and 0.396 (0.314 to 0.479) in MVP, and 0.390 (0.341 to 0.438) and 0.372 (0.290 to 0.455) in FinnGen. Mendelian randomisation (MR) estimates were positive for all seven endpoints under both instrument-selection rules (Fig. 4b). The instruments showed heterogeneity, and the MR-Egger intercept suggested directional pleiotropy for MVP cerebral infarction, limiting causal interpretation (Extended Data Figure 5; Supplementary Table S9).

**Figure 4.**
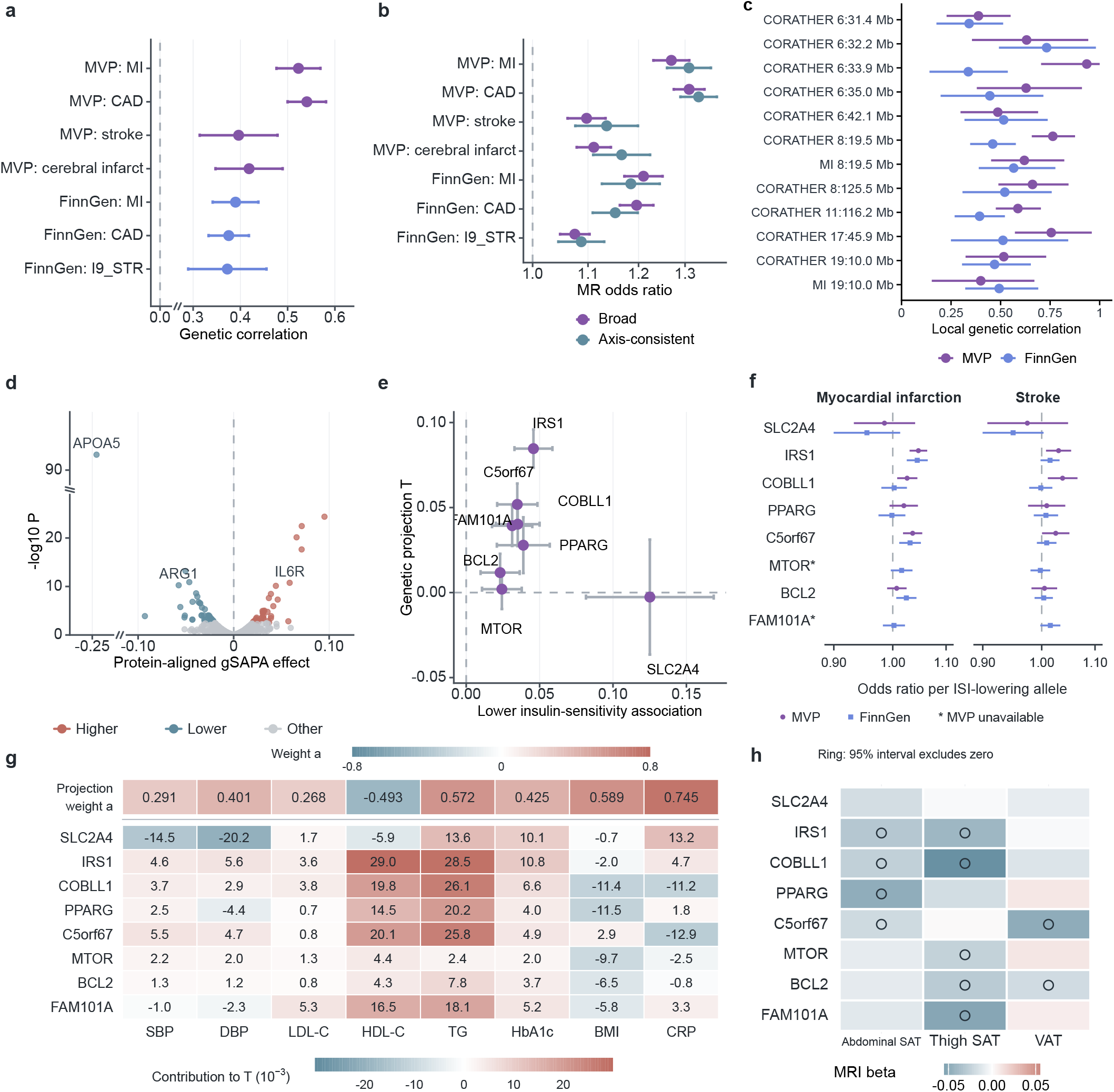
Global, local and functional genetic evidence **a**, Genetic SAPA (gSAPA) correlations with seven vascular endpoints and 95% intervals. MVP stroke is survey-reported; cerebral infarction is separate. FinnGen stroke excludes subarachnoid haemorrhage. **b**, Inverse-variance weighted random-effects Mendelian randomisation odds ratios and 95% intervals per genetic-factor unit. Broad (purple) and axis-consistent (blue-grey) instrument sets were clumped separately. **c**, Twelve local associations across ten genomic regions. Colours identify MVP/FinnGen. Bars are pointwise 95% Local Analysis of [co]Variant Association (LAVA) intervals, assuming zero cross-trait sampling-error correlation and without regional-selection adjustment. CORATHER denotes coronary atherosclerosis. **d**, Associations aligned to protein-increasing alleles at 966 cis protein quantitative trait locus (pQTL) anchors. Higher/Lower denote directions meeting false discovery rate (FDR) < 0.05 across 1,300 eligible assays; Other denotes remaining anchors. Marked breaks omit 0.025–0.28 in a and −0.23 to −0.11 horizontally/28–88 vertically in d. Intervals are retained across breaks. **e**, Eight representatives with BMI-unadjusted insulin sensitivity index (ISI) effects, aligned to ISI-lowering alleles. Bars show marginal reference 95% intervals for ISI and fixed genetic SAPA projection T. **f**, Marginal odds ratios for MI (left) and stroke (right) per ISI-lowering allele in MVP (purple circles) and FinnGen (blue squares). Bars show pointwise 95% intervals on a logarithmic odds-ratio axis. Asterisks mark the two variants with MVP imputation r² < 0.90 for both outcomes. **g**, Contributions for all representatives, in 10⁻³ T units per allele. The separate top row gives fixed genetic projection coefficients a, using its own colour scale. Each contribution in the lower matrix is aⱼβⱼ, where βⱼ is the representative variant’s marginal effect on marker j. Summing the unrounded contributions on the displayed scale and dividing by 1,000 gives T in e. These coefficients differ from the clinical scoring coefficients in Extended Data Fig. 2b. Blue-grey/coral indicate negative/positive values on the two separately labelled scales. **h**, MRI fat associations on original trait scales; rings denote marginal 95% intervals excluding zero. Gene names identify candidate regions. Supplementary Tables S9–S12 specify sources, sample sizes, instruments, selection and error assumptions. MVP, Million Veteran Program; CAD, coronary artery disease; BMI, body mass index; MRI, magnetic resonance imaging; SAT/VAT, subcutaneous/visceral adipose tissue; ASAT, abdominal SAT. <u>Source data</u>.

Regional analyses located 12 associations with MI or coronary atherosclerosis across 10 genomic regions (Fig. 4c). All remained positive when assumptions about correlated estimation errors were varied, although statistical support for individual associations depended on those assumptions (Supplementary Table S10A–D).

Among 1,300 eligible protein assays, 966 had a variant near the protein-coding gene that could be matched to gSAPA. Eighty-one matched protein anchors met the false discovery rate criterion after correction across all 1,300 assays (Fig. 4d; Supplementary Table S11).

We compared eight variants identified in a study of post-challenge insulin responses, aligning each to the allele associated with lower insulin sensitivity.^15^ The SAPA projection (T) was close to zero for rs117643180-A at *SLC2A4* (−0.00266, reference standard error 0.01726) and positive for rs2972144-G at *IRS1* (0.08466, 0.00573) (Fig. 4e). For the *IRS1*-region allele, MI odds ratios were 1.047 (95% CI, 1.031 to 1.063) in MVP and 1.045 (1.025 to 1.064) in FinnGen. For the *SLC2A4*-region allele, the MI and stroke intervals included 1 in both sources (Fig. 4f). At *SLC2A4*, lower blood pressure largely offset the contributions of higher TG, HbA1c and CRP to the projection. At *IRS1*, seven of the eight marker contributions pointed towards higher SAPA (Fig. 4g). The same alleles also differed in their associations with regional fat deposits (Fig. 4h, Supplementary Table S12).

Across the genome, the fixed SAPA direction captured an estimated 91.1% of the eight markers’ linear genetic information for MI and 81.9% for survey-reported stroke in MVP. In FinnGen, the estimates were 76.3% for MI and 71.8% for stroke excluding subarachnoid haemorrhage (Fig. 5b). Intervals were broad, particularly for stroke, and some upper limits exceeded 100%. Genetic PC1 (78.5–94.2%) and equal weights (79.8–96.6%) had higher point estimates, without a paired superiority test. An alternative LD score regression specification also placed most of the panel’s linear genetic information along SAPA (Supplementary Note 3).

**Figure 5.**
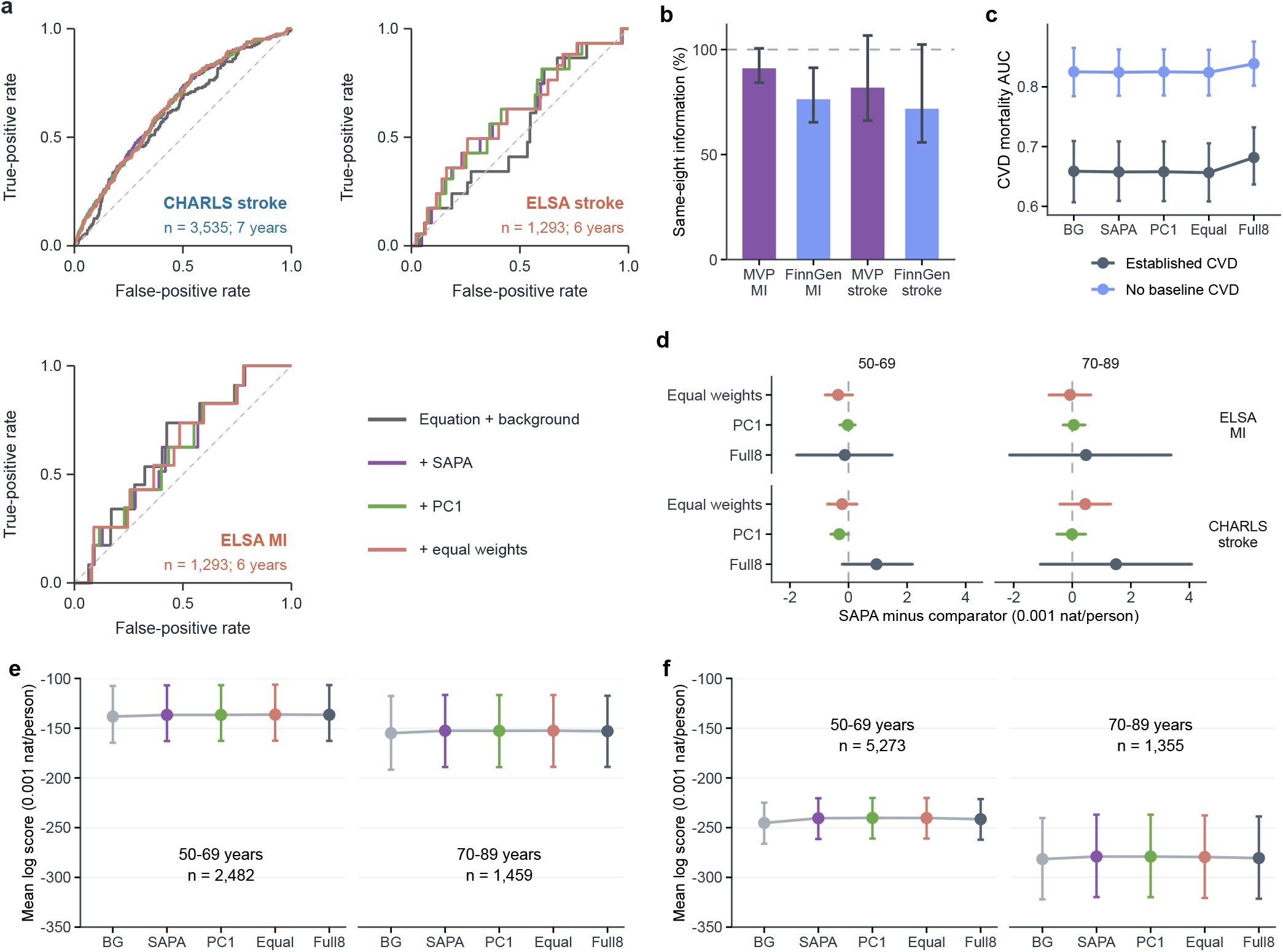
Predictive information across representations and outcomes **a**, Held-out, censoring-weighted receiver operating characteristic (ROC) curves within lower-risk categories: CHARLS stroke (n = 3,535; approximately seven years), ELSA stroke and myocardial infarction (MI; n = 1,293 each; six years). The equation/background model and score extensions share participants, folds and censoring weights. Curves use interval-midpoint event times; Fig. 2 and Supplementary Table S5 give uncertainty and timing comparisons. **b**, Bias-corrected fractions of same-panel linear genetic information retained by the SAPA direction; approximate reflected 95% intervals from 5,000 joint-error draws, without truncation at 100%. Stroke definitions follow Fig. 4. **c**, Optimism-corrected cardiovascular disease (CVD) mortality AUC in NHANES without baseline CVD (n = 6,573; 215 deaths) and with CVD (n = 1,043; 146 deaths); 95% percentile intervals from 500 within-cycle participant refits. **d**, Age-specific SAPA-minus-comparator log-score differences. **e,f**, Absolute log scores for ELSA MI observation records (e) and CHARLS stroke conditional on observed survival (f). Higher values favour prediction. Participant/report counts are 2,482/59 and 1,459/42 for ELSA; 5,273/332 and 1,355/103 for CHARLS, whose age-analysis sample includes 514 deaths. Models share background, targets and folds and average sexes equally. Bars in d–f are pointwise 95% intervals from 1,000 resamples conditional on predictions; units are 0.001 natural-log units per person. AUC, area under the ROC curve; PC1, first principal component; Full8, all eight markers entered separately. Supplementary Notes 2–3 and Supplementary Table S13 define the targets and comparisons. <u>Source data</u>.

### Clinical prediction gains varied across outcomes

The lower-risk comparisons in Fig. 5a assessed the value of adding SAPA to conventional risk estimates. We then examined whether entering the eight markers separately improved prediction of disease reports or cardiovascular death.

In the primary analyses of MI and stroke reports, entering the eight markers separately (Full8) gave no clear predictive improvement over a single SAPA score (Fig. 5d–f). For stroke, the models predicted disease reports conditional on the recorded survival history. The relative performance of Full8 and SAPA showed no consistent difference by age (Supplementary Table S13).

In NHANES, cardiovascular disease (CVD) mortality AUCs among 6,573 participants without baseline CVD (215 deaths) were 0.825 for background, 0.824 with SAPA and 0.839 with Full8 (Fig. 5c). Among 1,043 participants with prior CVD (146 deaths), they were 0.659, 0.658 and 0.682. The SAPA mortality hazard ratio in the CVD-free sample was 1.093 (95% CI, 0.857 to 1.394); all six mortality association intervals included 1 (Supplementary Figure 1a).

HbA1c offered a clinical example of the omissions examined in Fig. 1e. We compared predictions with and without measured HbA1c among participants who had a glucose measurement and no known diabetes (Supplementary Table S3F–H). In CHARLS, adding seven-marker SAPA to age, sex, smoking and glucose increased stroke AUC from 0.617 to 0.675 (n = 5,478, paired gain 0.057, 95% CI 0.033 to 0.089, Supplementary Table S3H). In ELSA participants with normal fasting glucose, the seven-marker score improved held-out prediction of MI reports. The gain for stroke was uncertain. Across these three comparisons, measured HbA1c gave no clear further improvement once the seven-marker score was included (Supplementary Table S3G).

### Marker clustering weakened in older age groups

The share of between-person marker variation along SAPA decreased by 2.907 percentage points per decade of age (simultaneous 95% interval, −3.856 to −1.957) in NHANES 2015–2018 and by 3.109 points (−3.811 to −2.407) in 1999–2010 (Fig. 6a–c). Older groups had less variation along SAPA and more outside it. The share was also lower at ages 70–89 than at 50–69 in ELSA and CHARLS. The HRS IL-1RA sample gave a less precise age comparison (Supplementary Table S8D).

**Figure 6.**
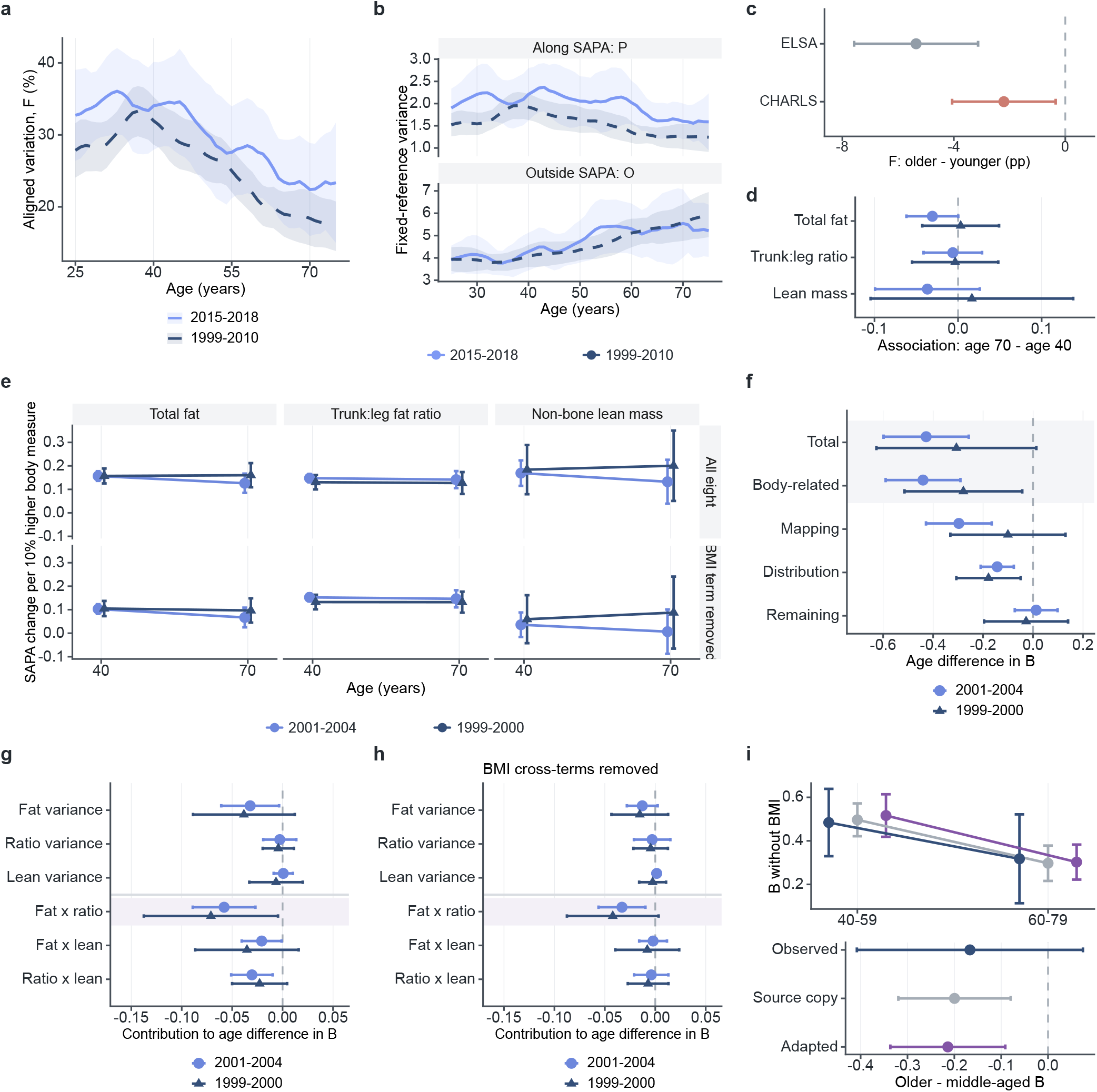
Age differences in marker clustering and their body composition context **a**, Fraction of marker variation along SAPA, F=P/(P+O). **b**, Variation along (P) and outside (O) SAPA. NHANES 2015–2018 (n = 4,000) and 1999–2010 (n = 11,053); simultaneous max-t bands from 500 survey resamples per period. **c**, F differences between ages 70–89 and 50–69 in ELSA (n = 3,941) and CHARLS (n = 6,628); six-comparison simultaneous intervals. pp, percentage points. **d**, Age-70-minus-age-40 differences in conditional SAPA associations per 10% higher total fat, trunk-to-leg fat ratio or non-bone lean mass. **e**, Associations at each age, including BMI-omitted SAPA. NHANES: n = 3,242 in 2001–2004 and 1,513 in 1999–2000. Colours/symbols distinguish periods; lines connect cross-sectional age estimates. **f**, Differences in shared cross-marker variation B between ages 60–79 and 40–59 (n = 2,081/986 across periods). Relationship and distribution terms sum to the body-related component; remaining covariance completes the total. **g,h**, Body measurement variance and covariance contributions, retaining all terms (g) or removing BMI cross-terms (h). Negative values indicate lower shared variation in older groups. Panels d–h: pointwise 95% intervals from five official imputations and 399 survey resamples per imputation, with Rubin pooling and Barnard–Rubin degrees of freedom. **i**, Model transfer from 2001–2004 (n = 2,057) to 1999–2000 (n = 978), excluding BMI cross-terms. Upper/lower plots show age-group values/contrasts. Dark blue: target observations; grey: source carry-over; purple: substitution of target BMI/waist covariance. Pointwise 95% t intervals use target-only uncertainty (14 design degrees of freedom), source-only uncertainty (30), or joint uncertainty with Welch degrees of freedom, respectively. BMI, body mass index. Extended Data Figures 6–9 and Supplementary Tables S14–S19, S22–S23 provide definitions, diagnostics and supporting comparisons. <u>Source data</u>.

Even the direction capturing the most variation at each age showed declines of 3.604 and 2.953 percentage points per decade, respectively. Its advantage over fixed SAPA had no clear age trend (Supplementary Note 7).

### Body composition helped explain age differences in marker clustering

The anatomical associations persisted across age groups. Two non-overlapping NHANES periods included 3,242 adults in 2001–2004 and 1,513 in 1999–2000. In the first period, a 10% higher trunk-to-leg fat ratio corresponded to SAPA differences of 0.148 at age 40 and 0.141 at age 70 (95% CIs, 0.130 to 0.165 and 0.105 to 0.178). The second period showed a similar pattern (Fig. 6d,e). Total fat was positively associated with SAPA in both periods. Removing BMI from the score weakened the association with lean mass. Age differences in the associations of body measures with individual markers appeared in the first period only (Extended Data Figure 6, Supplementary Tables S14–S16).

Older adults varied less in total fat, and total fat covaried less with its regional distribution. In both periods these differences contributed to the lower covariance among markers at ages 60–79 than at 40–59 (Fig. 6f–h, Supplementary Table S17). The contribution persisted after removing terms involving BMI.

Results based only on measured scans were smaller and less precise than those using the multiply imputed samples; the measured subsets also had lower BMI (Extended Data Figure 7). Analyses using observed BMI and waist showed similar distribution contributions, with weaker precision in the second period (Extended Data Figure 8; Supplementary Tables S16, S18). However, using the target population’s BMI and waist distributions did not improve predictions of marker relationships from the other period’s model (Fig. 6i; Supplementary Table S19).

## Discussion

SAPA measures a combination of routine abnormalities that recurred across populations and carried much of the panel’s linear genetic information for MI and stroke (Figs. 1, 4 and 5). Its clearest prognostic association within a conventional low-risk category was for stroke in CHARLS (Fig. 2). At similar total fat, participants with higher SAPA had more visceral and less leg fat. The association with regional fat distribution persisted at older ages, even as the markers clustered less closely (Figs. 3 and 6).

Metabolic syndrome is the clinical construct closest to SAPA. Its criteria group abnormalities associated with insulin resistance, and continuous scores quantify their severity across demographic groups.^5,8,14,16^ SAPA provides a common reference for studying what the recurring combination of test results represents across populations, extending work on biomarkers shared across diseases.^9^ The overlap was apparent for visceral fat. SAPA and Gurka’s score predicted it similarly, and combining them added little (Fig. 3e, Supplementary Table S6).

The genetic direction estimated from marker relationships captured 71.8–91.1% of the panel’s linear genetic information for MI and stroke (Fig. 5b). Earlier studies described genetic variation shared across diseases^17^ and genetically distinct forms of obesity.^18^ Two variants associated with lower post-challenge insulin sensitivity illustrate how different marker combinations map onto SAPA.^15^ At *SLC2A4*, lower blood pressure largely offset the contributions of higher TG, HbA1c and CRP, leaving the projection close to zero. At *IRS1*, most contributions pointed towards higher SAPA (Fig. 4e,g). In a separate comparison, the same *IRS1*-region allele was associated with higher MI odds in both sources (Fig. 4f).

Limited peripheral fat storage may favour lipid accumulation in organs and impair insulin action.^19^ This offers one explanation for the opposing associations with visceral and leg fat, consistent with studies of regional fat and cardiovascular risk^20^ and of regional fat depots and metabolic traits.^21,22^ The SAPA–VAT association persisted after BMI was removed from the score and included as a covariate (Supplementary Table S6). At a given total fat and after adjustment for background characteristics, SAPA retained 84.9% of the partial linear information all eight markers provided about VAT. The marker combination thus reflects differences in fat distribution among people with similar BMI.

Waist measurements allowed us to follow this relationship within individuals. In ELSA, a gain in waist circumference accompanied a rise in SAPA even after its BMI term was removed and BMI change was taken into account (Fig. 3d). Repeated DXA measurements have likewise linked changes in visceral fat to metabolic syndrome severity.^23^ In NHANES, fasting insulin and regional fat distribution each related to the six-marker score after adjustment for the other and for CRP (Supplementary Table S8G).

Adipose tissue can also secrete IL-1RA, an inhibitor of interleukin-1 (IL-1) activity.^24^ Previous studies have associated circulating IL-1RA with fat distribution and metabolic syndrome features.^25,26,27^ In HRS, blood pressure, lipid and glycaemic measurements helped predict IL-1RA after accounting for BMI and CRP (Supplementary Table S8E). Fat storage, insulin action and inflammatory responses may each contribute to the abnormalities seen on routine testing. The insulin and IL-1RA findings came from different populations. Following all three processes in the same people would help establish how they relate over time.

Previous work documented age differences in how routine markers cluster.^11^ In both NHANES periods, older adults varied less in total fat, and total fat covaried less with its regional distribution. A narrower spread in body composition can leave less variation for markers to share even when SAPA remains associated with regional fat. The distributional differences contributed to lower covariance between markers after removal of terms involving BMI (Fig. 6f–h). The overall clustering measure also includes each marker’s own variance. Evidence that individual marker associations with body measures differed by age was inconsistent across periods. Comparisons with all eight markers offered little evidence that age changed their predictive gain over SAPA (Fig. 5d–f).

Within the original low-risk China-PAR category, approximately seven-year stroke net probabilities were 2.560% and 9.351% in the groups with low and high SAPA, respectively. When participants had to qualify as low risk under four settings for unavailable China-PAR inputs, the continuous association and modest prediction gains persisted. Few stroke reports in the high-SAPA group made a three-group gradient difficult to establish in that subset (Fig. 2). A continuous metabolic syndrome score likewise showed prognostic associations for coronary events with little improvement in discrimination.^6^ Treatment selection or downward risk reclassification would require evidence on calibration, missed events and the resulting clinical decisions.^28^

SAPA, PC1 and equal weights gave similar prognostic discrimination (Figs. 2e and 5a). Fixed SAPA weights allow the same combination of tests to be studied across populations. PC1 maximises variation among the inputs along a single direction, which may help explain its modest advantage for IL-1RA. Adding SAPA to background characteristics did not improve cardiovascular mortality discrimination in NHANES (Fig. 5c).

With one test withheld, the other seven usually placed participants near their complete-score rank (Fig. 1e). Accuracy in reconstructing the missing test itself varied by marker (Extended Data Figure 3). Without HbA1c, SAPA retained prognostic information for CHARLS stroke reports and for MI reports in ELSA participants with normal fasting glucose (Supplementary Table S3G). Extending this finding to other omissions or to patients whose clinicians select which tests to order requires separate evaluation.

We defined the clinical subgroups after inspecting the cohorts. Unavailable China-PAR inputs affected classification, interview intervals left event dates uncertain, and later disease-free entry selected survivors. In the lower-risk analyses, incomplete mortality records precluded estimation of incidence accounting for competing death. ELSA subgroup estimates were imprecise, and ten-year composite risk differs from prognosis for a single endpoint over shorter follow-up. Clinical–genetic agreement depends partly on the selection of local marker relationships. NHANES scoring and DXA samples overlap, and we selected and evaluated IL-1RA in HRS. Routine markers informed DXA imputations, and participants with measured scans tended to have lower BMI. Cross-sectional fasting insulin reflects sensitivity, secretion and clearance. Waist measures adiposity indirectly, and CHARLS estimates depended on the waist range included. Measurement error and time-varying confounding can affect within-person comparisons. Treatment, birth cohort and survival may differ by age. Some Biobank Japan local parameters were unstable, genetic intervals were broad, and MR indicated possible pleiotropy (Supplementary Note 1).

Future intervention studies could test whether improvement follows the SAPA direction. Prediabetes remission without weight loss has been associated with a higher subcutaneous-to-visceral fat ratio,^29^ and lower IL-1RA after gastric bypass has accompanied reduced adiposity.^30^ In these settings, repeating the eight tests alongside measurements of fat distribution and IL-1RA, while recording medication use, would show which markers move together within an individual.

SAPA gives studies in different populations a common measure of how routine abnormalities cluster and how that pattern relates to physiology and prognosis. We provide a <u>spreadsheet calculator</u> for research use (Supplementary Data 1). The fixed score could also be tested as an interpretable input to artificial intelligence models of physiological change.

## Methods

### Study design and sources

This observational study combined clinical cohorts and genetic summary data, with analysis-specific populations and targets (Supplementary Tables S1–S2). NHANES supplied the clinical reference, linked DXA measurements and age-structure evaluations. ELSA supplied MI and stroke reports, CHARLS supplied stroke and broad heart disease reports, and HRS supplied a further stroke comparison and IL-1RA measurements.^31,32,33^ Genetic analyses used Pan-UK Biobank routine-trait data, an independently reconstructed Biobank Japan structure, MRI adiposity studies and MVP/FinnGen vascular outcomes.^34,35,36,37,38,39^ Supplementary Table S2 identifies each population, its time origin, the outcome evaluated and any overlap in participants or genetic inputs. Clinical prediction models were trained and tested within cohorts using NHANES scoring weights or cutoffs where specified. We evaluated IL-1RA in the cohort in which it was selected and tested the body composition model in a separate survey period.

### Fixed score and measurement scales

The panel comprised systolic and diastolic blood pressure, LDL cholesterol, HDL cholesterol, triglycerides, HbA1c, BMI and CRP. Before reference standardisation, we took the natural logarithm of triglycerides and CRP and kept the other six markers on their original scales. The NHANES reference was derived from 4,565 fasting participants in 2015 to 2018, including 4,265 with complete observed panels. The clinical SAPA score was Q=Σλⱼzⱼ, where each zⱼ used the reference transformation, mean and standard deviation. The clinical scoring coefficients were fixed after estimation in NHANES. Supplementary Note 1 gives the coefficients to six decimal places, the reference parameters and the SDs used to express Q for each analysis. For the genetic analyses, w denotes the direction estimated from trait covariances. Genomic structural equation modelling (Genomic SEM) estimates variant effects on the genetic SAPA factor (gSAPA). T projects marginal genome-wide association study (GWAS) effects directly onto fixed weights on the traits’ native scales.

The accompanying workbook (<u>Supplementary Data 1</u>) calculates observed and medication-adjusted Q using these constants and provides weighted reference percentiles for NHANES, ELSA and CHARLS where the input treatment is compatible. A separate calculator page supports one omitted measurement using the remaining fixed contributions and omission-specific reference ranks. A fasting glucose estimate of HbA1c was developed in NHANES 2015–2016 and evaluated without refitting in the next cycle and the existing ELSA and CHARLS panels (Supplementary Table S3). This option also supports one further omitted marker, retaining the corresponding seven-marker reference (Supplementary Table S3E).

The clinical factor model retained the three local relationships selected from the initial genetic correlation matrix: SBP–DBP, HDL–TG and BMI–CRP. The selection rule screened all 28 pairs using |r_g|>0.50 plus physiological rationale; these data-informed choices were then fixed for the cross-source comparisons.

For repeated NHANES blood pressure measurements, the first available reading was omitted when more than one was present, with the single-reading and diastolic-zero rules given in Supplementary Note 1.

Missing reference inputs were completed by a single random forest imputation before estimating the weighted reference scale and factor coefficients. Reference uncertainty was conditional on that completion. Supplementary Note 1 supplies the full measurement rule, predictor set, coefficients and structural-bootstrap diagnostics. A ten-seed sensitivity analysis of the identifier used in reference imputation is reported in Supplementary Table S20.

Observed and medication-adjusted inputs used the same reference. Principal clinical results use observed measurements and the score SD specified for each task. Age-comparable NHANES structure and missing-marker reconstruction use the reference blood pressure handling and reference parameters. External clinical and genetic structural comparisons retain their native scales.

Prognostic evaluation after omitting HbA1c used the post-measurement CHARLS and ELSA populations without known diabetes. Matched interval-observation models compared background information, reduced and complete SAPA, measured HbA1c and individual markers using five-fold prediction and 200 cluster resamples (Supplementary Note 2; Supplementary Table S3F–H).

### Clinical associations, risk strata and ranking

NHANES-reference ELSA analyses included 3,941 participants with 101 MI reports and 3,956 with 142 stroke reports. Risk sets required an endpoint-negative Wave-7 interview after the Wave-6 nurse examination and excluded earlier positive histories. Interval-observation Weibull models adjusted for age using a three-degree-of-freedom natural spline, sex and smoking, with household-cluster direct-score sandwich intervals. Score SDs were 1.426816 for MI and 1.425993 for stroke. Mortality ascertainment was incomplete.

CHARLS entry used the disease-negative 2013 interview after baseline blood measurement. First positive reports were identified from the harmonised diagnosis and disputed-prior-diagnosis fields and carried forward; pre-entry positives were excluded. Stroke analyses included 7,423 participants with positive follow-up and 461 reports from 7,555 eligible participants. Broad heart disease included 6,178 with follow-up and 611 reports from 6,282 eligible participants. Weighted illness-death models split baseline onset and disease-free mortality rates at three years, adjusted for age, sex and smoking, and used community-cluster sandwich intervals. Both used the score SD of 1.499031 from the original 8,601-person panel. Broad heart disease included the questionnaire’s coronary disease, heart failure and other heart problems.

The HRS stroke analysis included 2,282 participants and 82 reports, with score SD 1.513622. Its stroke-negative 2018 landmark followed the 2016 blood draw by a median 1.795 years. A four-state observation model integrated known and unknown disease paths around death. Supplementary Note 2 specifies cohort observation rules, time scales and covariates; Supplementary Table S4 gives all clinical associations.

Five-fold held-out four-year predictions retained households or communities within folds. ELSA MI and stroke used the same 3,941 and 3,956 people; CHARLS stroke and heart disease used 6,628 and 5,455 aged 50 to 89. Quartile cutoffs were calculated separately in each cohort and outcome-analysis sample from the fixed reference-scaled scores. ELSA summaries were Weibull event probabilities; CHARLS summaries were cumulative onset probabilities before death, including paths ending in death after onset. Five hundred cluster resamples kept predictions and score cutoffs fixed.

NHANES stroke history used survey-weighted logistic regression in 4,256 complete-panel participants, including 170 histories. The 1999 to 2010 mortality analyses included separate samples without baseline CVD (n = 6,573) and with prior CVD (n = 1,043). Eight-year survey-weighted Cox models adjusted for age, sex, smoking and cycle. Mortality prediction comparisons used a separate internal optimism correction.

### Analyses within lower published-risk categories

The principal lower-category display uses a post-measurement landmark analysis. CHARLS entry required a disease-free, diabetes-negative 2013 interview after the 2011–2012 blood-collection period. ELSA entry required a Wave-7 interview strictly after the Wave-6 nurse month and absence of earlier cardiovascular disease or diabetes. Original scores, age, other covariates and category membership were retained. China-PAR <5% defined the original CHARLS category, with northern-China and atherosclerotic cardiovascular disease (ASCVD) family-history indicators set to zero where unavailable; urban/rural status was measured. A sensitivity set required <5% under all four settings of the two unavailable inputs and used each participant’s maximum scenario probability. ELSA retained the age-specific lower SCORE2 categories. These comparisons condition on disease-free survival and attendance at the later visit; the published equations were not reassessed at that visit. HRS comparisons within lower categories of the Pooled Cohort Equations had only two stroke reports and insufficient fold support for a comparable prediction analysis; eligibility and the limited association estimates are reported in Supplementary Table S21.

Weighted 25th and 75th percentiles among 4,265 complete observed NHANES reference panels defined low SAPA (Q≤−1.4862), high SAPA (Q≥1.0250) and the intervening group. The unrounded cutoffs in Supplementary Note 2 were applied unchanged across cohorts and risk categories. Net event probabilities were one minus Kaplan–Meier survival at the approximately seven-year wave-based horizon in CHARLS and six years in ELSA. Events were assigned to interval midpoints or first positive reports; non-events were censored at their last explicit negative report. One thousand fold-stratified household/community resamples provided pointwise intervals for group probabilities and paired differences. Incomplete mortality information prevents identifying competing-risk incidence from these analyses. Supplementary Table S5A–E gives flow, observation support and both timing rules.

Five-fold prediction used 5,481 CHARLS and 2,110 ELSA training participants across the qualifying score range and evaluated their original lower categories. The calibrated baseline contained a three-degree-of-freedom spline of the published-score logit, age, sex and smoking. Matched extensions each added one linear term for SAPA, PC1 or equal weights. Training-fold censoring distributions were estimated within the relevant lower category; Supplementary Note 2 gives the full model specifications. Two hundred complete four-model refits were attempted per setting. After warnings arose, qualification required all four models to give finite estimates without fitting warnings. Reportable intervals required at least 180 qualified refits: CHARLS retained 194–199, ELSA midpoint stroke/MI 195/192, and ELSA first-positive stroke/MI only 77/130. Qualified percentile intervals are conditional on successful fitting. Qualification counts and the reasons for withholding unstable intervals are reported in Supplementary Table S5E.

### Regional adiposity, longitudinal change and biochemical measurements

The anatomical analysis linked observed routine measurements and observed DXA in 1,876 NHANES 2015–2018 participants aged 20 to 59. The earlier, multiply imputed DXA analyses described below addressed age and body composition in different survey cycles. Survey-aware models included total fat and its square, age and age squared, sex, race/ethnicity, education and cycle. The five regions were VAT, abdominal SAT, trunk, whole-leg and gynoid fat. Within-cycle measurement ordering was unavailable.

Genetic MRI comparisons related T to abdominal SAT, thigh SAT, VAT and liver proton-density fat fraction, conditional on a bioelectrical impedance analysis (BIA) whole-body-fat trait; the studies shared UK Biobank participants.

Matched construct comparisons and imaging-free prediction exchanged survey cycles for training and testing, with 60 survey jackknife replicates and t-based intervals. Supplementary Note 5 and Supplementary Table S6 specify their distinct populations, predictors and direct comparisons.

Liver-fat–disease genetic covariance was separated into a component along T, an additional Full8 component and the residual beyond Full8. Joint summary-error covariance supported delta-method intervals and 12-test Holm correction within each of two linkage disequilibrium score regression (LDSC) specifications (Supplementary Note 5; Supplementary Table S6).

We also related repeated waist measurements to changes in SAPA in ELSA waves 4 and 6 (n = 2,959), the earlier waves 2 and 4 (n = 2,246), and CHARLS waves 1 and 3. Separate weighted regressions of two-visit means and changes estimated between-person and within-person associations per 10 cm of waist.

Models included baseline age, sex, smoking and visit interval; the BMI-conditioned versions also included mean BMI or BMI change. Q used the fixed reference coefficients without further SD scaling; the BMI-omitted version removed only that fixed term. The primary CHARLS comparison required both waists to be 40–200 cm (n = 5,207), with all positive values retained in a separate check (n = 5,404). This broad review range was fixed before model fitting. We used inherited follow-up blood-sampling weights and 999 shared household-bootstrap resamples for pointwise 95% intervals and direct contrasts. ELSA timing came from nurse visits; CHARLS interview timing was a proxy. Supplementary Note 5 and Supplementary Table S7 specify the paired-window, medication and measurement checks.

Fasting insulin analyses used 2,475 NHANES 2001–2004 participants aged 20–79 years with complete markers, verified measured DXA, positive fasting weights and no reported diabetes diagnosis or glucose-lowering treatment. Borderline diabetes responses were eligible. Insulin assays were harmonised using the Centers for Disease Control and Prevention (CDC) equation. Survey-weighted models related insulin and regional fat distribution to a fixed projection excluding BMI and CRP, with adjustment for background characteristics, body composition and CRP. Three primary comparisons used Holm adjustment and pointwise intervals from 999 stratified primary sampling unit resamples. Cycle-specific, native-assay and laboratory-hyperglycaemia-exclusion estimates are reported in Supplementary Table S8F–J. Supplementary Note 6 specifies eligibility, scales and resampling.

Serum IL-1RA came from the HRS 2016 Venous Blood Study additional-assay release, version 2 (April 2025).^40^ The analysis included 3,464 participants from 2,680 families, with five family-based folds. Models for log IL-1RA shared age, sex, smoking, measurement interval, fasting and collection-time terms. SAPA, PC1 and equal-weight models used the same reference scale, matched flexibility and sex interactions. Two hundred family resamples repeated fitting for uncertainty and the specified Holm families.

A component comparison entered BMI and log CRP separately, then added each frozen projection with those two terms removed. The three remaining six-marker projections had matched flexibility; a comparator entered all six markers individually. Every added predictor used a three-df natural spline and linear sex interaction. Two hundred family resamples repeated all six model fits within the original folds; seven paired mean squared error (MSE) comparisons formed one Holm family (Supplementary Table S8E).

The median core-visit to venous blood interval was 58 days; HbA1c came from dried blood spots. The complete preceding six-marker screen is in Supplementary Table S8. The held-out analysis evaluated the selected IL-1RA model; marker selection was not repeated within training folds. Supplementary Note 6 also describes training at ages 55 to 69 and evaluation at ages 70 to 84, with target calibration and testing separated by family.

### Genetic information and multilevel genetic analyses

Cross-trait analyses used the fixed genetic direction and the genetic correlation and structural equation framework.^41,42^ For each vascular endpoint, linear genetic information represented by that direction was compared with the information expressible by all eight traits. The two information quantities were bias-corrected using 5,000 joint Gaussian summary-error draws, retaining the cross-covariances among trait and disease estimates. Their corrected ratio and paired reflected-draw percentile interval were reported without clipping at 100%. Supplementary Note 3 gives the corresponding numerator, denominator and genetic-PC1/equal-weight comparisons.

Genomic structural equation modelling estimated gSAPA variant effects using the multivariate sampling-error structure; the complete summary contained 6,666,346 variants. Functional-source projection T applied fixed weights to eight marginal effects on their native GWAS scales; covariance analyses used a genetically standardised projection, defined in Supplementary Note 5. Genetic correlations used the multivariate covariance and sampling-error specification documented in Supplementary Note 4.

MR used allele-harmonised outcomes and inverse-variance weighted multiplicative random-effects estimation.^43^ Both instrument rules began at *P* < 5×10⁻⁸; the axis-consistent rule also required seven-degree-of-freedom heterogeneity *P* ≥ 0.05 before separate linkage disequilibrium (LD) clumping (r²<0.001 within 10 Mb). The independently clumped instrument sets were not nested. Minimum instrument F statistics were 29.776 and 29.772 for the Broad and AxisConsistent sets. Cochran Q and results from four MR estimators are reported in Extended Data Figure 5 and Supplementary Table S9.

MR-Egger intercepts used the same harmonised inputs, with positive-exposure allele orientation, multiplicative dispersion floored at one, t-based intervals and seven-endpoint Holm correction within each set; correction across all 14 tests is also reported.^44^

Local genetic analyses used Local Analysis of [co]Variant Association (LAVA), its 2,495-region partition, a UK Biobank LD reference and two scale-equivalent sample size conventions.^45^ Fig. 4c presents the zero sampling-error-correlation reference. Sensitivity analyses used matched LDSC intercepts from the Default and Chi80 specifications, preserving eligibility and repeating cross-source selection. A post-fit diagnostic held the original replication correction fixed. Supplementary Note 4 and Supplementary Table S10 give the settings, selection criteria and pointwise intervals.

Using the deCODE plasma protein resource,^46^ the protein screen selected 1,300 assays with molecularly eligible cis anchors; 966 could be matched to complete gSAPA. Benjamini–Hochberg adjustment retained all 1,300 assays. Source eligibility and anchor selection are detailed in Supplementary Note 4 and Supplementary Table S11.

Eight representatives with BMI-unadjusted insulin sensitivity effects, drawn from a fixed 11-region list of post-challenge functional discoveries, were aligned to the insulin sensitivity index (ISI)-lowering allele.^15^ For each allele, T was the weighted sum of its eight marginal marker effects. For the same alleles, we report marginal MI and stroke odds ratios in MVP and FinnGen with pointwise 95% intervals (Fig. 4e–g, Supplementary Note 4). Reference errors and wider unknown-covariance bounds, all marker contributions and 24 marginal MRI associations are supplied with Supplementary Table S12.

MVP MI used PheCode 411.2, an electronic-record diagnosis grouping for MI. MVP stroke (GCST90476623) was the survey disease-diagnosis endpoint CircStrk, comprising 18,436 cases and 297,232 controls in the genetically inferred European-ancestry group. The public dictionary supplies no stroke-subtype or subarachnoid haemorrhage exclusion rule. FinnGen release 12 used I9_MI_STRICT and I9_STR, the latter excluding subarachnoid haemorrhage. Sources and endpoints were analysed separately, with shared routine-trait inputs and possible study overlap considered in interpretation.

### Age-related structure and disease information

For marker covariance Σ and the unit-normalised fixed direction u, aligned variation was *P*=u′Σu, remaining variation O=tr(Σ)−P and the aligned share F=P/(P+O). NHANES 2015 to 2018 and 1999 to 2010 analyses used the NHANES reference scale, survey designs and background adjustment, with simultaneous intervals for the specified age contrasts (Supplementary Note 7). For comparison, M=λmax(Σ)/tr(Σ) gave the largest share captured by any direction in each age-specific covariance matrix; M−F measured the gap from the fixed direction. All used the same reference metric. Correlation-spectrum and paired ELSA comparisons, using within-population correlation matrices, are shown separately in Extended Data Figure 9.

Disease-record models compared ages 50 to 69 with 70 to 89 in 3,941 ELSA participants and 6,628 CHARLS participants. Held-out log-prediction gains targeted the MI observation record and the stroke record conditional on observed survival. Direct age contrasts used 1,000 resamples conditional on fitted predictions; joint and survival-record quantities were secondary (Supplementary Note 2 and Supplementary Table S13).

### Body composition and age differences in clustering

Official NHANES 2001 to 2002 and 2003 to 2004 whole-body DXA releases were linked to observed eight-marker panels. (https://wwwn.cdc.gov/nchs/nhanes/dxa/Dxa.aspx) After excluding pregnancy and unusable scans, 3,242 participants aged 20 to 79 entered mean-relation analyses; 2,081 aged 40 to 79 entered structural comparisons. Predictors were log total fat mass, log trunk-to-bilateral-leg fat ratio and log total non-bone lean mass, using the published mass-corrected values. Two-year fasting-subsample weights divided by two defined the combined survey, with cycle-specific strata and primary sampling units. (https://wwwn.cdc.gov/nchs/nhanes/tutorials/weighting.aspx)

Five official DXA imputations were analysed separately (<u>CDC technical documentation</u>). Sex-specific weighted models included an age natural spline, height, cycle, the three body measures and their interactions with linear age. Contrasts represented 10% higher body measures at ages 40 and 70, holding the other measures constant and averaging sexes equally. Fixed weights converted marker effects into SAPA effects; the BMI-omitted version removed only its contribution. Three age differences were evaluated jointly using complete Rubin covariance and a finite-degree-of-freedom F approximation.^47^ Individual contrasts used Barnard–Rubin intervals;^48^ their P values were adjusted together using Holm’s method.

Structural comparisons separated ages 40 to 59 and 60 to 79 within sex and cycle. Identical weighted adjustment for age and height preceded the projection of markers onto body measures. Marker covariance C was decomposed as C=MSM′+R, with body covariance S, body-to-marker mapping M and remaining covariance R. Shared cross-marker variation was B(C)=u′Cu−Σuⱼ²Cⱼⱼ. The body-related age difference was partitioned into relationship and distribution terms, retaining its exact signed sum. Matrices were averaged equally across sexes and cycles. The distribution term was further separated into three body measurement variances and three pairwise covariances. Supplementary Note 8 gives the formulas and all comparison definitions.

A verified measured subset required no-imputation flags and invariance of required body values across all five versions. It comprised 2,668 participants for mean relations and 1,676 for structure. Twenty-three participants in the mean-relation sample, including 17 in the structural comparison, had zero imputation flags but arm/leg-overlap tissue-quality codes and varying values. The main analysis retained all five released versions; the measured subset excluded these participants. Requiring valid regional tissue-quality codes reproduced the same measured subset. The same 2,057 waist-complete participants supplied paired DXA and BMI/waist comparisons. Descriptive sample profiles used original fasting weights and observed sex/cycle composition. Supplementary Table S22 describes the primary and measured-subset populations.

The same rules were applied separately to NHANES 1999 to 2000: 1,513 participants for mean relations, 986 for structure, verified measured subsets of 1,215 and 769, and 978 for the matched body measurement comparison. Single-cycle fasting weights were used directly. The design comprised 13 strata and 27 primary sampling units, giving 14 complete-design degrees of freedom. The samples had no participants in common; their estimates were not pooled. A frozen transfer comparison from 2001 to 2004 to 1999 to 2000 used the waist-complete samples, holding source mappings and residual covariances fixed while substituting target BMI/waist covariance. Target marker values supplied evaluation and diagnostics. For quantities depending on both periods, source and target uncertainty were propagated jointly; squared-error improvement used an approximate finite-reference joint-ellipse projection interval. Supplementary Table S23 provides the corresponding replication-population profiles.

### Analysis planning, uncertainty and reproducibility

Analyses used previously examined datasets and were not preregistered. Reference parameters and comparison sets were fixed for the stated downstream analyses. The post-measurement landmark design followed review of baseline timing and was selected as the principal lower-category display to establish measurement before follow-up. The CHARLS observation model followed a time-fit diagnostic, and IL-1RA was evaluated in its selection resource. Supplementary Note 9 records these decisions and the scope of subsequent diagnostics.

Intervals apply to individual estimates unless labelled simultaneous. We handled multiplicity within each specified family, including six paired age comparisons, seven MR outcomes per instrument set and estimator, and 1,300 protein assays. Supplementary Note 9 also lists the anatomical, biochemical and body composition families. Where a Holm-adjusted P value accompanies a pointwise interval, the adjustment applies to the P value alone. We report percentile or basic-bootstrap intervals as specified for each analysis. Resampling fitted predictions holds the model fit fixed. We identify the approximate intervals used for genetic information and transfer between survey periods.

Body composition analyses used 399 survey-design bootstrap resamples drawn from the full positive-weight design universe before domain restriction, with identical primary sampling unit draws across imputations. Rubin pooling combined within- and between-imputation uncertainty. Finite-sample references followed the multivariate approximation and Barnard–Rubin interval procedures.^47,48^ The complete-design degrees of freedom were 30 for the combined 2001 to 2004 cycles, 15 for either individual cycle and 14 for 1999 to 2000. Supplementary Note 8 records the finite-reference approximation, numerical checks and transfer errors. Supplementary Tables S14–S19, S22–S23 give full body composition estimates, measured-subset comparisons, sample profiles, component contributions and transfer diagnostics. Aggregate estimates supporting the figures and tables are provided as Source Data.

### Artificial intelligence assistance

Large language models (LLMs) were used for code generation, suggestions for figure design and assistance with manuscript writing. Analyses of the original data were performed using locally executed statistical scripts. No image generation models were used. The authors reviewed the AI-assisted work and take responsibility for the final manuscript.

## Supporting information

Supplementary tables, figure source data and SAPA calculator

## Data availability

Aggregate results supporting the figures and supplementary tables are supplied as Source Data. The source-specific access routes and release identifiers are listed in <u>Data source and access details</u> and Supplementary Note 9. Participant-level cohort data and third-party GWAS files are obtained from their original providers under the applicable access conditions.

NHANES data for 1999–2010 and 2015–2018 are provided by the Centers for Disease Control and Prevention, National Center for Health Statistics, U.S. Department of Health and Human Services, Hyattsville, MD (<u>CDC/NCHS data portal</u>; accessed 19 October 2026), including the <u>DXA data resources</u>. ELSA data are available through the UK Data Service (Study 5050; analysed package: 50th Edition, https://doi.org/10.5255/UKDA-SN-5050-37).^31,49^ CHARLS data, including Harmonized CHARLS Version D (June 2021), are available through the <u>CHARLS portal</u>.^32^ HRS core and longitudinal data are available to registered users through the <u>HRS data portal</u>; the biomarker and venous blood files used here require approved sensitive-data access.^33,40^ These include the 2016 main Venous Blood Study version 3 and Additional Assays version 2 (April 2025), produced and distributed by the University of Michigan, Ann Arbor, MI, with funding from the National Institute on Aging (NIA U01AG009740; HRS data portal accessed 19 October 2026). HRS participant-level files are not redistributed with this paper. Product-specific HRS data citations are provided in Supplementary Note 9.

The eight Pan-UK Biobank source GWAS records are available through the NHGRI-EBI GWAS Catalog under GCST90691918, GCST90691891, GCST90691894, GCST90691577, GCST90691583, GCST90691576, GCST90691769 and GCST90691573.^35,50^ Biobank Japan sources are NBDC releases hum0014.v8.58qt.v1 and hum0014.v6.158k.v1.^38,39^ MVP outcomes use GCST90475932 (MI), GCST90475936 (coronary atherosclerosis), GCST90476623 (survey-reported stroke) and GCST90477991 (cerebral infarction).^36^ FinnGen outcomes use release 12, with I9_MI_STRICT, I9_CORATHER and I9_STR, through the FinnGen access route.^37^ The four MRI fat-distribution records are GCST90590314–GCST90590317; the distinct whole-body-fat source is GCST90691779.^34,35^ deCODE protein summary statistics are available through the access route accompanying the source publication.^46^ The post-challenge insulin sensitivity estimates came from the supplementary tables of Williamson et al.^15^ Source roles, identifiers and GWAS Catalog download dates are listed in the access table.

Summary statistics for 6,666,346 gSAPA variants are deposited in an unpublished Zenodo record, version 1.0 (reserved DOI: 10.5281/zenodo.21513053). The dataset is accessible to reviewers through a confidential preview link.

## Code availability

The macro-free SAPA spreadsheet calculator, with visible formulas and embedded reference constants, is provided in <u>Supplementary Data 1</u>. The analyses were implemented using R, Python and supporting command-line workflows.

Analysis code and documentation are held in an unpublished Zenodo Software record (version 1.0; reserved DOI: 10.5281/zenodo.23452518) and are available to journal editors and reviewers through a confidential link.

## Ethics and data use

This study used previously collected NHANES, ELSA, CHARLS and HRS data and genetic summary statistics. The investigators did not recruit or contact participants, collect biospecimens or perform laboratory assays. Genetic analyses used summary statistics rather than participant-level source-biobank data. All participant-level data were de-identified before use in this study.

NHANES was approved by the National Center for Health Statistics Research Ethics Review Board, ELSA by the relevant National Health Service research ethics committees, and CHARLS by the Peking University Institutional Review Board. Source studies obtained participant consent. HRS data were accessed under an approved sensitive-data agreement, with source-study consent procedures described in the HRS documentation.^31,32,33,40^ Source-study approval details are provided in Supplementary Note 9.

Ethics and consent procedures for the genetic resources are described in their source publications.^34,35,36,37,38,39,46^

## Author contributions

Junhua Wu: Conceptualization, Methodology, Formal analysis, Writing - original draft, Writing - review and editing. Qiuxia Chen: Methodology, Formal analysis, Data curation, Validation, Writing - original draft, Writing - review and editing. Xi Chen: Methodology, Formal analysis, Data curation, Validation, Writing - original draft, Writing - review and editing. Min Wu: Data curation, Formal analysis, Validation, Writing - review and editing. Shuai Mi: Data curation, Formal analysis, Validation, Writing - review and editing. Hyo Jung Shin: Methodology, Formal analysis, Validation, Writing - review and editing. Hee-Jung Song: Formal analysis, Data curation, Validation, Writing - review and editing. Dong Woon Kim: Conceptualization, Supervision, Project administration, Writing - review and editing. Zhenhua Zhou: Conceptualization, Methodology, Supervision, Project administration, Writing - original draft, Writing - review and editing.

## Data Availability

Aggregate results, figure source data, editable supplementary tables and the SAPA calculator are provided in the accompanying supplementary materials and supporting-data archive.
Participant-level cohort data and third-party genetic summary statistics must be obtained from their original providers under the applicable access conditions. NHANES data are publicly available through the CDC/NCHS data portal. ELSA data are available through the UK Data Service (Study 5050), CHARLS data through the CHARLS data portal, and HRS data through the HRS data portal. HRS biomarker and Venous Blood Study files require approved sensitive-data access. Data sources, release identifiers, GWAS accessions and access routes are detailed in the manuscript, Supplementary Note 9 and Source_Data/Data_Access_Identity.tsv.
The complete gSAPA summary statistics are deposited in an unpublished Zenodo record, version 1.0 (reserved DOI: 10.5281/zenodo.21513053), and are available for confidential journal review. Analysis code and documentation are held in a separate unpublished Zenodo Software record, version 1.0 (reserved DOI: 10.5281/zenodo.23452518), and are available to journal editors and reviewers through a confidential link. These records are not currently publicly accessible.

https://wwwn.cdc.gov/nchs/nhanes/

https://doi.org/10.5255/UKDA-SN-5050-37

https://charls.charlsdata.com/

https://hrsdata.isr.umich.edu/

https://www.ebi.ac.uk/gwas/

https://humandbs.dbcls.jp/en/research/hum0014/v8

https://www.finngen.fi/en/access_results

https://download.decode.is/form/folder/proteomics

## Acknowledgements

We thank the participants and staff of NHANES and the CDC National Center for Health Statistics for making the survey data available. We thank the ELSA participants and study team and the UK Data Service for access to Study 5050. We thank the China Center for Economic Research, the National School of Development of Peking University for providing the data.

Research reported in this publication was supported by the National Institute On Aging of the National Institutes of Health under Award Number R01AG017644. The content is solely the responsibility of the authors and does not necessarily represent the official views of the National Institutes of Health. ELSA is funded by the NIHR Policy Research Programme (HEI) 198_1074_03. The views expressed are those of the author(s) and not necessarily those of the NIHR or the Department of Health and Social Care.

This analysis uses data or information from the Harmonized CHARLS dataset and Codebook, Version D as of June 2021 developed by the Gateway to Global Aging Data. The development of the Harmonized CHARLS was funded by the National Institute on Aging (R01 AG030153, RC2 AG036619, R03 AG043052). For more information, please refer to https://g2aging.org/.

The HRS (Health and Retirement Study) is sponsored by the National Institute on Aging (grant number NIA U01AG009740) and is conducted by the University of Michigan.

The supplementary stroke analysis uses Early Release data from the Health and Retirement Study, 2022 HRS Exit (Early Version 1.0), sponsored by the National Institute on Aging (grant number NIA U01AG009740) and conducted by the University of Michigan. These data have not been cleaned and may contain errors that will be corrected in the Final Release version of the dataset.

We acknowledge the participants and investigators of the FinnGen study. We thank BioBank Japan and its participants and investigators, and the <u>NBDC Human Database</u>, for the publicly released summary statistics. We acknowledge the <u>Pan-UK Biobank project</u> and UK Biobank for the source GWAS results, the Million Veteran Program participants and investigators for vascular GWAS summary statistics, deCODE genetics for protein-QTL results, and the NHGRI-EBI GWAS Catalog for distributing the source records.

