## Supplementary information for "A shared cardiovascular and cerebrovascular risk marker phenotype: physiological context and prognosis"

The guide links each Results section to its methods and supporting tables. Methods are grouped by analytical domain. Supplementary Tables S1–S2 define resources, analysis populations, time origins and validation roles. The coordinate and effect-scale guides in Supplementary Note 1 distinguish the clinical and genetic quantities.

### Guide to the Supplementary Notes

| Results topic | Main figure | Supplementary Notes | Principal tables |
|---|---|---|---|
| Common profile across sources | Fig. 1 | Note 1 | Supplementary Tables S1–S3, S20 |
| Clinical associations and post-measurement risk categories | Fig. 2 | Note 2 | Supplementary Tables S4–S5, S21 |
| Fat distribution, waist change, insulin and IL-1RA | Fig. 3 | Notes 5 and 6 | Supplementary Tables S6–S8 |
| Global, local and functional genetic evidence | Fig. 4 | Note 4 | Supplementary Tables S9–S12 |
| Information across representations and outcomes | Fig. 5 | Notes 2 and 3 | Supplementary Tables S3F–H, S5 and S13 |
| Age differences and body composition | Fig. 6 | Notes 7 and 8 | Supplementary Tables S14–S19, S22–S23; Supplementary Table S8D |

### Abbreviations

| Abbreviation | Full name or definition |
|---|---|
| AI | artificial intelligence |
| ASAT | abdominal subcutaneous adipose tissue |
| AUC | area under the receiver operating characteristic curve |
| BBJ | Biobank Japan |
| BIA | bioelectrical impedance analysis |
| BMI | body mass index |
| CDC | Centers for Disease Control and Prevention |
| CFA | confirmatory factor analysis |
| CFI | comparative fit index |
| CHARLS | China Health and Retirement Longitudinal Study |
| CI | confidence interval |
| CRP | C-reactive protein |
| CVD | cardiovascular disease |
| DBP | diastolic blood pressure |
| DXA | dual-energy X-ray absorptiometry |
| ELSA | English Longitudinal Study of Ageing |
| FDR | false discovery rate |
| FMI | fraction of missing information |
| Full8 | model including all eight routine markers |
| gSAPA | genetic shared adverse physiological axis |

| Abbreviation | Full name or definition |
|---|---|
| GWAS | genome-wide association study |
| HbA1c | glycated haemoglobin |
| HDL | high-density lipoprotein |
| HR | hazard ratio |
| HRS | Health and Retirement Study |
| IL-10 | interleukin-10 |
| IL-1RA | interleukin-1 receptor antagonist |
| IL-6 | interleukin-6 |
| ISI | insulin sensitivity index |
| IVW | inverse-variance weighted |
| LAVA | Local Analysis of [co]Variant Association |
| LD | linkage disequilibrium |
| LDL | low-density lipoprotein |
| MAE | mean absolute error |
| MetS | metabolic syndrome |
| MI | myocardial infarction |
| MR | Mendelian randomization |
| MRI | magnetic resonance imaging |
| MSE | mean squared error |
| MVP | Million Veteran Program |
| NGSP | National Glycohemoglobin Standardization Program |
| NHANES | National Health and Nutrition Examination Survey |
| NT-proBNP | N-terminal pro-B-type natriuretic peptide |
| Pan-UKB | Pan-UK Biobank |
| PCE | Pooled Cohort Equations |
| PC1 | first principal component |
| pQTL | protein quantitative trait locus |
| PSU | primary sampling unit |
| RAPS | robust adjusted profile score |
| RMSE | root mean square error |
| RMSEA | root mean square error of approximation |
| ROC | receiver operating characteristic |
| SAH | subarachnoid haemorrhage |
| SAPA | shared adverse physiological axis |
| SAT | subcutaneous adipose tissue |
| SBP | systolic blood pressure |
| SD | standard deviation |
| SRMR | standardized root mean square residual |
| TG | triglycerides |
| TGF-β1 | transforming growth factor-β1 |
| TNF | tumour necrosis factor |
| VAT | visceral adipose tissue |

Symbols F, P, O, B, D, Q and T are defined at their first use in the relevant analysis; D denotes the frozen clinical reference when used as a reference label. Source variable names and accession identifiers retain their original spelling.

Supplementary Note 1 | Scoring, structural comparisons and marker reconstruction

For an input $x_j$, $z_j = (T_j(x_j) - \mu_{j,D}) / \sigma_{j,D}$, and the portable projection is $Q = \Sigma_j \lambda_j z_j$. Scoring uses the fixed coefficients below. The full-precision CFA loadings document the model from which they were derived. The unit vector $u = \lambda/||\lambda||$ is used for orthogonal variance decomposition. Analysis-specific score centring and SDs are given separately for the disease and biochemical tasks.

Observed and medication-adjusted inputs share the NHANES reference. For participants receiving treatment, the adjustments add 10 and 5 mmHg to systolic and diastolic pressure, respectively, multiply LDL by 1.3, and add 0.5 percentage points to HbA1c, as applicable to each treatment. HDL, triglycerides, BMI and CRP retain their measured values. The same adjustment applies to everyone receiving the relevant treatment. The main clinical analyses use observed inputs and follow each source analysis's rules for unknown medication status.

| indicator | calculator_reference_unit | transformation_before_standardisation | NHANES_reference_mean | NHANES_reference_sd | projection_loading_frozen |
|---|---|---|---|---|---|
| SBP | mmHg | identity | 125.889299739127 | 19.3283521104695 | 0.392562 |
| DBP | mmHg | identity | 72.3085558109613 | 11.8285951530586 | 0.2875 |
| LDL | mg/dL | identity | 117.720489114109 | 36.3155706912103 | 0.352646 |
| HDL | mg/dL | identity | 55.1045830570953 | 17.3360977057968 | -0.413997 |
| lnTG | mg/dL | natural log | 4.537647063786 | 0.592184147889648 | 0.629933 |
| HbA1c | NGSP % | identity | 5.74264956338763 | 1.08363802210029 | 0.442657 |
| BMI | kg/m^2 | identity | 29.5624364493185 | 7.13127073285116 | 0.448779 |
| lnCRP | mg/L | natural log | 0.596453219072588 | 1.23541639029462 | 0.423349 |

Spreadsheet implementation and reduced-input checks

Supplementary Data 1 is the editable SAPA calculator using the fixed NHANES reference. It provides measured-input and medication-adjusted scores, with interpretation guidance and the study-specific CHARLS stroke example (Supplementary Table S5A–C). Its calculation and reference sheets give the formulas, units, transformations and sources. Both complete-input branches use the same means, SDs and six-decimal loadings to calculate the portable coordinate Q. Unknown medication status suppresses only the adjusted branch.

Reference percentiles use blood-sample weights in participants with biomarker measurements. NHANES 2015–2018 includes 4,265 complete measured panels and 4,565 participants in the imputed NHANES reference, aged 20 or older (age 80 is top-coded), with WTSAF4YR. ELSA Wave 6 includes 4,260 participants aged 50–89 with positive w6bldwt; CHARLS baseline includes 8,335 participants aged 45–101 with positive Weight_Bio. ELSA provides percentiles for measured inputs only because its medication correction covered blood pressure alone. Regional reference percentiles are available for NHANES, ELSA and CHARLS. The HRS sample was selected for the stroke analysis.

Flexible inputs provides a measured seven-marker branch for exactly one unavailable measurement. For marker j, $Q_{-j} = \Sigma_{(k \neq j)} \lambda_k z_k$ retains the seven frozen contributions, without rescaling the sum or re-estimating weights. The missing laboratory value remains unfilled. The workbook supplies a separate weighted P1–P99 reference for each omission within each cohort (24 columns). It reports the omitted marker, reduced score and matching ranks. Complete-score clinical group probabilities are not assigned to these reduced scores. Zero or invalid laboratory values, incorrect units and multiple omissions do not activate this branch; the original blood pressure rule below is retained.

We assessed each omission by masking one measurement in complete panels (Supplementary Table S3A). Weighted Pearson correlations, score bias and RMSE describe agreement with full Q. Rank differences use each score's own within-cohort reference. Each integer rank is the greatest stored percentile cutoff at or below the score, with the tails represented by 0 and 99. Quartile disagreement counts participants whose relative position changed. Figure 1e shows the complementary percentage remaining in the same reference quartile, calculated as 100% minus the percentage changing quartile. The calculator itself displays <1 below P1 and >=99 at/above P99. These agreement estimates apply to simulated omissions in complete panels. Prognosis after HbA1c omission is examined in Supplementary Note 2.

For missing HbA1c, we specified a two-coefficient fasting glucose bridge before examining its performance. Survey-weighted least squares in 2,168 NHANES 2015–2016 adults gave estimated HbA1c (%) = 2.8439268321 + 0.0260297063 × fasting plasma glucose (mg/dL). The workbook stores full coefficient precision and uses 18.0182 mg/dL per mmol/L. It restores the estimated HbA1c contribution to $Q_{-HbA1c}$ using the existing NHANES loading, mean and SD. We applied the frozen equation to NHANES 2017–2018 (n=2,097), ELSA recorded fasting samples (n=2,438) and CHARLS recorded glucose (n=8,325). The NHANES reference uses both NHANES cycles; the temporal holdout applies to the bridge coefficients. ELSA fglu is labelled as glucose in fasting samples, with fastelig=1. CHARLS fasting-code labels could not be confirmed in the retained primary dictionary; raw codes 1 and 2 are reported without assigning clinical meanings.

The main implementation checks restrict glucose to the development range of 21–479 mg/dL; nine CHARLS values fall outside it. The limits are the lowest and highest training values. Supplementary

Table S3B–D reports calibration, HbA1c reconstruction and recorded treatment/diabetes strata. The omission comparator adds no HbA1c contribution, which algebraically corresponds to the D mean for the HbA1c error calculation. In ELSA the score using the glucose estimate correlated more closely with the complete score, yet its RMSE was higher. The fasting glucose section on the Flexible inputs sheet is labelled as a research trial and accepts only confirmed fasting plasma glucose. With only HbA1c missing, it gives an estimated eight-marker Q with the NHANES full-score reference. With HbA1c and one other marker j missing, it combines the six measured contributions with the estimated HbA1c contribution and uses the NHANES reference omitting j. The other marker remains unfilled. This is a separately named seven-marker estimate, without rescaling or cohort-specific refitting. The HbA1c percentage can be calculated from qualified glucose even when the remaining measurements are insufficient for a SAPA score. An invalid additional value is not treated as an omitted test. The glucose bridge used no random-glucose conversion, medication-corrected branch or external refit. Prognostic evaluation of the seven-marker omission is described in Supplementary Note 2.

Native Microsoft Excel checks covered 347 synthetic scenarios and 5,552 score/rank comparisons, including one-marker omissions, seven HbA1c-plus-one-other combinations, unit conversions, fasting status, invalid values and development-range limits. Twenty representative input workflows added 320 comparisons. All matched independent arithmetic, with maximum absolute difference below 10^−15. The workbook contains no participant records, macros or external workbook dependencies.

For the additional omission j, we masked HbA1c and j in the validation panels and compared the glucose estimate with both measured Q−j and complete Q (Supplementary Table S3E). We applied the fixed glucose model within its development range in NHANES 2017–2018 (n=2,097), ELSA recorded fasting samples (n=2,438) and CHARLS recorded glucose (n=8,316). For Q−j, the only estimation error comes from the HbA1c contribution. Its RMSE is therefore identical for all seven choices of j. Correlation, complete-Q error and rank movement depend on the other omitted marker. Ranks for the estimated and measured Q−j use the same cohort-specific omission reference. The calculator uses the NHANES reference. These comparisons quantify score agreement under simulated omissions.

Blood pressure measurements and reference completion

The clinical reference comprised 4,565 NHANES 2015–2018 fasting participants, of whom 4,265 had complete observed marker panels. Blood pressure readings were considered in their recorded order. Finite, non-negative readings were retained; a sole reading was used directly. When several readings were available, the first available reading was omitted and the remaining readings were averaged. For diastolic pressure, zero readings were removed from that average when positive retained readings existed. If only zeros remained after omitting the first reading but an original positive reading existed, the original positive reading was retained. All-zero diastolic records remained zero; this occurred in 14 reference participants. A separately saved sensitivity treated those records as missing.

Medication adjustments were applied before the log transformations and reference estimation. Missing transformed values were completed with the mice random forest method using ranger, one completed dataset, five iterations, ten trees per forest, seed 2026 and one computational thread. The predictor set contained the eight transformed marker variables, age, sex, race/ethnicity, education and the numeric participant identifier SEQN. SEQN was included as an auxiliary predictor in the saved implementation and has no physiological interpretation. The predictor matrix, run specification and logged predictor events document that implementation. Disease outcomes were absent from the predictor set. Observed values were preserved.

Survey-weighted means and population-denominator SDs defined the marker scale. The factor model allowed local relationships between SBP and DBP, HDL and log triglycerides, and BMI and log CRP. The fitted standardized loadings were rounded to six decimal places for the portable projection. Reference uncertainty used 60 stratified delete-one-PSU jackknife refits conditional on the single completed dataset. Between-imputation variation and uncertainty attributable to the imputation specification were not included. The blood pressure and completion rules apply to the NHANES reference; external cohort-native structures and genetic estimates retain their own measurement scales.

### Identifier sensitivity of the clinical reference

We assessed the influence of the numeric participant identifier SEQN on the random forest completion used to construct the clinical reference. The analysis retained the original 4,565 participants, transformations, medication adjustments, survey weights, auxiliary variables and factor model. The original seed-2026 completion was reproduced exactly. For each of ten prespecified seeds (2026–2035), completion was repeated with and without SEQN as a predictor of the eight markers, retaining SEQN as a record-linkage key. Both branches used one completed dataset, five iterations and ten trees per forest with the ranger backend and a single computational thread. Weighted reference means, population-denominator SDs and factor loadings were re-estimated; loadings were rounded to six decimal places for coordinate comparisons.

To assess propagation through the reference independently of changes in imputed individual values, both reference versions were applied to the same 4,265 complete marker panels. Observed and medication-adjusted inputs were evaluated separately. Directional comparisons expressed the revised coefficients in the original NHANES-standardised marker space. We calculated coordinate correlations, survey-weighted differences and changes in within-sample quartile membership. The ten-seed ranges describe algorithmic variation and are not sampling confidence intervals or pooled multiple-imputation inference.

At seed 2026, excluding SEQN changed the clinical direction by 0.436°. For observed inputs, the correlation with the original coordinate was 0.999989, and the weighted root-mean-square difference was 0.010 of the original coordinate SD. In each paired run, differences were divided by the weighted SD of that run's with-SEQN coordinate; at seed 2026 this equals the original NHANES-reference score SD. Across the ten paired runs, observed-coordinate correlations were 0.99970–0.999994 and root-mean-square differences were 0.005–0.024 SD. The corresponding maximum individual differences were 0.024–0.113 SD. The original reference was retained.

Differences between completed values were reported separately for the originally missing cells. Similar individual-level variation was also present when SEQN was retained and only the random seed changed. This diagnostic measures sensitivity of the reference and complete-case scores to the imputation settings. The original jackknife intervals condition on a single completion. Accuracy of the missing values was unavailable, and downstream outcome models were held fixed.

Supplementary Table S20 summarizes the diagnostic. Its linked aggregate tables contain all twenty parameter sets, ten paired comparisons and the random-seed context. The accompanying data contain aggregate summaries.

Coordinate definitions

| Symbol | Definition and input | Use |
|---|---|---|
| Q (clinical SAPA) | Sum of the eight formal clinical coefficients times NHANES-standardised, transformed measurements | Portable clinical coordinate; any analysis-specific centring and scaling are stated separately |
| u (clinical) | Clinical coefficient vector divided by its Euclidean norm | Fixed direction used to calculate F and B in the age analyses |
| w (genetic) | Frozen genetic direction on the eight genetically standardized traits | Same-panel genetic information comparison in Supplementary Note 3 |
| T | Fixed projection. Functional-source displays use native-scale, allele-aligned effects, $T = a'\beta$. Covariance analyses use a fixed direction on genetically standardised traits and standardise the resulting projection to unit genetic variance. | Functional-source comparisons in Supplementary Note 4; regional-adiposity and liver-fat covariance comparisons in Supplementary Note 5 |
| gSAPA | Factor-GWAS effect estimated by genomic structural equation modelling | Full genetic association, MR and local analyses; its estimation weights incorporate the sampling-error structure |

The clinical u and genetic w arise from different fitted structures. T uses a single frozen coefficient vector across variants; the gSAPA factor-GWAS does not use that same constant-weight calculation. Supplementary Note 4 supplies the T coefficients and uncertainty calculation.

### Reading the effect scales

The marker reference, total-score scaling and endpoint scale are separate choices. The clinical reference transformation stays fixed even when a disease model expresses Q per an analysis-specific SD. Unit changes do not change a ranking, but they change the numerical effect reported per unit.

| Reported quantity | Unit or denominator | Where used |
|---|---|---|
| Clinical association per SD | Q divided by the stated analysis SD: ELSA MI 1.426816, ELSA stroke 1.425993, CHARLS stroke/heart disease 1.499031; HRS stroke 1.513622 | Fig. 2a; Supplementary Table S4. CHARLS retains its original 8,601-person panel SD rather than recomputing it in each landmark sample |
| Stroke-history association | Q per complete observed-reference SD, 1.81544499486556 | NHANES, Supplementary Table S4; Supplementary Figure 1a |
| Mortality association | Q per the fixed SD within each mortality sample | Supplementary Table S4; Supplementary Note 2 |
| Lower-category continuous association and fixed group thresholds | Raw Q units; no total-score SD rescaling | Continuous associations, Supplementary Table S5B. Fixed groups, Fig. 2b–d and Supplementary Table S5C. Exact thresholds, Supplementary Note 2. |
| Waist-change association | Change in Q with the BMI term removed, per 10 cm of waist change | Fig. 3d; Supplementary Table S7 |
| IL-1RA mean association | Geometric-mean IL-1RA ratio per reference SD of SAPA | Fig. 3g; Supplementary Table S8 |
| Body composition mean association | Change in Q per 10% higher body measure, at specified ages | Fig. 6d,e; Supplementary Tables S14, S16 |
| gSAPA MR | Disease odds ratio per genetic-factor unit | Fig. 4b; Supplementary Table S9 |
| Genetic projection T and its marker contributions | Weighted sum of native-scale marginal marker effects per ISI-lowering allele | Fig. 4e (T) and 4g (marker contributions). Fig. 4f gives the marginal MI and stroke odds ratios per copy of the same allele. Supplementary Table S12 gives both scales. |
| Information-retention fraction | Information along the fixed direction divided by the same eight inputs' linear information for that target | Figs. 3 and 5; not a fraction of all disease risk or all heritability |
| F and M | Fractions of marker variation along the fixed direction (F) and the best age-specific direction (M) | Fig. 6a,c show F. Supplementary Note 7 reports F and M on the same scale |
| B and its age components | Shared cross-marker variation in the fixed reference metric, excluding individual marker variances | Fig. 6f–h; Supplementary Note 8 |

In the machine-readable files, D denotes the NHANES clinical reference and Frozen_gSHL denotes the fixed genetic SAPA direction.

### Cross-source and repeated-measurement structure

The Biobank Japan comparison combines two public summary-data releases. SBP, DBP, LDL cholesterol, HDL cholesterol, triglycerides, HbA1c and CRP came from the 58-quantitative-trait release hum0014.v8.58qt.v1 (1 May 2018), associated with Kanai et al.[1] BMI came from hum0014.v6.158k.v1 (8 September 2017), associated with Akiyama et al.[2] Trait-specific sample sizes and native scales were retained. Publication-wide totals, which can include additional traits or replication samples, were not substituted for the analysis-specific counts. The source publications identify the GWAS resources; the local reconstruction and its bootstrap specification are described below.

Fig. 1b,c and Extended Data Figures 1a and 2a,d compare the same clinical samples aged 40–54: 1,104 NHANES, 3,127 CHARLS and 413 ELSA participants. NHANES uses the blood pressure summary and single completed reference dataset specified above, with unweighted robust maximum-likelihood CFA and the three specified local relationships. Clinical CFI is the robust CFI; SRMR comes from the same fitted model. Tucker congruence compares each clinical loading profile with the fixed Pan-UKB genetic profile. NHANES intervals use 500 participant-bootstrap refits, conditional on the single reference completion and fixed genetic vector; all 500 converged and yielded finite congruence estimates. CHARLS and ELSA intervals also use 500 participant-bootstrap samples on their stated measurement scales. Covariance matrices, all 28 named marker pairs per source and model-fit summaries specify each comparison. ELSA between-wave correlations are descriptive point estimates without interval summaries.

Extended Data Figure 1 supplies SRMR, ELSA constrained-minus-unconstrained fit changes, and genetic and repeated-wave local relationships. NHANES SRMR uses the fixed reference dataset. Genetic and ELSA repeated-wave comparisons use their stated inputs; genetic intervals come from parametric bootstrap. The complete BBJ BMI–CRP interval is displayed with a labelled axis break.

The retained eight-loading Wald-type equality comparison between Pan-UKB and Biobank Japan gave $\chi^2 = 54.084$ with 8 degrees of freedom ($P = 6.65 \times 10^{-9}$). The joint test compares exact loadings. The cosine describes overall orientation and permits individual differences, including the near-zero LDL loading in Biobank Japan (Extended Data Fig. 2c).

Genetic structural-bootstrap diagnostics

The cross-source genetic comparison used 1,000 parametric draws per source from the saved joint summary-error distribution, followed by refitting the original factor model. Pan-UKB yielded 1,000 regular results. BBJ yielded 582 regular results, 353 finite results with a negative observed-trait residual variance (Heywood solutions), 52 missing model-result tables and 13 draws with a non-positive input diagonal. The original rule retained finite loadings from both regular and Heywood results, yielding 935 BBJ results for the reported intervals. The point solution was admissible.

Genetic loading and local-relationship intervals use the model output's fully standardized estimates (STD_All). The BBJ BMI–CRP local interval was −7.952 to 0.4956. Finite output from an unconstrained model can include unstable, inadmissible bootstrap solutions; the interval therefore needs the accompanying fit-status information. The paired direction-cosine interval was 0.736 to 0.962 using all 935 finite BBJ draws and 0.840 to 0.966 using only the 582 regular draws. The latter interval is conditional on a regular fit. Fit-status counts, local-parameter intervals and the regular-only diagnostic are provided together.

Missing-marker reconstruction

Extended Data Figure 3 uses the covariance matrix from the fixed NHANES confirmatory factor model, including the factor loadings, factor variance, residual variances and three local residual relationships. For each omitted indicator j, its conditional linear reconstruction is $\Sigma[j,-j]\Sigma[-j,-j]^{-1}z[-j]$. The reconstructed marker and the seven observed markers then enter the fixed NHANES scoring equation with coefficients given to six decimal places. The implementation normalizes its weight vector to unit length; this common positive scaling leaves the reported Pearson correlations unchanged. Performance is evaluated in the same six measurement panels: ELSA waves 2, 4, 6 and 8 and CHARLS waves 1 and 3, preserving participant eligibility and input preprocessing. All 48 marker-by-panel combinations are retained. Points and ranges are the mean and minimum–maximum Pearson correlations across these panels, separately for the omitted marker and completed coordinate. The ranges describe variation across panels. This conditional-reconstruction task differs from dropping one term from a score or substituting glucose for HbA1c.

Reference scales across analyses

NHANES structure, reduced-input comparisons and clinical prediction use the fixed NHANES reference. Cohort-native structures and genetic estimates use their stated measurement scales. NHANES correlation spectra in Extended Data Figure 9 include 1,017 participants aged 40–54, 1,286 aged 55–70 and 644 above 70, with observed inputs and positive weights. These spectra standardize within each population; Fig. 6 evaluates a fixed reference-scale direction. Clinical-association populations and post-measurement prediction populations are specified separately in Supplementary Tables S4–S5.

Selection of local relationships and uncertainty in structural comparisons

The initial genetic correlation screen considered all 28 marker pairs using $|r_g|>0.50$ together with physiological rationale. It selected SBP–DBP ($r_g=0.793$), HDL–TG (−0.616) and BMI–CRP (0.576). The retained specification was applied to the subsequent clinical and genetic comparisons. This is a data-informed selection rule; prospective preregistration is not claimed. The current source-specific fit comparisons and fixed clinical scoring parameters retain their separate roles.

The cross-source tests of equality give $\chi^2 = 54.084$ on 8 degrees of freedom ($P = 6.65 \times 10^{-9}$) for the standardised loading vector. The separate unit-direction test gives $\chi^2 = 51.524$ on 8 degrees of freedom ($P = 2.08 \times 10^{-8}$). The loading-vector statistic is a Wald quadratic form for the eight between-source loading differences, using their bootstrap covariance. Its degrees of freedom equal the retained covariance rank, eight under the original eigenvalue tolerance of $10^{-8}$ times the largest eigenvalue. This Wald statistic tests equality of loadings. The direction cosine measures their relative similarity. Fig. 1c's clinical intervals condition on the retained Pan-UKB comparison profile. ELSA reported intervals for unstandardised factor covariances and point estimates for standardised factor correlations. Fig. 1d displays the latter.

Supplementary Note 2 | Clinical associations, discrimination and age-specific information

CHARLS interval-observed stroke occurrence

The CHARLS analysis required baseline age at least 45 years, complete original biomarker measurements, no reported baseline stroke, and a stroke-negative actual 2013 interview. Retrospective evidence of pre-entry stroke was used in eligibility. First positive reports were restored from the harmonized dispute fields (code 6) before defining baseline, entry and follow-up histories. Once restored, a positive report was retained at later waves. Earlier-positive participants were excluded at entry. The updated stroke sample comprised 7,555 eligible participants, 7,423 with positive follow-up and 461 observed reports. The observed-input D projection was standardized using its mean and SD in the original 8,601-person panel (−0.388372345820215 and 1.49903099588137); these values were not re-estimated in the landmark sample. Baseline blood collection was in 2011–2012 at the study level; exact individual draw dates were unavailable. Entry used the actual 2013 interview, and subsequent Core and Exit records were handled as interval observations through the 2018 observation window. No fixed day was substituted for an unknown onset or death date.

The illness–death model allowed transitions from stroke-free survival to stroke, from stroke-free survival to death, and from stroke survival to death. Unknown stroke status before death was integrated over compatible paths. Baseline stroke and stroke-free death rates were split at three years, following a time-fit diagnostic. A time-homogeneous model provided a sensitivity comparison. Both adjusted for 2013 age, sex and baseline smoking and used normalized blood-sample weights, community-cluster sandwich variance and t intervals with 410 degrees of freedom. These models do not reproduce every component of the sampling design. Post-stroke mortality slopes were linked to the stroke-free death process; inference depends on observation-process and Markov assumptions. Observed positive reports are not a count of all latent transitions integrated by the model.

CHARLS broad heart disease occurrence

The endpoint comprises reported heart attack, coronary heart disease, angina, congestive heart failure and other heart problems. Eligibility uses a heart-disease-negative actual 2013 interview and the same first-positive-history principle. The eligible sample has 6,282 participants, including 6,178 with positive follow-up and 611 reports; 104 supply no positive follow-up interval. Observed SAPA scores use the original 8,601-person mean and SD specified above. The two-period, weighted illness-death model shares the stroke model's background, three-year baseline split and community-cluster sandwich framework. Endpoint-specific records and state paths are constructed separately.

ELSA post-examination myocardial infarction reports

The analysis included participants aged 50–89 with complete measured biomarkers, no prior reported MI in the available history, and an MI-negative Wave-7 interview after the Wave-6 nurse examination. CRP and triglycerides were restricted to at most 10 mg/L and 10 mmol/L, respectively. Participants needed subsequent definite disease information. The 3,941-person analysis included 101 positive reports and 3,231 households. In this population, the measured-input SAPA score had mean 0.111078840050 and SD 1.426815895356. Smoking was recorded at Wave 6 and age at follow-up entry.

MI onset was bounded by the last recorded negative and first recorded positive report. Participants without a positive report were right-censored at their last definite negative report. The unpenalized Weibull interval model adjusted for a three-degree-of-freedom natural spline of age, sex and baseline smoking. We converted the accelerated-failure-time coefficient to a proportional-hazards coefficient using the fitted scale parameter. Household sandwich variance was obtained from direct individual log-likelihood derivatives, including coefficient–log-scale covariance, with $G/(G-1)$ correction and t intervals on $G-1=3{,}230$ degrees of freedom. Direct derivatives were used because the local survival implementation's log-scale residual for interval records did not agree with the independently differentiated likelihood.

A linked death year was available for 199 participants. Exact death dates and any unknown pre-death MI status remained missing. Incomplete death ascertainment precluded estimation of MI cumulative incidence accounting for competing death. The association applies to participants with available follow-up and assumes conditionally noninformative observation and censoring.

ELSA post-examination stroke reports

The endpoint-specific Wave-7 stroke-negative population includes 3,956 participants aged 50–89 with complete observed inputs, no earlier positive stroke history and subsequent definite information. There were 142 positive reports. The score SD is 1.425993. Risk entry, measurement support restrictions, interval construction, age spline, sex and smoking adjustment, and household direct-score sandwich follow the MI framework. Mortality ascertainment remains incomplete, so the Weibull event probability is not a competing-risk cumulative incidence.

HRS post-blood stroke occurrence

The HRS analysis included 2,282 participants from 1,851 households whose stroke-negative 2018 interview followed the 2016 blood draw. Eligibility required valid measurements and subsequent definite disease information. Blood collection preceded entry by at least 0.777 years and a median of 1.795 years. The score mean was 0.371955400818 and SD was 1.513622190263. Observation records included explicit stroke-negative Exit reports and unknown stroke status before death.

The four-state model distinguished stroke-free survival, survival after stroke, death without stroke and death after stroke. Compatible paths were integrated when observations did not determine the state. Stroke-onset and stroke-free-death intensities each included an intercept, a three-degree-of-freedom natural spline of age, sex, baseline smoking and the fixed standardized SAPA coordinate. Post-stroke death shared the death-process slopes and had an additional rate multiplier. Stroke and death baseline intensities could each change after three years, under the specified observation model. All 17 parameters were estimated without a penalty in the stated target sample.

The reported hazard ratio refers to the stroke-onset transition, not cumulative incidence or joint predictive gain. The household sandwich variance used the individual likelihood scores and observed information, $G/(G-1)$ correction and t intervals with 1,850 degrees of freedom. Interpretation depends on the specified transition and observation models, including the shared post-stroke mortality slopes and available-follow-up selection. The analysis was unweighted and does not claim national representativeness.

NHANES stroke history and mortality

The observed-input stroke-history analysis includes 4,256 complete-panel participants with valid disease and smoking data, including 170 positive histories. It uses the frozen NHANES loading projection, standardized with the mean −0.204824576561296 and SD 1.81544499486556 from 4,265 complete observed reference panels. Survey-weighted quasibinomial logistic regression includes age, sex, race/ethnicity and current smoking; the displayed intervals use the design-t reference with 21 residual degrees of freedom.

Mortality analyses use NHANES 1999–2010 participants aged at least 40 years with complete observed inputs, positive fasting weights and valid eight-year linked mortality data. Four complete baseline disease-history items define the samples without CVD (6,573 people) and with established CVD (1,043). All-cause, CVD and heart disease deaths number 804/215/174 and 344/146/115. Cause-specific survey-weighted Cox models adjust for age, sex, smoking and cycle, with score SD fixed within each mortality sample. Normal intervals use the survey-robust variance. Prediction comparisons retain eight-year weighted logistic models with the same background. Five displayed models are background, SAPA, PC1, equal weights and Full8. Five hundred within-cycle participant bootstrap samples repeat model fitting and the optimism correction; these intervals describe internal validation, separately from the survey-design association intervals. This mortality PC1 retains the original unweighted PCA of the recent complete observed-input NHANES-standardised panel.

Four-year probabilities by score quartile

Five-fold models keep each household or community together. ELSA MI and stroke evaluate 3,941 and 3,956 people; CHARLS stroke and heart disease evaluate the age-50–89 populations of 6,628 and 5,455. Each person's predicted probability comes from a model fitted outside their fold. Quartile cutoffs are calculated separately within each cohort and outcome-analysis sample using frozen NHANES-reference SAPA values, with counts 985/985/985/986, 989/989/989/989, 1,657 in each quartile, and 1,364/1,364/1,363/1,364. ELSA uses Weibull event probability at four years; CHARLS sums the illness-death probabilities for onset while alive, including paths ending in death after onset. Model backgrounds retain the cohort-specific terms specified for disease prediction. Quartile means summarise model-predicted four-year probabilities. The 500 household or community resamples keep the fitted predictions and quartile cutoffs fixed.

Clinical association intervals are pointwise and conditional on the fixed score reference, analysis populations and specified models. They do not incorporate reference-estimation uncertainty, unmeasured confounding, informative observation or disease-report misclassification. The cohorts had been used in exploratory project analyses. ORs and HRs with different score scales and endpoint definitions were not pooled; differences in statistical significance were not treated as tests of cohort heterogeneity.

HRS lower-PCE supplement

The HRS analysis retained the existing 2,282-person stroke-negative 2018 landmark and evaluated eligibility for the original Pooled Cohort Equations using contemporaneous 2016 age, blood pressure, cholesterol, smoking and antihypertensive-treatment fields.[3] It required ages 40 to 79, non-Hispanic White or Black race/ethnicity, total cholesterol 130 to 320 mg/dl, HDL 20 to 100 mg/dl and systolic pressure 90 to 200 mmHg.[4] Values outside these ranges were excluded rather than clipped. Published sex/race coefficients and unrounded probabilities were used. Both the 2016 and 2018 records had to explicitly deny broad heart disease and diabetes; raw records from earlier waves excluded positive or disputed histories available by entry. Harmonised ever-disease fields that could be retrospectively revised by later answers were not used for this new eligibility rule. It does not change the earlier overall HRS risk set.

Of 800 eligible people with 16 stroke reports, 218 had PCE <5%, with two subsequent stroke reports and two deaths. The latter included one path with unknown stroke status. The median interval from blood draw to entry was 1.812 years, and median last observation was 3.833 years after entry. Four-year status had one established positive, 76 supported negatives and 141 records that did not directly resolve status by that time. These unresolved records were retained in the observation likelihood. The two total reports are not two known four-year cases.

Five household-preserving folds applied training-only 75th-percentile SAPA cutoffs. Three test folds had no stroke reports, limiting the fold comparisons to descriptive counts. Continuous SAPA associations used the four-state observation probabilities from a model with constant onset and death rates, the same death rate before and after stroke, and no death covariates. The onset log-rate included either SAPA alone or SAPA and centred logit(PCE). SAPA was scaled by the original 2,282-person SD of 1.513622 raw D units. Approximate 95% profile-likelihood intervals depended on these working assumptions and asymptotic likelihood calibration. Only two stroke reports contributed to these intervals. Supplementary Table S21 presents both estimates, including their directions, alongside the sparse observation support.

PCE calculations were independently checked for all 800 eligible people, and likelihood values and gradients were checked with an independent numerical implementation. The public rounded coefficients reproduced the four worked examples within 0.1 percentage point; the White-male example was 5.3844% versus the displayed 5.3%. Coefficients and the 5% threshold were not changed to force agreement. Twelve eligible people lay between 4.9% and 5.1%, five of them below 5%. Four lower-PCE participants had LDL ≥190 mg/dl and six had HbA1c ≥6.5%, reinforcing the distinction between a low score and absence of other clinical indications. HRS had been examined previously in this project and was not an untouched confirmatory sample.

Age-specific disease observation-record models

ELSA includes 2,482 participants aged 50–69 with 59 MI reports and 1,459 aged 70–89 with 42 reports. Its completed five-fold household predictions use the interval Weibull observation model, age spline, sex and smoking. Full8 ridge tuning was internal to the outer training sample. CHARLS includes 5,273 younger participants with 332 stroke reports and 1,355 older participants with 103 reports; all are in the specified 50–89 subset. Its matched five-fold community models use age spline, sex, smoking and the original reported blood pressure/glucose treatment terms. Death processes share background variables across all candidate representations. Full8 ridge tuning uses the fixed four-value grid and internal community folds. These predictive CHARLS analyses are unweighted, distinct from the weighted clinical HR.

The ELSA target is the observed interval/right-censored MI record. The CHARLS primary target is log P(joint stroke–death record|X) minus log P(the same survival record|X). Joint and survival targets are also reported in Supplementary Table S13. Sex-standardized mean gains weight women and men equally within each age group. One thousand household/community resamples retain fixed disease predictions while refitting the structural background and covariance. Six principal older-minus-younger contrasts comprise F, SAPA-minus-background gain and Full8-minus-SAPA gain in each cohort. Their simultaneous intervals use the joint critical value 2.642686, conditional on the fitted disease models. CHARLS includes 6,628 participants, 435 stroke reports and 514 deaths. The age-70 division was fixed before this analysis and was not estimated as a biological change point.

Post-measurement comparisons within baseline lower-risk categories

Disease follow-up began at a documented disease-free visit after biomarker measurement. This design was chosen after identifying ambiguity in measurement-to-interview ordering, before fitting its outcome models. It was not preregistered. Baseline equation values and category membership were fixed, so the estimand concerns later disease-free survivors from those categories.

Fixed SAPA groups used 4,265 complete observed NHANES 2015–2018 reference panels. Scores were ordered and survey weights accumulated. Each cutoff was the first observed score reaching a cumulative weight fraction of 0.25 or 0.75, without interpolation. The analysis cutoffs, shown at full floating-point precision, were -1.4861684541779769 and 1.0250198531818573. Low SAPA was $Q \leq -1.4861684541779769$; high SAPA was $Q \geq 1.0250198531818573$; the middle group lay strictly between them. These thresholds were unchanged across cohorts and score categories.

China-PAR used the northern-China and ASCVD family-history indicators set to zero because complete current-residence and family-history fields were unavailable. All four binary settings were evaluated without changing measured inputs. The intersection required a probability below 5% in every setting and used the maximum probability as its model input. SCORE2 used the published low-risk-region recalibration: scale1/scale2 were −0.5699/0.7476 for men and −0.7380/0.7019 for women.[5,6] These baseline categories follow the equations' ten-year composite targets; the local outcomes and post-measurement horizons differ.

CHARLS entry required a 2013-wave interview in a year after 2012 with a valid month, explicit absence of stroke and broad heart disease after restoring earlier positive histories, and no reported diabetes at that visit. Baseline scores and China-PAR inputs were fixed; the four-setting intersection used the maximum scenario probability. Eligible entry interviews occurred during July–November 2013. Interview months established measurement-before-entry ordering; event durations used the common wave clock rather than exact individual anniversary dates. The nominal wave clock ran from 15 July 2013 to 15 July 2020 (7.000684 years). Negative reports after a conflicting recorded death date were excluded. Non-events were censored at the last explicit negative report.

Baseline SCORE2 eligibility was age 40–69 in the source score implementation, with recorded absence of cardiovascular disease and reported diabetes. The lower category was <2.5% below age 50 and <5% at ages 50–69. The 1,293-person evaluation set had baseline ages 42–69, including eight participants below 50. ELSA entry required a Wave-7 interview month strictly after the Wave-6 nurse month, explicit absence of stroke, MI, broad heart disease and diabetes, and no restored earlier positive cardiovascular history. Follow-up used actual interview months through Wave 10, with a fixed six-year evaluation horizon. Participants without any positive follow-up duration were counted in the flow and excluded from fitting. The original lower SCORE2 membership, baseline age and other covariates were not updated at entry. These are conditional comparisons among later disease-free survivors from the baseline-defined categories.

Each endpoint retained interval-midpoint and first-positive-report assignment. Continuous-Q Cox models adjusted for original equation logit, age, sex and smoking, with community/household sandwich intervals. Holm values across the three original-category endpoints were reported separately for each timing; the overlapping CHARLS intersection was a sensitivity comparison. Fixed external SAPA groups used 1,000 fold-stratified cluster resamples. Five-fold prediction retained the original cluster allocation and compared the flexible equation/background model with the same model plus SAPA, PC1 or equal weights. Two hundred complete four-model refits were attempted per setting. Preprocessing, model fitting and censoring estimation used training folds.

All four prediction models were unpenalised Cox models with Breslow handling of ties. The baseline used a natural cubic spline (three degrees of freedom) of $\log[p/(1-p)]$, linear age, sex and smoking. Spline knots, column means and sample standard deviations were estimated in each training fold. Each score extension added one linear degree of freedom. PC1 was the leading unit eigenvector of the survey-weighted covariance of the 4,565 completed NHANES reference panels on the fixed marker scale, oriented to agree with SAPA. Equal weights were $+1/\sqrt{8}$ for seven markers and $-1/\sqrt{8}$ for HDL. Both directions were frozen before cohort prediction; the added score was standardised with its training-fold mean and SD.

| Model | Predictors | Fitting population | Evaluation and censoring population |
|---|---|---|---|
| Equation + background | Three-df score-logit spline, age, sex, smoking | All eligible training-fold participants across score categories | Relevant lower category |

| Model | Predictors | Fitting population | Evaluation and censoring population |
|---|---|---|---|
| + SAPA | Baseline plus one linear SAPA term | Same training participants | Same lower category |
| + PC1 | Baseline plus one linear fixed-reference PC1 term | Same training participants | Same lower category |
| + equal weights | Baseline plus one linear adverse-direction equal-weight term | Same training participants | Same lower category |

Censoring survival was estimated from the relevant lower-category participants in the training fold, separately for each endpoint and timing rule. It supplied inverse-probability-of-censoring weights for the corresponding held-out lower-category participants. Outcome models used the full qualifying training-fold score range. Thus the outcome-fitting and censoring-estimation populations deliberately differed.

An explicitly negative observation at the evaluation horizon was a control. Censoring weights used the left limit of training-fold censoring survival at event times and the horizon; cases and controls required support of at least 0.10. No CHARLS event interval crossed approximately seven years. In ELSA, 12 stroke and six MI intervals crossed six years, contributing to timing sensitivity. ROC curves use these same held-out probabilities and censoring weights, with tied predictions grouped before calculating cumulative case and control weights.

After fitting warnings were observed, a numerical qualification rule required finite estimates without warnings for all four models in a resample. Intervals required at least 180 of 200 qualified refits. CHARLS retained 194–199; ELSA midpoint stroke/MI retained 195/192, and first-positive stroke/MI retained 77/130. The latter intervals were not used for inference. The first-positive ELSA continuous-stroke model also issued a possible infinite-coefficient warning for smoking. Other continuous fits and all point prediction fits completed without these warnings. Qualified intervals remain conditional on successful fitting. Group probabilities, AUCs, Brier losses and censoring weights were checked with an independent implementation. Supplementary Table S5A–E gives the results and numerical support.

Prognosis after omitting HbA1c

The post-measurement risk sets described above were used across all conventional-risk categories. Eligibility retained absence of known diabetes and cardiovascular disease at the qualifying visits. Actual HbA1c values were not used to exclude potentially undiagnosed hyperglycaemia. CHARLS included 5,478 participants with positive recorded glucose and 381 subsequent stroke reports. Its sampling-status codes entered as categorical covariates; eligibility did not require verified fasting. Normal-fasting-glucose analyses were limited to ELSA. ELSA models were trained among 1,323 participants with positive fasting glucose and the recorded fasting-eligibility flag. Primary evaluation used the 983 participants with glucose 70 to <100 mg/dL. The 70 to <110 mg/dL and all-eligible fasting groups were specified supporting populations. Follow-up horizons were approximately seven years in CHARLS and six years in ELSA (Supplementary Table S3F).

Seven-marker SAPA removed only the fixed HbA1c contribution from observed SAPA. The remaining weights, input transformations and reference scale were unchanged. Seven models used the same training and evaluation participants. Background included linear age, sex, smoking and log2 glucose, with sampling-code categories in CHARLS. Models added measured HbA1c, seven-marker SAPA, both terms with separate coefficients, complete SAPA, the seven individual markers, or all eight markers. Continuous terms other than glucose were linear. All non-intercept coefficients were standardised within the training fold and given a fixed ridge penalty of one-half their squared sum added to the negative log likelihood. The intercept and Weibull scale were unpenalised. No outcome-driven tuning or threshold search was used.

Weibull accelerated-failure-time models used the recorded last-negative and first-positive bounds directly. The likelihood contribution was the probability of an event within its interval, or survival through the last negative observation for right-censored records. Primary performance was mean held-out log probability of these records. Higher values indicated better prediction; paired differences are reported in units of 10^-3 nat per participant. The likelihood models the disease report, treating observation and censoring as ignorable. Death was not modelled separately, so incidence accounting for competing mortality could not be estimated.

Predictions used the existing five community or household folds. Two hundred within-fold cluster resamples repeated all model fits, training standardisation and censoring-weight estimation. The same draws were used for every model in a comparison. Pointwise intervals were the 2.5th and 97.5th percentiles. The principal contrasts added HbA1c to background information or to background plus seven-marker SAPA. Five supporting contrasts compared complete with seven-marker SAPA, Full8 with Full7, seven-marker SAPA with background or background plus HbA1c, and Full8 with seven-marker SAPA plus HbA1c. No equivalence margin or multiplicity-adjusted confirmatory test was specified.

Auxiliary AUC, Brier and observed-to-predicted risk comparisons used midpoint or first-positive event timing. Inverse censoring weights came from the training fold within the relevant glucose group; a required censoring-survival probability below 0.10 would make that metric unavailable. All original-sample censoring weights met this criterion. ELSA normal-glucose stroke included four reports definitely before six years and 14 intervals crossing six years; MI included nine and four, respectively. These intervals were retained in the primary likelihood. Four resamples had no first-positive-timed stroke before six years, leaving 196 AUC estimates for that timing rule. All interval-score comparisons used 200 completed refits. Numerical solutions were checked against gradients and independent Weibull probability calculations. The comparison concerns prognosis when a recorded HbA1c is masked. Diabetes screening and clinical decisions about who should be tested require evidence from routine care.

Supplementary Note 3 | Same-panel genetic information

Let G be the eight-trait genetic covariance matrix, c the vector of covariances between those traits and one disease component, and h the disease genetic variance. The fixed direction w is defined on genetically standardized traits. For each input matrix, let d = w / sqrt(diag(G)), with elementwise division. The two primitive quantities are

$A\_full = c'G^{-1}c / h$

$A\_axis = (d'c)^2 / (d'Gd\ h)$.

Their ratio A_axis/A_full describes the linear information retained by the fixed direction relative to all eight traits. The factor h cancels in the ratio. It is not a proportion of all disease risk or all genetic variance. The fixed genetic directions are supplied at full precision. This calculation holds w fixed and recalculates d from each sampled G; it differs from holding the native-scale T coefficients fixed across variants.

The error vector contained the 36 distinct entries of G, eight trait–disease covariances for each of six outcomes and the six disease genetic variances. Its full sampling covariance V retained cross-cell and cross-outcome error terms from the joint LD score regression output. The two input specifications used the default LD score regression chi-square filter and a chi-square cap of 80. The genetic direction was fixed across these specifications. For each specification, 5,000 Gaussian perturbations were generated as 2,500 antithetic error pairs. G was inverted by Cholesky factorization. Draws with non-positive trait or disease variances or a non-positive-definite G were rejected; no ridge term or nearest-positive-definite replacement was applied. Both completed specifications retained all 5,000 draws, as recorded in the numerical diagnostics.

Bias correction was applied to the primitive quantities before forming the point ratio. For a primitive vector f, with observed estimate f0 and bootstrap estimate fb, the reflected vector was fb* = 2f0 − fb. The corrected point estimate of each primitive was the mean of its reflected values, equivalent to 2f0 minus the bootstrap mean. The reported retention point estimate was the corrected A_axis divided by the corrected A_full. Its 95% interval was the 2.5th to 97.5th percentile of the ratios of the paired reflected primitives, using R's type-7 quantiles. Values were not clipped to zero or one. A denominator was flagged stable when the reflected A_full interval lay above zero and the rejected-draw fraction did not exceed 1%.

These are approximate intervals under a Gaussian summary-error model with fixed V and a fixed genetic direction. They propagate uncertainty in G, c and h, including their cross-terms. The calibration assessed the interval procedure under this error model. Each fraction uses the same eight traits in its denominator and the disease definition from its source.

The figure source table identifies the exact frozen direction, outcome row, estimate and interval in the completed retention output. MVP MI is PheCode 411.2. MVP stroke (GCST90476623) maps to CircStrk in the genetically inferred ancestry (GIA) European group of the official MVP data dictionary, document phd008759.1. Its category is Surveys / Disease Diagnosis. The 18,436 cases, 297,232 controls and total of 315,668 agree exactly with the local GWAS. The HARE version has different counts (18,606 cases and 301,029 controls) and was not used. CircStrkMed is a separate medication item. The dictionary does not supply stroke-subtype coding, a subarachnoid haemorrhage exclusion rule, the exact questionnaire wording or missing-response handling. The endpoint is therefore labelled survey-reported stroke with subtype unspecified; its controls retain the source-defined status. MVP cerebral infarction (GCST90477991; PheCode 433.21) is a separate endpoint. The endpoint clarification table accompanies Source Data. The FinnGen results use Data Freeze 12, publicly released on 4 November 2024, with endpoint codes I9_MI_STRICT and I9_STR. I9_STR excludes subarachnoid haemorrhage. Release provenance is given in the R12 documentation and R12 results browser; the source publication describes the cohort.[7] The local outcome label R12_I9_STR denotes I9_STR in that release, while I9_MI_STRICT is saved without a release prefix. The source identity table preserves these names. Different sources and endpoints are not pooled. The shared routine-trait inputs and potential cohort overlap limit interpretation as independent replications.

Same-panel genetic information retained by SAPA and comparison directions (Fig. 5b)

| Endpoint | Direction | Bias-corrected A_axis [95% interval] | Bias-corrected A_full [95% interval] | Remaining quantity [95% interval] | Retained fraction, % [95% interval] |
|---|---|---|---|---|---|
| MVP MI | Fixed SAPA direction | 0.287 [0.234, 0.336] | 0.316 [0.259, 0.367] | 0.028 [-0.002, 0.050] | 91.072 [84.246, 100.573] |
| MVP MI | Adverse equal weights | 0.305 [0.250, 0.355] | 0.316 [0.259, 0.367] | 0.011 [-0.013, 0.026] | 96.608 [91.627, 104.580] |
| MVP MI | Genetic PC1 | 0.297 [0.243, 0.347] | 0.316 [0.259, 0.367] | 0.018 [-0.010, 0.037] | 94.237 [88.195, 103.282] |
| MVP survey stroke | Fixed SAPA direction | 0.154 [0.085, 0.210] | 0.188 [0.102, 0.258] | 0.034 [-0.009, 0.067] | 81.899 [66.178, 106.699] |
| MVP survey stroke | Adverse equal weights | 0.165 [0.092, 0.223] | 0.188 [0.102, 0.258] | 0.024 [-0.018, 0.053] | 87.453 [72.959, 113.382] |
| MVP survey stroke | Genetic PC1 | 0.168 [0.095, 0.226] | 0.188 [0.102, 0.258] | 0.021 [-0.017, 0.048] | 89.091 [75.294, 113.294] |
| FinnGen MI | Fixed SAPA direction | 0.156 [0.111, 0.196] | 0.205 [0.147, 0.257] | 0.048 [0.014, 0.076] | 76.340 [65.382, 91.326] |
| FinnGen MI | Adverse equal weights | 0.174 [0.125, 0.217] | 0.205 [0.147, 0.257] | 0.031 [$-3.70 \times 10^{-4}$, 0.054] | 85.041 [74.955, 100.224] |
| FinnGen MI | Genetic PC1 | 0.169 [0.120, 0.211] | 0.205 [0.147, 0.257] | 0.036 [0.004, 0.061] | 82.350 [71.909, 97.596] |
| FinnGen stroke, excluding subarachnoid haemorrhage | Fixed SAPA direction | 0.138 [0.073, 0.191] | 0.192 [0.094, 0.270] | 0.054 [-0.003, 0.096] | 71.833 [55.807, 102.402] |
| FinnGen stroke, excluding subarachnoid haemorrhage | Adverse equal weights | 0.154 [0.083, 0.211] | 0.192 [0.094, 0.270] | 0.039 [-0.013, 0.076] | 79.788 [64.521, 111.377] |
| FinnGen stroke, excluding subarachnoid haemorrhage | Genetic PC1 | 0.151 [0.083, 0.207] | 0.192 [0.094, 0.270] | 0.041 [-0.012, 0.079] | 78.470 [63.047, 110.281] |

Fig. 5b displays the SAPA rows; equal weights and genetic PC1 provide comparison directions. Ratios use the unrounded bias-corrected numerator and denominator on the scale defined above; the remaining quantity is A_full minus A_axis. Comparisons concern the same eight-trait panel, and point differences between directions are not paired superiority tests. Negative remaining-quantity limits and fraction limits above 100% are reported. All source rows and the compact table give full precision. Amounts are displayed to three decimal places; the small nonzero limit uses scientific notation. Differences calculated from the rounded amounts can differ from the reported remaining quantity.

Genetic SAPA retention under two LD score regression input specifications

| Endpoint | Default filter: retained fraction, % [95% interval] | Chi-square cap 80: retained fraction, % [95% interval] |
|---|---|---|
| MVP MI | 91.072 [84.246, 100.573] | 91.790 [85.941, 100.181] |
| MVP survey-reported stroke | 81.899 [66.178, 106.699] | 82.978 [67.304, 110.501] |
| FinnGen MI | 76.340 [65.382, 91.326] | 77.077 [65.932, 91.629] |
| FinnGen stroke excluding subarachnoid haemorrhage | 71.833 [55.807, 102.402] | 78.683 [62.241, 104.560] |

Both specifications use the same frozen genetic SAPA direction and all eight traits as the denominator. The cap-80 specification changes the SNP chi-square filter in LD score regression, not an intercept constraint. Both retained all 5,000 perturbations. The cap-80 estimates ranged from 77.077% to 91.790%; the largest point-estimate shift was 6.850 percentage points for FinnGen stroke. Intervals remain broad and are not clipped at 100%. Full-precision sensitivity results.

### Supplementary Note 4 | Global, local and functional genetic analyses

The complete gSAPA association file contains 6,666,346 variants. Global genetic correlations use multivariate summary-data covariance and sampling-error estimates from the genomic structural equation modelling framework.[8,9] The projection T used in functional and MRI comparisons is a fixed linear combination on its stated input scale, distinct from factor-GWAS gSAPA.

For MR, genome-wide significance was $P < 5\times10^{-8}$. The Broad instrument set included all qualifying variants; AxisConsistent applied the seven-degree-of-freedom Wald heterogeneity $P \geq 0.05$ rule before clumping. Each set was clumped separately with a European GRCh37 reference, $r^2<0.001$ and a 10,000-kb window, yielding 266 and 127 instruments with 48 shared. Outcome-specific allele harmonization retained 263/126 in MVP and 249/119 in FinnGen. Fig. 4b shows the inverse-variance weighted multiplicative random-effects estimates. Extended Data Figure 5 displays all 56 IVW, weighted-median, Egger and robust-adjusted-profile-score estimates under this same specification.[10,11,12,13] The specified-model Cochran Q table contains 28 rows across seven endpoints, two instrument sets and IVW/Egger methods. Minimum F statistics before outcome harmonisation were 29.776 (Broad) and 29.772 (AxisConsistent). Heterogeneity was substantial in some comparisons: for Broad-set MVP survey stroke, Q=409.538 on 262 degrees of freedom ($P = 1.42 \times 10^{-8}$). MR-Egger intercepts used the same harmonised inputs (Supplementary Table S9). Holm adjustment covered seven outcomes per instrument set and estimator. Causal interpretation under either rule requires the exclusion restriction. The axis-consistency filter tests agreement among marker effects and remains compatible with horizontal pleiotropy and more than one underlying biological process.

Local analysis used LAVA with its original 2,495-region partition.[14] The reference analysis used UK Biobank linkage disequilibrium, assumed zero cross-trait sampling-error correlation and evaluated two scale-equivalent effective sample sizes. The two gSAPA effective-N conventions were 46,009 (unit variance) and 2,443,498 (reference scale). Both traits had to pass local univariate $P < 0.05/2{,}495$. Within a discovery family and scale, the bivariate threshold was 0.05 divided by the number of eligible regions. The union significant in either discovery scale defined the replication family. A displayed result passed discovery and replication under both scales, with matching locus dimensions, a common non-zero direction, finite estimates and no boundary cap ($|\text{rho}|<0.999999$); the replication threshold was 0.05 divided by the full discovery-union count. The display reproduces the 12 retained family–region results across ten physical regions with the corresponding MVP and FinnGen estimates paired. The original discovery and replication outputs retain pointwise 95% LAVA intervals. The complete results contain 48 rows across 12 family–region pairs, two sources and two scales. Fig. 4c and Supplementary Table S10 show the 24 unit-scale intervals; the complete two-scale output also retains the reference scale. These intervals retain the original LAVA parameter-boundary treatment and are not adjusted for the selection of regions. Replication was assessed in the regions selected at discovery, and a physical region can contribute to more than one disease family. Univariate-ineligible regions and coverage limitations remain in the qualification tables.

The deCODE resource measured plasma proteins with 4,907 aptamers in 35,559 Icelanders.[15] Source selection used the included human-assay list and conditional cis-pQTL results in Supplementary Tables 1 and 8 of Ferkingstad et al. (worksheets ST01 and ST08), and corrected variant annotations. ST08 fields included conditional rank, −log10(P) (adj.), beta in SD (adj.) and correlated PAV annotations. Eligible signals met the official conditional $P < 0.05/27{,}200{,}000$ threshold, had a standard rsID and exact annotation match, and passed the common biallelic source rules. These required resolved metadata and aligned alleles, minor-allele frequency ≥0.01, conditional $F>10$, no source-exclusion flag, and no same-gene or different-gene protein-altering-variant (PAV) annotation. The strongest eligible conditional association defined the anchor; conditional rank and rsID resolved ties. Selection used molecular information.

The source-selection cascade retained 1,300 assays with at least one eligible non-PAV cis anchor. Of these, 966 anchors could be matched to the complete gSAPA summary. Their gSAPA effects were aligned to the protein-increasing allele; Benjamini–Hochberg adjustment retained the full 1,300-assay family, with 81 meeting FDR < 0.05. The remaining 334 assays stay in Supplementary Table S11 with their availability fields. The scatterplot displays anchor-level gSAPA associations.

Aggregate diagnostics provide MR estimates, heterogeneity, harmonization, instrument counts, local eligibility and unevaluable regions. MR source tables correspond to the complete gSAPA input and the instrument rules specified above.

Fixed insulin sensitivity representatives

The discovery list comprised 11 regions associated in the source study with post-challenge insulin sensitivity or insulin fold change, including four insulin fold change and eight insulin sensitivity regions with one shared region.[16] The main functional comparison used the eight representatives with an available BMI-unadjusted insulin sensitivity index (ISI) effect in the source supplement. MTNR1B, the primary PPP1R3B representative and C2CD4A lacked that particular effect field. All three had complete eight-marker input and calculable T projections. Their BMI-adjusted ISI effects were available in a different source table. Supplementary Table S12 lists the source trait, adjustment and available effect for each region. An additional PPP1R3B signal was recorded as a secondary representative outside the 11-region comparison.

For each SNP, the eight marginal effects β and SEs were aligned to one effect allele. The genetic direction w was frozen from the original eight-trait model. With reference covariance G0 and correlation R0, the fixed coefficients were a_j = w_j / sqrt(G0_jj) / sqrt(w′R0w), giving a′G0a = 1. These coefficients were then applied to native-scale effects: T = a′β. For the BMI-omitted form, a_BMI was set to zero and the remaining coefficients were unchanged. The table below gives the full-precision coefficients.

| Trait | Fixed T coefficient a | BMI-omitted coefficient |
|---|---|---|
| SBP | 0.291431475902566 | 0.291431475902566 |
| DBP | 0.401164495384613 | 0.401164495384613 |
| LDL | 0.267786578489337 | 0.267786578489337 |
| HDL | -0.492838946583687 | -0.492838946583687 |
| TG | 0.572092010629286 | 0.572092010629286 |
| HbA1c | 0.424616865781418 | 0.424616865781418 |
| BMI | 0.58901055257504 | 0 |
| CRP | 0.74457877042963 | 0.74457877042963 |

The reference error correlation E was obtained by standardizing the saved cross-trait LD score intercept matrix: E_jk = I_jk / sqrt(I_jj I_kk). For a SNP with SE vector s, its error covariance was Cβ = diag(s) E diag(s) and SE(T) = sqrt(a′Cβa). The labelled eight-by-eight matrix is supplied. The widest SE allowed by unrestricted error correlations was sum_j abs(a_j) s_j; this is a bound conditional on the marginal SEs, not the reference SE. Component contributions were a_j β_j, with normal-approximation intervals from abs(a_j) s_j. Summed-projection intervals used their respective reference SE or bound. Source effects, alleles, BMI-adjustment versions, sample sizes and source-table cells are retained in the functional input table.

Panels e–h of Fig. 4 use the ISI-lowering allele for all eight representatives. Panels f, g and h list the regions in the same order. In Fig. 4f, MVP MI (GCST90475932) and survey-reported stroke (GCST90476623, CircStrk) use the reported odds ratios and 95% intervals. When the source effect allele was reversed, we took the reciprocal of the odds ratio and reversed the reciprocal interval limits. FinnGen R12 MI (I9_MI_STRICT) and stroke excluding subarachnoid haemorrhage (I9_STR) use exp(β) and exp(β ± 1.96 SE) after allele alignment. Each interval applies to one variant and one outcome in one source. Fig. 4g gives the 64 signed marker contributions. The separate gSAPA factor GWAS uses the variant-specific sampling covariance. The disease odds ratios in Fig. 4f are marginal effects per allele.

Sampling-error sensitivity in the local genetic analysis

Sharing an LD reference is distinct from sharing GWAS participants. The MVP and FinnGen local comparisons also reuse the same gSAPA summary, which makes the two comparisons dependent through their exposure. Neither fact alone establishes the within-pair sampling-error correlation.

Two multivariate LDSC intercept matrices cover the same complete gSAPA input and seven vascular endpoints, under the default and chi-square-80 SNP-filter settings. Trait order was matched across the covariance matrices. LAVA converts the supplied overlap matrix to correlations, giving I_gy/√(I_gg I_yy), where I is the intercept matrix. The table gives these inputs alongside the zero sampling-error-correlation reference.

| Endpoint | Default normalised intercept | Chi80 normalised intercept | Zero sampling-error-correlation reference |
|---|---|---|---|
| MVP_MI | 0.035755 | 0.035897 | 0 |
| MVP_CAD | 0.039244 | 0.036416 | 0 |
| MVP_Stroke | 0.022547 | 0.022547 | 0 |
| MVP_CerebralInfarction | 0.016443 | 0.016443 | 0 |
| FG_MI | 0.017258 | 0.016192 | 0 |
| FG_CAD | 0.024381 | 0.022152 | 0 |
| FG_Stroke | 0.009292 | 0.009292 | 0 |

The complete compatibility table retains raw intercepts, diagonal values, source-object identities and endpoint accessions. The estimates range from about 0.009 to 0.039. These values alone leave participant overlap unresolved. The source reports sampling errors for genetic covariances but supplies no standard errors for the intercepts. HapMap3 selection and chi-square filters also differ from the full LAVA variant set. The local sensitivity analysis treated the two sets of normalised intercepts as fixed error settings.

We retained the original qualification records for four MVP endpoint families, two scales and 2,495 regions (19,960 records). Changing the error correlation leaves each trait's univariate parameters and eligibility unchanged when the diagonal entries remain fixed. The error-correlation comparison included all 166 family–region combinations eligible on either discovery scale, covering 151 physical regions. Both scale conventions, LD input, allele processing and regional dimensions were retained. The two nonzero settings used LAVA 0.1.5 and the same seed within each region. Regions outside the original univariate qualification were not newly fitted.

For each error setting, we reapplied the original discovery thresholds, formed the union across discovery scales and used its size for replication correction. Final selection required both discovery scales and both FinnGen scales to pass, with the original direction, dimension and boundary rules. Separate fixed-case comparisons retained every original selected association across three settings, two sources and two scales (144 rows), including estimates that no longer passed discovery. Supplementary Table S10B summarises selection, and S10C reports the original 12 cases.

After inspecting the completed fits, we also held the original replication-family sizes fixed at four for MI and 36 for coronary atherosclerosis. This post-fit diagnostic reused the computed P values and all other original gates, separating error-model changes from changes in the replication threshold (Supplementary Table S10D). Under the original procedure, each nonzero setting retained 11 associations. Holding the original correction fixed retained nine with Default and ten with Chi80. All original cases retained positive point estimates and pointwise interval lower limits; selection-sensitive cases remain in the comparison tables. These intervals condition on the regional model and error setting and do not adjust for selection. P values near a selection threshold can also reflect the simulation precision of LAVA.

Selection summaries, all final selection gates, original-case estimates and the fixed-correction diagnostic accompany Supplementary Table S10. The error-setting index distinguishes applied settings from the zero-error field in the compatibility file. The MVP survey-stroke family had one eligible discovery region and no discovery; cerebral infarction had none. Their sparse local coverage limits source comparisons for these endpoints.

#### MR-Egger intercept diagnostics

Fourteen intercept diagnostics used the complete gSAPA harmonised records passing the stated allele-quality exclusions, without repeating instrument selection. TwoSampleMR 0.6.15 reoriented each allele to positive exposure effect, fitted outcome effect on exposure effect with inverse-outcome-variance weights, and used multiplicative residual dispersion with an underdispersion floor of one. Intervals and tests used $n-2$ t degrees of freedom. Supplementary Table S9 reports nominal P, seven-endpoint Holm-adjusted P within each rule and Holm-adjusted P across all 14 tests. Under the within-rule families, the MVP cerebral-infarction intercept was positive in both sets; only Broad retained support across all 14. The instrument sets share variants. Intercept tests assess directional pleiotropy under the MR-Egger assumptions, including InSIDE.[11]

Supplementary Note 5 | Anatomical associations and matched prediction comparisons

The linked 1,876-person NHANES analysis retained 2015–2016 and 2017–2018 participants aged 20–59 with the qualified eight measurements and DXA regions. The primary cross-sectional models conditioned on total fat and its square, age and age squared, sex, race/ethnicity, education and cycle. All five depot readouts are shown. Scans and blood data are linked within person and cycle; exact paired timestamps are unavailable. Removing the BMI score term while including BMI separately is a sensitivity to score composition.

MRI abdominal SAT (GCST90590314), thigh SAT (GCST90590315), VAT (GCST90590316) and liver fat (GCST90590317) retain their original trait units and harmonized variant handling. Existing input audits give N=37,589, 37,584, 37,589 and 30,085, respectively. The catalogue maximum of 37,589 does not replace the lower thigh or liver input N. BIA whole-body fat (GCST90691779) has study-level N=412,524; the file does not provide a per-variant N. These sizes describe different, overlapping UK Biobank sources. Conditional genetic correlations use the completed same-scale covariance system and BIA whole-body fat; the studies share UK Biobank participants and originate from distinct studies. The whole-leg DXA and thigh-SAT MRI labels are preserved. Supplementary Table S6 presents the anatomical, matched-construct and imaging-free comparisons under their respective conditioning sets.

Liver-fat–disease covariance along the common direction

We used the joint 17-trait LD score regression system containing the eight routine traits, four vascular endpoints, four MRI traits and whole-body fat. LD scores and regression weights used the European-ancestry reference, with 232 common blocks and the full sampling covariance of the 153 unique covariance entries. The primary SNP chi-square cap followed the default max(80, 0.001 × maximum sample size); the sensitivity specification fixed it at 80. MRI liver fat (GCST90590317) had 30,085 participants. The MVP MI, MVP CircStrk survey-stroke, FinnGen I9_MI_STRICT and FinnGen I9_STR endpoints retained their distinct definitions from Supplementary Note 4. The component analysis uses genetic covariances without conditioning on whole-body fat. The correlations in Fig. 3b are conditional on whole-body fat.

Let X be the eight genetic components standardised to unit genetic variance, G their correlation matrix, and w the fixed genetic direction. For this analysis, $T = w'X / \sqrt{w'Gw}$. For genetically standardised liver fat L and disease Y, let $cL = Cov(X,L)$, $cY = Cov(X,Y)$ and $rLY = Cov(L,Y)$. The covariance represented by T is $C = Cov(L,T)Cov(T,Y)$. The covariance represented by the full panel is $H = cL'G^{-1}cY$. We report the additional component $K = H - C$ and the residual $U\_X = rLY - H$, giving $rLY = C + K + U\_X$. These signed components share the original L and Y genetic-SD scales. They are additive covariances, not partial correlations, proportions of risk or causal pathways. Negative components can offset positive components, so C/rLY is not an information-retention percentage.

A joint delta method propagated the full sampling covariance through genetic standardisation and all component calculations. Pointwise 95% normal intervals accompany Holm-adjusted P values for the 12 component tests (four endpoints × C, K and U_X), separately within each specification. Total genetic correlations are descriptive and outside these families. Two thousand joint Gaussian summary-error draws assessed nonlinearity and whether the projected quantities stayed within their mathematical domains. The full 17-trait covariance estimate had a small negative eigenvalue and was not smoothed. The matrices used for these endpoint projections were valid, and the joint sampling covariance was positive definite. The interpretation is specific to these projections, which use overlapping UK Biobank participants and source-specific GWAS measurements.

Same-panel partial information for visceral fat

Within the same 1,876 participants, weighted regressions residualized the eight NHANES-standardised measurements, their frozen clinical score Q and visceral-fat mass against an intercept, age and age squared, sex, race/ethnicity, education, cycle, total fat and total fat squared. With residual eight-marker covariance Gx, marker–VAT covariance cx and residual VAT variance vy, the full-panel partial $R^2$ was $cx'Gx^{-1}cx/vy$. The SAPA partial $R^2$ was the squared weighted correlation of residual Q with residual VAT. The reported fraction divided the SAPA quantity by the full-panel quantity. These are fitted, same-sample linear summaries; the held-out prediction comparisons below have different estimands.

All residualization, covariance and ratio calculations were repeated in the 60 stratified delete-one-PSU jackknife replicates. Replicate weights removed one PSU and doubled the other within the relevant stratum. The variance was one half the sum of squared replicate departures from the point estimate; 95% intervals used a t distribution with 30 design degrees of freedom and were not clipped. Supplementary Table S6 reports the numerator, denominator and their directly propagated ratio. These clinical partial $R^2$ quantities are separate from the genetic information fractions in Supplementary Note 3.

Imaging-free anatomical prediction

The main support was the original anatomy sample with positive measured waist and an absent or non-1 waist-quality flag (BMIWAIST): 1,869 participants, comprising 1,060 and 809 by cycle. Seven original participants lacked eligible waist values; six also had the flagged code, so these were not counted as separate exclusions. We retained measured VAT and whole-leg mass in kg as outcomes, while excluding DXA total fat, other depots and all imaging-derived quantities from the predictors.

The main common background contained the population terms, restricted cubic splines for BMI (knots 20, 25, 30 and 40 kg/m²) and waist (70, 90, 110 and 130 cm), sex interactions with their linear terms, and a linear BMI-by-waist interaction. Each spline had three degrees of freedom. Fixed centres and scales were (BMI−25)/5 and (waist−90)/10; knots were not chosen from outcome fits or data quantiles. One prespecified diagnostic removed the four nonlinear spline columns but retained the interactions. It was not used to select the primary result.

Five unpenalized weighted regressions were compared: background alone; background plus SAPA; background plus log triglycerides, HDL and HbA1c as three separate NHANES-standardised terms; background plus both; and background plus Full8. With BMI already in the background, its fixed score contribution was removed algebraically; in the combined model, the fixed contributions of TG, HDL and HbA1c were also removed as an algebraically equivalent parameterization. Predictions were checked against those from the corresponding full score. Full8 contained seven additional free terms because BMI was already present. These reparameterizations do not define new scores.

The two cycles exchanged training and test roles. Each participant contributed one held-out prediction, with survey-weighted errors pooled across cycles. MSE differences in kg² were the primary estimands. We also reported RMSE, MAE, their paired differences and relative MSE improvement. Four prespecified primary contrasts comprised SAPA versus background and adding SAPA to the three-term metabolic comparator, for each of two outcomes; two-sided P values were adjusted together by Holm's method. Other paired comparisons and the simplified-background analysis were descriptive. There was no equivalence margin. Negative predictions were not clipped. Calibration intercepts and slopes were estimated separately within each test cycle for diagnosis only and were not used to alter predictions.

The existing 1,465-person MetS-eligible subset supplied a limited sensitivity comparison using the fixed 2014 Gurka waist-based equations. The common background allowed their six sex-by-race/ethnicity waist slopes and required group intercepts, so an apparent score increment could not merely exploit those predefined waist relationships. No equations were extrapolated to other groups. Original total-fat-conditioned MetS, waist and clinical PhenoAge comparisons remain separate from this no-imaging-input analysis.

Vascular contrasts and matched construct comparisons

The four VAT-minus-thigh-SAT vascular contrasts in Fig. 3c use partial genetic correlations conditional on whole-body fat. Delta-method contrast errors include covariance between the two correlation estimates. Normal-approximation intervals are pointwise; Holm adjustment retains the complete 20-comparison family. MRI, BIA and vascular GWAS identities remain source-specific, with no pooling of source-specific P values.

Fig. 3e and Extended Data Figure 4 retain the original matched construct comparisons. The continuous MetS equations retain their sex- and race/ethnicity-specific eligibility (n = 1,465). The clinical PhenoAge comparison uses the corresponding complete population of 1,867, with the clinical score developed before the DNA-methylation surrogate and the corrected clinical formula.[17,18,19] Both exchange the 2015–2016 and 2017–2018 cycles for training and testing. Total-fat-conditioned models share age and age squared, sex, race/ethnicity, education and total fat and its square; direct BMI-conditioned additions have a separate baseline denominator. Sixty survey jackknife replicates refit both training directions, with pointwise t intervals with 30 degrees of freedom. Prespecified MetS-minus-SAPA, SAPA-beyond-waist and SAPA-minus-PhenoAge comparisons keep their original separate Holm families. The no-imaging task is specified in this section and uses a different 1,869-person population. These comparisons evaluate anatomical outcome-model transfer with the externally frozen scoring rules; the NHANES reference contains both NHANES cycles.

Clinical PhenoAge implementation

The saved implementation uses age and nine blood measurements: albumin, creatinine, glucose, CRP, lymphocyte percentage, mean corpuscular volume, red-cell distribution width, alkaline phosphatase and white-cell count.[17,18] Ordinary serum glucose is used here. The frozen coefficient and unit table records the NHANES fields and conversions: albumin from g/dL to g/L (×10), creatinine from mg/dL to μmol/L (×88.4), glucose from mg/dL to mmol/L (×0.05551), and CRP from mg/L to mg/dL (×0.1) before its natural logarithm. Other inputs retain the units listed in that table.

Let $xb = -19.907 + \Sigma\, c_j x_j$, using those transformed inputs and coefficients. The saved calculation implements the published correction[19] through $H = 1.51714 \times \exp(xb) / 0.0076927$ and $M = 1 - \exp(-H)$. Clinical PhenoAge is $141.50 + \ln[-0.00553 \times \ln(1 - M)] / 0.09165$. For numerical stability, the implementation evaluates the algebraically equivalent expression 141.50 + {ln[0.00553 × 1.51714 / 0.0076927] + xb} / 0.09165. This is the clinical blood-based score; DNA methylation is not an input.

Longitudinal waist and marker changes

Fig. 3d displays the BMI-conditioned within-person estimates for the BMI-omitted projection in the two ELSA windows and both CHARLS waist ranges. Supplementary Figure 2 and Supplementary Table S7 give the complete comparisons.

This complementary analysis asked whether change in waist accompanied change in the same fixed clinical profile within individuals. It used observed measurements and the frozen D transformation, means, SDs and six-decimal loadings. For marker j at visit t, $z_{ijt}=(T_j(x_{ijt})-\mu_{j,D})/\sigma_{j,D}$ and $Q_{it}=\Sigma_j\lambda_j z_{ijt}$. Q without BMI removed λBMI zBMI while retaining all other coefficients; neither coordinate was rescaled within a cohort or wave. The blood pressure contribution was λSBP zSBP+λDBP zDBP; the other-five contribution contained LDL, HDL, log TG, HbA1c and log CRP. Their sum equals Q without BMI. HDL retained its negative scoring coefficient and original measurement direction.

The primary ELSA window linked nurse visits at waves 4 and 6 in 2,959 participants from 2,517 households. The median interval was 3.917 years. Eligibility required the saved complete observed eight-marker panel, positive valid waist measurements at both visits with the source measurement-status rule, BMI, ordered visits and complete model background fields. The earlier waves 2 and 4 included 2,246 participants. Among 1,416 people with all three visits, both windows were fitted using the same wave-6 blood weights and wave-4 household identities. The windows overlap at wave 4 and belong to one cohort.

CHARLS waves 1 and 3 supplied 5,404 participants with linked valid positive waist measurements and complete model fields. Before fitting, the principal comparison was restricted to both waist values between 40 and 200 cm, giving 5,207 participants from 3,901 households. The threshold was an investigator-defined broad review range. All 197 excluded participants had at least one waist below 40 cm; none exceeded 200 cm. The full positive-value sample was analysed separately, without converting presumed units or selecting another threshold. CHARLS blood files did not supply individual collection dates. Interview dates provided an approximate interval, with median four years. ELSA timing used nurse-visit months for body and blood measurements. The saved ELSA LDL values were calculated, whereas CHARLS LDL was directly measured.

For each marker, the two regressions were:

mean(z) = αB + βB mean(waist)/10 + γB C + εB

change(z) = αW + βW change(waist)/10 + γW C + εW

C contained baseline age, male sex, smoking and the observed visit interval. Age was centred at 65 and divided by 10; interval was centred at four years. Smoking status follows the baseline definition for each paired sample. In BMI-conditioned models, mean BMI entered the first equation and BMI change entered the second. The change intercept absorbed the common visit shift; background terms allowed differences in mean change by background characteristics. The estimands were the difference between people in their two-visit mean profile and the association of same-person marker changes with waist changes. For two balanced visits these equations are equivalent to a stacked mean/deviation model with background-by-visit terms. The change model estimates change over the observed visit interval and adjusts for its duration.

Scalar coefficients were obtained by applying the same frozen weights to the eight marker coefficients. In the BMI-conditioned regression, the BMI outcome is also a predictor, so its partial waist coefficient is zero by construction. This gives identical conditional waist coefficients for Q and Q without BMI. All units in Supplementary Table S7 are fixed Q units per 10 cm, or fixed NHANES-reference marker SD per 10 cm for constituent markers. They differ from the task-specific score-SD units of the disease analyses.

Weights were inherited from the follow-up blood-sampling frames: ELSA wave 6 w6bldwt for waves 4 to 6, wave 4 w4bldwt for waves 2 to 4, and CHARLS wave 3 r3wtrespbiob. The three-visit comparison used wave-6 weights in both windows. Weights were normalized within each model, which leaves coefficients unchanged. They were not newly estimated weights for longitudinal selection. Uncertainty used 999 household-level multinomial bootstrap resamples with seed 4852026. Related models and subsets shared household draws so that within-minus-between, medication-adjustment and paired-window contrasts retained joint error. Intervals are the 2.5th and 97.5th percentiles for each estimate separately. The bootstrap preserves household clustering but does not reproduce the full original complex survey design. No simultaneous coverage is claimed.

Prespecified checks retained participants with complete medication status at both visits, added the means or changes in antihypertensive, lipid-lowering and diabetes medication indicators, restricted to unchanged medication states, and restricted ELSA to its fasting-eligible subset at both visits. The CHARLS coding subset required qc1_va003=1 at wave 1 and bl_fasting=1 at wave 3; the unverified meaning of the wave-1 code prevents identifying this as a confirmed fasting subset. ELSA also used the directly measured-weight subset. Wave-2 ELSA lipid-medication status was unavailable and was not coded as untreated. CHARLS additionally excluded records flagged for triglyceride top-coding at wave 3. All marker inputs remained observed; medication covariates did not alter measured values. Supplementary Table S7 gives all sample counts and estimates, including the unadjusted comparison in the medication-complete sample.

The ELSA main estimates were also evaluated by deleting one household at a time without changing the eligibility rules. In CHARLS, the 197 range-flagged records accounted for 3.139% of sampling weight and 73.940% of weighted residual waist-change sum of squares after the conditional model's other predictors were removed. These records had a large influence on the estimated waist-change association. Their true values and units could not be verified. The full-sample check includes all positive values.

For a supplementary direction diagnostic, squared cosine was $(\beta'\lambda)^2/[(\beta'\beta)(\lambda'\lambda)]$ on the frozen D scale. It measures alignment of an eight-marker association vector with the fixed weights. It is not a fraction of disease risk, genetic information or individual change variance explained. No disease model, age interaction or new set of score weights was fitted in this unit. Aggregate estimates, designs, weights, paired contrasts and checks accompany Supplementary Table S7 in Source Data.

### Supplementary Note 6 | Biochemical correspondence: fasting insulin and IL-1RA

#### Fasting insulin, regional fat distribution and the six-marker projection

The NHANES 2001–2002 and 2003–2004 fasting insulin analyses included participants aged 20–79 years with complete observed construction markers, background covariates and positive fasting-subsample weights. DXA measurements required the no-imputation quality flag and identical values across all five supplied imputation files. No imputed DXA values entered these models. Eligibility required DIQ010 responses of no or borderline diabetes and no reported glucose-lowering treatment. All 2,475 included participants explicitly denied insulin use (DIQ050); oral-treatment responses (DIQ070) were structurally missing under the questionnaire skip pattern. They were not recoded as explicit denials. The two cycles contributed 1,327 and 1,148 participants. These samples are separate survey cycles, not external validation cohorts. Selection was material: weighted mean BMI was 27.009 in the included sample and 32.702 among 506 otherwise eligible participants whose DXA was imputed or not verified as measured (Supplementary Table S8F).

Insulin and fasting glucose were obtained from L10AM_B and L10AM_C. The 2003–2004 Tosoh insulin measurements were converted to the Pharmacia-equivalent scale using the CDC equation, 0.96006 × Tosoh + 3.23663. Insulin entered models on the log2 scale. The native-assay sensitivity retained the released values in each cycle. Because the calibration includes an intercept, doubling on the converted scale differs from doubling on the native scale. Uncertainty in the published conversion parameters was not available for propagation. Fasting weights correspond to the source examination and fasting eligibility rules. The combined analysis used the two-year weights divided by two.

All eight markers used the fixed NHANES 2015–2018 reference transformations, means, standard deviations and loadings. The six-marker projection Q6 was the sum of the original weighted SBP, DBP, LDL, HDL, log TG and HbA1c terms after removing BMI and log CRP. It was not refitted or rescaled; coefficients are in the original projection units, not per SD of Q6. This scale differs from the separately standardised six-marker HRS prediction model. Background adjustment comprised centred age in decades, age squared, sex, race/ethnicity, smoking, height and survey cycle. Body composition terms were log total fat mass, log trunk-to-whole-leg fat ratio and log non-bone lean mass.

Model A related each reference-standardised marker and Q6 to log2 insulin plus background characteristics. Model B added the three body composition terms. Model C added log CRP and evaluated the six retained markers and Q6. Model D related log2 insulin to body composition and background characteristics. Three pooled comparisons were specified before estimation: insulin–Q6 and regional-fat-ratio–Q6 associations in model C, and insulin–CRP in model B. They formed one Holm family. Ratio effects were converted to a 10% higher trunk-to-whole-leg fat ratio. For CRP, the standardised log-concentration coefficient was first multiplied by its reference SD (1.23541639029462), then exponentiated to obtain a percentage difference. The insulin–fat-ratio and CRP–Q6 comparisons were supporting analyses, with pointwise intervals (Supplementary Table S8G).

Uncertainty used 999 Rao–Wu resamples of primary sampling units within survey strata. Within each stratum containing m units, m−1 were sampled with replacement and replicate weights multiplied by m/(m−1). Resampling used the full positive-fasting-weight population before restricting to the analytic domain. Coefficient intervals used replicate standard errors and t quantiles with 30 pooled or 15 cycle-specific degrees of freedom. Directional similarity was the cosine between the insulin-associated marker coefficients and the corresponding fixed SAPA loadings; its intervals used bootstrap percentiles. A joint proportionality test evaluated whether the coefficient vector was a scalar multiple of the fixed loadings. The cosine and squared projection quantify alignment of the insulin-associated marker effects with the fixed loadings. The two specified sensitivities retained native assay values and excluded fasting glucose ≥126 mg/dL or HbA1c ≥6.5%. All resample fits completed. The coefficients describe cross-sectional associations conditional on the stated covariates.

### IL-1RA selection, prediction and age comparisons

IL-1RA was selected after the earlier same-resource six-marker screen. The current family-fold analysis evaluates the selected marker model; neither cross-validation nor resampling repeats biomarker selection. Its precision is therefore conditional on that selection.

The IL-1RA measurement is PIL1RA (pil1ra in the imported Stata file), from VBS 2016 Additional Assays, version 2, released in April 2025 as vbs16aa.dta.[20] The accompanying data description identifies serum measurement by a Simple Plex Human Interleukin 1 Receptor Antagonist assay on the ELLA system (ProteinSimple). This supplement is linked by HHID and PN to the main HRS 2016 venous blood release, version 3 (vbs16v3.dta), the 2016 core examination and the dried-blood-spot HbA1c measurement, PA1C_ADJ. Lipids and CRP come from venous blood; HbA1c comes from dried blood spots. Thus the panel combines linked 2016 measurements with a recorded visit-to-blood interval. File, field and manual identities are listed in the module provenance table.

The NHANES-reference analysis uses 3,464 people in 2,680 HRS families and the original five family folds. SAPA, PC1 and adverse-direction equal weights are estimated or defined in the external NHANES reference; each is standardized there using its own weighted mean and SD. PC1 is the leading eigenvector of the NHANES-standardised weighted covariance. HDL has the opposite orientation to the other equal-weight inputs. Their transformations and score units remain fixed across test folds.

The shared background includes age-by-sex terms, smoking, the measurement interval, fasting and collection-time band. Each score uses a three-degree-of-freedom natural spline and sex interaction. Interior score knots use the training one-third and two-thirds quantiles, with training extremes as boundaries. The original age-mapping comparison adds the prespecified age interaction. Whole-model held-out $R^2$, paired MSE differences and fixed one-reference-SD contrasts are separate estimands.

The 200 family resamples keep the original family folds, repeat training knots and coefficients, and jointly support the saved basic-bootstrap intervals. Two-sided normal-approximation P values use the bootstrap SE. The five-comparison performance family comprises the three score age-interaction increments and the two direct M0 score comparisons. The separate three-comparison age family retains its original definition. Individual basic intervals are not simultaneous intervals. S8 reproduces all six biomarkers of the initial screening step at its reported precision, including the component-controlled analysis. Reusing the selection resource is stated in the manuscript.

### Composition information beyond BMI and CRP

The composition comparison used the same 3,464 participants, 2,680 families, five folds and log IL-1RA outcome. We specified these models after examining the whole-score results and six-marker molecular screen, and before fitting the component comparison. Each participant contributed equally. Resampling families accounted for dependence within households.

For each frozen eight-marker direction, we removed the BMI and log CRP terms. The remaining terms were SBP, DBP, LDL, HDL, log TG and HbA1c. Their weight ratios were preserved. Projection means and SDs used the original NHANES reference and its weights, without using HRS outcomes. The six-marker PC1 projection therefore retains weights from the eight-marker PC1; it is not a newly fitted six-marker principal component. The weights and units identify these scores.

The background retained age spline by sex, smoking, visit-to-blood interval, fasting and collection-time terms. The age spline used fixed interior knots at 60 and 75 years and boundary knots at 40 and 100. The component background added BMI and log CRP, each with a three-df natural spline and linear sex interaction. Separate extensions added one of the three six-marker projections using the same four-coefficient form. A fuller comparator entered the six markers individually, each with that form, adding 24 coefficients. The latter is an additive model with more parameters; it is not generally nested with a nonlinear projection model. All non-age spline knots were recomputed from each training sample's one-third and two-thirds quantiles, with training extremes as boundary knots.

All models predicted the same held-out participants. Whole-model $R^2$ was 1 minus MSE divided by log IL-1RA variance in the evaluation sample. The variance denominator was common across models and was not used for training. $R^2$ increments describe prediction improvement under these fitted models, not a causal or mediating contribution of the remaining markers.

Two hundred whole-family bootstrap resamples retained the original family folds and repeated training knots and all model coefficients. Seven paired MSE comparisons formed one Holm family: each of the four extensions against the BMI/CRP background, SAPA versus each alternative projection, and the individual-marker model versus SAPA. Positive improvement favours the named extended or first model. Basic-bootstrap 95% intervals are pointwise; P values use a two-sided normal approximation with the bootstrap standard error. $R^2$-difference intervals use the same paired resamples and a common denominator within each resample, without a second testing family. A repeat was eligible only if all six models were finite, full rank and warning-free. All 200 qualified; the predefined minimum was 180. Supplementary Table S8E reports all models and contrasts.

Within-HRS transfer across age

Supplementary Figure 3 and Supplementary Table S8 report the age-transfer comparisons.

The source has 1,757 people aged 55–69; the target has 1,287 aged 70–84, with 2,417 unique families and 102 families spanning age groups. Family folds prevent every target test family's members from entering either source or target training. The background has the saved age, sex, interaction, smoking, interval, fasting and collection-time columns. Source score splines use source-training quantiles and are centred at Q=0.

T0 applies the younger relation directly. T1 re-estimates target-training background terms with the fixed source score contribution as an offset. T2 additionally estimates one multiplier on that contribution. Two hundred family resamples repeat both stages; family members at both ages are sampled jointly. The primary improvement MAE(T1)−MAE(T2) uses a basic-bootstrap interval. T0−T1 and the two alternative-score comparisons use the retained three-comparison Bonferroni normal intervals. Mean error, geometric-mean calibration, MSE and $R^2$ remain diagnostics in the completed source output. No equivalence margin was defined.

Age structure in the IL-1RA source population

Supplementary Table S8D presents the structural comparison in the HRS IL-1RA sample. Background regression used all 3,464 participants; residual covariances were then estimated in 3,044 people aged 55–84 across six five-year age bands, with sexes equally weighted. The frozen SAPA, PC1 and adverse-direction equal-weight directions define P, O and F as in Supplementary Note 7. The fourth quantity is the off-diagonal contribution $u'Ru-\Sigma u_j^2$ of the residual correlation matrix R. Linear trends across the six band midpoints are expressed per decade. Two hundred paired family resamples gave approximate simultaneous intervals from the maximum standardized error across 12 score-by-quantity trends. Normal-approximation two-sided P values use bootstrap standard errors and Holm adjustment across the same 12 trends. The table reports this complete family, with F in percentage points; individual basic intervals are available in the aggregate source file. These summaries concern the selected IL-1RA sample and are cross-sectional age comparisons.

Supplementary Note 7 | Age-related marker structure

NHANES recent (2015–2018, n = 4,000) and earlier (1999–2010, n = 11,053) samples retain complete observed panels, valid stroke questionnaires and positive fasting weights at ages 20–79. These are not disease-free incident-risk sets. The age grid has 51 points from 25 to 75 years with a five-year kernel. Within cycle and sex, the original five-column mean-model basis is fitted with the saved survey weights; weighted, centred residual covariance matrices are combined with equal sex weights and then equal cycle weights.

On the fixed D metric, $P=u'C(a)u$, $O=tr(C(a))-P$ and $F=P/(P+O)$. The best-direction proportion $M=\lambda max(C(a))/tr(C(a))$ and M−F were retained as diagnostics. Each of the two periods used all 500 original survey resamples and re-estimated the required mean and covariance quantities. Linear trends per decade summarize the whole grid. The simultaneous summary intervals use the saved maximum standardized errors across two periods, nine quantities and two summaries (36 entries); the nonlinear template was previously observed and then frozen. These intervals condition on the reference and loadings.

The algebraic decomposition separates $\Sigma_j u_j^2 C_{jj}$ from $2\Sigma_{j<k} u_j u_k C_{jk}$. B is the latter additive cross-marker quantity on the fixed-reference covariance scale; it is distinct from the variance share F and its components are not a partition of F. It establishes a contribution of cross-marker covariance to the age pattern. It does not identify a personal feedback network or a within-person ageing rate. Fixed-direction proportions have a panel-variance denominator, unlike disease-information retention.

#### Fixed versus age-adapted directions on the same reference metric

Allowing a different direction at every age separates the concentration of marker variation from the suitability of the fixed SAPA weights. M is the largest achievable share for a single direction on the same NHANES reference metric; M−F is the additional share available by changing direction. The following saved linear trends use all 51 age-grid points and the original 36-summary simultaneous-error family.

| NHANES period | Quantity | Change per decade, percentage points | Simultaneous 95% interval |
|---|---|---|---|
| 2015–2018 | Fixed SAPA share F | −2.907 | −3.856 to −1.957 |
| 2015–2018 | Best-direction share M | −3.604 | −4.936 to −2.272 |
| 2015–2018 | Gap M−F | −0.697 | −1.817 to 0.423 |
| 1999–2010 | Fixed SAPA share F | −3.109 | −3.811 to −2.407 |
| 1999–2010 | Best-direction share M | −2.953 | −3.677 to −2.229 |
| 1999–2010 | Gap M−F | 0.156 | −0.746 to 1.058 |

Both F and M declined. The simultaneous intervals for the linear trend in M−F included zero in both periods. This test measures the age trend in the gap between variance shares. The complete summaries retain all 36 estimates, pointwise and simultaneous intervals, and the frozen nonlinear-template summary. The age curves retain the full grid. Extended Data Figure 9 describes the eigenvalues of within-population correlation matrices.

#### Within-population correlation spectra

Extended Data Figure 9 presents weighted correlation spectra across three cohorts, disease-free and medication-minimal subsets, and 751 ELSA participants with paired measurements. PC1 share and entropy rank use within-population correlation matrices. Intervals are 500-resample percentiles; raw and rank-normal transformations and retention weighting are compared. These quantities differ from the fixed reference-scale SAPA variation in Supplementary Note 7.

### Supplementary Note 8 | Body composition and age-related clustering

#### Population, measurements and scoring

The NHANES 2001 to 2002 and 2003 to 2004 whole-body DXA releases were linked by participant identifier to the existing observed-marker records. Among 3,443 eligible marker-panel candidates aged 20 to 79, 187 pregnant participants and 14 participants with other unusable records were excluded. The final population comprised 1,161 aged 20 to 39, 1,115 aged 40 to 59 and 966 aged 60 to 79. The two official releases each contained five imputations for the same participants. No high-variability supplementary-file candidate occurred in this analysis domain.

Body measures were total fat mass, trunk fat divided by the sum of left and right leg fat, and total non-bone lean mass. Masses were expressed in kilograms before natural logarithmic transformation. Published values already incorporated CDC's mass correction and were not corrected again. Early DXA trunk fat and non-bone lean mass retain these measurement definitions; they are distinct from VAT and directly measured skeletal muscle. The observed eight-marker inputs used the same D transformations, constants and six-decimal loadings as Supplementary Note 1. SAPA remained in its fixed projection units without a new sample-SD normalization.

Two-year fasting-subsample weights, WTSAF2YR, were divided by two when combining the cycles. Strata and primary sampling units retained cycle-specific identities. We used survey weights within each domain. The main summaries standardise to equal weights for the sexes and, for covariance matrices, for the cycles. The official CDC DXA page, technical documentation and weighting tutorial are cited in the main Methods.

#### Conditional mean contrasts

Separate female and male weighted models were fitted for each of the eight transformed NHANES-standardised markers. The design contained an intercept, an age natural spline with internal knots at 40 and 60 and boundaries at 20 and 79, survey cycle, height, the three log body measures, and each body measure's interaction with (age−50)/10. All three body measures were included together. Primary contrasts represented a 10% higher value of one measure, with the other two fixed, evaluated at ages 40 and 70. Effects and direct age differences were averaged equally across sexes.

The fixed loading vector transformed constituent-marker effects into SAPA effects. A separate linear contrast removed only the BMI contribution and did not retrain or renormalize the remaining coefficients. All eight constituent effects, including HDL in its original measurement direction, are retained. Sex-specific and cycle-specific estimates are available in the aggregate source table. The contrasts describe the fitted local conditional relationships; they do not define an intervention on body composition or longitudinal change within one person.

Three primary SAPA age differences were compared jointly with a Wald statistic based on the Rubin total covariance and the finite-reference approximation specified below. Each individual age difference also had a small-sample interval and t test, with Holm correction across the three tests. The multivariate and univariate degrees of freedom are calculated for their respective targets. A post-estimation contrast adding the total-fat and lean-mass differences was saved to explain the joint result. It is labelled as an explanatory re-expression, not an additional prespecified primary test.

#### Exact descriptive covariance decomposition

The structural comparison used ages 40 to 59 and 60 to 79, separately within sex and cycle, giving 2,081 participants overall. In each stratum, identical weighted regressions removed the intercept, linear within-group age and height from the eight-marker vector X and body measurement vector Z. The same weights and covariance denominator were used throughout. An unpenalized weighted multivariate projection gave

$$C_g = M_g S_g M_g' + R_g,$$

where C is the background-adjusted marker covariance, S is body measurement covariance, M maps body measures to markers, and R is the residual covariance. The implementation verified both projection orthogonality and the full matrix identity. The unadjusted marker-covariance difference was also retained; its change after age/height adjustment was recorded separately and was not assigned to a body composition component.

Let $S_{bar}=(S_{old}+S_{mid})/2$. Within each sex and cycle, the body-related covariance difference was partitioned as

$$\text{Relationship} = M_{old} S_{bar} M_{old}' - M_{mid} S_{bar} M_{mid}'$$

$$\text{Distribution} = [M_{old} (S_{old}-S_{mid}) M_{old}' + M_{mid} (S_{old}-S_{mid}) M_{mid}']/2.$$

Their sum equals the older-minus-middle-age difference in fitted covariance. Adding $R_{old}-R_{mid}$ recovers the complete background-adjusted difference. Corresponding matrices were averaged with equal sex and cycle weights before the summary quantities were calculated.

For the unit-normalized frozen loading vector u, the cross-marker contribution was

$$B_{cov}(C) = u'Cu - \Sigma_j u_j^2 C_{jj} = 2\Sigma_{j<k} u_j u_k C_{jk}.$$

Fig. 6 shortens $B_{cov}$ to B. It is linear in the covariance matrix, so the relationship, distribution and remaining terms add exactly on this scale. It excludes each marker's own variance. Its fixed-reference covariance units differ from the same-panel genetic information quantities in Supplementary Note 3. The accompanying summaries give aligned variance P, remaining variance O and their corresponding panel-variance share F. We calculated F from the averaged matrix.

This decomposition separates second-moment distribution and projection relationships. Signed terms were retained, including contributions with opposite signs. No causal mediation fraction, full-distribution standardization or explanation-percentage success threshold was imposed. Removing BMI meant subtracting its fixed cross-marker contributions while keeping all other u coefficients unchanged.

Measured subset and routine body measurement comparison

CDC's imputation models used sex and age groups, body measures and several routine measurements including blood pressure and HDL. Before association estimation, a measured-subset definition required zero relevant imputation flags and identical required body values across the five released versions. It yielded 2,668 participants overall and 1,676 in the age-group comparison (915 aged 40 to 59 and 761 aged 60 to 79).

In the 2003 to 2004 release, 80 participants across the released age range had zero overall and regional imputation flags but varying body values. All 80 had completed scans and at least one regional soft-tissue quality code of 4, defined as arm/leg overlap in the official codebook (DXX_C, documentation pages 12–13). Among participants with zero imputation flags, the regions with this quality code matched exactly the regions with varying fat and non-bone lean values: left arm, 61; right arm, 70; left leg, 60; and right leg, 69. Head, trunk, bone and area values were invariant. Regional fat and lean sums reproduced the corresponding total values within release rounding. This pattern localizes the discrepancy to overlap-flagged soft tissue and derived totals. We found no explanation for the retained zero imputation indicators in the public documentation reviewed. We did not infer that an entire participant record was invalid or relabel these as officially confirmed imputations. Independent R and Python reads recovered the same 80 identifiers, and the saved model inputs matched the raw files exactly. Twenty-three participants entered the 20–79-year mean-relation analysis (6 aged 20–39, 10 aged 40–59 and 7 aged 60–79); 17 entered the 40–79-year structural comparison. All five official versions remained in the primary analysis. Excluding the 23 from the measured subset was already part of the frozen invariance rule; adding a requirement for valid regional soft-tissue quality codes selected exactly the same 2,668/1,676 participants. No estimates or sample memberships changed. The 1999–2000 and 2001–2002 releases contained no flag-zero discrepancies for the required measurements. Aggregate cycle, region and analytic-sample checks accompany Source Data. Selection into the measured subset remains relevant when comparing its estimates with the primary analysis.

The same 2,057 waist-complete participants supplied a paired comparison between the three DXA variables and log BMI/log waist. Background adjustment, projection, weighting and uncertainty were identical. The main comparison was the DXA-minus-BMI/waist difference in the fitted B age contrast; remaining and internal component differences were also retained. This is a comparison of shared-variation descriptions and has no predefined equivalence margin or clinical-outcome threshold.

Imputation pooling and uncertainty

Each of the five released versions was analysed separately. Using the fixed seed 4142026, 399 survey-design bootstrap replicates were generated from the full positive-weight design universe, preserving strata and cycle identities; the analysis domain was applied subsequently. All imputations shared the same PSU draws and the survey procedure's scaling factors.

Within-imputation sampling variances and covariances were combined with between-imputation variation using Rubin rules. Pointwise 95% intervals used Barnard-Rubin degrees of freedom, with complete-design degrees of freedom of 30 for both cycles and 15 for one cycle. The three-dimensional comparison uses the finite-degree-of-freedom reference below. Figures show pointwise intervals. We compared the magnitudes and precision of estimates across cycles and measured-scan subsets.

The aggregate source files contain complete constituent-marker contrasts, sex/cycle summaries, observed/background-adjusted covariance differences, imputation variance components, direct paired comparisons and the explicitly labelled post-estimation re-expression. Participant-level intermediate files are excluded from the shared data files.

Finite multivariate reference and descriptive sample profiles, 2001 to 2004

The finite-reference calculation uses the five point-estimate vectors and their 399 survey replicates. For each imputation, replicate contrasts are centred on the original point estimate and combined with the saved survey scale and replicate multipliers. Let Ubar be their mean within-imputation covariance, B_MI the sample covariance of the five point vectors, and T=Ubar+(1+1/m)B_MI, with m=5. B_MI here denotes between-imputation uncertainty, not the cross-marker quantity B_cov.

For p=3 contrasts, the unrestricted-covariance statistic is $W=qbar'T^{-1}qbar$ and the F statistic is W/p. Following the small-sample proposal of Marchenko and Reiter (2009), define

$gamma=(1+1/m)\ trace(B\_MI\ T^{-1})/p$

$nu\_star=(m-1)/gamma^2$

$nu\_obs=(1-gamma)\ nu\_com\ (nu\_com+1)/(nu\_com+3)$

nu_BR=1/(1/nu_star+1/nu_obs).

The upper tail of F(p,nu_BR) supplies the joint P value, with nu_com=30. If gamma=0, nu_star is infinite. Applying the same rule to the verified measured subset gives nu_BR=28.1818. This retains unrestricted T but uses an average-FMI approximation for the denominator. It is not an exact multivariate test under arbitrary heterogeneity of missing information. Only five imputations are available. The three eigenvalues of the T-whitened matrix (1+1/m)B_MI describe directional FMI: 0.005, 0.027 and 0.124 in the primary sample, with average 0.052; all are zero in the verified measured subset. No covariance terms are discarded or truncated.

Supplementary Table S22 describes the fixed eligible populations, using original WTSAF2YR/2 weights. Weighted SD is sqrt(sum(w*(x−weighted_mean)$^2$)/sum(w)). For body measures, mean and SD are calculated separately in each released dataset and then averaged across all five; source tables retain the minimum and maximum. They are descriptive summaries, not Rubin intervals. Original age, BMI, waist, SAPA, sex, cycle, eligibility and measured-subset identity were verified to be invariant across versions. Waist uses variable-specific valid records. No outcome model, selection correction or comparison P value was added.

Marchenko and Reiter's small-sample proposal, especially pp. 390–391, supplies the multivariate degrees-of-freedom approximation used here; Barnard and Rubin supply the univariate interval reference.[21,22] The survey replicate design, chosen contrasts and Holm comparison families are specified separately above.

Separate-cycle replication in NHANES 1999 to 2000

The replication applied the preceding definitions to the official 1999 to 2000 DXX release. Of 1,618 eligible marker-panel candidates, 103 pregnant participants and two participants with other unusable records were excluded. The resulting 1,513 participants comprised 527 aged 20 to 39, 492 aged 40 to 59 and 494 aged 60 to 79. The 986 participants in the structural comparison were disjoint from the original 2,081. The replication remained within the same NHANES system and shared the fixed NHANES reference and official DXA imputation methods.

Single-cycle WTSAF2YR weights were used directly. The full positive-weight design universe comprised 3,428 participants, 13 strata and 27 primary sampling units, giving 14 complete-design degrees of freedom. One stratum contained three primary sampling units. With seed 4182026, 399 survey-design bootstrap draws were generated before domain restriction and shared across the five imputations. The mean models omitted the constant survey-cycle term; body measurement definitions, age spline, interactions and contrasts remained unchanged. Structural matrices were averaged equally across sexes. The periods were not merged, so the special weighting rules for combined 1999 to 2002 data did not enter these separate estimates.

The verified measured subset contained 1,215 participants, including 388 aged 40 to 59 and 381 aged 60 to 79. All required measured values were identical across the five releases; no eligible record had a no-imputation flag paired with changing required values. This replication added no cases to the 2003 to 2004 flag–quality discrepancy described above. The paired routine-body comparison used 978 participants, with identical inclusion for the DXA and BMI/waist representations. Supplementary Table S23 gives weighted sample profiles under the same descriptive convention as Supplementary Table S22, including female proportion, trunk and leg fat, and the trunk-to-leg fat ratio. Each ratio was calculated within person before weighting; it is not the ratio of the two weighted means. Female proportion uses the original survey weights. Body measures were summarized separately in each of the five releases and their means and SDs were averaged. The single-cycle table has no varying cycle indicator.

Pointwise intervals and joint inference followed the preceding rules with complete-design degrees of freedom of 14. For the three-contrast primary joint test, average FMI was 0.064, the denominator degrees of freedom were 11.429 and F was 0.045 ($P = 0.987$). The measured-subset FMI was zero, giving denominator degrees of freedom of $14\times15/17=12.353$ and $F=1.125$ ($P = 0.377$). Pointwise intervals and the paired comparator accompany the joint result in Supplementary Table S16 and the aggregate source files.

Six additive body-distribution components

The analysis reused the separately prepared 2001 to 2004 and 1999 to 2000 structural samples and their original survey replicate weights. Within each age, sex and cycle cell, identical weighted regression on an intercept, linear age and height residualized the eight-marker vector X and three log-body measures Z. Weighted covariances used the population denominator. Let $M=\mathrm{Cov}(X,Z)S^{-1}$ with $S=\mathrm{Cov}(Z)$, and $R=C-MSM'$. For the older-minus-middle-aged difference $\Delta S$, the original distribution component is $D=\frac{1}{2}(M_o\Delta S M_o'+M_m\Delta S M_m')$. Each diagonal entry of $\Delta S$ defined one variance component; each symmetric pair of off-diagonal entries defined one covariance component. Substituting those six matrices separately into D preserved its exact matrix sum. Applying $B(C)=u'Cu-\Sigma u_j^2C_{jj}$ gave six additive scalar contributions. For the BMI-omitted form, only the BMI entry of u was set to zero. Cell matrix products were evaluated before equal averaging over sexes and cycles.

Each imputation used all 399 saved design replicates, the original replicate scale and mean-squared-error centring. Rubin total variances and Barnard-Rubin degrees of freedom retained complete-design values of

30 and 14. Intervals describe each prespecified term; no family-level detection or rank-order claim was made. The six-term identity, original distribution estimates and every original distribution replicate were checked. The components use the selected body coordinates; total fat and the regional ratio have overlapping anatomy.

Frozen BMI/waist transfer between periods

The direction of transfer was fixed as 2001 to 2004 to 1999 to 2000. The required observed marker, body and design fields, including row identity, were identical across all five prepared versions. The 2,057 and 978 waist-complete participants were therefore analysed once, without adding multiple-imputation uncertainty to these observed inputs. Source log-BMI/log-waist mappings M and residual matrices R remained specific to age group, sex and cycle. The adapted prediction was the equal-cell average of $M_sS_tM_s'+R_s$. The direct source prediction used $S_s$ in the same expression. Each target-sex body covariance was paired with both source cycles before averaging. Target X entered only observed outcomes and diagnostics; perturbing target X summaries left predictions unchanged.

The primary quantity was the age difference in B with BMI cross-terms removed. Age-group levels and the full eight-marker B were also reported. Define $e_0$ as source-copy minus observed target and $e_1$ as adapted minus observed target. Positive $H=e_0^2-e_1^2$ favours adaptation. Source weights were perturbed with the target fixed, then target weights with the source fixed, using each period's saved 399-replicate design. Their joint design covariance matrices were added. This is a first-order two-sample replication approximation; predictions and observed target quantities remained paired within every perturbation. Scalar intervals used Welch-Satterthwaite degrees of freedom, $(V_s+V_t)^2/(V_s^2/30+V_t^2/14)$, or the single contributing design's degrees of freedom. Observed-only estimates retained their complete-design reference without an additional missing-information reduction.

Because a first-order squared-error approximation can degenerate near zero errors, the H interval was obtained by projecting a joint error ellipse through $e_0^2-e_1^2$. The prespecified radius was $\sqrt{(2F_{2,14}(0.95))}$; the full-rank covariance and numerical extrema were checked. This finite-reference projection is an approximate uncertainty interval, not an exact small-sample coverage result. No Wald P value for H was used.

For a frozen source mapping, target residuals $e=X-M_sZ$ gave $K=\mathrm{Cov}(Z,e)$ and the identity $C_t=M_sS_tM_s'+R_t+M_sK+K'M_s'$. The observed-minus-adapted gap was decomposed into the residual covariance shift and cross-term contribution, evaluated for each source cell before averaging. These diagnostics did not update predictions. Marginal range checks described target BMI/waist values outside source support; no participants were removed using those checks. Full source and target uncertainty, exact sample lineage, and error covariance accompany Supplementary Table S19.

### Supplementary Note 9 | Source data and reproducibility

Six main figures, nine Extended Data figures and three Supplementary Figures organise the evidence. Their legends and linked aggregate source tables specify analysis populations, reference scales and uncertainty definitions. Individual participant records and predictions are excluded from the shared data files.

#### Resampling units and conditioning

Resampling follows the dependence structure of each source. Participant, household, family and survey-design resamples serve different sampling targets; the replicate count alone does not make their uncertainty interchangeable. The table summarises the main procedures. The error-control map below states which intervals are pointwise or simultaneous.

| Analysis | Unit and count | Repeated estimation | Held fixed |
|---|---|---|---|
| Clinical loading congruence, Fig. 1c | Participants; 500 per cohort | Factor-model fitting and congruence | Genetic comparison vector, retained local relationships and source completion |
| Lower-category prediction increments, Fig. 2e | Household/community clusters; 200 attempted, qualified counts in Fig. 2e | Training transformations, models and censoring distributions | Original outer folds, model menu and external SAPA thresholds |
| Within-person waist change, Fig. 3d | Households; 999 shared draws | Regression and paired contrasts | Reference score, visit definitions and measured inputs |
| IL-1RA, Fig. 3f,g | Families; 200 | Training knots, coefficients and held-out evaluation | Family folds and selection of IL-1RA |
| Matched anatomical prediction, Fig. 3e,h | Survey jackknife; 60 replicates | Reciprocal-cycle training and paired evaluation | Model definitions and cycle roles |
| Body composition and age, Fig. 6d–h | Survey strata/primary sampling units; 399 per imputation | Conditional models and covariance components in each of five imputations, then pooling | Official imputed values and analysis definitions; imputations are not regenerated |
| Genetic information fractions, Fig. 5b | Joint summary-error draws; 5,000 | Derived information fractions and their contrasts | Fixed direction and the specified joint-error model; no participant resampling |
| Fasting insulin and body composition; Supplementary Table S8F–J | Survey strata/primary sampling units; 999 Rao–Wu draws | All specified regression coefficients and directional comparisons | Reference transformations/loadings, eligibility, model sets and assay equation |
| HbA1c omission and measured-test increment; S3F–H | Communities/households within five folds; 200 draws | All seven models, training standardisation and censoring weights | Reference marker weights, eligibility, fold assignments, model terms and penalty |

#### Analysis status and error-control map

The analyses were not preregistered. Local relationships were chosen using an initial genetic correlation screen before application to the stated clinical comparisons; the NHANES reference overlaps the 2015–

2018 anatomical sample. The CHARLS observation model followed a time-fit diagnostic, and IL-1RA was selected and evaluated in HRS without repeating biomarker selection within prediction folds. Review of baseline timing motivated the landmark design, specified before its effects were fitted and subsequently made the principal lower-category display. Fourteen MR-Egger intercept diagnostics were added to the saved harmonised inputs after the original MR estimates; instrument selection and the reported correction families were retained. Numerical qualification rules for the landmark refits were adopted after fitting warnings. The fasting insulin comparison set, three-test family and two sensitivity definitions were fixed before estimating their associations. The HbA1c-omission model set and glucose-group definitions were fixed before estimating their outcome comparisons. These decisions limit confirmatory interpretation. The map below specifies the complete comparison families and their error control; intervals are 95% unless stated otherwise.

| Analysis and display | Comparison family or status | Error control and interval interpretation |
|---|---|---|
| Clinical HR/OR associations; Fig. 2a, Supplementary Figure 1a; Supplementary Table S4 | Separate cohort and endpoint estimates | Pointwise model/sandwich intervals, conditional on the clinical reference; no pooled association or family-wide claim |
| Age/disease paired comparisons; Figs. 5d–f and 6c; Supplementary Table S13 | Six older-minus-younger contrasts across ELSA/CHARLS: F, SAPA gain, and Full8-minus-SAPA gain | Simultaneous family intervals; absolute model-score intervals remain marginal; predictions fixed in 1,000 resamples |
| Genetic information fractions; Fig. 5b | Four source-specific endpoint estimates | Paired reflected-draw percentile intervals from 5,000 joint summary-error draws; approximate, not clipped at 100% |
| MR; Fig. 4b, Extended Data Figure 5; Supplementary Table S9 | Seven outcomes per instrument set and estimator | Holm adjustment within each seven-outcome family; estimator-specific pointwise intervals |
| Local genetic analysis; Fig. 4c; Supplementary Table S10 | Qualified regions within each disease family and scale | Univariate threshold 0.05/2,495; discovery bivariate threshold 0.05/eligible-region count; replication uses the full discovery-union count; selected local display uses pointwise local-model intervals without adjustment for regional selection |
| Protein-anchor screen; Fig. 4d; Supplementary Table S11 | 1,300 molecularly eligible assays; 966 match gSAPA | Benjamini-Hochberg FDR over all 1,300 assays; 81 meet FDR<0.05; unavailable matches remain in the table |
| Functional profiles and vascular outcomes (Fig. 4e–h, Supplementary Table S12) | Eight of 11 regions had BMI-unadjusted ISI effects. Of 32 allele, endpoint and source combinations, 28 passed quality criteria. | Pointwise intervals for functional effects, T, disease ORs and MRI traits. Alternative covariance bounds apply to T. The variant-level disease and MRI comparisons are descriptive and have no family-wide multiplicity adjustment. |
| MRI vascular contrasts; Fig. 3c | Original 20-comparison family | Holm adjustment; normal-approximation intervals propagate covariance between correlations and remain pointwise |
| Matched anatomical constructs; Fig. 3e, Extended Data Figure 4; Supplementary Table S6 | Separate original families for MetS-minus-SAPA, SAPA-beyond-waist and SAPA-minus-PhenoAge questions | Original Holm families; 60 survey jackknife refits, pointwise t intervals with 30 degrees of freedom; comparisons use their own matched participants |

| Analysis and display | Comparison family or status | Error control and interval interpretation |
|---|---|---|
| No-imaging anatomical prediction; Fig. 3h; Supplementary Table S6 | Four primary contrasts across VAT/whole-leg fat; other displayed pairs descriptive | Holm adjustment for four primary MSE contrasts; paired RMSE displays retain pointwise t intervals with 30 degrees of freedom; no equivalence margin |
| Longitudinal waist/profile associations; Fig. 3d, Supplementary Figure 2; Supplementary Table S7 | Primary window plus specified sensitivity and direct comparisons | Pointwise percentile intervals from 999 shared household resamples; no simultaneous-coverage claim |
| IL-1RA prediction and age relation; Fig. 3f,g; Supplementary Table S8 | Five performance comparisons and a separate three-comparison age family | Holm adjustment within each family; marginal basic-bootstrap intervals from 200 family resamples with refitting |
| IL-1RA composition after BMI/CRP; S8E | Same 3,464 people and original five family folds | 200 complete family refits; seven paired MSE contrasts with Holm correction; pointwise basic intervals; conditional on prior marker selection |
| IL-1RA age transfer; Supplementary Figure 3; Supplementary Table S8 | Primary MAE improvement; three additional transfer contrasts | Primary basic-bootstrap interval; original three-comparison Bonferroni normal intervals for additional contrasts |
| NHANES age-structure summaries; Fig. 6a,b | Thirty-six entries: two periods, nine quantities and two summaries | Simultaneous intervals from maximum standardised errors in 500 survey resamples; nonlinear template was examined before being fixed |
| Body-association age contrasts; Fig. 6d,e; Supplementary Tables S14–S16 | Three individual age contrasts per period and their joint comparison | Holm-adjusted individual t tests; Barnard-Rubin intervals and joint finite-degree-of-freedom F approximation after Rubin pooling |
| Body-distribution decomposition; Fig. 6f–h; Supplementary Tables S17–S18 | Signed terms in an exact descriptive partition | Pointwise Rubin/Barnard-Rubin intervals; component magnitudes do not establish a family-wise ranking |
| Cross-period body-distribution transfer; Fig. 6i; Supplementary Table S19 | Squared-error improvement between two specified predictions | Approximate interval from a joint finite-reference error ellipse; no Wald P value for the squared-error contrast |
| Post-measurement lower categories; Fig. 2b–e; S5A–E | Two overlapping CHARLS sets and two ELSA endpoints, each with two timings | Three-endpoint Holm association families; 1,000 fixed-group resamples; 200 attempted complete refits, with convergence qualification reported separately; pointwise, not simultaneous intervals |
| HRS lower PCE; S21 | Two predefined continuous association models | Approximate profile-likelihood intervals under the simplified observation model; two reports |
| Fasting insulin, regional fat and the six-marker projection; S8F–J | Three primary pooled comparisons; cycle-specific and two specified sensitivities | Holm adjustment for the three pooled tests; pointwise t intervals from 999 survey resamples; percentile intervals for cosine similarity; supporting estimates are descriptive |
| HbA1c omission and measured-test increment; S3F–H | Two principal incremental-HbA1c comparisons and five supporting contrasts in three settings; two additional ELSA glucose groups | Exploratory, pointwise 95% percentile intervals from 200 paired cluster refits; no simultaneous-coverage or equivalence claim |

### Error control for liver-fat genetic comparisons

| Analysis and display | Comparison family or status | Error control and interval interpretation |
|---|---|---|
| Liver-fat–disease components; Supplementary Table S6 | Four endpoints × three components, separately in two specifications | Joint delta-method pointwise normal intervals; Holm correction across 12 component tests per specification |

Display details for clinical figures

Fig. 2a uses SAPA SDs of 1.426816 for ELSA MI, 1.425993 for ELSA stroke and 1.499031 for both CHARLS endpoints. Mortality-event counts without/with baseline CVD were 804/344 for all-cause death, 215/146 for CVD death and 174/115 for heart disease death. Complete observation and adjustment rules are given in Supplementary Note 2.

Supplementary Figure 1b,c quartile counts are 985/985/985/986 for ELSA MI, 989/989/989/989 for ELSA stroke, 1,657 in each CHARLS stroke quartile and 1,364/1,364/1,363/1,364 for CHARLS heart disease. These four-year populations were aged 50–89; probabilities integrate each cohort's stated observation and mortality model.

Source-study ethics and access

The study used existing cohort data and summary statistics, without new participant recruitment, biospecimen collection or laboratory work. Source-specific access information is provided in the data access table. All participant-level data were de-identified before use in this study.

NHANES was approved by the National Center for Health Statistics Research Ethics Review Board. For the 2015–2016 files, the source protocol was #2011-17. The 2017–2018 cycle continued #2011-17 through 26 October 2017 and used #2018-01 thereafter. These identifiers describe those two cycles; they are not assigned to the earlier 1999–2010 survey cycles.

The ELSA source approvals recorded for waves 6–10 were 11/SC/0374, 13/SC/0532, 15/SC/0526, 17/SC/0588 and 21/SC/0030, respectively; written consent was obtained for nurse assessments and blood sampling. These identifiers describe waves 6–10, not the earlier repeated-measurement waves. CHARLS household and anthropometric procedures were approved by the Peking University Institutional Review Board under IRB00001052-11015 and biomarker collection under IRB00001052-11014. The source study obtained separate signed consent for fieldwork, non-blood biomarkers, blood collection and blood storage. HRS data were accessed under an approved sensitive-data agreement; source-study consent is described in the HRS documentation.[23,24,25,20]

Genetic source-study ethics and consent procedures are documented in the publications for the reused resources.[26,27,28,7,1,2,15] Access to these summary statistics does not imply that this project held individual-level access to the original genetic cohorts. The archived source approvals and grant identifiers are not approvals or project-funding awards for the current secondary analysis.

### Data-product citations

The CHARLS national baseline data are described in the 2011–2012 baseline user guide.[29]

The HRS IL-1RA analyses used the 2016 Core, 2022 Tracker final version 1.0, 2016 Blood-Based Biomarker data, and the 2016 Venous Blood Study main and Additional Assays releases.[30,31,32,33] The module provenance table identifies the linked measurements and assay releases.

GWAS Catalog study identifiers, download dates and original publications are specified in the data access table; the database resource is cited separately.[34]

The supplementary HRS stroke analysis additionally used the 2018, 2020 and 2022 Core and Exit releases.[35,36,37,38,39,40] The 2022 Exit file was Early Version 1.0 (January 2025); the remaining Core and Exit releases cited here were final releases.

### Supplementary tables

#### Guide to the supplementary tables

Tables are arranged by topic and give the population, quantity estimated and uncertainty for each analysis. The companion Excel workbook contains Tables S1–S23 and tabular definitions from the Supplementary Notes. Linked source files provide the underlying aggregate results.

| Table | Content | Related display or purpose |
|---|---|---|
| Supplementary Table S1 | Clinical resources and principal study populations | Fig. 1a: data sources |
| Supplementary Table S2 | Analysis-specific populations and targets | Figs. 1–6: analysis populations and estimands |
| Supplementary Table S3 | Reduced-input scoring, fasting glucose bridge and prognosis | Fig. 1e and Extended Data Figure 3: reduced-input scoring; clinical HbA1c example |
| Supplementary Table S4 | Clinical associations with SAPA | Fig. 2a and Supplementary Figure 1a: disease associations |
| Supplementary Table S5 | Clinical information within lower published-risk categories | Figs. 2b–e and 5a: prognosis within risk categories |
| Supplementary Table S6 | Anatomical associations and matched prediction comparisons | Figs. 3a–c,e,h and Extended Data Figure 4: fat distribution and comparator models |
| Supplementary Table S7 | Longitudinal waist change and the shared marker profile | Fig. 3d and Supplementary Figure 2: within-person change |
| Supplementary Table S8 | Biochemical characterization, fasting insulin and IL-1RA | Fig. 3f,g and Supplementary Figure 3: insulin and IL-1RA |
| Supplementary Table S9 | Mendelian randomization estimates and diagnostics | Fig. 4b and Extended Data Figure 5: MR estimates and diagnostics |
| Supplementary Table S10 | Local genetic associations passing the specified cross-source criteria | Fig. 4c: local genetic associations |
| Supplementary Table S11 | Protein-anchor screen and assay coverage | Fig. 4d: protein anchors and coverage |
| Supplementary Table S12 | Functional discovery regions, insulin sensitivity projections and vascular outcomes | Fig. 4e–h: insulin sensitivity, vascular outcomes, marker contributions and regional fat associations |
| Supplementary Table S13 | Age-specific disease observation-record information | Fig. 5d–f: age-specific prognostic information |
| Supplementary Table S14 | Body composition associations in NHANES 2001 to 2004 | Fig. 6d,e and Extended Data Figure 6: primary body composition analysis |

| Table | Content | Related display or purpose |
|---|---|---|
| Supplementary Table S15 | Joint age comparisons and imputation uncertainty | Fig. 6d,e: joint age contrasts and imputation uncertainty |
| Supplementary Table S16 | Body composition results in NHANES 1999 to 2000 | Fig. 6d,e and Extended Data Figure 6: replication body composition analysis |
| Supplementary Table S17 | Body-distribution moments and their six additive contributions | Fig. 6g,h: body-distribution moments |
| Supplementary Table S18 | Body-related age differences in shared marker variation | Fig. 6f–h and Extended Data Figures 7–8: decomposition of age differences |
| Supplementary Table S19 | Frozen cross-period transfer and diagnostic comparisons | Fig. 6i: cross-period transfer |
| Supplementary Table S20 | Identifier sensitivity of the fixed clinical reference | Fixed-score reference sensitivity |
| Supplementary Table S21 | HRS lower-PCE observation support and limited association analysis | HRS lower-risk category: observation support |
| Supplementary Table S22 | Primary sample composition and the verified measured subset | Extended Data Figure 7: primary measured-scan subset |
| Supplementary Table S23 | Replication sample composition and the verified measured subset | Extended Data Figure 7: replication measured-scan subset |

Supplementary Table S1 | Clinical resources and principal study populations

| Resource | Population and design | Main analytical samples | Measurements and study role |
|---|---|---|---|
| NHANES: clinical reference | United States; cross-sectional health survey | 4,565 reference participants | Eight routine markers; fixed input transformations and clinical coordinate |
| NHANES 2015–2018 | United States; repeated cross-sectional survey | 4,000 for age-related structure; 1,876 for regional fat analyses | Routine-marker clustering; same-person DXA regional fat in adults aged 20–59 |
| NHANES 1999–2010 | United States; earlier survey periods | 11,053 for age-related structure | Comparison of the clustering pattern in a separate survey period |
| NHANES 2001–2004 and 1999–2000 | United States; two non-overlapping DXA samples within the earlier surveys | 3,242 and 1,513 for mean relationships; 2,081 and 986 for structural comparisons | Total fat, trunk-to-leg fat ratio and non-bone lean mass; mean relationships at ages 20–79 and clustering at ages 40–59 versus 60–79 |
| ELSA | England; longitudinal ageing cohort | 3,941; 101 myocardial infarction reports; 3,956; 142 stroke reports | Common structure, prospective disease association and age-related disease information |
| CHARLS | China; longitudinal ageing cohort | 7,423; 461 stroke reports; 6,178; 611 broad heart disease reports | Common structure, prospective disease association, age comparisons |
| HRS | United States; longitudinal ageing cohort | 2,282; 82 stroke reports; separate IL-1RA analysis of 3,464 participants from 2,680 families | Stroke association, biochemical correspondence and age-related information |

NHANES, National Health and Nutrition Examination Survey; DXA, dual-energy X-ray absorptiometry; ELSA, English Longitudinal Study of Ageing; CHARLS, China Health and Retirement Longitudinal Study; HRS, Health and Retirement Study; IL-1RA, interleukin-1 receptor antagonist. Sample sizes describe specific analyses and overlap within each resource. The reference included 4,265 complete observed panels with the retained reference handling. The CHARLS age–stroke comparison used a separate 6,628-person risk set. Full analysis-specific counts, age ranges, temporal definitions and supporting samples are retained in Supplementary Table S2. Genetic sources are mapped in Fig. 1 and documented with their trait-specific sample sizes in the relevant legends and Methods.

CHARLS association risk sets exclude any prior positive report before constructing follow-up. For stroke, 7,555 participants met entry criteria and 7,423 had positive follow-up time; the corresponding broad-heart-disease counts were 6,282 and 6,178. The post-measurement lower-category prediction populations follow the distinct entry rules in Supplementary Table S5A–E.

Supplementary Table S2 | Analysis-specific populations and targets

| Component | Resource | n | Target | Interpretation |
|---|---|---|---|---|
| NHANES reference | NHANES | 4565 | Frozen input transformations and loadings | 4265 complete observed panels; retained handling in reference |
| MI association | ELSA | 3941 | 101 reports; cohort-specific HR | Post-blood risk entry; incomplete mortality linkage |
| Stroke association | ELSA | 3956 | 142 reports; interval-observation Weibull HR | Stroke-negative Wave-7 entry following nurse examination |
| Broad heart disease association | CHARLS | 6178 | 611 reports; weighted illness-death model | 6282 eligible; 104 without positive follow-up; first positive histories restored |
| Four-year probability strata | ELSA MI / stroke | 3941 / 3956 | Five-fold held-out event probabilities | Same endpoint-specific samples as the current associations |
| Four-year probability strata | CHARLS stroke / broad heart disease | 6628 / 5455 | Cumulative onset before death | Age 50–89; current first-positive history rules |
| Stroke history | NHANES 2015–2018 | 4256 | 170 histories; survey-weighted OR | Complete observed D inputs; cross-sectional |
| Mortality without baseline CVD | NHANES 1999–2010 | 6573 | 804 all-cause / 215 CVD / 174 heart deaths | Eight-year associations and separate internal prediction validation |
| Mortality with established CVD | NHANES 1999–2010 | 1043 | 344 all-cause / 146 CVD / 115 heart deaths | Eight-year associations and separate internal prediction validation |
| Stroke association | CHARLS | 7423 | 461 reports; interval illness-death model | 7555 eligible; 132 without positive follow-up; first positive histories restored |
| Stroke association | HRS | 2282 | 82 reports; four-state observation model | Blood precedes 2018 negative entry; death/unknown paths retained |
| DXA anatomy | NHANES | 1876 | Five regions conditional on total fat | Ages20–59; same-cycle measurement; survey design |
| IL-1RA mapping | HRS | 3464 | Log concentration; five family folds | 2680 families; selected evaluation following the initial screen |
| Recent age structure | NHANES | 4000 | Fixed-axis variance decomposition | Population age comparison; not within-person decline |
| Earlier age structure | NHANES | 11053 | Fixed-axis variance decomposition | Separate survey period |
| MI age task | ELSA | 3941 | MI observation record | Ages50–69 versus70–89 |
| Stroke age task | CHARLS | 6628 | Conditional stroke record | 435 stroke reports; 514 deaths; original age 50–89 frame and community folds retained |
| IL-1RA transfer: source | HRS | 1757 | Train ages55–69 | Target calibration/test split by family |
| IL-1RA transfer: target | HRS | 1287 | Evaluate ages70–84 | Not an independent external cohort |
| Body composition mean relations | NHANES 2001 to 2004 | 3242 | Ages 20 to 79; 10% body measurement contrasts | Five official imputations; frozen D units |
| Body-related clustering | Same body composition sample | 2081 | Ages 40 to 59 versus 60 to 79 | 1115 and 966 participants; equal sex/cycle matrix weighting |
| Verified measured subset | Same sample | 2668 / 1676 | Mean / structural comparisons | No-imputation flags plus cross-version invariance |

| Component | Resource | n | Target | Interpretation |
|---|---|---|---|---|
| Routine-body comparator | Same sample | 2057 | Paired DXA versus BMI/waist | Identical waist-complete participants |
| Fasting insulin and regional fat distribution | NHANES 2001–2004 | 2475 | Ages 20–79; same-person conditional associations | No diagnosed diabetes; complete markers; positive fasting weights; verified measured DXA only |
| Prognosis after HbA1c omission | CHARLS / ELSA | 5478 / 1323 | Post-measurement stroke / MI and stroke | No known diabetes; 983 ELSA participants with glucose 70 to <100 mg/dL formed the primary evaluation subgroup; S3F–H |

| Component | Resource | n | Target | Interpretation |
|---|---|---|---|---|
| Replication mean relations | NHANES 1999 to 2000 | 1513 | Ages 20 to 79; same contrasts | Separate cycle; same frozen reference |
| Replication clustering | Same replication sample | 986 | 492 aged 40 to 59; 494 aged 60 to 79 | Equal sex matrix weighting |
| Replication verified measured | Same replication sample | 1215 / 769 | Mean / structural comparisons | Same flag and invariance definition |
| Replication routine-body comparator | Same replication sample | 978 | Paired DXA versus BMI/waist | Identical waist-complete participants |

| Component | Resource | n | Target | Interpretation |
|---|---|---|---|---|
| Six body-distribution terms | NHANES 2001 to 2004 / 1999 to 2000 | 2081 / 986 | Original structural samples | Same participants as existing body comparison |
| Frozen structural transfer | Source 2001 to 2004; target 1999 to 2000 | 2057 / 978 | BMI/waist-complete structural samples | Observed inputs analysed once; source and target error propagated |

Some participants contributed to several analyses. Sample sizes are given separately for each clinical analysis and for each genetic trait and source.

### Time origin, target and evaluation in the clinical analyses

Each row specifies the entry visit and observation rules for an analysis. Event counts refer to disease reports or linked deaths. Rows from the same cohort can include the same participants.

| Question and display | Analysis population | Time origin and target | Evaluation and uncertainty | Detailed specification |
|---|---|---|---|---|
| Prospective associations; Fig. 2a | ELSA MI: 3,941/101 reports; stroke: 3,956/142 | Endpoint-negative Wave 7 after Wave-6 nurse examination; interval-observed MI or stroke | Cohort-specific Weibull associations; household-cluster intervals | Methods, Clinical associations; Supplementary Note 2; Supplementary Table S4 |
| Prospective associations; Fig. 2a | CHARLS stroke: 7,423/461 reports; broad heart disease: 6,178/611 | Disease-negative 2013 interview after baseline blood measurement; earlier positives excluded | Two-period illness-death models; community-cluster intervals | Supplementary Note 2; Supplementary Table S4 |
| Additional stroke association; Supplementary Table S4 | HRS: 2,282/82 reports | Stroke-negative 2018 landmark after 2016 blood draw; median separation 1.795 years | Four-state observation model with death/unknown paths; household-cluster intervals | Supplementary Note 2; Supplementary Table S4 |
| Post-measurement lower categories; main Fig. 2b–e | CHARLS: 3,535/166 original, 2,195/81 intersection; ELSA: 1,293 per endpoint | 2013 CHARLS or Wave-7 ELSA disease-negative visit after measurement; approximately 7 or 6 years | Original baseline categories retained; 1,000 group resamples and 200 attempted complete refits per setting; numerical qualification reported | Supplementary Table S5A–E |

| Question and display | Analysis population | Time origin and target | Evaluation and uncertainty | Detailed specification |
|---|---|---|---|---|
| Mortality; Fig. 5c | NHANES without baseline CVD: 6,573/215 CVD deaths; with CVD: 1,043/146 | Examination to linked mortality over eight years in the 1999–2010 surveys | Survey-weighted associations; separate internally optimism-corrected prediction comparisons | Supplementary Table S4; Supplementary Figure 1a |
| Disease information by age; Fig. 5d–f | ELSA: 3,941/101 MI reports; CHARLS: 6,628/435 stroke reports and 514 deaths | MI observation record; stroke observation record conditional on observed survival | Held-out record likelihood, not an eight-year binary outcome; intervals conditional on fitted predictions | Supplementary Table S13; Supplementary Note 2 |

External NHANES cutoffs define phenotype groups. The principal lower-category analyses use the baseline risk category and start follow-up at the qualifying visit after blood measurement. Their Kaplan–Meier net probabilities differ from the four-year onset-before-death probabilities in the interval-observation models.

### Evidence roles across the physiological and genetic comparisons

| Question and display | Population or input | What was compared or evaluated | Scope of validation | Detailed specification |
|---|---|---|---|---|
| Common direction; Fig. 1b,c | Age 40–54 clinical samples: NHANES 1,104, CHARLS 3,127, ELSA 413; Pan-UKB and BBJ genetic profiles | Source-specific loadings and their similarity | Cross-source structural comparison; distinct from disease-model validation | Supplementary Note 1; Extended Data Figures 1 and 2 |
| Regional anatomy; Fig. 3a,e,h | Observed NHANES 2015–2018 DXA association sample: 1,876; matched prediction samples: 1,465, 1,867 or 1,869 | Same-person regional fat associations; reciprocal-cycle prediction for matched constructs and information sets | Between-cycle testing within NHANES; sample changes reflect required inputs | Supplementary Table S6; Supplementary Note 5 |
| Within-person change; Fig. 3d | ELSA waves 4/6: 2,959; overlapping waves 2/4: 2,246; CHARLS: 5,207 in the primary waist range | Change in waist and the BMI-omitted fixed profile, conditional on BMI change | Two-visit association; the two ELSA windows are not independent replications | Supplementary Table S7; Supplementary Figure 2 |
| Biochemical correspondence (Fig. 3f,g for IL-1RA) | NHANES, 2,475 participants without known diabetes. HRS, 3,464 people from 2,680 families. | NHANES fasting insulin and six-marker SAPA. HRS IL-1RA prediction in five family-separated folds after marker selection. | Cross-sectional associations in NHANES. Internal prediction in HRS, with marker selection outside the folds. | Supplementary Table S8 and Supplementary Note 6 |
| Genetic relationships; Figs. 3b,c, 4 and 5b | Eight routine-trait summary inputs, MRI/BIA and MVP/FinnGen endpoints; trait-specific sample sizes | Genetic correlations, MR, regional and protein associations, fixed marker projections, variant-level MI and stroke odds ratios, and panel-wide information fractions | Sources and endpoints remain distinct; shared participants and input traits preclude treating all layers as independent replications | Supplementary Notes 3–5; Supplementary Tables S6, S9–S12 |
| Age and body composition; Fig. 6d–h | NHANES 2001–2004: 3,242 mean/2,081 structural; 1999–2000: 1,513 mean/986 structural | The same mean-relation and age-decomposition definitions in two non-overlapping survey periods | Separate participants, with shared survey procedures and official imputation methods | Supplementary Tables S14–S18, S22–S23; Supplementary Note 8 |
| Fixed-model transfer; Fig. 6i | Waist-complete source/target samples: 2,057/978 | Substitute target BMI/waist covariance into the fixed source model; evaluate target marker covariance | Cross-period prediction test using routine body proxies; distinct from the DXA decomposition | Supplementary Table S19; Extended Data Figure 8 |

Supplementary Table S3 | Reduced-input scoring, fasting glucose bridge and prognosis

### A. One-at-a-time omission

| Cohort | Omitted marker | n | Pearson r | RMSE (Q) | Median / 90th absolute rank shift | Quarter disagreement (%) |
|---|---|---|---|---|---|---|
| NHANES | SBP | 4265 | 0.983 | 0.358 | 3 / 9 | 14.9 |
| NHANES | DBP | 4265 | 0.989 | 0.283 | 2 / 7 | 12.4 |
| NHANES | LDL | 4265 | 0.983 | 0.349 | 3 / 9 | 15.2 |
| NHANES | HDL | 4265 | 0.978 | 0.412 | 3 / 9 | 16.2 |
| NHANES | TG | 4265 | 0.958 | 0.601 | 5 / 15 | 24.9 |
| NHANES | HbA1c | 4265 | 0.979 | 0.392 | 2 / 6 | 10.1 |
| NHANES | BMI | 4265 | 0.975 | 0.449 | 3 / 10 | 16.5 |
| NHANES | CRP | 4265 | 0.977 | 0.423 | 4 / 11 | 15.9 |
| ELSA | SBP | 4260 | 0.974 | 0.358 | 4 / 11 | 17.3 |
| ELSA | DBP | 4260 | 0.986 | 0.262 | 3 / 8 | 12.7 |
| ELSA | LDL | 4260 | 0.966 | 0.393 | 4 / 13 | 20.6 |
| ELSA | HDL | 4260 | 0.962 | 0.491 | 5 / 13 | 22.3 |
| ELSA | TG | 4260 | 0.957 | 0.545 | 5 / 15 | 24.0 |
| ELSA | HbA1c | 4260 | 0.982 | 0.284 | 2 / 7 | 10.3 |
| ELSA | BMI | 4260 | 0.981 | 0.327 | 3 / 9 | 14.9 |
| ELSA | CRP | 4260 | 0.980 | 0.325 | 4 / 10 | 17.4 |
| CHARLS | SBP | 8335 | 0.963 | 0.444 | 5 / 13 | 20.7 |
| CHARLS | DBP | 8335 | 0.984 | 0.309 | 3 / 9 | 14.7 |
| CHARLS | LDL | 8335 | 0.976 | 0.332 | 3 / 11 | 16.0 |
| CHARLS | HDL | 8335 | 0.976 | 0.377 | 3 / 11 | 17.2 |
| CHARLS | TG | 8335 | 0.934 | 0.622 | 6 / 18 | 29.6 |
| CHARLS | HbA1c | 8335 | 0.978 | 0.383 | 2 / 7 | 10.6 |
| CHARLS | BMI | 8335 | 0.980 | 0.476 | 2 / 7 | 11.7 |
| CHARLS | CRP | 8335 | 0.981 | 0.357 | 4 / 10 | 15.9 |

Ranks are percentile points relative to each version's own weighted cohort reference. Q is the unrescaled NHANES-reference coordinate. Quartile disagreement is the percentage of participants who changed score quartile when one measured test was masked. Entries are descriptive point estimates.

### B. Frozen glucose bridge within its development range

| Validation panel | Version | n | Score r | Score bias | Score RMSE | HbA1c RMSE (%) | 90th absolute rank shift |
|---|---|---|---|---|---|---|---|
| NHANES holdout | Omission | 2097 | 0.980 | 0.032 | 0.369 | 0.903 | 6 |
| NHANES holdout | Glucose estimate | 2097 | 0.993 | 0.019 | 0.204 | 0.499 | 5 |
| ELSA fasting | Omission | 2438 | 0.989 | -0.002 | 0.212 | 0.518 | 6 |
| ELSA fasting | Glucose estimate | 2438 | 0.993 | -0.160 | 0.236 | 0.578 | 8 |
| CHARLS recorded glucose | Omission | 8316 | 0.978 | 0.204 | 0.380 | 0.931 | 7 |
| CHARLS recorded glucose | Glucose estimate | 8316 | 0.983 | 0.176 | 0.334 | 0.817 | 9 |

Bias is estimated minus complete Q. HbA1c RMSE is in percentage points. Score ranks for omission use the reduced reference; glucose estimates use the complete reference. External correlation can improve while bias worsens agreement. The CHARLS estimates apply to recorded glucose values. Fasting status could not be verified from the sampling codes.

### C. Recorded treatment, diabetes and sampling-code strata

| Panel and recorded grouping | Status/code | Version | n | Score bias | Score RMSE |
|---|---|---|---|---|---|
| NHANES / glucose-lowering medication | 0 | Omission | 1758 | 0.113 | 0.243 |
| NHANES / glucose-lowering medication | 0 | Glucose estimate | 1758 | 0.036 | 0.158 |
| NHANES / glucose-lowering medication | 1 | Omission | 339 | -0.586 | 0.849 |
| NHANES / glucose-lowering medication | 1 | Glucose estimate | 339 | -0.118 | 0.411 |
| ELSA / reported diabetes | 0 | Omission | 2318 | 0.015 | 0.179 |
| ELSA / reported diabetes | 0 | Glucose estimate | 2318 | -0.153 | 0.224 |
| ELSA / reported diabetes | 1 | Omission | 76 | -0.484 | 0.655 |
| ELSA / reported diabetes | 1 | Glucose estimate | 76 | -0.339 | 0.472 |
| CHARLS / glucose-lowering medication | 0 | Omission | 8001 | 0.230 | 0.350 |
| CHARLS / glucose-lowering medication | 0 | Glucose estimate | 8001 | 0.176 | 0.325 |
| CHARLS / glucose-lowering medication | 1 | Omission | 315 | -0.339 | 0.786 |
| CHARLS / glucose-lowering medication | 1 | Glucose estimate | 315 | 0.173 | 0.480 |
| CHARLS / raw fasting code 1 | See panel | Omission | 7490 | 0.198 | 0.380 |
| CHARLS / raw fasting code 1 | See panel | Glucose estimate | 7490 | 0.167 | 0.316 |
| CHARLS / raw fasting code 2 | See panel | Omission | 694 | 0.229 | 0.351 |
| CHARLS / raw fasting code 2 | See panel | Glucose estimate | 694 | 0.280 | 0.484 |

Medication and reported diabetes use their source 0/1 coding. Binary strata include participants with known status. The ELSA groups use reported diabetes history. CHARLS results are shown by recorded sampling code because its clinical meaning could not be verified.

### D. Range support and model identity

| Panel | Positive glucose n | Within 21–479 mg/dL | Outside range |
|---|---|---|---|
| NHANES_holdout | 2097 | 2097 | 0 |
| ELSA_fasting | 2438 | 2438 | 0 |
| CHARLS_recorded_glucose | 8325 | 8316 | 9 |

The two-coefficient weighted model was developed once in NHANES 2015–2016 (n=2,168), with no cohort-specific recalibration. The fixed NHANES reference itself includes both NHANES cycles. Source Data provides all omission metrics, all glucose metrics including out-of-range observations, subgroups, range support, input support and the frozen model. Sections A–E assess score agreement; sections F–H evaluate prognosis after HbA1c omission.

### E. HbA1c plus one additional unavailable measurement

The frozen glucose estimate was combined with six measured contributions. The additional omitted marker remained absent. Agreement with the matching measured seven-marker score and with complete Q is reported separately. The model was not refitted.

| Panel | Other omission | n | r, matching seven | RMSE to complete Q: six measured only | RMSE to complete Q: with estimated HbA1c | Absolute rank shift: median / P90 |
|---|---|---|---|---|---|---|
| NHANES 2017–2018 | SBP | 2097 | 0.992 | 0.583 | 0.444 | 2 / 5 |
| NHANES 2017–2018 | DBP | 2097 | 0.993 | 0.475 | 0.346 | 2 / 5 |
| NHANES 2017–2018 | LDL | 2097 | 0.993 | 0.505 | 0.412 | 2 / 5 |
| NHANES 2017–2018 | HDL | 2097 | 0.992 | 0.571 | 0.426 | 2 / 5 |
| NHANES 2017–2018 | TG | 2097 | 0.989 | 0.783 | 0.645 | 2 / 6 |
| NHANES 2017–2018 | BMI | 2097 | 0.991 | 0.641 | 0.510 | 2 / 5 |
| NHANES 2017–2018 | CRP | 2097 | 0.992 | 0.587 | 0.456 | 2 / 5 |
| ELSA fasting | SBP | 2438 | 0.991 | 0.432 | 0.466 | 3 / 9 |
| ELSA fasting | DBP | 2438 | 0.992 | 0.330 | 0.384 | 3 / 8 |
| ELSA fasting | LDL | 2438 | 0.992 | 0.433 | 0.501 | 3 / 8 |
| ELSA fasting | HDL | 2438 | 0.990 | 0.567 | 0.486 | 4 / 9 |
| ELSA fasting | TG | 2438 | 0.988 | 0.564 | 0.600 | 4 / 10 |
| ELSA fasting | BMI | 2438 | 0.991 | 0.414 | 0.363 | 4 / 9 |
| ELSA fasting | CRP | 2438 | 0.991 | 0.415 | 0.372 | 3 / 8 |
| CHARLS recorded glucose | SBP | 8316 | 0.978 | 0.571 | 0.525 | 4 / 11 |
| CHARLS recorded glucose | DBP | 8316 | 0.980 | 0.464 | 0.423 | 4 / 10 |
| CHARLS recorded glucose | LDL | 8316 | 0.982 | 0.529 | 0.496 | 3 / 10 |
| CHARLS recorded glucose | HDL | 8316 | 0.978 | 0.514 | 0.446 | 4 / 10 |

| Panel | Other omission | n | r, matching seven | RMSE to complete Q: six measured only | RMSE to complete Q: with estimated HbA1c | Absolute rank shift: median / P90 |
|---|---|---|---|---|---|---|
| CHARLS recorded glucose | TG | 8316 | 0.973 | 0.712 | 0.617 | 4 / 12 |
| CHARLS recorded glucose | BMI | 8316 | 0.980 | 0.727 | 0.674 | 3 / 10 |
| CHARLS recorded glucose | CRP | 8316 | 0.980 | 0.609 | 0.549 | 3 / 10 |

For the matching measured seven-marker target, six-measurement omission versus glucose-estimated RMSE was 0.369 versus 0.204 in NHANES, 0.212 versus 0.236 in ELSA and 0.380 versus 0.334 in CHARLS, identical across the seven other omissions by algebra. In contrast, the complete-Q error also includes the missing contribution of the other marker. Rank shifts use the matching seven-marker reference for both estimates and targets; the six-measurement-only comparator has no deployed percentile reference. These are descriptive masked-complete-panel checks without resampling intervals. CHARLS fasting-code meaning remains unresolved. Aggregate results.

### F. Prognostic evaluation populations after HbA1c omission

All participants had no known diabetes at the qualifying visits. CHARLS used positive recorded glucose without requiring verified fasting. ELSA training used all 1,323 eligible fasting participants; primary evaluation used glucose 70 to <100 mg/dL. The wider glucose range and all-eligible ELSA group were supporting comparisons. Each row counts distinct participants. Some participants appear in several rows.

| Population / endpoint | Glucose group | n | All subsequent reports | Reports definitely by horizon | Intervals crossing horizon | Negative through horizon |
|---|---|---|---|---|---|---|
| CHARLS Stroke | All eligible without known diabetes | 5478 | 381 | 381 | 0 | 4576 |
| ELSA Stroke | Fasting glucose 70 to <100 mg/dL | 983 | 18 | 4 | 14 | 734 |
| ELSA Stroke | Fasting glucose 70 to <110 mg/dL | 1228 | 22 | 4 | 18 | 912 |
| ELSA Stroke | All eligible without known diabetes | 1323 | 25 | 4 | 21 | 971 |
| ELSA MI | Fasting glucose 70 to <100 mg/dL | 983 | 13 | 9 | 4 | 740 |
| ELSA MI | Fasting glucose 70 to <110 mg/dL | 1228 | 18 | 13 | 5 | 917 |
| ELSA MI | All eligible without known diabetes | 1323 | 23 | 15 | 8 | 976 |

Horizons were approximately seven years for CHARLS and six years for ELSA. Seven-marker SAPA omits only the fixed HbA1c term. Full7 and Full8 enter the corresponding markers separately. Every prediction model includes the common background terms described in Supplementary Note 2. These are complete-panel masking analyses, not observations of clinically selected missing tests.

### G. Paired gains in interval-record prediction

Values are improvements in held-out mean log probability, in 10^-3 nat per participant [pointwise 95% interval]. Positive values favour the added or first-named information. Every contrast uses the same participants and five folds. Intervals use 200 complete cluster refits. The first two contrasts directly evaluate measured HbA1c; the remaining five are supporting comparisons. No multiplicity-adjusted or equivalence claim is made.

| Comparison | CHARLS stroke | ELSA stroke | ELSA MI |
|---|---|---|---|
| HbA1c added to background | 0.247 [-0.409, 0.913] | 0.890 [-2.820, 4.398] | -0.287 [-3.025, 1.087] |

| Comparison | CHARLS stroke | ELSA stroke | ELSA MI |
|---|---|---|---|
| HbA1c added to seven-marker SAPA | 0.083 [-0.519, 0.605] | 0.998 [-2.535, 4.781] | -0.088 [-2.815, 1.764] |
| Complete versus seven-marker SAPA | 0.172 [-0.299, 0.622] | 0.082 [-0.620, 0.863] | -0.439 [-1.679, 0.371] |
| HbA1c added to Full7 | 0.050 [-0.588, 0.555] | 0.840 [-7.173, 5.759] | -0.419 [-4.981, 2.096] |
| Seven-marker SAPA added to background | 6.494 [3.490, 10.427] | -3.291 [-12.556, 1.319] | 5.518 [0.267, 12.350] |
| Seven-marker SAPA added after HbA1c | 6.330 [3.384, 10.136] | -3.182 [-13.557, 1.122] | 5.717 [0.291, 12.445] |
| Full8 versus seven-marker SAPA plus HbA1c | -0.307 [-2.761, 0.632] | 0.846 [-46.302, 8.391] | -3.542 [-20.664, 2.177] |

The seven contrasts are repeated for glucose subgroups of the same ELSA cohort using the same fitted models.

| ELSA glucose group | Comparison | Stroke | MI |
|---|---|---|---|
| Fasting glucose 70 to <110 mg/dL | HbA1c added to background | 0.768 [-3.114, 3.830] | -0.404 [-3.706, 0.952] |
| Fasting glucose 70 to <110 mg/dL | HbA1c added to seven-marker SAPA | 0.797 [-2.795, 4.101] | -0.411 [-3.065, 1.044] |
| Fasting glucose 70 to <110 mg/dL | Complete versus seven-marker SAPA | 0.111 [-0.478, 0.756] | -0.193 [-1.119, 0.353] |
| Fasting glucose 70 to <110 mg/dL | HbA1c added to Full7 | 0.694 [-5.706, 4.606] | -0.418 [-4.957, 2.022] |
| Fasting glucose 70 to <110 mg/dL | Seven-marker SAPA added to background | -2.553 [-10.717, 1.037] | 3.503 [-1.106, 8.094] |
| Fasting glucose 70 to <110 mg/dL | Seven-marker SAPA added after HbA1c | -2.524 [-10.831, 1.137] | 3.496 [-1.033, 8.742] |
| Fasting glucose 70 to <110 mg/dL | Full8 versus seven-marker SAPA plus HbA1c | 1.728 [-35.430, 6.787] | -4.096 [-21.396, 0.790] |
| All eligible without known diabetes | HbA1c added to background | 0.337 [-32.710, 2.783] | -0.450 [-5.410, 1.970] |
| All eligible without known diabetes | HbA1c added to seven-marker SAPA | 0.286 [-84.871, 2.880] | -0.459 [-4.868, 2.506] |
| All eligible without known diabetes | Complete versus seven-marker SAPA | 0.038 [-0.656, 0.733] | -0.369 [-2.756, 0.387] |
| All eligible without known diabetes | HbA1c added to Full7 | -1.465 [-2454.897, 2.717] | -0.509 [-13.909, 4.427] |
| All eligible without known diabetes | Seven-marker SAPA added to background | -2.768 [-10.157, 1.014] | 3.647 [-1.529, 8.521] |
| All eligible without known diabetes | Seven-marker SAPA added after HbA1c | -2.819 [-50.034, 1.320] | 3.638 [-1.357, 9.231] |
| All eligible without known diabetes | Full8 versus seven-marker SAPA plus HbA1c | -1.167 [-2390.623, 5.647] | -4.579 [-33.829, -0.063] |

### H. Fixed-horizon discrimination and prediction error

These auxiliary comparisons use the same interval-model predictions. Main rows below use midpoint event timing and training-fold inverse censoring weights. AUC is unitless; Brier values and their improvements are multiplied by 1,000. Each cell gives estimate [pointwise 95% interval]. CHARLS includes all eligible recorded-glucose participants; ELSA rows use glucose 70 to <100 mg/dL.

| Population / endpoint | Model | AUC | Brier × 1,000 |
|---|---|---|---|
| CHARLS stroke | Background | 0.617 [0.571, 0.641] | 67.650 [61.583, 74.122] |
| CHARLS stroke | Background + HbA1c | 0.623 [0.573, 0.648] | 67.644 [61.656, 74.084] |
| CHARLS stroke | Background + seven-marker SAPA | 0.675 [0.638, 0.698] | 66.701 [60.841, 73.130] |

| Population / endpoint | Model | AUC | Brier × 1,000 |
| --- | --- | --- | --- |
| CHARLS stroke | Background + seven-marker SAPA + HbA1c | 0.675 [0.638, 0.699] | 66.704 [60.888, 73.173] |
| CHARLS stroke | Background + complete SAPA | 0.676 [0.637, 0.699] | 66.681 [60.888, 73.143] |
| CHARLS stroke | Background + Full7 | 0.672 [0.627, 0.693] | 66.785 [61.081, 73.686] |
| CHARLS stroke | Background + Full8 | 0.673 [0.627, 0.692] | 66.791 [61.163, 73.630] |
| ELSA stroke | Background | 0.517 [0.224, 0.680] | 21.229 [13.136, 33.018] |
| ELSA stroke | Background + HbA1c | 0.564 [0.278, 0.717] | 21.214 [13.123, 32.944] |
| ELSA stroke | Background + seven-marker SAPA | 0.469 [0.194, 0.649] | 21.338 [13.151, 33.105] |
| ELSA stroke | Background + seven-marker SAPA + HbA1c | 0.510 [0.242, 0.683] | 21.335 [13.141, 32.997] |
| ELSA stroke | Background + complete SAPA | 0.474 [0.208, 0.654] | 21.337 [13.151, 33.112] |
| ELSA stroke | Background + Full7 | 0.624 [0.353, 0.761] | 21.557 [13.354, 33.663] |
| ELSA stroke | Background + Full8 | 0.644 [0.353, 0.771] | 21.637 [13.281, 33.721] |
| ELSA MI | Background | 0.524 [0.200, 0.720] | 13.367 [6.343, 18.762] |
| ELSA MI | Background + HbA1c | 0.507 [0.214, 0.681] | 13.388 [6.408, 18.827] |
| ELSA MI | Background + seven-marker SAPA | 0.665 [0.344, 0.836] | 13.122 [6.314, 18.505] |
| ELSA MI | Background + seven-marker SAPA + HbA1c | 0.666 [0.351, 0.829] | 13.199 [6.382, 18.576] |
| ELSA MI | Background + complete SAPA | 0.661 [0.320, 0.821] | 13.131 [6.322, 18.541] |
| ELSA MI | Background + Full7 | 0.646 [0.312, 0.836] | 13.434 [6.688, 20.035] |
| ELSA MI | Background + Full8 | 0.639 [0.294, 0.819] | 13.510 [6.672, 20.425] |

| Paired AUC gain, midpoint timing | CHARLS stroke | ELSA stroke | ELSA MI |
| --- | --- | --- | --- |
| HbA1c added to background | 0.005 [-0.002, 0.014] | 0.047 [-0.031, 0.200] | -0.016 [-0.090, 0.040] |
| HbA1c added to seven-marker SAPA | 0.001 [-0.003, 0.005] | 0.041 [-0.033, 0.140] | 0.000 [-0.046, 0.031] |
| Complete versus seven-marker SAPA | 0.001 [-0.002, 0.005] | 0.005 [-0.010, 0.020] | -0.005 [-0.029, 0.009] |
| HbA1c added to Full7 | 0.000 [-0.003, 0.005] | 0.019 [-0.027, 0.081] | -0.008 [-0.051, 0.016] |
| Seven-marker SAPA added to background | 0.057 [0.033, 0.089] | -0.048 [-0.182, 0.076] | 0.142 [0.015, 0.349] |
| Seven-marker SAPA added after HbA1c | 0.053 [0.030, 0.085] | -0.054 [-0.163, 0.066] | 0.158 [0.016, 0.377] |
| Full8 versus seven-marker SAPA plus HbA1c | -0.003 [-0.018, 0.003] | 0.134 [-0.071, 0.296] | -0.027 [-0.248, 0.144] |

| Paired AUC gain, first-positive timing | CHARLS stroke | ELSA stroke | ELSA MI |
| --- | --- | --- | --- |
| HbA1c added to background | 0.005 [-0.002, 0.014] | 0.045 [-0.081, 0.232] | -0.028 [-0.137, 0.057] |
| HbA1c added to seven-marker SAPA | 0.001 [-0.003, 0.005] | 0.072 [-0.052, 0.278] | 0.000 [-0.065, 0.026] |
| Complete versus seven-marker SAPA | 0.001 [-0.002, 0.005] | -0.009 [-0.028, 0.021] | -0.004 [-0.025, 0.013] |
| HbA1c added to Full7 | 0.000 [-0.003, 0.005] | 0.016 [-0.029, 0.106] | -0.008 [-0.064, 0.017] |

| Paired AUC gain, first-positive timing | CHARLS stroke | ELSA stroke | ELSA MI |
|---|---|---|---|
| Seven-marker SAPA added to background | 0.057 [0.033, 0.089] | -0.174 [-0.419, 0.064] | 0.105 [0.000, 0.338] |
| Seven-marker SAPA added after HbA1c | 0.053 [0.030, 0.085] | -0.147 [-0.389, 0.067] | 0.133 [-0.003, 0.382] |
| Full8 versus seven-marker SAPA plus HbA1c | -0.003 [-0.018, 0.003] | 0.048 [-0.265, 0.512] | 0.023 [-0.168, 0.195] |

First-positive ELSA stroke AUC intervals use 196 evaluable resamples because four contained no reports by six years; other displayed intervals use 200. ELSA timing differences are material because most stroke-event intervals crossed six years. The complete model metrics and paired contrasts include both timing rules, all glucose groups, Brier comparisons and observed-to-predicted ratios. The absolute-risk estimates depend on how events were dated and censoring handled. Here HbA1c was evaluated for prediction of these vascular outcomes.

### Supplementary Table S4 | Clinical associations with SAPA

Analysis-specific standard-deviation scales and outcome definitions follow Fig. 2a and Supplementary Figure 1a. HR, hazard ratio; OR, odds ratio.

| Cohort and outcome | Measure | n | Reports or deaths | Estimate [95% CI] |
|---|---|---|---|---|
| ELSA: MI | HR | 3,941 | 101 | 1.588 [1.287, 1.960] |
| ELSA: stroke | HR | 3,956 | 142 | 1.456 [1.202, 1.763] |
| CHARLS: stroke | HR | 7,423 | 461 | 1.534 [1.344, 1.749] |
| CHARLS: heart disease | HR | 6,178 | 611 | 1.132 [1.007, 1.272] |
| HRS: stroke | HR | 2,282 | 82 | 1.146 [0.929, 1.414] |
| NHANES: stroke history | OR | 4,256 | 170 | 1.440 [1.091, 1.899] |
| No CVD: all-cause death | HR | 6,573 | 804 | 1.086 [0.986, 1.196] |
| No CVD: CVD death | HR | 6,573 | 215 | 1.093 [0.857, 1.394] |
| No CVD: heart death | HR | 6,573 | 174 | 1.161 [0.901, 1.497] |
| Prior CVD: all-cause death | HR | 1,043 | 344 | 1.145 [0.979, 1.340] |
| Prior CVD: CVD death | HR | 1,043 | 146 | 1.154 [0.893, 1.491] |
| Prior CVD: heart death | HR | 1,043 | 115 | 1.239 [0.888, 1.728] |

HRS uses observed-input NHANES-reference SAPA, scaled by SD 1.513622 in 2,282 participants at a stroke-negative 2018 landmark following the 2016 venous blood draw; 82 stroke histories were observed. Its piecewise illness-death model includes age (natural spline, three degrees of freedom), sex and baseline smoking, with household-cluster direct-score sandwich and t-based 95% intervals (1,850 degrees of freedom). The sample includes 112 deaths, with 15 unresolved stroke paths. The HRS HR is 1.146 (95% CI 0.929-1.414). HRS biochemical associations are shown in Fig. 3f,g; the age-transfer comparisons are in Supplementary Figure 3.

Supplementary Table S5 | Clinical information within lower published-risk categories

All results in this table use a disease-free entry visit after blood measurement. Sections A–E describe population support, continuous associations, fixed SAPA groups, matched prediction and numerical qualification. Interval-midpoint and first-positive-report assignments are evaluated in the same eligible populations.

### A. Post-measurement landmark populations and observation support

| Population | Original lower n | Eligible lower at landmark | Analysed lower n | Training n | All later reports | Horizon, years |
|---|---|---|---|---|---|---|
| CHARLS original | 4054 | 3621 | 3535 | 5481 | 166 | 7.001 |
| CHARLS intersection | 2520 | 2256 | 2195 | 5481 | 81 | 7.001 |
| ELSA stroke | 1530 | 1375 | 1293 | 2110 | 17 | 6.000 |
| ELSA MI | 1530 | 1375 | 1293 | 2110 | 15 | 6.000 |

CHARLS excluded 170 of 5,651 landmark-eligible training participants without positive follow-up duration; ELSA excluded 133 of 2,243. Lower-category exclusions without follow-up were 86 and 61 for the two CHARLS sets and 82 for ELSA. Eligibility required the later disease-negative, diabetes-negative visit and documented ordering after blood/nurse measurement. Scores, risk categories and covariates remained defined at the original baseline.

| Population and timing | Assigned events by horizon | Event intervals crossing horizon | Confirmed negative through horizon | Non-events last negative before horizon |
|---|---|---|---|---|
| CHARLS original; midpoint | 166 | 0 | 3113 | 256 |
| CHARLS original; first positive | 166 | 0 | 3113 | 256 |
| CHARLS intersection; midpoint | 81 | 0 | 1983 | 131 |
| CHARLS intersection; first positive | 81 | 0 | 1983 | 131 |
| ELSA stroke; midpoint | 16 | 12 | 959 | 317 |
| ELSA stroke; first positive | 5 | 12 | 959 | 317 |
| ELSA MI; midpoint | 11 | 6 | 961 | 317 |
| ELSA MI; first positive | 9 | 6 | 961 | 317 |

### B. Continuous SAPA associations after the landmark

| Population and timing | n / events | HR per raw Q unit [95% CI] | Nominal P | Three-endpoint Holm-adjusted P | Qualification |
|---|---|---|---|---|---|
| CHARLS original; midpoint | 3535 / 166 | 1.440 [1.270, 1.634] | $1.44 \times 10^{-8}$ | $4.32 \times 10^{-8}$ | No fitting warning |
| CHARLS original; first positive | 3535 / 166 | 1.436 [1.268, 1.626] | $1.18 \times 10^{-8}$ | $3.55 \times 10^{-8}$ | No fitting warning |
| CHARLS intersection; midpoint | 2195 / 81 | 1.484 [1.255, 1.755] | $4.00 \times 10^{-6}$ | Overlapping sensitivity | No fitting warning |
| CHARLS intersection; first positive | 2195 / 81 | 1.484 [1.255, 1.755] | $3.94 \times 10^{-6}$ | Overlapping sensitivity | No fitting warning |
| ELSA stroke; midpoint | 1293 / 16 | 2.009 [0.969, 4.169] | 0.061 | 0.122 | No fitting warning |

| Population and timing | n / events | HR per raw Q unit [95% CI] | Nominal P | Three-endpoint Holm-adjusted P | Qualification |
|---|---|---|---|---|---|
| ELSA stroke; first positive | 1293 / 5 | 1.140 [0.211, 6.157] | 0.879 | 0.987 | Smoking separation warning; diagnostic only |
| ELSA MI; midpoint | 1293 / 11 | 1.299 [0.531, 3.178] | 0.566 | 0.566 | No fitting warning |
| ELSA MI; first positive | 1293 / 9 | 0.751 [0.332, 1.703] | 0.494 | 0.987 | No fitting warning |

Models adjust for the original equation logit, baseline age, sex and smoking. Intervals use community/household sandwich errors. These are conditional associations among later disease-free attendees. SAPA proportional-hazards P values were 0.772–0.859 for CHARLS; the full term-level diagnostic table is supplied in Source Data. The ELSA first-positive stroke fit does not support a stable adjusted effect interpretation.

### C. Fixed SAPA groups after the landmark

| Population and timing | SAPA group | n / assigned events | Kaplan–Meier net probability, % [95% interval] |
|---|---|---|---|
| CHARLS original; midpoint | Low | 1267 / 31 | 2.560 [1.742, 3.431] |
| CHARLS original; midpoint | Middle | 2030 / 114 | 5.871 [4.773, 6.989] |
| CHARLS original; midpoint | High | 238 / 21 | 9.351 [5.906, 13.037] |
| CHARLS original; first positive | Low | 1267 / 31 | 2.564 [1.746, 3.436] |
| CHARLS original; first positive | Middle | 2030 / 114 | 5.877 [4.776, 6.995] |
| CHARLS original; first positive | High | 238 / 21 | 9.351 [5.906, 13.037] |
| CHARLS intersection; midpoint | Low | 932 / 18 | 2.007 [1.199, 2.880] |
| CHARLS intersection; midpoint | Middle | 1178 / 59 | 5.192 [3.841, 6.712] |
| CHARLS intersection; midpoint | High | 85 / 4 | 4.872 [1.136, 10.071] |
| CHARLS intersection; first positive | Low | 932 / 18 | 2.007 [1.199, 2.880] |
| CHARLS intersection; first positive | Middle | 1178 / 59 | 5.196 [3.848, 6.718] |
| CHARLS intersection; first positive | High | 85 / 4 | 4.872 [1.136, 10.071] |
| ELSA stroke; midpoint | Low | 298 / 2 | 0.756 [0.000, 2.095] |
| ELSA stroke; midpoint | Middle | 811 / 8 | 1.253 [0.474, 2.113] |
| ELSA stroke; midpoint | High | 184 / 6 | 3.970 [1.242, 7.293] |
| ELSA stroke; first positive | Low | 298 / 1 | 0.362 [0.000, 1.172] |
| ELSA stroke; first positive | Middle | 811 / 2 | 0.249 [0.000, 0.636] |
| ELSA stroke; first positive | High | 184 / 2 | 1.189 [0.000, 3.010] |
| ELSA MI; midpoint | Low | 298 / 2 | 0.699 [0.000, 1.826] |
| ELSA MI; midpoint | Middle | 811 / 6 | 0.758 [0.245, 1.380] |
| ELSA MI; midpoint | High | 184 / 3 | 2.012 [0.000, 4.514] |
| ELSA MI; first positive | Low | 298 / 2 | 0.737 [0.000, 1.927] |

| Population and timing | SAPA group | n / assigned events | Kaplan–Meier net probability, % [95% interval] |
|---|---|---|---|
| ELSA MI; first positive | Middle | 811 / 6 | 0.778 [0.250, 1.420] |
| ELSA MI; first positive | High | 184 / 1 | 0.588 [0.000, 1.875] |

| Population and timing | Direct group comparison | Difference, percentage points [95% interval] |
|---|---|---|
| CHARLS original; midpoint | High minus Low | 6.791 [3.164, 10.517] |
| CHARLS original; midpoint | High minus other | 4.752 [1.307, 8.491] |
| CHARLS original; midpoint | Low minus other | -3.670 [-4.942, -2.356] |
| CHARLS original; first positive | High minus Low | 6.787 [3.163, 10.517] |
| CHARLS original; first positive | High minus other | 4.747 [1.304, 8.484] |
| CHARLS original; first positive | Low minus other | -3.673 [-4.948, -2.353] |
| CHARLS intersection; midpoint | High minus Low | 2.865 [-1.068, 8.026] |
| CHARLS intersection; midpoint | High minus other | 1.087 [-2.893, 6.367] |
| CHARLS intersection; midpoint | Low minus other | -3.165 [-4.720, -1.750] |
| CHARLS intersection; first positive | High minus Low | 2.865 [-1.068, 8.026] |
| CHARLS intersection; first positive | High minus other | 1.085 [-2.893, 6.365] |
| CHARLS intersection; first positive | Low minus other | -3.168 [-4.723, -1.753] |
| ELSA stroke; midpoint | High minus Low | 3.214 [0.074, 6.565] |
| ELSA stroke; midpoint | High minus other | 2.861 [-0.098, 6.154] |
| ELSA stroke; midpoint | Low minus other | -1.010 [-2.307, 0.490] |
| ELSA stroke; first positive | High minus Low | 0.827 [-0.714, 2.773] |
| ELSA stroke; first positive | High minus other | 0.910 [-0.381, 2.740] |
| ELSA stroke; first positive | Low minus other | -0.062 [-0.748, 0.798] |
| ELSA MI; midpoint | High minus Low | 1.313 [-0.774, 3.961] |
| ELSA MI; midpoint | High minus other | 1.270 [-0.735, 3.819] |
| ELSA MI; midpoint | Low minus other | -0.298 [-1.408, 1.010] |
| ELSA MI; first positive | High minus Low | -0.149 [-1.594, 1.461] |
| ELSA MI; first positive | High minus other | -0.179 [-1.149, 1.262] |
| ELSA MI; first positive | Low minus other | -0.006 [-1.046, 1.302] |

Intervals are pointwise percentiles from 1,000 paired cluster resamples. Among consistently low-risk CHARLS participants, the high-SAPA group had 85 participants and four reports. Its point estimate was slightly below that of the middle group. The interval for high SAPA versus the remainder included zero, and the interval for low SAPA versus the remainder excluded zero.

D. Post-landmark matched prediction comparisons

| Population and timing | Model | AUC [95% interval] | Brier [95% interval] | Paired AUC gain | Paired Brier reduction |
|---|---|---|---|---|---|
| CHARLS original; midpoint | Flexible equation + background | 0.614 [0.541, 0.655] | 0.046421 [0.038910, 0.054225] | Reference | Reference |
| CHARLS original; midpoint | + SAPA | 0.645 [0.577, 0.682] | 0.046161 [0.038633, 0.053866] | 0.031 [0.006, 0.063] | 0.000260 [0.000033, 0.000484] |
| CHARLS original; midpoint | + PC1 | 0.643 [0.575, 0.681] | 0.046176 [0.038617, 0.053885] | 0.029 [0.006, 0.061] | 0.000245 [0.000034, 0.000479] |
| CHARLS original; midpoint | + equal weights | 0.644 [0.580, 0.680] | 0.046184 [0.038676, 0.053898] | 0.030 [0.007, 0.061] | 0.000238 [0.000034, 0.000450] |
| CHARLS original; first positive | Flexible equation + background | 0.614 [0.541, 0.655] | 0.046439 [0.038902, 0.054236] | Reference | Reference |
| CHARLS original; first positive | + SAPA | 0.645 [0.577, 0.678] | 0.046180 [0.038619, 0.053885] | 0.031 [0.006, 0.063] | 0.000259 [0.000032, 0.000481] |
| CHARLS original; first positive | + PC1 | 0.643 [0.575, 0.678] | 0.046196 [0.038605, 0.053901] | 0.029 [0.006, 0.061] | 0.000244 [0.000032, 0.000478] |
| CHARLS original; first positive | + equal weights | 0.644 [0.579, 0.677] | 0.046202 [0.038659, 0.053913] | 0.030 [0.007, 0.061] | 0.000237 [0.000035, 0.000450] |
| CHARLS intersection; midpoint | Flexible equation + background | 0.589 [0.502, 0.638] | 0.036715 [0.028050, 0.044431] | Reference | Reference |
| CHARLS intersection; midpoint | + SAPA | 0.642 [0.542, 0.685] | 0.036533 [0.027898, 0.044257] | 0.054 [0.015, 0.097] | 0.000183 [0.000030, 0.000377] |
| CHARLS intersection; midpoint | + PC1 | 0.645 [0.546, 0.689] | 0.036529 [0.027878, 0.044227] | 0.056 [0.017, 0.100] | 0.000186 [0.000028, 0.000378] |
| CHARLS intersection; midpoint | + equal weights | 0.651 [0.555, 0.695] | 0.036515 [0.027875, 0.044214] | 0.062 [0.025, 0.107] | 0.000200 [0.000044, 0.000407] |
| CHARLS intersection; first positive | Flexible equation + background | 0.589 [0.502, 0.638] | 0.036717 [0.028043, 0.044427] | Reference | Reference |
| CHARLS intersection; first positive | + SAPA | 0.642 [0.541, 0.685] | 0.036535 [0.027888, 0.044254] | 0.053 [0.014, 0.097] | 0.000182 [0.000031, 0.000374] |
| CHARLS intersection; first positive | + PC1 | 0.645 [0.544, 0.689] | 0.036532 [0.027885, 0.044224] | 0.056 [0.016, 0.100] | 0.000185 [0.000027, 0.000375] |
| CHARLS intersection; first positive | + equal weights | 0.651 [0.555, 0.695] | 0.036518 [0.027872, 0.044211] | 0.062 [0.025, 0.107] | 0.000199 [0.000044, 0.000406] |
| ELSA stroke; midpoint | Flexible equation + background | 0.536 [0.255, 0.667] | 0.015007 [0.007664, 0.023128] | Reference | Reference |

| Population and timing | Model | AUC [95% interval] | Brier [95% interval] | Paired AUC gain | Paired Brier reduction |
|---|---|---|---|---|---|
| ELSA stroke; midpoint | + SAPA | 0.607 [0.271, 0.732] | 0.014977 [0.007664, 0.023057] | 0.071 [-0.124, 0.234] | 0.000030 [-0.000185, 0.000238] |
| ELSA stroke; midpoint | + PC1 | 0.606 [0.268, 0.737] | 0.014975 [0.007663, 0.023077] | 0.070 [-0.122, 0.235] | 0.000032 [-0.000174, 0.000328] |
| ELSA stroke; midpoint | + equal weights | 0.608 [0.269, 0.733] | 0.014970 [0.007660, 0.022970] | 0.072 [-0.122, 0.238] | 0.000037 [-0.000169, 0.000301] |
| ELSA stroke; first positive | Flexible equation + background | 0.519; interval not qualified | 0.004093; interval not qualified | Reference | Reference |
| ELSA stroke; first positive | + SAPA | 0.487; interval not qualified | 0.004098; interval not qualified | -0.032; interval not qualified | -0.000004; interval not qualified |
| ELSA stroke; first positive | + PC1 | 0.484; interval not qualified | 0.004098; interval not qualified | -0.035; interval not qualified | -0.000004; interval not qualified |
| ELSA stroke; first positive | + equal weights | 0.500; interval not qualified | 0.004097; interval not qualified | -0.019; interval not qualified | -0.000003; interval not qualified |
| ELSA MI; midpoint | Flexible equation + background | 0.633 [0.375, 0.767] | 0.009225 [0.003367, 0.014143] | Reference | Reference |
| ELSA MI; midpoint | + SAPA | 0.601 [0.317, 0.760] | 0.009251 [0.003377, 0.014214] | -0.032 [-0.099, 0.088] | -0.000026 [-0.000168, 0.000009] |
| ELSA MI; midpoint | + PC1 | 0.605 [0.321, 0.762] | 0.009252 [0.003377, 0.014218] | -0.028 [-0.104, 0.094] | -0.000027 [-0.000159, 0.000020] |
| ELSA MI; midpoint | + equal weights | 0.616 [0.314, 0.783] | 0.009247 [0.003378, 0.014223] | -0.017 [-0.097, 0.106] | -0.000023 [-0.000133, 0.000021] |
| ELSA MI; first positive | Flexible equation + background | 0.592; interval not qualified | 0.007397; interval not qualified | Reference | Reference |
| ELSA MI; first positive | + SAPA | 0.593; interval not qualified | 0.007397; interval not qualified | 0.001; interval not qualified | -0.000001; interval not qualified |
| ELSA MI; first positive | + PC1 | 0.591; interval not qualified | 0.007397; interval not qualified | -0.0008; interval not qualified | -0.000001; interval not qualified |
| ELSA MI; first positive | + equal weights | 0.590; interval not qualified | 0.007398; interval not qualified | -0.002; interval not qualified | -0.000002; interval not qualified |

All four models use the same training participants, cluster folds and censoring estimates. Brier reduction equals baseline error minus extended-model error. Qualified intervals exclude any replicate with a fitting warning or a nonfinite result for any model. Unqualified point estimates are retained as descriptive outputs. Pairwise SAPA-versus-PC1/equal-weight superiority was not tested in this comparison.

### E. Numerical qualification of post-landmark prediction intervals

| Population and timing | Attempted refits | Finite refits | Refits with warnings | Clean finite refits | Qualified interval |
|---|---|---|---|---|---|
| CHARLS original; midpoint | 200 | 200 | 3 | 197 | Yes |
| CHARLS original; first positive | 200 | 200 | 6 | 194 | Yes |
| CHARLS intersection; midpoint | 200 | 200 | 1 | 199 | Yes |
| CHARLS intersection; first positive | 200 | 200 | 1 | 199 | Yes |
| ELSA stroke; midpoint | 200 | 200 | 5 | 195 | Yes |
| ELSA stroke; first positive | 200 | 183 | 122 | 77 | No |
| ELSA MI; midpoint | 200 | 200 | 8 | 192 | Yes |
| ELSA MI; first positive | 200 | 196 | 70 | 130 | No |

The qualification rule was added after fitting warnings were observed and applies equally to all models and settings. An interval requires at least 180 of 200 clean finite refits. ELSA first-positive stroke and MI had only 77 and 130, so their resampling intervals are withheld; finite point estimates remain descriptive.

Source Data: flow, continuous associations, groups, group intervals, qualified predictions, qualification, horizon support, proportional hazards. ROC coordinates.

### Supplementary Table S6 | Anatomical associations and matched prediction comparisons

The numerical summaries below reproduce estimates. Their populations, conditioning sets and uncertainty procedures differ by task, as specified in Supplementary Note 5.

#### Same-person DXA associations

| Region | Partial correlation [95% CI] |
|---|---|
| Abdominal VAT | 0.335 [0.267, 0.399] |
| Abdominal SAT | 0.065 [0.006, 0.125] |
| Trunk fat | 0.354 [0.289, 0.415] |
| Whole-leg fat | -0.371 [-0.430, -0.309] |
| Gynoid fat | -0.307 [-0.354, -0.259] |

n=1,876, ages 20–59; total fat and background adjusted; survey-aware intervals. With the BMI term removed from SAPA and BMI added to the conditioning set, the saved VAT partial correlation was 0.311 (95% CI, 0.246 to 0.373; n=1,876). This diagnostic uses the same survey jackknife procedure with 30 degrees of freedom. Full-precision diagnostic.

Full-precision values

#### Same-panel partial information for VAT

All 1,876 participants contribute to each row. The condition set includes age and its square, sex, race/ethnicity, education, cycle, total fat and its square. The first two rows use residual VAT variance as their denominator; the last divides the SAPA partial $R^2$ by the Full8 partial $R^2$.

| Quantity | Estimate | 95% interval |
|---|---|---|
| Full8 partial $R^2$ | 0.132 | 0.083 to 0.180 |
| SAPA partial $R^2$ | 0.112 | 0.068 to 0.156 |
| SAPA / Full8 partial $R^2$ | 0.849 | 0.738 to 0.960 |

Thus, the 84.9% fraction has a 95% interval of 73.8% to 96.0%. The interval comes from the ratio recomputed within each of 60 survey jackknife replicates, with a t30 reference. It is a same-sample partial linear summary; cross-cycle prediction is reported separately. Full partial-information output.

#### MRI genetic associations

| Region | Partial genetic correlation [95% CI] |
|---|---|
| Abdominal VAT | 0.459 [0.365, 0.553] |
| Abdominal SAT | 0.357 [0.188, 0.526] |
| Liver fat | 0.541 [0.434, 0.648] |
| Thigh SAT | -0.144 [-0.240, -0.049] |

Fixed genetic projection T conditional on whole-body fat. UK Biobank sources overlap; intervals use the original delta-method error.

Full-precision values

#### VAT-minus-thigh-SAT vascular contrasts

| Endpoint | Difference in partial genetic correlations [95% CI] |
|---|---|
| MVP: MI | 0.308 [0.169, 0.447] |
| MVP: stroke | 0.366 [0.154, 0.577] |
| FinnGen: MI | 0.354 [0.198, 0.509] |
| FinnGen: I9_STR | 0.045 [-0.139, 0.228] |

Normal intervals retain covariance between the two depot estimates. Source and stroke definitions remain separate.

Full-precision values

### Matched construct comparisons

| Comparison population | Target and model | MSE improvement, % [95% CI] |
|---|---|---|
| MetS (n=1,465) | VAT / SAPA | 9.853 [4.801, 14.904] |
| MetS (n=1,465) | VAT / MetS | 9.475 [4.501, 14.448] |
| MetS (n=1,465) | Leg fat / SAPA | 13.050 [7.851, 18.248] |
| MetS (n=1,465) | Leg fat / MetS | 13.426 [8.842, 18.010] |
| PhenoAge (n=1,867) | VAT / SAPA | 10.440 [5.583, 15.297] |
| PhenoAge (n=1,867) | VAT / PhenoAge | -0.461 [-2.975, 2.054] |
| PhenoAge (n=1,867) | Leg fat / SAPA | 13.569 [7.772, 19.366] |
| PhenoAge (n=1,867) | Leg fat / PhenoAge | 1.015 [-2.016, 4.046] |
| Waist (n=1,465) | VAT / Waist | 13.501 [7.877, 19.124] |
| Waist (n=1,465) | VAT / Waist + SAPA | 19.020 [12.387, 25.652] |
| Waist (n=1,465) | Leg fat / Waist | 25.826 [18.762, 32.890] |
| Waist (n=1,465) | Leg fat / Waist + SAPA | 32.224 [25.964, 38.483] |

Reciprocal-cycle prediction with total fat and background variables. Improvements use the respective background MSE; population and denominator are matched within each comparison.

Full-precision values

### Direct MetS–SAPA comparison in matched participants

The two scores were compared within the same 1,465 participants, using the same background and total-fat terms and exchanging cycles for training and testing. Positive differences favour MetS. Units are percentage points of MSE improvement relative to the common background model.

| Target | MetS minus SAPA | Paired 95% interval | Holm-adjusted P (two-target family) |
|---|---|---|---|
| VAT | -0.378 | -3.112 to 2.356 | 1.000 |
| Whole-leg fat | 0.376 | -2.948 to 3.701 | 1.000 |

Intervals use 60 paired survey jackknife refits and a t30 reference. The study specified no equivalence margin. Complete matched results.

### Mutual increments of SAPA and continuous metabolic syndrome severity

| Target | Addition | MSE improvement, percentage points | Paired 95% interval | Holm-adjusted P (four-comparison family) |
|---|---|---|---|---|
| VAT | SAPA added to MetS | 0.836 | -1.225 to 2.896 | 0.828 |
| VAT | MetS added to SAPA | 0.458 | -1.119 to 2.034 | 0.828 |
| Whole-leg fat | SAPA added to MetS | 1.198 | -0.510 to 2.906 | 0.487 |
| Whole-leg fat | MetS added to SAPA | 1.574 | -0.389 to 3.537 | 0.447 |

Each row compares the two-score model with the model containing the named starting score, using the same 1,465 participants, total-fat terms and background covariates. Improvements are expressed relative to the common background MSE. Intervals use 60 paired survey jackknife refits and a t30 reference; Holm adjustment covers the four additions. These comparisons do not establish an increment for either score or equivalence between scores.

### SAPA added after waist and total fat

| Target and conditioning | Paired MSE improvement, percentage points [95% CI] |
|---|---|
| VAT / Total fat | 5.519 [2.004, 9.035] |
| VAT / Total fat + BMI | 6.025 [2.202, 9.848] |
| Leg fat / Total fat | 6.398 [2.556, 10.239] |
| Leg fat / Total fat + BMI | 7.341 [3.464, 11.219] |

Direct paired improvements, in percentage points of background MSE. The BMI-adjusted comparison has its own denominator; n=1,465.

Full-precision values

### Imaging-free prediction: paired improvements

| Target | Comparison | RMSE reduction, g [95% CI] |
|---|---|---|
| VAT | SAPA added to body shape | 6.466 [2.805, 10.127] |
| VAT | SAPA added after lipids/glycaemia | 0.041 [-1.480, 1.563] |
| VAT | Lipids/glycaemia versus SAPA | -0.178 [-2.335, 1.980] |
| VAT | Full8 versus SAPA | -0.906 [-3.758, 1.947] |
| VAT | Full8 versus combined model | -0.770 [-2.355, 0.816] |
| Whole-leg fat | SAPA added to body shape | 23.139 [-2.064, 48.342] |
| Whole-leg fat | SAPA added after lipids/glycaemia | -2.411 [-15.361, 10.539] |
| Whole-leg fat | Lipids/glycaemia versus SAPA | 21.946 [-0.360, 44.252] |
| Whole-leg fat | Full8 versus SAPA | 11.694 [-43.340, 66.728] |
| Whole-leg fat | Full8 versus combined model | -7.841 [-42.768, 27.085] |

n=1,869; body shape includes BMI and waist. Positive values favour the added or first-named information. Sixty survey jackknife replicates refit both training directions; pointwise t30 intervals.

Full-precision values

### Imaging-free prediction: absolute errors

| Target | Model | RMSE, kg [95% CI] |
|---|---|---|
| VAT | Body shape | 0.161 [0.148, 0.175] |
| VAT | Body shape + SAPA | 0.155 [0.141, 0.168] |
| VAT | Body shape + lipids/glycaemia | 0.155 [0.142, 0.168] |
| VAT | Body shape + both | 0.155 [0.142, 0.168] |
| VAT | Body shape + Full8 | 0.156 [0.142, 0.170] |
| Whole-leg fat | Body shape | 1.988 [1.816, 2.160] |
| Whole-leg fat | Body shape + SAPA | 1.965 [1.800, 2.129] |
| Whole-leg fat | Body shape + lipids/glycaemia | 1.943 [1.786, 2.100] |
| Whole-leg fat | Body shape + both | 1.945 [1.788, 2.102] |
| Whole-leg fat | Body shape + Full8 | 1.953 [1.796, 2.110] |

These ten rows use the same imaging-free task above. Full-precision values and original row identifiers are supplied in Source Data.

### Liver-fat–disease genetic covariance components

Liver-fat genetic correlations with each disease are separated into covariance along the fixed common direction (C), covariance represented by the rest of Full8 (K), and residual covariance beyond Full8 (U_X). Values are estimates with pointwise 95%

confidence intervals on standardised genetic covariance scales. C + K + U_X equals the total correlation before rounding. No whole-body-fat adjustment is applied in this decomposition.

| Specification | Endpoint | Total | Along T (C) | Additional Full8 (K) | Beyond Full8 (U_X) |
|---|---|---|---|---|---|
| Default | MVP MI | 0.375 (0.262 to 0.487) | 0.368 (0.312 to 0.424) | −0.028 (−0.047 to −0.009) | 0.035 (−0.067 to 0.136) |
| Default | MVP survey-reported stroke | 0.311 (0.153 to 0.469) | 0.272 (0.205 to 0.338) | −0.007 (−0.032 to 0.017) | 0.047 (−0.102 to 0.196) |
| Default | FinnGen strict MI | 0.293 (0.178 to 0.407) | 0.272 (0.221 to 0.323) | −0.028 (−0.049 to −0.007) | 0.049 (−0.053 to 0.150) |
| Default | FinnGen stroke (I9_STR) | 0.214 (0.074 to 0.355) | 0.257 (0.194 to 0.321) | −0.042 (−0.072 to −0.012) | −0.001 (−0.131 to 0.130) |
| Chi-square cap 80 | MVP MI | 0.377 (0.264 to 0.490) | 0.359 (0.305 to 0.413) | −0.019 (−0.037 to −0.001) | 0.037 (−0.065 to 0.139) |
| Chi-square cap 80 | MVP survey-reported stroke | 0.311 (0.153 to 0.469) | 0.273 (0.208 to 0.337) | −0.004 (−0.027 to 0.020) | 0.042 (−0.108 to 0.192) |
| Chi-square cap 80 | FinnGen strict MI | 0.292 (0.178 to 0.406) | 0.259 (0.211 to 0.307) | −0.015 (−0.036 to 0.005) | 0.049 (−0.053 to 0.150) |
| Chi-square cap 80 | FinnGen stroke (I9_STR) | 0.214 (0.074 to 0.355) | 0.251 (0.189 to 0.312) | −0.028 (−0.052 to −0.004) | −0.008 (−0.139 to 0.123) |

Holm-adjusted P values below use all 12 component tests within each specification. Total correlations are descriptive. The negative K components were sensitive to the SNP chi-square specification, and residual intervals allow associations in either direction. Components are not causal contributions or percentages of disease risk. FinnGen I9_STR excludes subarachnoid haemorrhage; the MVP survey endpoint has no specified subtype or subarachnoid haemorrhage exclusion.

| Specification | Endpoint | C: Holm P | K: Holm P | U_X: Holm P |
|---|---|---|---|---|
| Default | MVP MI | $4.15 \times 10^{-37}$ | 0.027 | 1.000 |
| Default | MVP survey-reported stroke | $1.52 \times 10^{-14}$ | 1.000 | 1.000 |
| Default | FinnGen strict MI | $2.67 \times 10^{-24}$ | 0.0497 | 1.000 |
| Default | FinnGen stroke (I9_STR) | $1.78 \times 10^{-14}$ | 0.040 | 1.000 |
| Chi-square cap 80 | MVP MI | $2.22 \times 10^{-37}$ | 0.304 | 1.000 |
| Chi-square cap 80 | MVP survey-reported stroke | $9.07 \times 10^{-16}$ | 1.000 | 1.000 |
| Chi-square cap 80 | FinnGen strict MI | $2.57 \times 10^{-25}$ | 0.816 | 1.000 |
| Chi-square cap 80 | FinnGen stroke (I9_STR) | $9.82 \times 10^{-15}$ | 0.162 | 1.000 |

Full-precision liver-fat components and uncertainty. Supplementary Note 5 gives the projection, joint-error calculations and specification definitions.

### Supplementary Table S7 | Longitudinal waist change and the shared marker profile

All estimates reuse the completed weighted paired-visit analysis. Parentheses give pointwise 95% percentile intervals from 999 household bootstrap resamples. Scalar units are fixed Q units per 10 cm of waist, without within-sample SD scaling. Mean and change regressions include baseline age, sex, smoking and visit interval. BMI-conditioned versions additionally include the corresponding mean or change in BMI. The same bootstrap draws support direct within-minus-between contrasts.

#### A. Main and overlapping-window populations

| Population | Participants | Households | Median interval, years |
|---|---|---|---|
| ELSA W4 to W6 | 2959 | 2517 | 3.917 |
| ELSA W2 to W4 | 2246 | 1884 | 3.917 |
| ELSA three visits: W2 to W4 | 1416 | 1236 | 3.917 |
| ELSA three visits: W4 to W6 | 1416 | 1236 | 3.833 |
| CHARLS, 40–200 cm at both visits | 5207 | 3901 | 4.000 |
| CHARLS, all positive waists | 5404 | 4045 | 4.000 |

ELSA intervals use nurse-visit months; CHARLS uses interview months because individual blood dates were unavailable. The 1,416-person ELSA windows share wave 4 and wave-6 weights. The CHARLS 40–200 cm range was fixed before fitting and is an investigator-defined review range.

#### B. Between-person and within-person associations

| Population | Outcome | BMI conditioned | Between | Within | Within − between |
|---|---|---|---|---|---|
| ELSA W4 to W6 | Complete Q | No | 0.678 (0.641 to 0.718) | 0.450 (0.344 to 0.552) | −0.228 (−0.326 to −0.125) |
| ELSA W4 to W6 | Q without BMI | No | 0.455 (0.420 to 0.493) | 0.351 (0.264 to 0.437) | −0.104 (−0.193 to −0.017) |
| ELSA W4 to W6 | Two blood pressures | No | 0.080 (0.065 to 0.096) | 0.075 (0.030 to 0.122) | −0.005 (−0.049 to 0.042) |
| ELSA W4 to W6 | Other five markers | No | 0.375 (0.344 to 0.405) | 0.276 (0.217 to 0.340) | −0.099 (−0.159 to −0.034) |
| ELSA W4 to W6 | Q without BMI | Yes | 0.341 (0.263 to 0.415) | 0.217 (0.128 to 0.312) | −0.124 (−0.223 to −0.019) |
| ELSA W4 to W6 | Two blood pressures | Yes | 0.035 (0.004 to 0.067) | 0.035 (−0.011 to 0.087) | 0.000 (−0.050 to 0.058) |
| ELSA W4 to W6 | Other five markers | Yes | 0.305 (0.238 to 0.370) | 0.181 (0.113 to 0.250) | −0.124 (−0.218 to −0.040) |
| CHARLS, 40–200 cm at both visits | Complete Q | No | 0.841 (0.804 to 0.875) | 0.275 (0.207 to 0.340) | −0.566 (−0.641 to −0.488) |
| CHARLS, 40–200 cm at both visits | Q without BMI | No | 0.646 (0.611 to 0.678) | 0.198 (0.132 to 0.264) | −0.448 (−0.520 to −0.372) |
| CHARLS, 40–200 cm at both visits | Two blood pressures | No | 0.172 (0.152 to 0.190) | 0.031 (−0.005 to 0.068) | −0.140 (−0.181 to −0.099) |
| CHARLS, 40–200 cm at both visits | Other five markers | No | 0.474 (0.443 to 0.505) | 0.167 (0.112 to 0.219) | −0.307 (−0.367 to −0.247) |
| CHARLS, 40–200 cm at both visits | Q without BMI | Yes | 0.541 (0.473 to 0.614) | 0.171 (0.105 to 0.234) | −0.370 (−0.469 to −0.274) |
| CHARLS, 40–200 cm at both visits | Two blood pressures | Yes | 0.128 (0.085 to 0.165) | 0.013 (−0.022 to 0.048) | −0.115 (−0.162 to −0.059) |

| Population | Outcome | BMI conditioned | Between | Within | Within − between |
|---|---|---|---|---|---|
| CHARLS, 40–200 cm at both visits | Other five markers | Yes | 0.413 (0.351 to 0.475) | 0.158 (0.103 to 0.210) | −0.255 (−0.346 to −0.166) |

In the BMI-conditioned models, complete Q and Q without BMI give the same coefficient by construction. The other-five sum comprises LDL, HDL, log TG, HbA1c and log CRP. Summing component coefficients gives the total coefficient; interval endpoints are not additive.

### C. Constituent-marker within-person associations

| Marker | ELSA, no BMI condition | ELSA, BMI conditioned | CHARLS, no BMI condition | CHARLS, BMI conditioned |
|---|---|---|---|---|
| SBP | 0.090 (0.022 to 0.160) | 0.024 (−0.045 to 0.098) | 0.056 (−0.008 to 0.116) | 0.030 (−0.028 to 0.087) |
| DBP | 0.137 (0.068 to 0.211) | 0.091 (0.013 to 0.180) | 0.032 (−0.015 to 0.082) | 0.004 (−0.045 to 0.055) |
| LDL | 0.059 (−0.001 to 0.118) | 0.018 (−0.043 to 0.075) | 0.049 (0.005 to 0.093) | 0.036 (−0.008 to 0.080) |
| HDL | −0.080 (−0.117 to −0.041) | −0.052 (−0.091 to −0.002) | −0.088 (−0.124 to −0.049) | −0.077 (−0.111 to −0.038) |
| lnTG | 0.212 (0.164 to 0.266) | 0.155 (0.099 to 0.209) | 0.146 (0.103 to 0.189) | 0.139 (0.093 to 0.185) |
| HbA1c | 0.064 (0.034 to 0.096) | 0.043 (0.014 to 0.074) | −0.066 (−0.140 to −0.006) | −0.040 (−0.109 to 0.018) |
| BMI | 0.221 (0.165 to 0.269) | 0.000 (0.000 to 0.000) | 0.171 (0.144 to 0.197) | 0.000 (0.000 to 0.000) |
| lnCRP | 0.143 (0.102 to 0.191) | 0.087 (0.029 to 0.142) | 0.118 (0.075 to 0.163) | 0.103 (0.057 to 0.147) |

Units are fixed NHANES-reference marker SD per 10 cm. HDL is shown in its measured direction. The BMI coefficient is zero by construction when BMI itself is included as a covariate.

### D. Planned population and measurement checks

| Comparison | n | Q without BMI | Q without BMI, BMI conditioned |
|---|---|---|---|
| ELSA primary | 2959 | 0.351 (0.264 to 0.437) | 0.217 (0.128 to 0.312) |
| ELSA medication complete, no medication terms | 2913 | 0.350 (0.261 to 0.438) | 0.219 (0.130 to 0.316) |
| ELSA medication complete, medication terms | 2913 | 0.362 (0.273 to 0.448) | 0.227 (0.139 to 0.324) |
| ELSA unchanged medications | 2338 | 0.326 (0.231 to 0.428) | 0.201 (0.110 to 0.303) |
| ELSA fasting at both visits | 1623 | 0.407 (0.321 to 0.493) | 0.263 (0.094 to 0.367) |
| ELSA directly measured weight | 2951 | 0.351 (0.263 to 0.438) | 0.216 (0.127 to 0.313) |
| ELSA earlier window | 2246 | 0.466 (0.384 to 0.543) | 0.273 (0.116 to 0.373) |
| ELSA same 1,416: earlier window | 1416 | 0.424 (0.331 to 0.519) | 0.242 (0.023 to 0.368) |
| ELSA same 1,416: later window | 1416 | 0.438 (0.339 to 0.530) | 0.308 (0.153 to 0.418) |
| CHARLS 40–200 cm | 5207 | 0.198 (0.132 to 0.264) | 0.171 (0.105 to 0.234) |
| CHARLS all positive values | 5404 | 0.034 (0.007 to 0.063) | 0.0261 (−0.0004 to 0.0552) |
| CHARLS medication complete, no medication terms | 4577 | 0.200 (0.129 to 0.272) | 0.173 (0.101 to 0.245) |
| CHARLS medication complete, medication terms | 4577 | 0.209 (0.147 to 0.274) | 0.181 (0.113 to 0.247) |
| CHARLS unchanged medications | 3710 | 0.212 (0.147 to 0.276) | 0.174 (0.110 to 0.237) |
| CHARLS source codes = 1 at both visits | 4018 | 0.173 (0.091 to 0.250) | 0.138 (0.063 to 0.213) |
| CHARLS no TG top-coding | 5137 | 0.198 (0.131 to 0.266) | 0.171 (0.103 to 0.235) |

For CHARLS all-positive waists, the BMI-conditioned estimate at four decimals is 0.0261 (−0.0004 to 0.0552). The ELSA fasting subset uses the recorded eligibility indicator. The CHARLS subset uses source codes equal to 1 at both visits; its wave-1 fasting meaning remains unverified. Medication groups are defined by three recorded treatment classes. The comparison did not match participants on dose or adherence.

### E. Direct paired changes in the within-person coefficient

| Contrast | n | BMI conditioned | Difference in Q without BMI coefficient |
|---|---|---|---|
| ELSA: medication adjusted − unadjusted | 2913 | No | 0.012 (−0.005 to 0.030) |
| ELSA: medication adjusted − unadjusted | 2913 | Yes | 0.008 (−0.011 to 0.026) |
| CHARLS: medication adjusted − unadjusted | 4577 | No | 0.009 (−0.002 to 0.030) |
| CHARLS: medication adjusted − unadjusted | 4577 | Yes | 0.008 (−0.004 to 0.029) |
| ELSA same people: later − earlier window | 1416 | No | 0.013 (−0.109 to 0.144) |
| ELSA same people: later − earlier window | 1416 | Yes | 0.066 (−0.066 to 0.238) |

### F. Influence and range diagnostics

| ELSA specification | Full estimate | Household-deletion minimum | Maximum | Largest absolute change |
|---|---|---|---|---|
| ELSA, without BMI conditioning | 0.351 | 0.347 | 0.391 | 0.040 |
| ELSA, with BMI conditioning | 0.217 | 0.186 | 0.252 | 0.035 |

CHARLS: 197 of 5,404 participants had at least one waist below 40 cm. Their sampling-weight share was 3.139%; their share of weighted residual waist-change sum of squares was 72.500% without BMI conditioning and 73.940% with it. Observed changes in these records ranged from −70.9 to +80.0 cm. No presumed unit conversion was applied. These are leverage diagnostics, not confirmations that particular records are erroneous.

### G. Direction diagnostic

| Population | BMI conditioned | Squared cosine of within-person association vector |
|---|---|---|
| ELSA W4 to W6 | No | 0.867 (0.766 to 0.905) |
| ELSA W4 to W6 | Yes | 0.690 (0.408 to 0.758) |
| CHARLS, 40–200 cm at both visits | No | 0.603 (0.352 to 0.777) |
| CHARLS, 40–200 cm at both visits | Yes | 0.491 (0.198 to 0.695) |

Squared cosine describes vector orientation on the fixed D marker scale. It is not explained individual change, disease information or causal mediation. Full between-person diagnostics and all saved marker/component estimates are supplied in Source Data.

**Source Data:** Body Marker Estimates, Paired Contrasts, Sample Support, Vector Alignment, Family Influence, CHARLS Range Leverage, Weight Identity, Design Matrices.

Supplementary Table S8 | Biochemical characterization, fasting insulin and IL-1RA

A. Initial six-marker screen

| Marker | n | $R^2$ gain, percentage points | Lower limit | Upper limit | No-CRP score gain after CRP and BMI | Lower limit | Upper limit |
|---|---|---|---|---|---|---|---|
| IL-6 | 3464 | 6.926 | 5.250 | 8.629 | -0.060 | -0.812 | 0.377 |
| IL-10 | 3464 | 2.246 | 1.166 | 3.411 | 0.132 | -0.546 | 0.624 |
| IL-1RA | 3464 | 15.122 | 12.391 | 17.333 | 2.408 | 1.289 | 3.376 |
| sTNFR1 | 3464 | 4.382 | 3.170 | 5.787 | 0.557 | -0.121 | 1.209 |
| Activated TGF-beta1 | 3464 | 1.090 | 0.310 | 1.834 | 0.871 | 0.149 | 1.586 |
| NT-proBNP | 3406 | 0.109 | -0.276 | 0.4 | 0.231 | -0.3 | 0.678 |

Units: held-out $R^2$ increment in percentage points, with original marginal intervals. The final three columns add the fixed no-CRP score to a background including CRP and BMI. Screening-stage scoring and same-resource selection are retained.

### B. NHANES-reference IL-1RA models

The same HRS follow-up sample contains 3,464 participants from 2,680 families. All models use log IL-1RA, a common background, five family folds and 200 family resamples. Values below are whole-model $R^2$, not the increments in section A.

| Model | Held-out $R^2$, % [95% interval] |
|---|---|
| Background | 0.643 [0.032, 2.086] |
| SAPA | 16.452 [14.061, 19.630] |
| PC1 | 16.914 [14.434, 20.152] |
| Equal weights | 14.521 [12.283, 17.622] |

| Paired comparison | MSE improvement, $10^{-3}$ log-concentration² [95% basic interval] | Holm-adjusted P (five comparisons) |
|---|---|---|
| SAPA age interaction versus no interaction | −0.015 [−0.789, 0.923] | 1.000 |
| PC1 age interaction versus no interaction | 0.001 [−0.794, 0.947] | 1.000 |
| Equal-weight age interaction versus no interaction | 0.193 [−0.674, 1.214] | 1.000 |
| SAPA versus PC1, no age interaction | −1.115 [−1.724, −0.489] | 0.004 |
| SAPA versus equal weights, no age interaction | 4.662 [3.534, 5.583] | $6.29 \times 10^{-20}$ |

Positive paired MSE improvements favour the first-named model. $R^2$ intervals are marginal bootstrap intervals; MSE intervals are marginal basic-bootstrap intervals. Holm-adjusted P values control the prespecified five-comparison family. Model $R^2$ and main paired estimates; all direct comparisons.

### C. Age-specific IL-1RA associations and age transfer

| Score | Age, years | Geometric-mean ratio per reference SD [95% interval] |
|---|---|---|
| SAPA | 60 | 1.321 [1.275, 1.371] |
| SAPA | 80 | 1.265 [1.209, 1.314] |
| PC1 | 60 | 1.311 [1.265, 1.364] |
| PC1 | 80 | 1.254 [1.198, 1.305] |
| Equal weights | 60 | 1.302 [1.252, 1.359] |
| Equal weights | 80 | 1.233 [1.178, 1.281] |

| Direct age contrast | Log-ratio difference [95% marginal interval] | Holm-adjusted P (three comparisons) |
|---|---|---|
| SAPA, age 80 minus 60 | −0.043 [−0.084, −0.002] | 0.082 |
| SAPA minus PC1 in the age difference | 0.001 [−0.003, 0.005] | 0.587 |
| SAPA minus equal weights in the age difference | 0.011 [0.006, 0.017] | 0.002 |

The separate age-transfer task trained at ages 55–69 (n=1,757) and evaluated ages 70–84 (n=1,287). Target-test families were excluded from source fitting and target calibration. T0 carried the younger model over; T1 updated target background terms; T2 also updated score strength. The 200 family resamples repeated both fitting stages.

| Age-transfer comparison | MAE improvement, $10^{-3}$ log-concentration [interval] | Interval definition |
|---|---|---|
| Score-strength update: MAE(T1) − MAE(T2) | −0.157 [−2.802, 1.625] | 95% basic bootstrap (primary) |
| Background update: MAE(T0) − MAE(T1) | −1.756 [−13.815, 10.304] | Three-comparison normal-Bonferroni family interval |
| SAPA versus PC1 after background calibration | −0.769 [−2.283, 0.745] | Three-comparison normal-Bonferroni family interval |
| SAPA versus equal weights after background calibration | 4.869 [2.684, 7.053] | Three-comparison normal-Bonferroni family interval |

Positive MAE improvements favour the update or SAPA. These transfer intervals have different definitions from the marginal age-association intervals above. Age-specific effects; four transfer comparisons. Fig. 3f,g and Supplementary Figure 3 show these distinct evaluations; Supplementary Note 6 describes the models.

### D. Age structure in the HRS IL-1RA source population

All 3,044 participants aged 55–84 from the 3,464-person IL-1RA source are included. Background regression uses the full source, followed by age-band residual covariance with equal weighting of sexes. Estimates are cross-sectional linear changes per decade across six age-band midpoints. F is multiplied by 100; other quantities retain their fixed-reference or correlation units.

| Direction | Quantity | Change per decade | Simultaneous 95% interval | Holm-adjusted P (12 trends) |
|---|---|---|---|---|
| SAPA | Aligned variation P | -0.1138 | [-0.2317, 0.0041] | 0.198 |
| PC1 | Aligned variation P | -0.1127 | [-0.2346, 0.0092] | 0.238 |
| Equal weights | Aligned variation P | -0.1078 | [-0.2322, 0.0166] | 0.314 |

| Direction | Quantity | Change per decade | Simultaneous 95% interval | Holm-adjusted P (12 trends) |
|---|---|---|---|---|
| SAPA | Remaining variation O | -0.0600 | [-0.3160, 0.1961] | 1.000 |
| PC1 | Remaining variation O | -0.0611 | [-0.3189, 0.1967] | 1.000 |
| Equal weights | Remaining variation O | -0.0660 | [-0.3163, 0.1844] | 1.000 |
| SAPA | Aligned share F (percentage points) | -1.1295 | [-2.5884, 0.3295] | 0.491 |
| PC1 | Aligned share F (percentage points) | -1.0857 | [-2.6207, 0.4493] | 0.491 |
| Equal weights | Aligned share F (percentage points) | -1.0053 | [-2.5065, 0.4959] | 0.491 |
| SAPA | Off-diagonal correlation contribution | -0.0748 | [-0.1916, 0.0421] | 0.491 |
| PC1 | Off-diagonal correlation contribution | -0.0894 | [-0.2098, 0.0310] | 0.491 |
| Equal weights | Off-diagonal correlation contribution | -0.0916 | [-0.2118, 0.0287] | 0.491 |

Age-band counts (55–59 through 80–84 years): 535, 670, 552, 415, 518, 354.

Intervals use 200 paired family resamples and an approximate maximum-standardized-error calibration across the complete 12-trend family. Two-sided normal-approximation P values use the bootstrap standard error and Holm correction. The simultaneous intervals allow a range of age trends. See the full-precision trends and age/sex group counts.

### E. IL-1RA prediction beyond BMI and CRP using the NHANES reference

The original 3,464 people, 2,680 families and five family folds were retained. BMI and log CRP were entered separately before the remaining six-marker terms. All values below concern log IL-1RA in this sample. The comparison was specified before its new fits, after the earlier IL-1RA findings were known.

| Model | Number of fitted coefficients (including intercept) | Whole-model $R^2$, % [95% interval] | MSE ×1,000 [95% interval] | $R^2$ gain versus BMI/CRP background, percentage points [95% interval] |
|---|---|---|---|---|
| Background | 14 | 0.643 [0.032, 2.086] | 239.827 [225.605, 253.422] | Not applicable |
| Background + BMI + log CRP | 22 | 18.224 [15.789, 21.675] | 197.390 [179.789, 209.268] | Reference |
| + six-marker SAPA projection | 26 | 21.224 [18.586, 25.050] | 190.147 [174.762, 200.877] | 3.000 [1.955, 4.190] |
| + six-marker PC1 projection | 26 | 21.042 [18.380, 24.748] | 190.586 [174.980, 201.210] | 2.819 [1.778, 3.967] |
| + six-marker equal-weight projection | 26 | 20.256 [17.394, 23.864] | 192.484 [176.852, 203.583] | 2.032 [1.106, 3.028] |
| + six markers individually | 46 | 24.632 [22.211, 29.096] | 181.922 [162.359, 193.323] | 6.408 [4.791, 8.872] |

Each added projection uses four coefficients; the individual-marker extension adds 24. All three six-marker projections remove BMI and log CRP from the original frozen directions. They use externally fixed units. PC1 uses the corresponding weights from the eight-marker solution.

| Paired comparison | MSE reduction ×1,000 [95% basic interval] | $R^2$ difference, percentage points [95% basic interval] | Holm-adjusted P, seven comparisons |
|---|---|---|---|
| SAPA six-marker projection versus BMI/CRP background | 7.242 [4.448, 10.059] | 3.000 [1.955, 4.190] | $1.15 \times 10^{-6}$ |
| PC1 six-marker projection versus BMI/CRP background | 6.804 [4.033, 9.647] | 2.819 [1.778, 3.967] | $2.79 \times 10^{-6}$ |
| Equal-weight six-marker projection versus BMI/CRP background | 4.906 [2.576, 7.316] | 2.032 [1.106, 3.028] | $1.98 \times 10^{-4}$ |
| Individual six markers versus BMI/CRP background | 15.468 [12.152, 21.608] | 6.408 [4.791, 8.872] | $4.63 \times 10^{-10}$ |
| SAPA versus PC1 six-marker projections | 0.438 [0.094, 0.728] | 0.182 [0.028, 0.305] | 0.011 |
| SAPA versus equal-weight six-marker projections | 2.336 [1.569, 2.859] | 0.968 [0.655, 1.190] | $7.18 \times 10^{-11}$ |
| Individual six markers versus SAPA six-marker projection | 8.225 [5.082, 13.515] | 3.408 [2.054, 5.588] | $2.36 \times 10^{-4}$ |

Positive differences favour the first-named model. All 200 complete family refits were finite, full rank and warning-free. Individual intervals are pointwise, and Holm-adjusted P values use bootstrap standard errors with a normal approximation. Selection of IL-1RA was not repeated, so the inference remains conditional on its earlier selection within HRS. These prediction increments do not quantify mediation or an independent biological information fraction. Different nonlinear model spaces and parameter counts also preclude interpreting the SAPA/individual-marker gain ratio as a formal information-retention fraction. Section A's screening-stage 2.408-point estimate uses a different score and model; it is not an external replication of section E.

Source Data: whole-model estimates, paired MSE contrasts, paired $R^2$ contrasts, model specification, family folds and refit qualification.

### F. Fasting insulin population and measured-DXA selection

Participants were aged 20–79 years with complete construction markers and background covariates. No diagnosed diabetes includes borderline questionnaire responses. All included participants denied insulin use; oral-treatment fields were structurally skipped. Other eligible participants had imputed or unverified DXA. Counts are unweighted; descriptive means below are survey-weighted.

| Cycle | Complete-marker DXA panel | Eligible before measured-DXA restriction | Verified measured DXA | Other eligible DXA |
|---|---|---|---|---|
| 2001-2002 | 1705 | 1580 | 1327 | 253 |
| 2003-2004 | 1537 | 1401 | 1148 | 253 |
| Combined | 3242 | 2981 | 2475 | 506 |

Weighted mean BMI was 27.009 in the analysed sample and 32.702 in the other eligible DXA group. Mean ages were 42.995 and 46.302 years, respectively. Waist was not a covariate in these models and was missing in 14 included participants. The source description retains observed denominators for every descriptive variable.

### G. Three primary comparisons and two supporting associations

| Comparison | Status | n | Estimate [95% CI] | Holm-adjusted P |
|---|---|---|---|---|
| Six-marker projection per doubling of insulin | Primary | 2475 | 0.408 [0.310, 0.506] | $3.56 \times 10^{-9}$ |
| Six-marker projection per 10% higher trunk-to-whole-leg fat ratio | Primary | 2475 | 0.103 [0.085, 0.121] | $2.02 \times 10^{-12}$ |
| CRP percentage difference per doubling of insulin | Primary | 2475 | 10.664 [-0.267, 22.793] | 0.056 |
| Insulin percentage difference per 10% higher trunk-to-whole-leg fat ratio | Supporting | 2475 | 4.698 [3.949, 5.452] | Not in primary family |

| Comparison | Status | n | Estimate [95% CI] | Holm-adjusted P |
|---|---|---|---|---|
| Six-marker projection per doubling of CRP | Supporting | 2475 | 0.059 [0.025, 0.094] | Not in primary family |

Six-marker projection estimates use the fixed weighted sum excluding BMI and CRP; they are not SD-scaled. All models adjusted for age, age squared, sex, race/ethnicity, smoking, height and cycle. The projection models included total fat, regional fat ratio, lean mass and log CRP; insulin was included when estimating the fat-ratio and CRP terms. The CRP–insulin model included body composition but no CRP covariate. The insulin–fat-ratio model included the other body composition terms. Holm adjustment covers the three primary pooled tests; all intervals are pointwise. CRP percentages were calculated after restoring the raw log-concentration scale.

### H. Eight-marker associations with fasting insulin

| Marker | Background-adjusted [95% CI] | Body-composition-adjusted [95% CI] |
|---|---|---|
| SBP | 0.165 [0.104, 0.226] | 0.082 [0.003, 0.161] |
| DBP | 0.168 [0.097, 0.239] | 0.101 [0.020, 0.183] |
| LDL | 0.144 [0.079, 0.209] | -0.020 [-0.113, 0.074] |
| HDL | -0.463 [-0.513, -0.414] | -0.280 [-0.351, -0.209] |
| lnTG | 0.480 [0.431, 0.528] | 0.339 [0.277, 0.401] |
| HbA1c | 0.119 [0.082, 0.156] | 0.074 [0.024, 0.125] |
| BMI | 0.523 [0.475, 0.571] | 0.010 [-0.002, 0.022] |
| lnCRP | 0.405 [0.334, 0.477] | 0.082 [-0.002, 0.166] |

Differences are in marker-specific NHANES 2015–2018 reference SD units per doubling of harmonised fasting insulin (n=2,475). TG and CRP were log transformed before standardisation. Body composition adjustment adds total fat, trunk-to-whole-leg fat ratio and non-bone lean mass. These descriptive marker estimates were not added to the three-test primary family. Differences between adjustment sets are not mediation fractions.

### I. Cycle-specific and specified sensitivity estimates

| Analysis | Cycle | n | Six-marker projection per insulin doubling [95% CI] |
|---|---|---|---|
| Harmonised assay | pooled | 2475 | 0.408 [0.310, 0.506] |
| Harmonised assay | 2001-2002 | 1327 | 0.414 [0.270, 0.557] |
| Harmonised assay | 2003-2004 | 1148 | 0.416 [0.263, 0.569] |
| Native assay | pooled | 2475 | 0.320 [0.241, 0.398] |
| Native assay | 2001-2002 | 1327 | 0.414 [0.270, 0.557] |
| Native assay | 2003-2004 | 1148 | 0.276 [0.175, 0.377] |
| Exclude laboratory hyperglycaemia | pooled | 2384 | 0.412 [0.317, 0.506] |
| Exclude laboratory hyperglycaemia | 2001-2002 | 1275 | 0.417 [0.276, 0.558] |
| Exclude laboratory hyperglycaemia | 2003-2004 | 1109 | 0.421 [0.271, 0.571] |

Each estimate uses body composition and CRP adjustment. The 2003–2004 harmonisation was 0.96006 × native insulin + 3.23663; the intercept changes the doubling contrast. The hyperglycaemia sensitivity excludes fasting glucose ≥126 mg/dL or HbA1c ≥6.5%. Cycles are nonoverlapping NHANES samples. All 999 resample fits completed for each analysis. The intervals quantify uncertainty for each cycle and assay scale separately.

### J. Similarity of insulin-associated marker patterns to the fixed direction

| Cycle | Marker set and adjustment | n | Cosine similarity [95% interval] | Proportionality P |
|---|---|---|---|---|
| pooled | Eight markers; background adjusted | 2475 | 0.923 [0.893, 0.941] | $1.30 \times 10^{-14}$ |
| pooled | Eight markers; body composition adjusted | 2475 | 0.787 [0.675, 0.843] | $3.78 \times 10^{-10}$ |
| pooled | Six markers; body composition and CRP adjusted | 2475 | 0.840 [0.688, 0.915] | $2.27 \times 10^{-6}$ |
| 2001-2002 | Eight markers; background adjusted | 1327 | 0.915 [0.849, 0.946] | $2.02 \times 10^{-7}$ |
| 2001-2002 | Eight markers; body composition adjusted | 1327 | 0.773 [0.589, 0.847] | $1.53 \times 10^{-6}$ |
| 2001-2002 | Six markers; body composition and CRP adjusted | 1327 | 0.840 [0.570, 0.944] | 0.003 |
| 2003-2004 | Eight markers; background adjusted | 1148 | 0.924 [0.895, 0.938] | $1.82 \times 10^{-9}$ |
| 2003-2004 | Eight markers; body composition adjusted | 1148 | 0.793 [0.624, 0.846] | $1.08 \times 10^{-7}$ |
| 2003-2004 | Six markers; body composition and CRP adjusted | 1148 | 0.830 [0.660, 0.884] | $5.30 \times 10^{-8}$ |

Cosine similarity compares the regression coefficient vector with the corresponding frozen SAPA loadings; 1 denotes the same direction. Percentile intervals use 999 survey resamples. The proportionality test examines an exact common-multiplier restriction and is descriptive. Geometric similarity is not a percentage of insulin variation or disease-risk information recovered. The full source table also retains squared vector projections for reproducibility; these have the same geometric interpretation.

Source Data: Primary Interpretable Units, Supporting Interpretable Units, Coefficient Results, Fixed Sensitivity Results, Profile Alignment, Frozen Reference, Selection and Missingness Description, Model Integrity, Eligibility counts.

### Supplementary Table S9 | Mendelian randomization estimates and diagnostics

All seven vascular endpoints and both instrument rules are shown for the complete gSAPA input using inverse-variance weighted multiplicative random effects.

| Outcome identifier | Source label | Instrument rule | Instruments | OR [95% CI] | Holm-adjusted P |
|---|---|---|---|---|---|
| GCST90476623 | MVP Stroke | Broad | 263 | 1.097 [1.062, 1.134] | $3.93 \times 10^{-8}$ |
| GCST90476623 | MVP Stroke | AxisConsistent | 126 | 1.136 [1.076, 1.199] | $7.22 \times 10^{-6}$ |
| GCST90477991 | MVP cerebral infarction | Broad | 263 | 1.111 [1.079, 1.144] | $6.88 \times 10^{-12}$ |
| GCST90477991 | MVP cerebral infarction | AxisConsistent | 126 | 1.166 [1.110, 1.224] | $2.51 \times 10^{-9}$ |
| GCST90475936 | Coronary atherosclerosis | Broad | 263 | 1.309 [1.275, 1.344] | $6.63 \times 10^{-88}$ |
| GCST90475936 | Coronary atherosclerosis | AxisConsistent | 126 | 1.331 [1.290, 1.373] | $1.71 \times 10^{-70}$ |
| GCST90475932 | Myocardial infarction | Broad | 263 | 1.270 [1.231, 1.310] | $3.76 \times 10^{-51}$ |
| GCST90475932 | Myocardial infarction | AxisConsistent | 126 | 1.309 [1.260, 1.359] | $1.60 \times 10^{-43}$ |
| R12_I9_STR | Stroke excluding SAH | Broad | 249 | 1.075 [1.048, 1.103] | $3.93 \times 10^{-8}$ |
| R12_I9_STR | Stroke excluding SAH | AxisConsistent | 119 | 1.087 [1.045, 1.131] | $3.19 \times 10^{-5}$ |
| R12_I9_CORATHER | Coronary atherosclerosis | Broad | 249 | 1.196 [1.162, 1.231] | $2.44 \times 10^{-33}$ |
| R12_I9_CORATHER | Coronary atherosclerosis | AxisConsistent | 119 | 1.153 [1.109, 1.198] | $3.09 \times 10^{-12}$ |
| R12_I9_MI_STRICT | Myocardial infarction | Broad | 249 | 1.210 [1.171, 1.251] | $2.81 \times 10^{-29}$ |
| R12_I9_MI_STRICT | Myocardial infarction | AxisConsistent | 119 | 1.184 [1.127, 1.245] | $1.04 \times 10^{-10}$ |

MR estimates provide all four estimators under the specified complete-gSAPA and allele-quality rules. Extended Data Figure 5 displays the 56 estimates.

### Instrument-strength and heterogeneity diagnostics

The table gives all 14 IVW Cochran Q tests across seven endpoints and two instrument sets; the linked source also includes the 14 MR-Egger Q tests.

| Outcome | Instrument rule | Q | df | Nominal P |
|---|---|---|---|---|
| GCST90476623 | Broad | 409.538 | 262 | $1.42 \times 10^{-8}$ |
| GCST90476623 | AxisConsistent | 166.967 | 125 | 0.007 |
| GCST90477991 | Broad | 378.745 | 262 | $3.07 \times 10^{-6}$ |
| GCST90477991 | AxisConsistent | 151.641 | 125 | 0.053 |
| GCST90475936 | Broad | 1355.841 | 262 | $8.27 \times 10^{-147}$ |
| GCST90475936 | AxisConsistent | 273.549 | 125 | $4.15 \times 10^{-13}$ |
| GCST90475932 | Broad | 749.112 | 262 | $1.83 \times 10^{-48}$ |
| GCST90475932 | AxisConsistent | 165.037 | 125 | 0.010 |
| R12_I9_STR | Broad | 404.647 | 248 | $1.23 \times 10^{-9}$ |

| Outcome | Instrument rule | Q | df | Nominal P |
|---|---|---|---|---|
| R12_I9_STR | AxisConsistent | 147.650 | 118 | 0.034 |
| R12_I9_CORATHER | Broad | 756.144 | 248 | $8.56 \times 10^{-53}$ |
| R12_I9_CORATHER | AxisConsistent | 205.592 | 118 | $1.07 \times 10^{-6}$ |
| R12_I9_MI_STRICT | Broad | 561.104 | 248 | $2.68 \times 10^{-26}$ |
| R12_I9_MI_STRICT | AxisConsistent | 192.506 | 118 | $1.78 \times 10^{-5}$ |

All 28 Q tests.

### MR-Egger intercept diagnostics on the retained inputs

| Endpoint | Rule | n | Intercept [95% CI] | Nominal P | Holm within seven | Holm across 14 |
|---|---|---|---|---|---|---|
| GCST90475932 | Broad | 263 | 0.00613 [0.00136, 0.01090] | 0.012 | 0.060 | 0.132 |
| GCST90475936 | Broad | 263 | 0.00360 [-0.00050, 0.00769] | 0.085 | 0.256 | 0.681 |
| GCST90476623 | Broad | 263 | 0.00680 [0.00172, 0.01189] | 0.009 | 0.054 | 0.107 |
| GCST90477991 | Broad | 263 | 0.00879 [0.00428, 0.01329] | $1.54 \times 10^{-4}$ | 0.001 | 0.002 |
| R12_I9_MI_STRICT | Broad | 249 | 0.00232 [-0.00296, 0.00759] | 0.388 | 0.775 | 1.000 |
| R12_I9_CORATHER | Broad | 249 | -0.00073 [-0.00535, 0.00389] | 0.756 | 0.775 | 1.000 |
| R12_I9_STR | Broad | 249 | 0.00497 [0.00096, 0.00898] | 0.015 | 0.062 | 0.154 |
| GCST90475932 | AxisConsistent | 126 | 0.00093 [-0.00728, 0.00914] | 0.823 | 1.000 | 1.000 |
| GCST90475936 | AxisConsistent | 126 | 0.00021 [-0.00674, 0.00715] | 0.953 | 1.000 | 1.000 |
| GCST90476623 | AxisConsistent | 126 | 0.01101 [-0.00044, 0.02247] | 0.059 | 0.356 | 0.535 |
| GCST90477991 | AxisConsistent | 126 | 0.01436 [0.00408, 0.02465] | 0.007 | 0.046 | 0.086 |
| R12_I9_MI_STRICT | AxisConsistent | 119 | 0.00598 [-0.00496, 0.01691] | 0.281 | 1.000 | 1.000 |
| R12_I9_CORATHER | AxisConsistent | 119 | 0.00039 [-0.00817, 0.00896] | 0.928 | 1.000 | 1.000 |
| R12_I9_STR | AxisConsistent | 119 | -0.00130 [-0.01002, 0.00742] | 0.769 | 1.000 | 1.000 |

The intercept is in outcome log-odds units after positive-exposure orientation. Intervals use n−2 t degrees of freedom. MVP cerebral infarction is GCST90477991, distinct from the survey-reported stroke endpoint GCST90476623. Its positive intercept survives the seven-endpoint family in both rules, and the 14-test family only in Broad. Other null intercept tests do not exclude pleiotropy. The two rules share instruments. Complete diagnostics and slope cross-checks.

Supplementary Table S10 | Local genetic associations passing the specified cross-source criteria

### A. Zero sampling-error-correlation reference shown in Fig. 4c

The zero sampling-error-correlation reference retained 12 family–region associations across ten physical regions. Correlations and original pointwise 95% LAVA intervals below use the unit-variance scale shown in Fig. 4c. Intervals are conditional on the regional model and are not adjusted for selection. CORATHER denotes coronary atherosclerosis. The same physical region may occur in more than one disease family.

| Family | Region | Chr | Start, source coordinates | MVP local r_g [95% CI] | FinnGen local r_g [95% CI] |
|---|---|---|---|---|---|
| MI | 1264 | 8 | 19488889 | 0.620 [0.452, 0.821] | 0.566 [0.390, 0.777] |
| MI | 2318 | 19 | 10028841 | 0.400 [0.153, 0.671] | 0.493 [0.322, 0.690] |
| CORATHER | 961 | 6 | 31427210 | 0.388 [0.227, 0.550] | 0.341 [0.176, 0.513] |
| CORATHER | 962 | 6 | 32208902 | 0.631 [0.356, 0.942] | 0.732 [0.492, 0.981] |
| CORATHER | 970 | 6 | 33864263 | 0.935 [0.704, 1.000] | 0.337 [0.141, 0.536] |
| CORATHER | 971 | 6 | 34979271 | 0.629 [0.380, 0.911] | 0.446 [0.197, 0.716] |
| CORATHER | 977 | 6 | 42103739 | 0.486 [0.298, 0.689] | 0.515 [0.318, 0.738] |
| CORATHER | 1264 | 8 | 19488889 | 0.764 [0.657, 0.876] | 0.460 [0.346, 0.577] |
| CORATHER | 1351 | 8 | 125453323 | 0.661 [0.489, 0.843] | 0.521 [0.308, 0.758] |
| CORATHER | 1722 | 11 | 116247378 | 0.587 [0.476, 0.703] | 0.394 [0.268, 0.522] |
| CORATHER | 2209 | 17 | 45883902 | 0.756 [0.572, 0.960] | 0.512 [0.249, 0.841] |
| CORATHER | 2318 | 19 | 10028841 | 0.516 [0.323, 0.730] | 0.470 [0.305, 0.653] |

Two-scale intervals give the marginal limits and P values. Qualification counts and unevaluable regions specify coverage. Supplementary Note 4 describes the zero sampling-error-correlation assumption and cross-source criteria.

### B. Selection under matched sampling-error settings

Eligibility is unchanged across error settings. Discovery thresholds use the eligible count for each family and scale; replication uses the discovery union for that setting. Final selection requires both scales at each stage. Zero is the original reference; Default and Chi80 use the corresponding matched LDSC normalised intercepts. U/R denotes unit-variance/reference scale. The stroke families retain their distinct MVP definitions.

| Family | Error setting | Eligible U/R | Discovery U/R | Discovery union | Final associations |
|---|---|---|---|---|---|
| MI | Zero | 14/14 | 4/3 | 4 | 2 |
| MI | Default | 14/14 | 1/1 | 1 | 1 |
| MI | Chi80 | 14/14 | 1/1 | 1 | 1 |
| Coronary atherosclerosis | Zero | 151/150 | 36/36 | 36 | 10 |
| Coronary atherosclerosis | Default | 151/150 | 29/29 | 29 | 10 |
| Coronary atherosclerosis | Chi80 | 151/150 | 29/29 | 29 | 10 |
| Survey-reported stroke | Zero | 1/1 | 0/0 | 0 | 0 |
| Survey-reported stroke | Default | 1/1 | 0/0 | 0 | 0 |
| Survey-reported stroke | Chi80 | 1/1 | 0/0 | 0 | 0 |
| Cerebral infarction | Zero | 0/0 | 0/0 | 0 | 0 |
| Cerebral infarction | Default | 0/0 | 0/0 | 0 | 0 |
| Cerebral infarction | Chi80 | 0/0 | 0/0 | 0 | 0 |

Both nonzero settings retained the same 11 original associations across ten physical regions and added none. MI region 2318 no longer passed discovery. Its unit-scale MVP estimate changed from 0.400 (95% interval, 0.153 to 0.671; P = 0.002) to 0.325

(0.075 to 0.587; P = 0.011) under Default. The discovery threshold remained 0.05/14. The survey-stroke and cerebral-infarction rows describe local qualification, not the absence of genome-wide stroke association.

Complete selection summary and all final selection gates.

### C. Original selected associations across error settings

Each cell is a unit-scale local correlation with its pointwise 95% interval. Cases were fixed at the original 12 associations, regardless of selection under the new settings. The full 144-row file also retains the reference scale, P values and processing fields. All displayed directions and interval lower limits remained positive. These intervals are conditional on the error setting and are not adjusted for regional selection.

| Family | Region | Source | Zero | Default | Chi80 |
|---|---|---|---|---|---|
| MI | 1264 | MVP | 0.620 [0.452, 0.821] | 0.594 [0.426, 0.789] | 0.593 [0.426, 0.789] |
| MI | 1264 | FinnGen | 0.566 [0.390, 0.777] | 0.554 [0.378, 0.763] | 0.554 [0.379, 0.764] |
| MI | 2318 | MVP | 0.400 [0.153, 0.671] | 0.325 [0.075, 0.587] | 0.325 [0.074, 0.586] |
| MI | 2318 | FinnGen | 0.493 [0.322, 0.690] | 0.473 [0.302, 0.668] | 0.474 [0.304, 0.670] |
| CORATHER | 961 | MVP | 0.388 [0.227, 0.551] | 0.365 [0.205, 0.525] | 0.367 [0.206, 0.527] |
| CORATHER | 961 | FinnGen | 0.341 [0.176, 0.513] | 0.327 [0.163, 0.498] | 0.328 [0.164, 0.499] |
| CORATHER | 962 | MVP | 0.631 [0.356, 0.942] | 0.615 [0.337, 0.917] | 0.616 [0.339, 0.918] |
| CORATHER | 962 | FinnGen | 0.732 [0.492, 0.981] | 0.724 [0.484, 0.969] | 0.725 [0.485, 0.970] |
| CORATHER | 970 | MVP | 0.935 [0.704, 1.000] | 0.876 [0.649, 1.000] | 0.881 [0.653, 1.000] |
| CORATHER | 970 | FinnGen | 0.337 [0.141, 0.536] | 0.317 [0.122, 0.514] | 0.319 [0.124, 0.516] |
| CORATHER | 971 | MVP | 0.629 [0.380, 0.911] | 0.570 [0.324, 0.840] | 0.575 [0.328, 0.845] |
| CORATHER | 971 | FinnGen | 0.446 [0.197, 0.716] | 0.413 [0.164, 0.678] | 0.416 [0.167, 0.682] |
| CORATHER | 977 | MVP | 0.486 [0.298, 0.689] | 0.436 [0.249, 0.634] | 0.439 [0.252, 0.639] |
| CORATHER | 977 | FinnGen | 0.515 [0.318, 0.738] | 0.483 [0.286, 0.702] | 0.486 [0.289, 0.705] |
| CORATHER | 1264 | MVP | 0.764 [0.658, 0.876] | 0.744 [0.638, 0.854] | 0.745 [0.640, 0.856] |
| CORATHER | 1264 | FinnGen | 0.460 [0.346, 0.577] | 0.450 [0.336, 0.566] | 0.451 [0.337, 0.567] |
| CORATHER | 1351 | MVP | 0.661 [0.489, 0.843] | 0.616 [0.446, 0.794] | 0.619 [0.449, 0.797] |
| CORATHER | 1351 | FinnGen | 0.521 [0.308, 0.758] | 0.486 [0.272, 0.718] | 0.489 [0.275, 0.722] |
| CORATHER | 1722 | MVP | 0.587 [0.476, 0.703] | 0.567 [0.456, 0.682] | 0.568 [0.457, 0.684] |
| CORATHER | 1722 | FinnGen | 0.394 [0.268, 0.522] | 0.382 [0.255, 0.509] | 0.383 [0.256, 0.510] |
| CORATHER | 2209 | MVP | 0.756 [0.572, 0.960] | 0.711 [0.529, 0.909] | 0.714 [0.532, 0.912] |
| CORATHER | 2209 | FinnGen | 0.512 [0.250, 0.841] | 0.473 [0.211, 0.793] | 0.477 [0.214, 0.797] |
| CORATHER | 2318 | MVP | 0.516 [0.323, 0.730] | 0.458 [0.269, 0.667] | 0.462 [0.272, 0.672] |
| CORATHER | 2318 | FinnGen | 0.470 [0.305, 0.653] | 0.445 [0.281, 0.625] | 0.447 [0.283, 0.627] |

Compact unit-scale comparison, complete original-case estimates and effect-change summaries on both scales.

### D. Original replication correction held fixed

The discovery union decreased from four to one for MI and from 36 to 29 for coronary atherosclerosis. The procedure in B therefore uses less stringent replication thresholds in the nonzero settings. To assess their contribution, this diagnostic retained the original replication denominators, using the completed P values and unchanged discovery, direction, dimension and boundary gates. It was added after fitting and was not a prespecified primary comparison.

| Family | Error setting | Scenario replication denominator | Fixed original denominator | Retained: original procedure | Retained: fixed correction |
|---|---|---|---|---|---|
| MI | Zero | 4 | 4 | 2 | 2 |
| Coronary atherosclerosis | Zero | 36 | 36 | 10 | 10 |

| Family | Error setting | Scenario replication denominator | Fixed original denominator | Retained: original procedure | Retained: fixed correction |
|---|---|---|---|---|---|
| MI | Default | 1 | 4 | 1 | 1 |
| Coronary atherosclerosis | Default | 29 | 36 | 10 | 8 |
| MI | Chi80 | 1 | 4 | 1 | 1 |
| Coronary atherosclerosis | Chi80 | 29 | 36 | 10 | 9 |

The total retained counts were 12 for Zero, nine for Default and ten for Chi80 when the original correction was fixed. The following table gives membership for every original case.

| Family | Region | Default: original procedure | Default: fixed correction | Chi80: original procedure | Chi80: fixed correction |
|---|---|---|---|---|---|
| MI | 1264 | Yes | Yes | Yes | Yes |
| MI | 2318 | No | No | No | No |
| CORATHER | 961 | Yes | Yes | Yes | Yes |
| CORATHER | 962 | Yes | Yes | Yes | Yes |
| CORATHER | 970 | Yes | No | Yes | Yes |
| CORATHER | 971 | Yes | No | Yes | No |
| CORATHER | 977 | Yes | Yes | Yes | Yes |
| CORATHER | 1264 | Yes | Yes | Yes | Yes |
| CORATHER | 1351 | Yes | Yes | Yes | Yes |
| CORATHER | 1722 | Yes | Yes | Yes | Yes |
| CORATHER | 2209 | Yes | Yes | Yes | Yes |
| CORATHER | 2318 | Yes | Yes | Yes | Yes |

MI region 2318 failed discovery in both nonzero settings. Coronary atherosclerosis region 970 had Default FinnGen P = 0.001449/0.001427 across scales, between 0.05/36 and 0.05/29. Its Chi80 values (0.001360/0.001339) passed the original threshold. Region 971 had maximum replication P = 0.001667 under Default and 0.001544 under Chi80, also between the thresholds. These changes in selection accompany positive estimates; the threshold proximity and simulation precision should inform their interpretation.

Case-level fixed-correction diagnostic and family totals.

Supplementary Table S11 | Protein-anchor screen and assay coverage

| Screen category | Assays |
|---|---|
| Assays with molecularly eligible cis anchors | 1300 |
| Anchors matched to complete gSAPA | 966 |
| FDR <0.05 | 81 |
| Not evaluable against complete gSAPA | 334 |

The molecular selection cascade precedes gSAPA matching and is detailed in Supplementary Note 4. The 1,300-assay table provides assay identities, alleles and complete-gSAPA estimates, including unevaluable assays. Fig. 4d uses gSAPA_beta × orientation × protein_direction, gSAPA_P and gSAPA_FDR_pass. The orientation field aligns the GWAS allele with the protein-reference allele; protein_direction then aligns the result to higher protein concentration. Benjamini–Hochberg adjustment uses all 1,300 assays. The 966-anchor plot data give the plotted effect, −log10(P) and group. These variant-anchor associations alone do not establish regional colocalization.

### Supplementary Table S12 | Functional discovery regions, insulin sensitivity projections and vascular outcomes

Each variant is oriented to the ISI-lowering allele. The ISI decrease is shown as the absolute BMI-unadjusted effect. T is the fixed genetic projection on the original marker scales. Gene names identify the discovery regions.[16]

| Fixed source | Effect allele | ISI decrease (SE) | T (reference SE) | Candidate gene(s) |
|---|---|---|---|---|
| SLC2A4 | rs117643180-A | 0.125 (0.022) | -0.00266 (0.01726) | SLC2A4 |
| IRS1 | rs2972144-G | 0.046 (0.007) | +0.08466 (0.00573) | IRS1 |
| COBLL1 | rs12692738-T | 0.035 (0.008) | +0.04022 (0.00660) | COBLL1 / GRB14 |
| PPARG | rs11128603-A | 0.039 (0.009) | +0.02785 (0.00838) | PPARG |
| C5orf67 | rs459193-G | 0.035 (0.007) | +0.05184 (0.00628) | MAP3K1 / ANKRD55 |
| MTOR | rs2295080-T | 0.024 (0.007) | +0.00199 (0.00597) | MTOR |
| BCL2 | rs12454712-T | 0.023 (0.007) | +0.01174 (0.00565) | BCL2 |
| FAM101A | rs1906937-A | 0.031 (0.007) | +0.03933 (0.00609) | FAM101A |

Reference SEs use the original cross-trait error assumption. The aligned projections give the reference SE and its upper bound under unknown covariance. The discovery list identifies the original effect allele. For the BMI-unadjusted ISI rows in the functional comparison inputs, q equals the negative of the published ISI coefficient for that allele. The reference SE is not a covariance-robust bound. Marker-level effects, errors and all 64 signed contributions are in the complete marker table. Source labels sharing a representative are not additional independent observations. MTNR1B (rs1387153), PPP1R3B (rs7012814) and C2CD4A (rs7167878) are absent from this unadjusted-ISI comparison because the required functional effect was not provided. All three have calculable T values, supplied in the complete projection table below.

The vascular-outcome section of this table reports the Fig. 4f odds ratios. The underlying vascular outcome data list all 32 variant, source and outcome combinations, including endpoint identifiers, quality fields and interval definitions. Twenty-eight had eligible estimates. The MVP imputation $r^2$ was 0.8963 for *MTOR* and 0.8893 for *FAM101A*, below the 0.90 threshold for both MI and stroke. The SLC2A4 component table gives the eight contributions shown in Fig. 4g and their sum. The 24 MRI associations include the alleles, standard errors and intervals for abdominal SAT, thigh SAT and VAT (Fig. 4h).

#### Coverage of the original discovery list

The 11 regions combine source-study discoveries for ISI and insulin fold change (IFC).[16] The eight regions shown above were selected on availability of an unadjusted ISI effect.

| Region / representative | Discovery trait (BMI-adjusted) | T available | BMI-unadjusted ISI available |
|---|---|---|---|
| MTNR1B / rs1387153 | IFC | yes | no |
| PPP1R3B / rs7012814 | IFC | yes | no |
| C2CD4A / rs7167878 | IFC | yes | no |
| SLC2A4 / rs117643180 | IFC + ISI | yes | yes |
| IRS1 / rs2972144 | ISI | yes | yes |
| COBLL1 / rs12692738 | ISI | yes | yes |
| PPARG / rs11128603 | ISI | yes | yes |
| C5orf67 / rs459193 | ISI | yes | yes |
| MTOR / rs2295080 | ISI | yes | yes |
| BCL2 / rs12454712 | ISI | yes | yes |
| FAM101A / rs1906937 | ISI | yes | yes |

BMI-unadjusted ISI effects came from the EUR rows of Supplementary Table 6 of Williamson et al., worksheet "ST6 - primary associations", columns M–P (Beta, SE, P-value and N). They were cross-checked against the Beta_noBMI, SE_noBMI, P_noBMI and N_noBMI fields in Supplementary Table 12 ("ST12 - GRS weights"). Three representatives lacked these unadjusted ISI fields. Their BMI-adjusted ISI estimates were available in the EUR block of Supplementary Table 7 ("ST7 - associations ancestry"), columns AF–AI, and were not substituted for unadjusted effects. The second PPP1R3B marker, rs2126259, was an additional signal outside the main 11-region list. The fixed list, all 12 calculable T projections and field-specific coverage provide the complete accounting. The original alleles in those files precede the ISI-lowering reorientation. Supplementary Note 4 provides all eight coefficients and the reference error matrix.

### MI and stroke at the eight representative variants (Fig. 4f)

| Region | Variant and allele | MVP MI | FinnGen MI | MVP stroke | FinnGen stroke |
|---|---|---|---|---|---|
| SLC2A4 | rs117643180-A | 0.985 [0.933, 1.041] | 0.955 [0.900, 1.013] | 0.975 [0.907, 1.048] | 0.950 [0.900, 1.003] |
| IRS1 | rs2972144-G | 1.047 [1.031, 1.063] | 1.045 [1.025, 1.064] | 1.031 [1.008, 1.054] | 1.015 [0.998, 1.032] |
| COBLL1 | rs12692738-T | 1.026 [1.008, 1.045] | 1.002 [0.980, 1.026] | 1.038 [1.011, 1.066] | 0.998 [0.978, 1.019] |
| PPARG | rs11128603-A | 1.020 [0.994, 1.046] | 0.999 [0.975, 1.023] | 1.009 [0.976, 1.043] | 1.008 [0.986, 1.030] |
| C5orf67 | rs459193-G | 1.036 [1.019, 1.054] | 1.031 [1.012, 1.052] | 1.025 [1.001, 1.051] | 1.009 [0.991, 1.027] |
| MTOR | rs2295080-T | — | 1.016 [0.996, 1.036] | — | 0.998 [0.980, 1.015] |
| BCL2 | rs12454712-T | 1.007 [0.990, 1.025] | 1.025 [1.006, 1.043] | 1.005 [0.982, 1.028] | 1.004 [0.987, 1.020] |
| FAM101A | rs1906937-A | — | 1.002 [0.983, 1.022] | — | 1.016 [0.998, 1.034] |

Values are odds ratios [pointwise 95% confidence interval] per ISI-lowering allele. A dash indicates that the MVP imputation $r^2$ was below 0.90. MVP stroke was survey-reported. The FinnGen R12 stroke definition excludes subarachnoid haemorrhage. No multiplicity adjustment or between-variant comparison was applied to these descriptive estimates.

Supplementary Table S13 | Age-specific disease observation-record information

A. Model gains and age contrasts

| Cohort | Observation target | Age group or contrast | Model contrast | Estimate | Lower | Upper | Interval |
|---|---|---|---|---|---|---|---|
| ELSA | MI observation record | 70–89 minus 50–69 | SAPA minus background | 0.769 | -6.228 | 7.767 | six-comparison simultaneous interval |
| ELSA | MI observation record | 70–89 minus 50–69 | Full8 minus SAPA | -0.596 | -4.863 | 3.670 | six-comparison simultaneous interval |
| CHARLS | Stroke conditional on survival | 70–89 minus 50–69 | SAPA minus background | -2.146 | -9.377 | 5.084 | six-comparison simultaneous interval |
| CHARLS | Stroke conditional on survival | 70–89 minus 50–69 | Full8 minus SAPA | -0.540 | -4.477 | 3.397 | six-comparison simultaneous interval |
| ELSA | MI observation record | 50-69 | SAPA minus background | 1.659 | -1.406 | 4.389 | marginal 95% interval |
| ELSA | MI observation record | 50-69 | Full8 minus SAPA | 0.128 | -1.480 | 1.762 | marginal 95% interval |
| ELSA | MI observation record | 70-89 | SAPA minus background | 2.428 | -1.736 | 6.771 | marginal 95% interval |
| ELSA | MI observation record | 70-89 | Full8 minus SAPA | -0.469 | -3.366 | 2.132 | marginal 95% interval |
| CHARLS | Stroke conditional on survival | 50-69 | SAPA minus background | 4.738 | 2.026 | 7.374 | marginal 95% interval |
| CHARLS | Stroke conditional on survival | 50-69 | Full8 minus SAPA | -0.953 | -2.178 | 0.209 | marginal 95% interval |
| CHARLS | Joint stroke–death record | 50-69 | SAPA minus background | 4.944 | 2.068 | 7.565 | marginal 95% interval |
| CHARLS | Joint stroke–death record | 50-69 | Full8 minus SAPA | -0.612 | -1.887 | 0.660 | marginal 95% interval |
| CHARLS | Survival record | 50-69 | SAPA minus background | 0.206 | -0.213 | 0.572 | marginal 95% interval |
| CHARLS | Survival record | 50-69 | Full8 minus SAPA | 0.341 | 0.101 | 0.589 | marginal 95% interval |
| CHARLS | Stroke conditional on survival | 70-89 | SAPA minus background | 2.591 | -2.080 | 7.479 | marginal 95% interval |
| CHARLS | Stroke conditional on survival | 70-89 | Full8 minus SAPA | -1.493 | -4.070 | 1.085 | marginal 95% interval |
| CHARLS | Joint stroke–death record | 70-89 | SAPA minus background | 2.956 | -1.966 | 8.301 | marginal 95% interval |
| CHARLS | Joint stroke–death record | 70-89 | Full8 minus SAPA | -0.104 | -2.832 | 2.386 | marginal 95% interval |
| CHARLS | Survival record | 70-89 | SAPA minus background | 0.364 | -1.156 | 1.805 | marginal 95% interval |
| CHARLS | Survival record | 70-89 | Full8 minus SAPA | 1.389 | 0.284 | 2.391 | marginal 95% interval |

The CHARLS analysis uses first-positive-report histories and the 6,628-person risk set. All six simultaneous age-contrast intervals use the joint critical value of 2.642686. Conditional stroke, joint-record and survival-record targets are reported separately. The primary displays evaluate MI observation records and stroke records conditional on observed survival. Values are in 10^−3 natural-log units per person. Age-contrast estimates are older minus younger; age-specific estimates are model differences within the stated group.

### B. Paired age-specific contrasts shown in Fig. 5d

| Cohort | Target | Age | Comparison | Estimate | Lower | Upper |
|---|---|---|---|---|---|---|
| ELSA | MI observation record | 50-69 | SAPA minus Full8 | -0.128 | -1.762 | 1.480 |
| ELSA | MI observation record | 50-69 | SAPA minus PC1 | -0.026 | -0.290 | 0.220 |
| ELSA | MI observation record | 50-69 | SAPA minus equal weights | -0.357 | -0.788 | 0.130 |
| ELSA | MI observation record | 70-89 | SAPA minus Full8 | 0.469 | -2.132 | 3.366 |
| ELSA | MI observation record | 70-89 | SAPA minus PC1 | 0.050 | -0.302 | 0.429 |
| ELSA | MI observation record | 70-89 | SAPA minus equal weights | -0.076 | -0.785 | 0.628 |
| CHARLS | Stroke conditional on survival | 50-69 | SAPA minus Full8 | 0.953 | -0.209 | 2.178 |
| CHARLS | Stroke conditional on survival | 50-69 | SAPA minus PC1 | -0.313 | -0.599 | -0.007 |
| CHARLS | Stroke conditional on survival | 50-69 | SAPA minus equal weights | -0.220 | -0.711 | 0.282 |
| CHARLS | Stroke conditional on survival | 70-89 | SAPA minus Full8 | 1.493 | -1.085 | 4.070 |
| CHARLS | Stroke conditional on survival | 70-89 | SAPA minus PC1 | -0.010 | -0.521 | 0.448 |
| CHARLS | Stroke conditional on survival | 70-89 | SAPA minus equal weights | 0.442 | -0.419 | 1.305 |

All contrasts are SAPA minus the comparator, in 10^−3 natural-log units per person, with pointwise 95% paired intervals. For Full8, the source estimate and [L,U] interval were transformed to −estimate and [−U,−L]. These age-specific intervals differ from the simultaneous age-contrast intervals in part A. Plot values and original contrast directions. Full-precision part A values.

Supplementary Table S14 | Body composition associations in NHANES 2001 to 2004

| Population | Per 10% higher body measure | Age 40 [95% CI] | Age 70 [95% CI] | Age 70 minus 40 [95% CI] | Holm-adjusted P |
|---|---|---|---|---|---|
| Primary | Total fat | 0.157 [0.136, 0.177] | 0.126 [0.085, 0.167] | -0.0307 [-0.0618, 0.0003] | 0.156 |
| Primary | Trunk-to-leg fat ratio | 0.148 [0.130, 0.165] | 0.141 [0.105, 0.178] | -0.0063 [-0.0413, 0.0288] | 0.715 |
| Primary | Non-bone lean mass | 0.169 [0.115, 0.223] | 0.132 [0.039, 0.226] | -0.0367 [-0.0993, 0.0260] | 0.481 |
| Verified measured | Total fat | 0.160 [0.140, 0.181] | 0.129 [0.084, 0.174] | -0.0314 [-0.0684, 0.0057] | Not a separate primary family |
| Verified measured | Trunk-to-leg fat ratio | 0.156 [0.139, 0.172] | 0.159 [0.123, 0.194] | 0.0030 [-0.0320, 0.0380] | Not a separate primary family |
| Verified measured | Non-bone lean mass | 0.121 [0.070, 0.173] | 0.083 [-0.015, 0.181] | -0.0383 [-0.1105, 0.0338] | Not a separate primary family |

Units are fixed SAPA projection units. Other body measures are held constant; sexes are weighted equally. Pointwise intervals combine five released versions and 399 survey-design resamples. Finite joint age-comparison results:

| Population (mean analysis) | Wald / 3 | Numerator df | Denominator df | Finite P |
|---|---|---|---|---|
| DXA sample (n=3242) | 11.919 | 3 | 26.245 | $4.13 \times 10^{-5}$ |
| Verified measured scans (n=2668) | 3.922 | 3 | 28.182 | 0.019 |

The joint test uses a finite-degree-of-freedom F reference. Individual contrasts and their Holm-adjusted P values appear above. The full source file includes all eight markers, fixed BMI-removed SAPA, sex/cycle contrasts and the direct age differences. The fat-plus-lean contrast was calculated after model estimation.

### Supplementary Table S15 | Joint age comparisons and imputation uncertainty

| Population (mean analysis) | Wald / 3 | Numerator df | Denominator df | Finite P |
|---|---|---|---|---|
| DXA sample (n=3242) | 11.919 | 3 | 26.245 | $4.13 \times 10^{-5}$ |
| Verified measured scans (n=2668) | 3.922 | 3 | 28.182 | 0.019 |

| Population | Mean FMI | Directional FMI range |
|---|---|---|
| DXA sample | 0.052 | 0.005 to 0.124 |
| Verified measured scans | 0.000 | 0.000 to 0.000 |

Both comparisons use the full three-contrast covariance. Complete survey-design degrees of freedom are 30. The average-FMI small-sample rule is approximate, especially with five imputations; directional FMI values describe heterogeneity and are not missing-person percentages. The measured subset has zero between-imputation covariance. Under the same rule its denominator is 30×31/33=28.1818. No rule was selected using the new P values.

### Supplementary Table S16 | Body composition results in NHANES 1999 to 2000

NHANES 1999 to 2000. All estimates retain the frozen D scale. Conditional mean effects are per 10% higher body measure.

| Body measure | Age 40 | Age 70 | Age 70 minus 40 |
|---|---|---|---|
| Total fat | 0.157 [0.125, 0.189] | 0.160 [0.109, 0.212] | 0.003 [-0.043, 0.049] |
| Trunk-to-leg fat ratio | 0.130 [0.100, 0.161] | 0.127 [0.080, 0.174] | -0.003 [-0.055, 0.048] |
| Non-bone lean mass | 0.184 [0.079, 0.289] | 0.201 [0.051, 0.350] | 0.017 [-0.105, 0.138] |

| Population | Quantity | Component | Older minus middle-aged, 95% CI |
|---|---|---|---|
| Primary | Full fixed direction | adjusted | -0.307 [-0.627, 0.013] |
| Primary | Full fixed direction | fitted | -0.279 [-0.515, -0.043] |
| Primary | Full fixed direction | mapping | -0.101 [-0.331, 0.130] |
| Primary | Full fixed direction | distribution | -0.178 [-0.307, -0.050] |
| Primary | Full fixed direction | residual | -0.028 [-0.196, 0.140] |
| Primary | Fixed BMI contribution removed | adjusted | -0.176 [-0.415, 0.064] |
| Primary | Fixed BMI contribution removed | fitted | -0.157 [-0.285, -0.028] |
| Primary | Fixed BMI contribution removed | mapping | -0.077 [-0.202, 0.048] |
| Primary | Fixed BMI contribution removed | distribution | -0.080 [-0.148, -0.012] |
| Primary | Fixed BMI contribution removed | residual | -0.019 [-0.188, 0.150] |
| Verified measured | Full fixed direction | adjusted | -0.152 [-0.553, 0.249] |
| Verified measured | Full fixed direction | fitted | -0.112 [-0.340, 0.116] |
| Verified measured | Full fixed direction | mapping | -0.004 [-0.231, 0.224] |
| Verified measured | Full fixed direction | distribution | -0.108 [-0.230, 0.014] |
| Verified measured | Full fixed direction | residual | -0.040 [-0.247, 0.167] |
| Verified measured | Fixed BMI contribution removed | adjusted | -0.134 [-0.456, 0.189] |
| Verified measured | Fixed BMI contribution removed | fitted | -0.090 [-0.237, 0.057] |
| Verified measured | Fixed BMI contribution removed | mapping | -0.037 [-0.181, 0.107] |
| Verified measured | Fixed BMI contribution removed | distribution | -0.053 [-0.131, 0.025] |
| Verified measured | Fixed BMI contribution removed | residual | -0.044 [-0.249, 0.162] |

| Period | Population | Complete df | F (3 parameters) | Denominator df | Finite P |
|---|---|---|---|---|---|
| 2001-2004 | DXA sample | 30 | 11.919 | 26.245 | $4.13 \times 10^{-5}$ |
| 2001-2004 | Verified measured scans | 30 | 3.922 | 28.182 | 0.019 |
| 1999-2000 | DXA sample | 14 | 0.045 | 11.429 | 0.987 |
| 1999-2000 | Verified measured scans | 14 | 1.125 | 12.353 | 0.377 |

In the same 978 waist-complete participants, the DXA-minus-BMI/waist difference in the fitted B age contrast was 0.009 [-0.071, 0.090]. These separately estimated survey periods were not pooled or subjected to a direct heterogeneity test. Intervals are pointwise; individual primary age-contrast Holm values and all constituent-marker results are in the aggregate source files.

### Routine BMI/waist distribution contribution in the matched population

| Metric | Matched n | Distribution contribution [95% CI] |
|---|---|---|
| B | 978 | -0.093 [-0.175, -0.011] |
| B_without_BMI | 978 | -0.033 [-0.070, 0.003] |

In the 1999–2000 sample, the BMI and waist estimates use the participants with matched DXA data. The signed values give the contribution to the age difference in covariance between markers (B). Component estimates.

### Supplementary Table S17 | Body-distribution moments and their six additive contributions

Body variables are on natural-log scales. Body moments and signed contributions describe within-cell background-adjusted covariance, with equal sex/cycle averaging. Intervals are pointwise 95% Rubin/Barnard-Rubin intervals; they do not test a ranking of components.

#### Underlying body moments

| Body term | Age contrast or group | 2001 to 2004, 95% CI | 1999 to 2000, 95% CI |
|---|---|---|---|
| Total-fat variance | 40 to 59 | 0.128 [0.113, 0.142] | 0.131 [0.103, 0.159] |
| Total-fat variance | 60 to 79 | 0.103 [0.090, 0.116] | 0.103 [0.085, 0.122] |
| Total-fat variance | Older minus middle-aged | -0.025 [-0.043, -0.007] | -0.028 [-0.048, -0.007] |
| Trunk-to-leg ratio variance | 40 to 59 | 0.072 [0.062, 0.083] | 0.067 [0.052, 0.083] |
| Trunk-to-leg ratio variance | 60 to 79 | 0.070 [0.063, 0.078] | 0.064 [0.052, 0.076] |
| Trunk-to-leg ratio variance | Older minus middle-aged | -0.002 [-0.014, 0.010] | -0.003 [-0.020, 0.013] |
| Non-bone lean variance | 40 to 59 | 0.020 [0.017, 0.022] | 0.022 [0.017, 0.027] |
| Non-bone lean variance | 60 to 79 | 0.016 [0.014, 0.018] | 0.018 [0.014, 0.021] |
| Non-bone lean variance | Older minus middle-aged | -0.00347 [-0.00651, -0.00043] | -0.00451 [-0.00991, 0.00089] |
| Total fat / trunk-to-leg ratio covariance | 40 to 59 | 0.041 [0.033, 0.048] | 0.038 [0.024, 0.052] |
| Total fat / trunk-to-leg ratio covariance | 60 to 79 | 0.021 [0.013, 0.030] | 0.01264 [-0.00092, 0.02619] |
| Total fat / trunk-to-leg ratio covariance | Older minus middle-aged | -0.019 [-0.028, -0.011] | -0.025 [-0.043, -0.008] |
| Total fat / lean covariance | 40 to 59 | 0.039 [0.033, 0.044] | 0.042 [0.032, 0.052] |
| Total fat / lean covariance | 60 to 79 | 0.029 [0.025, 0.033] | 0.031 [0.024, 0.039] |
| Total fat / lean covariance | Older minus middle-aged | -0.010 [-0.016, -0.003] | -0.011 [-0.018, -0.003] |
| Trunk-to-leg ratio / lean covariance | 40 to 59 | 0.017 [0.014, 0.019] | 0.013 [0.007, 0.018] |
| Trunk-to-leg ratio / lean covariance | 60 to 79 | 0.007 [0.004, 0.010] | 0.006 [0.001, 0.011] |
| Trunk-to-leg ratio / lean covariance | Older minus middle-aged | -0.010 [-0.013, -0.006] | -0.007 [-0.012, -0.002] |

#### Contributions to the distribution component of B

| Summary | Component | 2001 to 2004, 95% CI | 1999 to 2000, 95% CI |
|---|---|---|---|
| All eight markers | Total-fat variance | -0.032 [-0.061, -0.003] | -0.038 [-0.089, 0.012] |
| All eight markers | Trunk-to-leg ratio variance | -0.003 [-0.019, 0.014] | -0.004 [-0.020, 0.011] |
| All eight markers | Non-bone lean variance | 0.00067 [-0.00886, 0.01020] | -0.007 [-0.033, 0.020] |
| All eight markers | Total fat / trunk-to-leg ratio covariance | -0.058 [-0.089, -0.027] | -0.071 [-0.138, -0.005] |
| All eight markers | Total fat / lean covariance | -0.02076 [-0.04055, -0.00097] | -0.035 [-0.087, 0.016] |
| All eight markers | Trunk-to-leg ratio / lean covariance | -0.030 [-0.051, -0.010] | -0.023 [-0.050, 0.004] |
| All eight markers | Distribution total | -0.143 [-0.210, -0.077] | -0.178 [-0.307, -0.050] |
| BMI cross-terms removed | Total-fat variance | -0.013 [-0.028, 0.002] | -0.015 [-0.043, 0.013] |
| BMI cross-terms removed | Trunk-to-leg ratio variance | -0.003 [-0.021, 0.015] | -0.005 [-0.022, 0.013] |
| BMI cross-terms removed | Non-bone lean variance | 0.001 [-0.003, 0.006] | -0.003 [-0.016, 0.011] |
| BMI cross-terms removed | Total fat / trunk-to-leg ratio covariance | -0.033 [-0.056, -0.010] | -0.042 [-0.088, 0.003] |
| BMI cross-terms removed | Total fat / lean covariance | -0.002 [-0.016, 0.012] | -0.008 [-0.040, 0.024] |

| Summary | Component | 2001 to 2004, 95% CI | 1999 to 2000, 95% CI |
| --- | --- | --- | --- |
| BMI cross-terms removed | Trunk-to-leg ratio / lean covariance | -0.004 [-0.021, 0.013] | -0.007 [-0.027, 0.013] |
| BMI cross-terms removed | Distribution total | -0.054 [-0.090, -0.017] | -0.080 [-0.148, -0.012] |

The two periods used 2,081 and 986 participants. Only the BMI entry of the fixed unit direction was set to zero in the BMI-omitted summary; other entries were unchanged. All six terms sum to the original distribution component. Full precision and variance components are in DXA_Component_Results.csv.

Supplementary Table S18 | Body-related age differences in shared marker variation

| Metric | Component | Primary, n=2081 [95% CI] | Verified measured, n=1676 [95% CI] |
|---|---|---|---|
| B | Overall adjusted | -0.428 [-0.598, -0.257] | -0.240 [-0.427, -0.054] |
| B | Body-related total | -0.440 [-0.590, -0.291] | -0.267 [-0.399, -0.136] |
| B | Body-marker relationship | -0.297 [-0.429, -0.165] | -0.196 [-0.314, -0.078] |
| B | Body measurement second moments | -0.143 [-0.210, -0.077] | -0.071 [-0.139, -0.004] |
| B | Remaining covariance | 0.013 [-0.073, 0.098] | 0.027 [-0.078, 0.131] |
| B_without_BMI | Overall adjusted | -0.208 [-0.326, -0.089] | -0.107 [-0.252, 0.038] |
| B_without_BMI | Body-related total | -0.226 [-0.313, -0.140] | -0.137 [-0.225, -0.050] |
| B_without_BMI | Body-marker relationship | -0.173 [-0.254, -0.091] | -0.111 [-0.191, -0.031] |
| B_without_BMI | Body measurement second moments | -0.054 [-0.090, -0.017] | -0.026 [-0.065, 0.013] |
| B_without_BMI | Remaining covariance | 0.019 [-0.070, 0.108] | 0.030 [-0.075, 0.135] |

All differences compare ages 60 to 79 with 40 to 59. Units are fixed-reference covariance units. Signed components sum exactly; this is a descriptive projection decomposition.

| Paired DXA minus BMI/waist, n=2057 | Component | Difference [95% CI] |
|---|---|---|
| B | fitted | -0.011 [-0.059, 0.036] |
| B | mapping | 0.038 [-0.021, 0.096] |
| B | distribution | -0.049 [-0.096, -0.002] |
| B | residual | 0.011 [-0.036, 0.059] |
| B_without_BMI | fitted | -0.019 [-0.068, 0.030] |
| B_without_BMI | mapping | 0.003 [-0.049, 0.054] |
| B_without_BMI | distribution | -0.022 [-0.052, 0.009] |
| B_without_BMI | residual | 0.019 [-0.030, 0.068] |

The paired population is identical for both representations. Individual 95% intervals are shown, with no equivalence margin. Full age-group quantities, unadjusted differences, cycle-specific results and imputation variance components are retained in AgeBody_Pooled_Results.csv.

### Aggregate source files

Source files: AgeBody_Analysis_Sample_Sizes.csv; AgeBody_Intersection_and_Imputation.csv; AgeBody_Joint_SAPA_Age_Comparison.csv; AgeBody_JointResult_Explanatory_Reexpression.csv; AgeBody_Paired_BodyRepresentation_Contrasts.csv; AgeBody_Pooled_Results.csv

### Routine BMI/waist distribution contribution in the matched population

| Metric | Matched n | Distribution contribution [95% CI] |
|---|---|---|
| B | 2057 | -0.093 [-0.147, -0.039] |
| B_without_BMI | 2057 | -0.032 [-0.057, -0.007] |

The 2001–2004 comparison uses measured BMI and waist in the same participants as DXA. Both rows give contributions to the age difference in covariance between markers (B). The second row excludes terms involving BMI as a constituent marker. Component estimates.

### Supplementary Table S19 | Frozen cross-period transfer and diagnostic comparisons

Source: 2001 to 2004, n=2,057. Target: 1999 to 2000, n=978. These are the original BMI/waist-complete structural samples, not the full DXA samples. Observed-target intervals use target-only design uncertainty (14 degrees of freedom); source-copy intervals use source-only uncertainty (30). Other scalar intervals combine source and target errors using Welch degrees of freedom. Squared-error improvement uses a finite-reference joint-ellipse projection interval, as specified in Supplementary Notes. All 54 saved quantities are reproduced below.

| Summary | Quantity | Age contrast or group | Estimate [95% interval] | Interval method |
|---|---|---|---|---|
| BMI cross-terms removed | Observed target | 40 to 59 | 0.484 [0.329, 0.639] | target-only design t, df=14 |
| BMI cross-terms removed | Observed target | 60 to 79 | 0.317 [0.113, 0.522] | target-only design t, df=14 |
| BMI cross-terms removed | Observed target | Older minus middle-aged | -0.167 [-0.408, 0.075] | target-only design t, df=14 |
| BMI cross-terms removed | Source-period copy | 40 to 59 | 0.497 [0.421, 0.572] | source-only design t, df=30 |
| BMI cross-terms removed | Source-period copy | 60 to 79 | 0.297 [0.216, 0.378] | source-only design t, df=30 |
| BMI cross-terms removed | Source-period copy | Older minus middle-aged | -0.200 [-0.320, -0.080] | source-only design t, df=30 |
| BMI cross-terms removed | Target-body adapted | 40 to 59 | 0.516 [0.419, 0.614] | joint source–target; finite Welch t |
| BMI cross-terms removed | Target-body adapted | 60 to 79 | 0.302 [0.222, 0.383] | joint source–target; finite Welch t |
| BMI cross-terms removed | Target-body adapted | Older minus middle-aged | -0.214 [-0.337, -0.091] | joint source–target; finite Welch t |
| BMI cross-terms removed | Target frozen residual minus source residual | 40 to 59 | -0.027 [-0.156, 0.103] | joint source–target; finite Welch t |
| BMI cross-terms removed | Target frozen residual minus source residual | 60 to 79 | -0.024 [-0.199, 0.150] | joint source–target; finite Welch t |
| BMI cross-terms removed | Target frozen residual minus source residual | Older minus middle-aged | 0.003 [-0.195, 0.200] | joint source–target; finite Welch t |
| BMI cross-terms removed | Target cross-term | 40 to 59 | -0.005 [-0.119, 0.109] | joint source–target; finite Welch t |
| BMI cross-terms removed | Target cross-term | 60 to 79 | 0.039 [-0.037, 0.116] | joint source–target; finite Welch t |
| BMI cross-terms removed | Target cross-term | Older minus middle-aged | 0.045 [-0.096, 0.185] | joint source–target; finite Welch t |
| BMI cross-terms removed | Target body component under frozen source mapping | 40 to 59 | 0.286 [0.197, 0.376] | joint source–target; finite Welch t |
| BMI cross-terms removed | Target body component under frozen source mapping | 60 to 79 | 0.065 [0.027, 0.103] | joint source–target; finite Welch t |
| BMI cross-terms removed | Target body component under frozen source mapping | Older minus middle-aged | -0.221 [-0.309, -0.134] | joint source–target; finite Welch t |
| BMI cross-terms removed | Source copy minus observed | 40 to 59 | 0.012 [-0.156, 0.181] | joint source–target; finite Welch t |
| BMI cross-terms removed | Adapted minus observed | 40 to 59 | 0.032 [-0.136, 0.200] | joint source–target; finite Welch t |
| BMI cross-terms removed | Squared-error improvement, positive favours adaptation | 40 to 59 | -0.00087 [-0.03061, 0.02358] | 95% joint finite-reference ellipse projection; approximate |
| BMI cross-terms removed | Source copy minus observed | 60 to 79 | -0.020 [-0.236, 0.196] | joint source–target; finite Welch t |
| BMI cross-terms removed | Adapted minus observed | 60 to 79 | -0.015 [-0.231, 0.201] | joint source–target; finite Welch t |

| Summary | Quantity | Age contrast or group | Estimate [95% interval] | Interval method |
|---|---|---|---|---|
| BMI cross-terms removed | Squared-error improvement, positive favours adaptation | 60 to 79 | 0.00019 [-0.00805, 0.00973] | 95% joint finite-reference ellipse projection; approximate |
| BMI cross-terms removed | Source copy minus observed | Older minus middle-aged | -0.033 [-0.296, 0.230] | joint source–target; finite Welch t |
| BMI cross-terms removed | Adapted minus observed | Older minus middle-aged | -0.047 [-0.310, 0.216] | joint source–target; finite Welch t |
| BMI cross-terms removed | Squared-error improvement, positive favours adaptation | Older minus middle-aged | -0.001 [-0.043, 0.032] | 95% joint finite-reference ellipse projection; approximate |
| All eight markers | Observed target | 40 to 59 | 0.796 [0.583, 1.008] | target-only design t, df=14 |
| All eight markers | Observed target | 60 to 79 | 0.498 [0.240, 0.755] | target-only design t, df=14 |
| All eight markers | Observed target | Older minus middle-aged | -0.298 [-0.622, 0.026] | target-only design t, df=14 |
| All eight markers | Source-period copy | 40 to 59 | 0.808 [0.699, 0.918] | source-only design t, df=30 |
| All eight markers | Source-period copy | 60 to 79 | 0.393 [0.301, 0.485] | source-only design t, df=30 |
| All eight markers | Source-period copy | Older minus middle-aged | -0.415 [-0.587, -0.244] | source-only design t, df=30 |
| All eight markers | Target-body adapted | 40 to 59 | 0.854 [0.682, 1.026] | joint source–target; finite Welch t |
| All eight markers | Target-body adapted | 60 to 79 | 0.411 [0.309, 0.514] | joint source–target; finite Welch t |
| All eight markers | Target-body adapted | Older minus middle-aged | -0.443 [-0.633, -0.253] | joint source–target; finite Welch t |
| All eight markers | Target frozen residual minus source residual | 40 to 59 | -0.024 [-0.154, 0.106] | joint source–target; finite Welch t |
| All eight markers | Target frozen residual minus source residual | 60 to 79 | -0.028 [-0.200, 0.144] | joint source–target; finite Welch t |
| All eight markers | Target frozen residual minus source residual | Older minus middle-aged | -0.004 [-0.201, 0.193] | joint source–target; finite Welch t |
| All eight markers | Target cross-term | 40 to 59 | -0.034 [-0.229, 0.161] | joint source–target; finite Welch t |
| All eight markers | Target cross-term | 60 to 79 | 0.115 [-0.027, 0.257] | joint source–target; finite Welch t |
| All eight markers | Target cross-term | Older minus middle-aged | 0.149 [-0.120, 0.418] | joint source–target; finite Welch t |
| All eight markers | Target body component under frozen source mapping | 40 to 59 | 0.629 [0.457, 0.802] | joint source–target; finite Welch t |
| All eight markers | Target body component under frozen source mapping | 60 to 79 | 0.178 [0.101, 0.254] | joint source–target; finite Welch t |
| All eight markers | Target body component under frozen source mapping | Older minus middle-aged | -0.452 [-0.620, -0.283] | joint source–target; finite Welch t |
| All eight markers | Source copy minus observed | 40 to 59 | 0.013 [-0.220, 0.246] | joint source–target; finite Welch t |
| All eight markers | Adapted minus observed | 40 to 59 | 0.058 [-0.160, 0.277] | joint source–target; finite Welch t |
| All eight markers | Squared-error improvement, positive favours adaptation | 40 to 59 | -0.003 [-0.084, 0.071] | 95% joint finite-reference ellipse projection; approximate |
| All eight markers | Source copy minus observed | 60 to 79 | -0.105 [-0.374, 0.165] | joint source–target; finite Welch t |
| All eight markers | Adapted minus observed | 60 to 79 | -0.087 [-0.354, 0.181] | joint source–target; finite Welch t |

| Summary | Quantity | Age contrast or group | Estimate [95% interval] | Interval method |
|---|---|---|---|---|
| All eight markers | Squared-error improvement, positive favours adaptation | 60 to 79 | 0.003 [-0.020, 0.043] | 95% joint finite-reference ellipse projection; approximate |
| All eight markers | Source copy minus observed | Older minus middle-aged | -0.117 [-0.475, 0.240] | joint source–target; finite Welch t |
| All eight markers | Adapted minus observed | Older minus middle-aged | -0.145 [-0.499, 0.209] | joint source–target; finite Welch t |
| All eight markers | Squared-error improvement, positive favours adaptation | Older minus middle-aged | -0.007 [-0.141, 0.102] | 95% joint finite-reference ellipse projection; approximate |

Units are B, except squared-error improvements ($B^2$). Positive improvement favours adaptation. Error covariance and marginal support checks are supplied separately: Transfer_Results.csv · Loss_ErrorCovariance.json · Transfer_Support_Description.csv · Comparison_Sample_Lineage.tsv.

### Supplementary Table S20 | Identifier sensitivity of the fixed clinical reference

The original reference comprised 4,565 participants; 300 required completion of 617 marker cells (1.689% of all eight-marker cells). Twenty completions used the paired with- and without-SEQN specifications at ten fixed seeds, 2026–2035. All twenty factor fits were admissible. Comparisons below apply the resulting references to the same 4,265 complete panels.

| Quantity | Seed 2026: excluding versus retaining SEQN | Range across ten same-seed pairs |
|---|---|---|
| Directional angle in the original D metric, degrees | 0.436 | 0.314–2.074 |
| Observed-coordinate Pearson correlation | 0.999989 | 0.99970–0.999994 |
| Observed-coordinate RMSE, comparator-coordinate SD units | 0.010 | 0.005–0.024 |
| Adjusted-coordinate RMSE, comparator-coordinate SD units | 0.009 | 0.005–0.024 |
| Observed-coordinate 95th percentile absolute difference, comparator SD units | 0.016 | 0.011–0.049 |
| Observed-coordinate maximum absolute difference, comparator SD units | 0.024 | 0.024–0.113 |

The comparator is the with-SEQN coordinate from the same seed; its weighted SD at seed 2026 equals that of the original NHANES-reference score. The seed-2026 completion and six-decimal loadings with SEQN exactly reproduced the original reference. The frozen NHANES reference and outcome estimates were retained. Seed ranges describe algorithmic variation, not confidence intervals. Individual missing-cell completions varied more than aggregate reference parameters; similar variation was present when SEQN was retained and only the seed changed. These comparisons do not measure the accuracy of the unobserved missing values. Reference jackknife intervals remain conditional on a single completion.

Complete coordinate comparisons · Reference parameters · Direction comparisons · Missing-cell and seed context · Data dictionary

Supplementary Table S21 | HRS lower-PCE observation support and limited association analysis

### A. Eligibility and observation support

| Population | n | All subsequent stroke reports | Deaths | Unknown-stroke death paths | Known positive by 4 years | Supported negative at 4 years | Unresolved at 4 years |
|---|---|---|---|---|---|---|---|
| Original 2018 stroke-negative risk set | 2282 | 82 | 112 | 15 | 65 | 1113 | 1104 |
| Age 40 to 79 in 2016 | 2018 | 67 | 71 | 8 | 55 | 949 | 1014 |
| 2016/2018 heart disease and diabetes explicitly denied | 1142 | 26 | 30 | 3 | 21 | 537 | 584 |
| Earlier raw histories also clear | 1056 | 24 | 25 | 3 | 20 | 493 | 543 |
| PCE race/ethnicity supported | 887 | 16 | 23 | 3 | 12 | 420 | 455 |
| All specified PCE inputs/ranges eligible | 800 | 16 | 15 | 2 | 12 | 371 | 417 |
| PCE below 5% | 218 | 2 | 2 | 1 | 1 | 76 | 141 |
| Lower PCE: SAPA upper quartile | 55 | 0 | 0 | 0 | 0 | 18 | 37 |
| Lower PCE: remaining participants | 163 | 2 | 2 | 1 | 1 | 58 | 104 |

Counts refer to the 2018 entry after the 2016 blood draw. Earlier raw histories through entry supplied the eligibility rule; later answers were not used to revise earlier qualification. The lower-PCE group included 204 women (93.6%), 192 non-Hispanic White and 26 non-Hispanic Black participants, and 213 households. Its age range in 2016 was 48 to 68, with mean 58.147 years. Median blood-to-entry interval was 1.812 years and median last observation was 3.833 years. The two total stroke reports do not both have known onset before four years. Unresolved status is not synonymous with loss to follow-up.

### B. Continuous SAPA associations

| Model | n / reports | HR per original HRS SD [approximate profile 95% interval] | Likelihood-ratio statistic | Asymptotic P |
|---|---|---|---|---|
| SAPA only | 218 / 2 | 0.574 [0.094, 2.975] | 0.425 | 0.515 |
| SAPA + centred logit(PCE) | 218 / 2 | 0.463 [0.075, 2.596] | 0.761 | 0.383 |

One SD is 1.513622 raw D units, fixed from the original 2,282-person HRS sample. This differs from the per-raw-unit scale in Supplementary Table S5. The model uses constant stroke and death rates and a common death rate before and after stroke. The approximate profile intervals reflect the two observed reports and the assumptions of the working model. Both point estimates are below one, with intervals spanning associations in either direction.

### C. Held-out fold support

| Fold | Training upper-quartile cutoff, raw D units | Test n | Stroke reports | Deaths |
|---|---|---|---|---|
| 1 | 0.491017 | 47 | 1 | 0 |

| Fold | Training upper-quartile cutoff, raw D units | Test n | Stroke reports | Deaths |
|---|---|---|---|---|
| 2 | 0.389447 | 45 | 0 | 0 |
| 3 | 0.469107 | 46 | 1 | 1 |
| 4 | 0.465261 | 40 | 0 | 0 |
| 5 | 0.460652 | 40 | 0 | 1 |

Three test folds had no stroke reports, leaving too few events to evaluate incremental prediction. None of the 55 people in the upper SAPA quartile reported stroke during the observed follow-up. Four lower-PCE people had LDL ≥190 mg/dl and six had HbA1c ≥6.5%; these are separate clinical considerations. HRS was a previously examined cohort.

Source Data: sample flow, continuous associations, timing/scale and fold support.

Supplementary Table S22 | Primary sample composition and the verified measured subset

### Primary structural population, ages 40–79

| Variable | All, n=2081 | Verified measured, n=1676 | Remaining, n=405 |
|---|---|---|---|
| Age, years | 54.828 (10.353) | 54.517 (10.166) | 56.198 (11.040) |
| Women, % | 51.978 | 51.791 | 52.805 |
| 2003–2004 cycle, % | 51.483 | 50.737 | 54.776 |
| BMI, kg/m² | 28.557 (5.974) | 27.614 (4.699) | 32.720 (8.632) |
| Waist, cm | 98.733 (14.760); n=2057 | 96.565 (12.602); n=1666 | 108.592 (19.181); n=391 |
| SAPA, fixed units | 0.346 (1.513) | 0.227 (1.447) | 0.870 (1.679) |
| Total fat, kg | 29.520 (11.068) | 27.755 (8.914) | 37.308 (15.429) |
| Trunk fat, kg | 14.838 (6.138) | 13.872 (4.940) | 19.101 (8.599) |
| Whole-leg fat, kg | 9.949 (4.133) | 9.408 (3.570) | 12.339 (5.404) |
| Trunk-to-leg fat ratio | 1.557 (0.480) | 1.547 (0.470) | 1.603 (0.518) |
| Non-bone lean mass, kg | 50.785 (12.398) | 49.584 (11.349) | 56.085 (15.139) |

### Ages 40–59

| Variable | All, n=1115 | Verified measured, n=915 | Remaining, n=200 |
|---|---|---|---|
| Age, years | 48.846 (5.518) | 48.772 (5.470) | 49.198 (5.730) |
| Women, % | 51.574 | 51.385 | 52.468 |
| 2003–2004 cycle, % | 50.118 | 49.698 | 52.110 |
| BMI, kg/m² | 28.609 (6.223) | 27.560 (4.756) | 33.585 (9.231) |
| Waist, cm | 97.790 (15.129); n=1103 | 95.430 (12.570); n=911 | 109.436 (20.384); n=192 |
| SAPA, fixed units | 0.239 (1.575) | 0.092 (1.473) | 0.935 (1.835) |
| Total fat, kg | 29.298 (11.467) | 27.345 (8.995) | 38.569 (16.355) |
| Trunk fat, kg | 14.653 (6.380) | 13.575 (4.976) | 19.773 (9.202) |
| Whole-leg fat, kg | 9.964 (4.180) | 9.373 (3.561) | 12.768 (5.538) |
| Trunk-to-leg fat ratio | 1.525 (0.466) | 1.514 (0.457) | 1.580 (0.504) |
| Non-bone lean mass, kg | 51.805 (12.682) | 50.425 (11.472) | 58.357 (15.737) |

### Ages 60–79

| Variable | All, n=966 | Verified measured, n=761 | Remaining, n=205 |
|---|---|---|---|
| Age, years | 67.631 (5.561) | 67.340 (5.396) | 68.743 (6.021) |
| Women, % | 52.845 | 52.697 | 53.409 |
| 2003–2004 cycle, % | 54.405 | 53.054 | 59.554 |
| BMI, kg/m² | 28.447 (5.402) | 27.733 (4.566) | 31.170 (7.184) |
| Waist, cm | 100.749 (13.723); n=954 | 99.105 (12.298); n=755 | 107.110 (16.760); n=199 |
| SAPA, fixed units | 0.575 (1.343) | 0.529 (1.337) | 0.753 (1.347) |
| Total fat, kg | 29.997 (10.146) | 28.672 (8.660) | 35.046 (13.313) |
| Trunk fat, kg | 15.233 (5.564) | 14.534 (4.793) | 17.898 (7.240) |
| Whole-leg fat, kg | 9.917 (4.032) | 9.484 (3.590) | 11.568 (5.064) |
| Trunk-to-leg fat ratio | 1.625 (0.502) | 1.620 (0.492) | 1.644 (0.539) |

| Variable | All, n=966 | Verified measured, n=761 | Remaining, n=205 |
|---|---|---|---|
| Non-bone lean mass, kg | 48.603 (11.465) | 47.709 (10.835) | 52.013 (13.047) |

### Whole conditional-mean population, ages 20–79

| Variable | All, n=3242 | Verified measured, n=2668 | Remaining, n=574 |
|---|---|---|---|
| Age, years | 44.332 (15.198) | 43.771 (15.051) | 47.168 (15.614) |
| Women, % | 50.646 | 50.143 | 53.181 |
| 2003–2004 cycle, % | 50.635 | 49.648 | 55.620 |
| BMI, kg/m² | 28.111 (6.187) | 27.140 (4.914) | 33.015 (8.991) |
| Waist, cm | 96.359 (15.439); n=3210 | 94.091 (13.079); n=2654 | 108.114 (20.551); n=556 |
| SAPA, fixed units | -0.035 (1.615) | -0.177 (1.539) | 0.685 (1.791) |
| Total fat, kg | 28.266 (11.676) | 26.411 (9.474) | 37.632 (16.321) |
| Trunk fat, kg | 13.876 (6.516) | 12.856 (5.299) | 19.026 (9.127) |
| Whole-leg fat, kg | 9.834 (4.214) | 9.263 (3.630) | 12.715 (5.561) |
| Trunk-to-leg fat ratio | 1.449 (0.461) | 1.434 (0.452) | 1.523 (0.501) |
| Non-bone lean mass, kg | 51.239 (12.465) | 50.019 (11.224) | 57.402 (16.074) |

Values are survey-weighted means (SD) or percentages; n is unweighted. Original fasting-subsample weights are used without equalizing sex or cycle. Each variable uses its valid records. The remaining group includes participants with imputation indicators and participants with zero indicators but overlap-flagged soft tissue and varying values. These categories are listed separately in the composition tables. Body summaries are calculated in each completed dataset and averaged over five versions; across-version ranges are in the composition summaries. The routine measurements are identical across these versions. These descriptive profiles differ from the equal-sex/equal-cycle population used for the structural decomposition. They quantify selection into the measured subset and do not identify an imputation effect.

### Source tables

Source files: Finite_Joint_Results.csv; Composition_Summary.csv; Composition_Counts.csv; Composition_By_Imputation.csv

Supplementary Table S23 | Replication sample composition and the verified measured subset

Survey-weighted mean (population-denominator SD), averaged across five completed datasets; n is variable-specific. Female percentage uses the original survey weights. The trunk-to-leg ratio is calculated within person before averaging. Profiles use the observed sex composition; the age-structure decomposition elsewhere averages sex-specific matrices equally. These are descriptive summaries without group-comparison tests.

### Ages 40 to 79

| Measure | All | Verified measured | Remaining |
|---|---|---|---|
| Age, years | 55.067 (10.641); n=986 | 55.057 (10.668); n=769 | 55.104 (10.534); n=217 |
| Female, weighted % | 52.694%; n=986 | 52.391%; n=769 | 53.890%; n=217 |
| BMI, kg/m² | 28.321 (6.083); n=986 | 27.161 (4.720); n=769 | 32.890 (8.294); n=217 |
| Waist, cm | 97.829 (15.439); n=978 | 94.955 (12.707); n=767 | 109.351 (19.514); n=211 |
| SAPA, frozen units | 0.508 (1.528); n=986 | 0.327 (1.453); n=769 | 1.224 (1.604); n=217 |
| Total fat, kg | 29.502 (11.378); n=986 | 27.272 (8.616); n=769 | 38.282 (15.821); n=217 |
| Trunk fat, kg | 15.068 (6.352); n=986 | 13.831 (4.888); n=769 | 19.938 (8.683); n=217 |
| Whole-leg fat, kg | 9.850 (4.256); n=986 | 9.138 (3.404); n=769 | 12.652 (5.829); n=217 |
| Trunk-to-leg fat ratio | 1.598 (0.490); n=986 | 1.586 (0.488); n=769 | 1.644 (0.494); n=217 |
| Non-bone lean mass, kg | 50.025 (12.688); n=986 | 48.527 (11.356); n=769 | 55.925 (15.603); n=217 |

### Ages 40 to 59

| Measure | All | Verified measured | Remaining |
|---|---|---|---|
| Age, years | 48.566 (5.705); n=492 | 48.564 (5.585); n=388 | 48.576 (6.167); n=104 |
| Female, weighted % | 51.517%; n=492 | 50.884%; n=388 | 54.080%; n=104 |
| BMI, kg/m² | 28.392 (6.364); n=492 | 27.079 (4.842); n=388 | 33.707 (8.619); n=104 |
| Waist, cm | 97.154 (16.126); n=487 | 93.742 (12.700); n=387 | 111.304 (20.521); n=100 |
| SAPA, frozen units | 0.395 (1.578); n=492 | 0.198 (1.478); n=388 | 1.194 (1.714); n=104 |
| Total fat, kg | 29.317 (11.882); n=492 | 26.772 (8.671); n=388 | 39.613 (16.611); n=104 |
| Trunk fat, kg | 14.885 (6.688); n=492 | 13.455 (4.936); n=388 | 20.671 (9.239); n=104 |
| Whole-leg fat, kg | 9.869 (4.333); n=492 | 9.080 (3.392); n=388 | 13.060 (5.956); n=104 |
| Trunk-to-leg fat ratio | 1.561 (0.480); n=492 | 1.544 (0.476); n=388 | 1.628 (0.486); n=104 |
| Non-bone lean mass, kg | 51.577 (13.082); n=492 | 49.771 (11.451); n=388 | 58.883 (16.321); n=104 |

### Ages 60 to 79

| Measure | All | Verified measured | Remaining |
|---|---|---|---|
| Age, years | 67.606 (5.488); n=494 | 67.783 (5.579); n=381 | 66.944 (5.076); n=113 |
| Female, weighted % | 54.965%; n=494 | 55.344%; n=381 | 53.546%; n=113 |
| BMI, kg/m² | 28.184 (5.496); n=494 | 27.323 (4.467); n=381 | 31.407 (7.444); n=113 |
| Waist, cm | 99.128 (13.935); n=491 | 97.333 (12.382); n=380 | 105.879 (17.036); n=111 |
| SAPA, frozen units | 0.727 (1.400); n=494 | 0.579 (1.368); n=381 | 1.279 (1.381); n=113 |
| Total fat, kg | 29.857 (10.328); n=494 | 28.251 (8.421); n=381 | 35.869 (13.953); n=113 |
| Trunk fat, kg | 15.419 (5.631); n=494 | 14.567 (4.707); n=381 | 18.609 (7.386); n=113 |
| Whole-leg fat, kg | 9.813 (4.104); n=494 | 9.253 (3.424); n=381 | 11.912 (5.513); n=113 |
| Trunk-to-leg fat ratio | 1.669 (0.502); n=494 | 1.669 (0.501); n=381 | 1.672 (0.503); n=113 |

| Measure | All | Verified measured | Remaining |
|---|---|---|---|
| Non-bone lean mass, kg | 47.032 (11.306); n=494 | 46.090 (10.760); n=381 | 50.560 (12.535); n=113 |

### Ages 20 to 79

| Measure | All | Verified measured | Remaining |
|---|---|---|---|
| Age, years | 43.620 (15.148); n=1,513 | 43.109 (15.199); n=1,215 | 46.011 (14.670); n=298 |
| Female, weighted % | 50.416%; n=1,513 | 49.045%; n=1,215 | 56.836%; n=298 |
| BMI, kg/m² | 27.583 (6.066); n=1,513 | 26.627 (4.881); n=1,215 | 32.062 (8.557); n=298 |
| Waist, cm | 94.273 (15.869); n=1,504 | 91.818 (13.195); n=1,213 | 106.042 (21.375); n=291 |
| SAPA, frozen units | 0.060 (1.631); n=1,513 | -0.115 (1.518); n=1,215 | 0.879 (1.876); n=298 |
| Total fat, kg | 27.581 (11.812); n=1,513 | 25.607 (9.462); n=1,215 | 36.822 (16.431); n=298 |
| Trunk fat, kg | 13.702 (6.633); n=1,513 | 12.577 (5.310); n=1,215 | 18.966 (9.184); n=298 |
| Whole-leg fat, kg | 9.531 (4.274); n=1,513 | 8.928 (3.638); n=1,215 | 12.355 (5.670); n=298 |
| Trunk-to-leg fat ratio | 1.478 (0.468); n=1,513 | 1.459 (0.462); n=1,215 | 1.566 (0.487); n=298 |
| Non-bone lean mass, kg | 50.401 (12.484); n=1,513 | 49.290 (11.153); n=1,215 | 55.600 (16.444); n=298 |

### Replication source files

Source files: Expanded descriptive summary; Descriptive summaries by imputation; Body composition estimates; Joint age comparison; Paired body measurement comparisons; Complete pooled replication estimates

### Extended Data figures and legends

### Extended Data Figure 1 | Local relationships and repeated-measurement structure

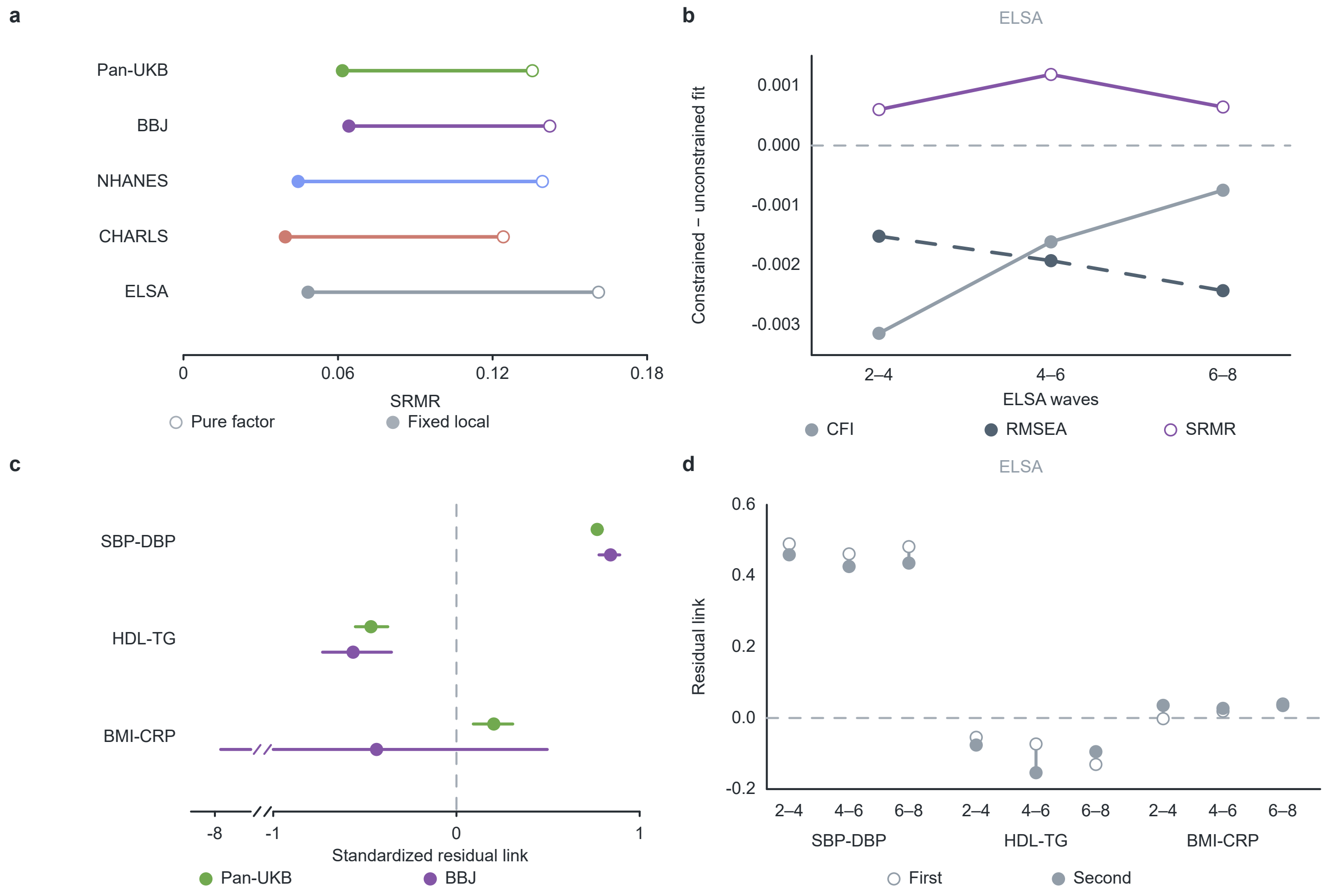


**a**, Standardised root mean square residual (SRMR) for the SAPA factor model without local relationships (open circles) and the model with three fixed local relationships (filled circles). These are systolic–diastolic blood pressure (SBP–DBP), high-density lipoprotein cholesterol–triglycerides (HDL–TG) and body mass index–C-reactive protein (BMI–CRP). Clinical samples match the NHANES reference and external cohort-native comparisons at ages 40–54 in Fig. 1b: NHANES, n = 1,104; CHARLS, n = 3,127; and ELSA, n = 413. Pan-UK Biobank (Pan-UKB) and Biobank Japan (BBJ) retain their original genetic populations and estimation scales. Extended Data Figure 2d gives the complementary comparative fit index (CFI).

**b**, Changes in robust CFI, robust root mean square error of approximation (RMSEA) and SRMR after constraining corresponding loadings across ELSA waves. Each value is constrained minus unconstrained fit for the named wave pair. **c**, The three standardised local relationships in Pan-UKB and BBJ, with their 95% parametric-bootstrap intervals. A marked axis break omits −7.7 to −1; intervals are not capped at the break. The interval uses all 935 finite BBJ bootstrap results, including 353 Heywood solutions; Supplementary Note 1 reports fit-status counts and the separate regular-only direction diagnostic. **d**, The same three local relationships at the first (open circles) and second (filled circles) assessments of each ELSA wave pair. Values in b and d are point estimates without interval summaries. These structural analyses use the stated marker transformations and sample definitions; repeated waves belong to one cohort. Clinical SAPA scoring coefficients come from the fixed NHANES reference. Source data.

Extended Data Figure 2 | Marker relationships, scoring and genetic structure

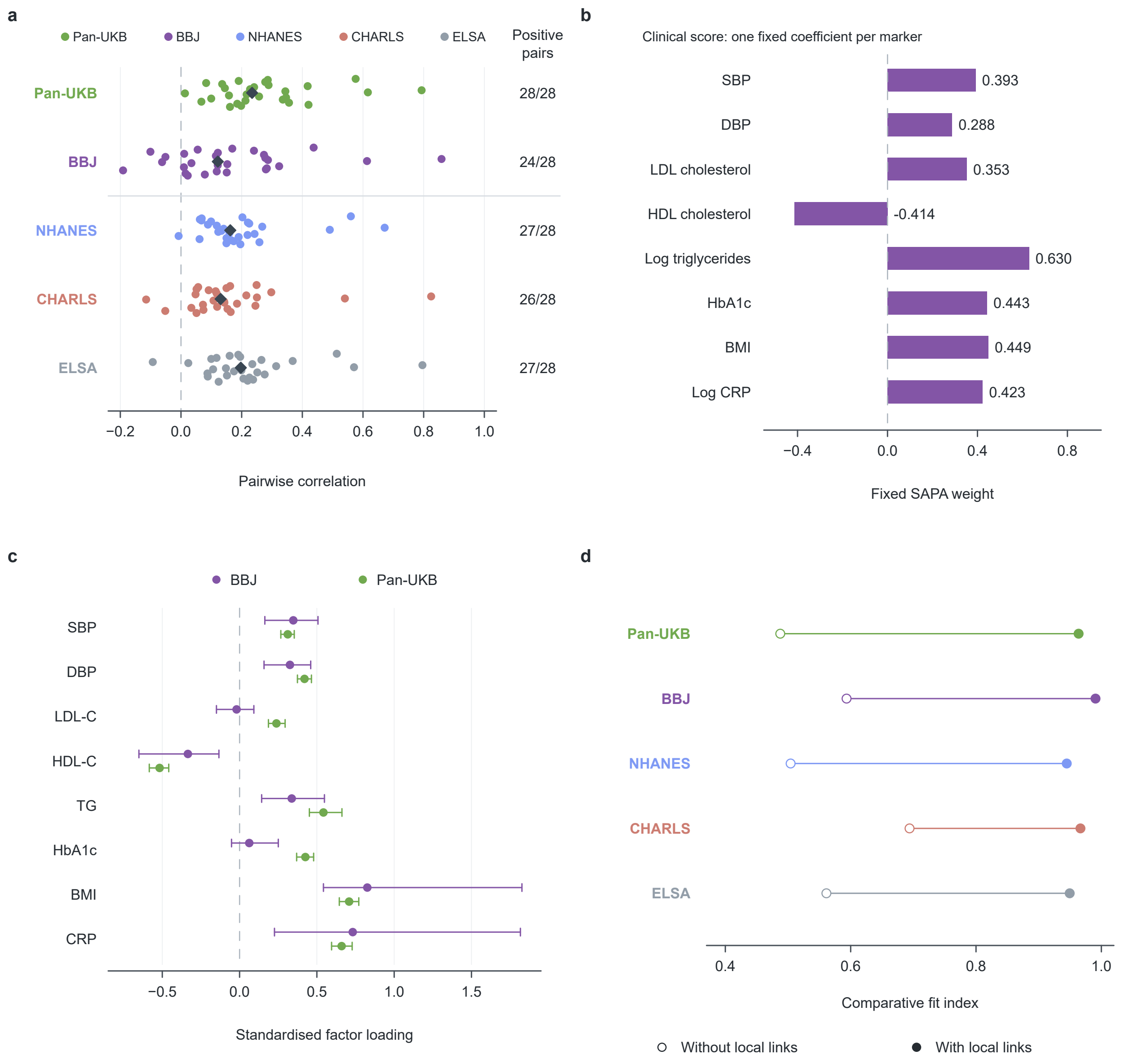


**a**, All 28 distinct pairs among eight markers in each source. Pan-UK Biobank (Pan-UKB) and Biobank Japan (BBJ) use their genetic correlation matrices; the National Health and Nutrition Examination Survey (NHANES), China Health and Retirement Longitudinal Study (CHARLS) and English Longitudinal Study of Ageing (ELSA) use the same clinical age range of 40–54 years (n = 1,104, 3,127 and 413). NHANES uses the repeated-reading blood pressure rule in Supplementary Note 1; external source matrices retain their cohort-specific processing. High-density lipoprotein (HDL) cholesterol is reversed for this panel so that higher values indicate the adverse direction. Each dot is one marker pair; vertical offsets separate points and diamonds show medians. The right column counts positive point estimates, with pair identities supplied in Source Data. **b**, Clinical SAPA scoring coefficients, estimated in 4,565 NHANES participants and displayed to three decimals. All coefficient bars use the SAPA colour. Cohort colours in a,c,d identify the labelled sources. **c**, The 16 standardised genetic loadings used in Fig. 1b and their parametric-bootstrap 95% intervals. All limits are displayed, including the BBJ BMI and CRP upper limits above one. Colours identify genetic sources; paired rows compare the same marker. Corresponding clinical intervals were unavailable for these age-specific estimates. **d**, Comparative fit index (CFI) without (open circles) and with (filled circles) the SBP–DBP, HDL –TG and BMI–CRP local relationships. These local relationships are residual correlations beyond the shared factor. Lines connect the two model fits within each source. Clinical robust CFI uses the age-matched samples in a; genetic estimates retain their source populations. Panels c,d describe model structure; b defines clinical scoring. SBP, systolic blood pressure; DBP, diastolic blood pressure; LDL-C, low-density lipoprotein cholesterol; HDL-C, HDL cholesterol; TG, triglycerides; HbA1c, glycated haemoglobin; BMI, body mass index; CRP, C-reactive protein. TG and CRP use the retained log transformations in the clinical coordinate. Source data.

Extended Data Figure 3 | Recovery of a missing marker and the complete SAPA score

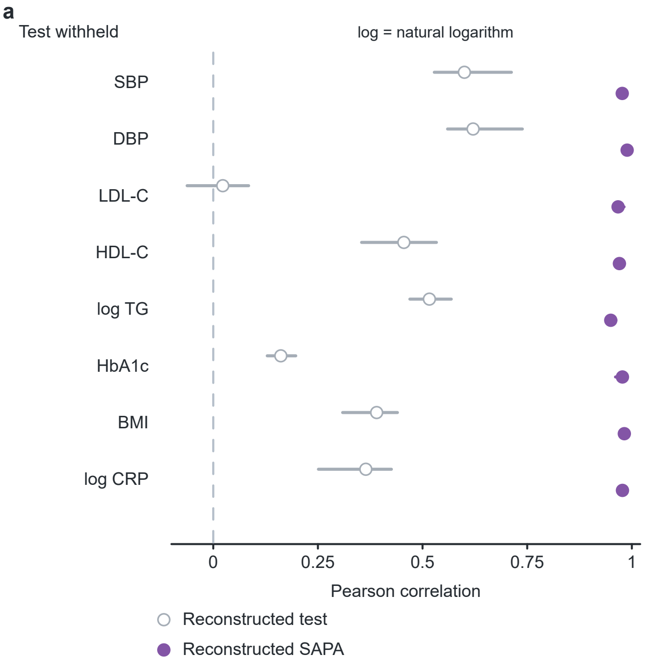


One of the eight tests was withheld in turn and estimated from the other seven. “Reconstructed test” (grey open circles) shows the Pearson correlation between estimated and measured values of the withheld test. “Reconstructed SAPA” (purple circles) shows the correlation between the score using that estimate and the score using all eight measured values. TG and CRP use their natural-log scales and the other markers their reference-standardised scales. Points are means across six measurement panels (ELSA waves 2, 4, 6 and 8 and CHARLS waves 1 and 3). Bars give the range across panels. Among the 16 estimates, the range for LDL cholesterol extends below zero.

Estimates use the covariance from the fixed NHANES confirmatory factor model, including its loadings, residual variances and three local residual relationships. Supplementary Note 1 gives the reconstruction and scoring equations, eligibility criteria and input transformations. Fig. 1e shows the separate analysis in which the missing term was dropped from the score. Source data.

Extended Data Figure 4 | Paired anatomical contributions after waist and total fat

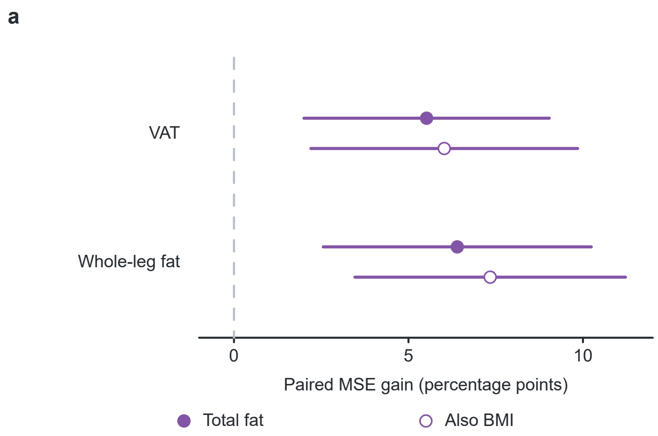


Direct paired improvements when SAPA is added to waist in 1,465 NHANES 2015–2018 participants, using the same eligible population as the continuous MetS comparison in Fig. 3e. Filled circles retain the total-fat-conditioned background; open circles additionally include BMI. Background terms include age, sex, race/ethnicity and education under the original specification. Targets are visceral adipose tissue (VAT) and whole-leg fat, summing both legs.

Points show percentage-point differences in mean squared error (MSE) improvement relative to each specification's background MSE. Positive values favour adding SAPA. Bars are pointwise 95% t intervals with 30 degrees of freedom from 60 survey jackknife replicates, each repeating both reciprocal-cycle training directions. Their denominators depend on the conditioning specification. The imaging-free task in Fig. 3h uses a separate population and predictor set. Supplementary Table S6 and Supplementary Note 5 retain the complete construct comparisons. Source data.

Extended Data Figure 5 | Complete genetic diagnostic-estimator comparison

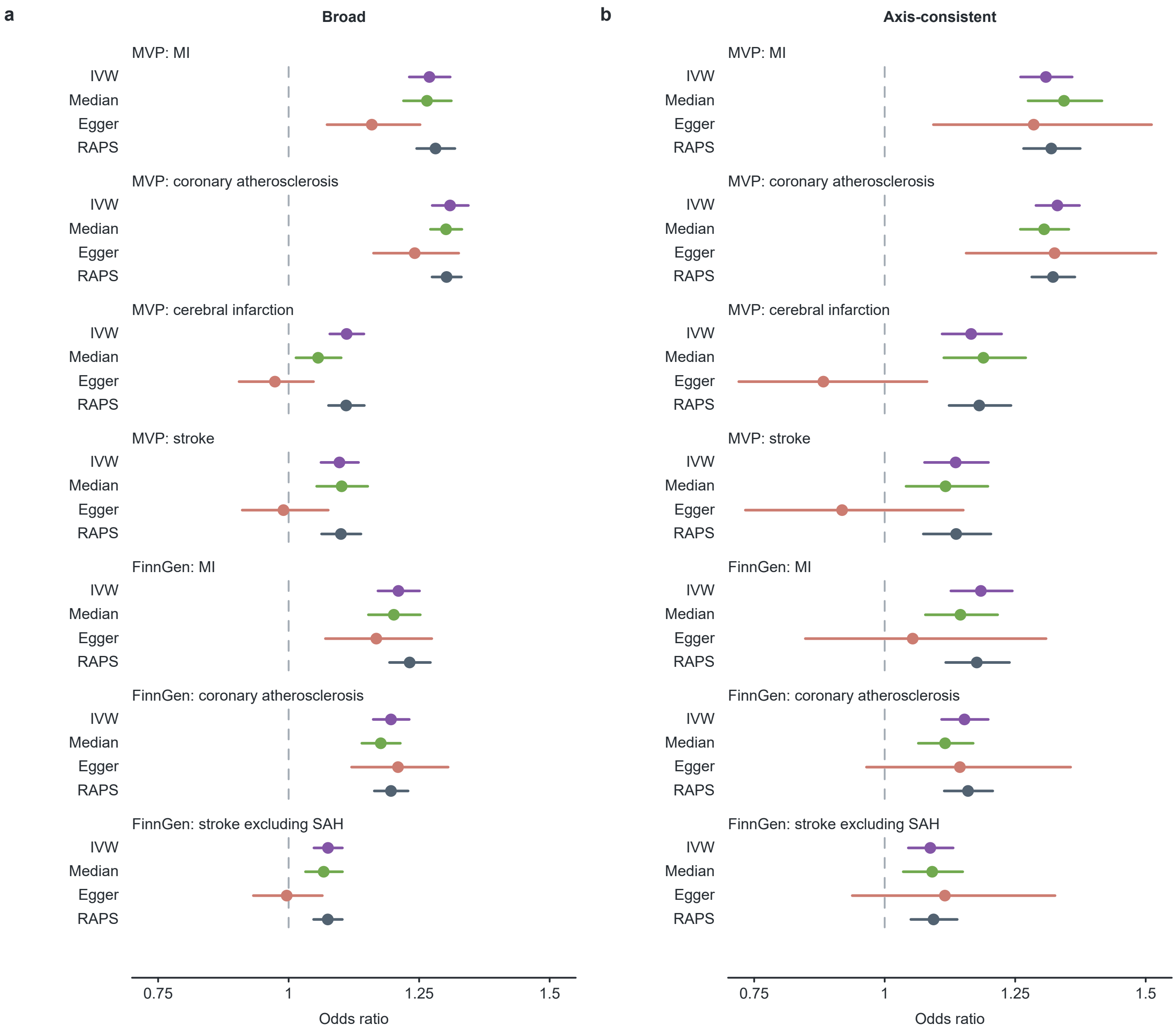


**a,b**, Broad and axis-consistent instrument rules, respectively. Each includes all seven retained vascular endpoints and four estimators: inverse-variance weighted multiplicative random effects (IVW; purple), weighted median (green), MR-Egger (coral) and robust adjusted profile score (RAPS; slate). Points and bars are odds ratios and 95% confidence intervals on the original genetic-factor scale. Dashed lines mark an odds ratio of one. The 56 estimates use the complete gSAPA input and the allele-quality rules in Supplementary Note 4.

Outcome identifiers are MVP MI (GCST90475932), coronary atherosclerosis (GCST90475936), cerebral infarction (GCST90477991), stroke (GCST90476623), and FinnGen R12 MI (I9_MI_STRICT), coronary atherosclerosis (I9_CORATHER) and stroke excluding subarachnoid haemorrhage (SAH; I9_STR). MVP stroke is the survey disease-diagnosis endpoint CircStrk in the GIA European group (18,436 cases; 297,232 controls), with subtype unspecified and no documented SAH exclusion rule. After harmonisation, Broad/axis-consistent instrument counts are 263/126 in MVP and 249/119 in FinnGen. These rules were clumped separately and their instrument sets are not nested. The four estimators use the same harmonised data within each instrument set and make different assumptions about instrument validity. Supplementary Table S9 gives all estimates, heterogeneity statistics and the seven-endpoint Holm adjustment within each estimator and rule. Source data.

Extended Data Figure 6 | Full marker associations with body composition in two survey periods

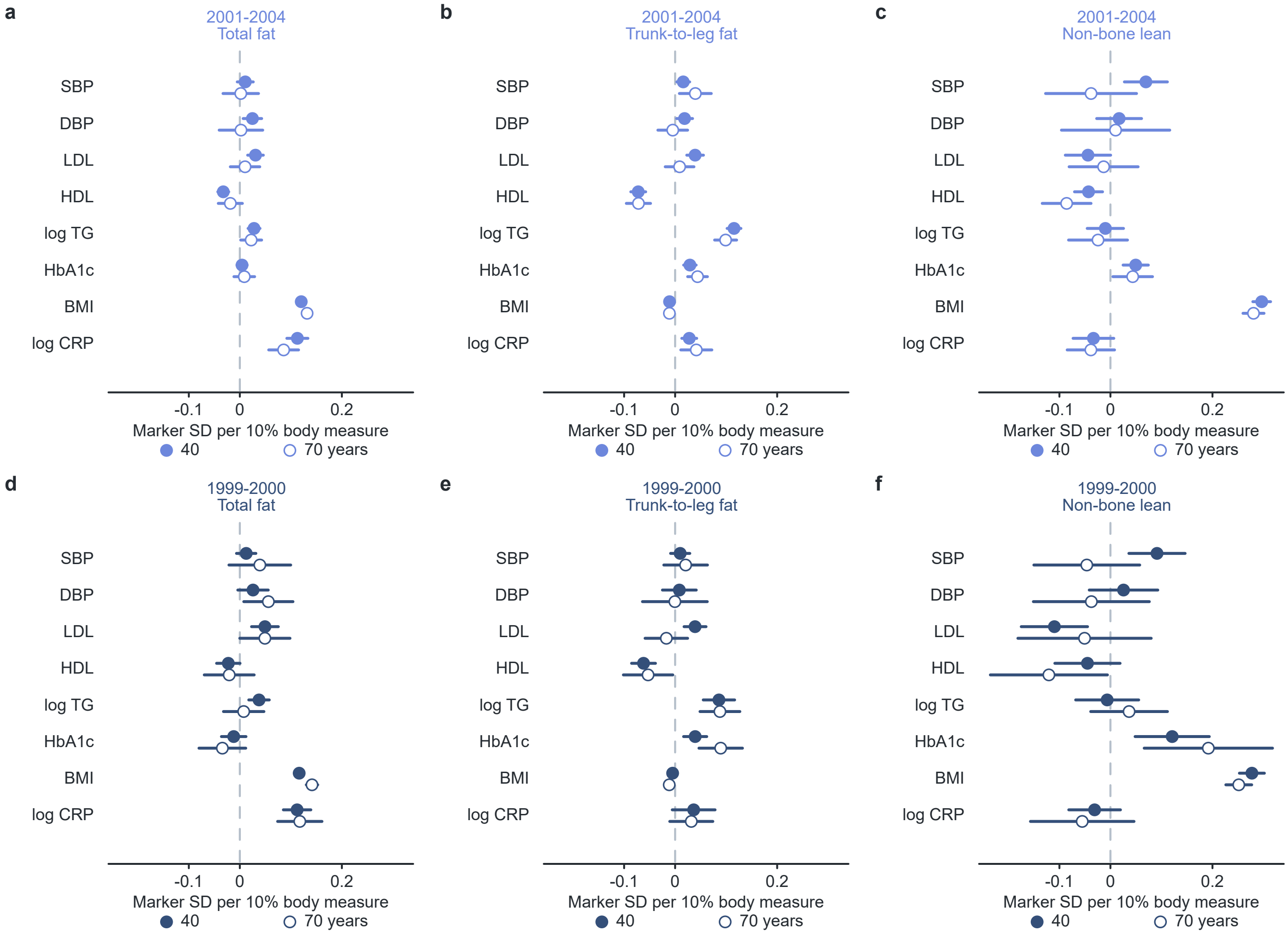


**a–c**, NHANES 2001–2004 (n = 3,242, ages 20–79). **d–f**, NHANES 1999–2000 (n = 1,513). Columns show total fat, trunk-to-leg fat ratio and non-bone lean mass. Within each panel, filled and open circles give the modelled associations at ages 40 and 70, respectively, for all eight constituent markers. Points show differences in fixed-reference marker SD per 10% higher body measure, holding the other two body measures constant. All six panels use a common horizontal scale. Colours identify the survey period.

Sex-specific models include age, height, the three log-body measures and linear age interactions; cycle terms apply in 2001–2004. The two sexes are averaged equally. Bars are pointwise 95% intervals from five official DXA imputations and 399 survey-design resamples per imputation, using Rubin pooling and Barnard–Rubin degrees of freedom. The periods are estimated separately. Triglycerides and CRP are log-transformed; HDL is shown on its original measurement direction. The 96 marker estimates complement SAPA and BMI-omitted associations in Fig. 6e. Supplementary Tables S14–S16 contain the direct age contrasts and complete results. Source data.

### Extended Data Figure 7 | Body-related clustering differences in primary and measured subsets

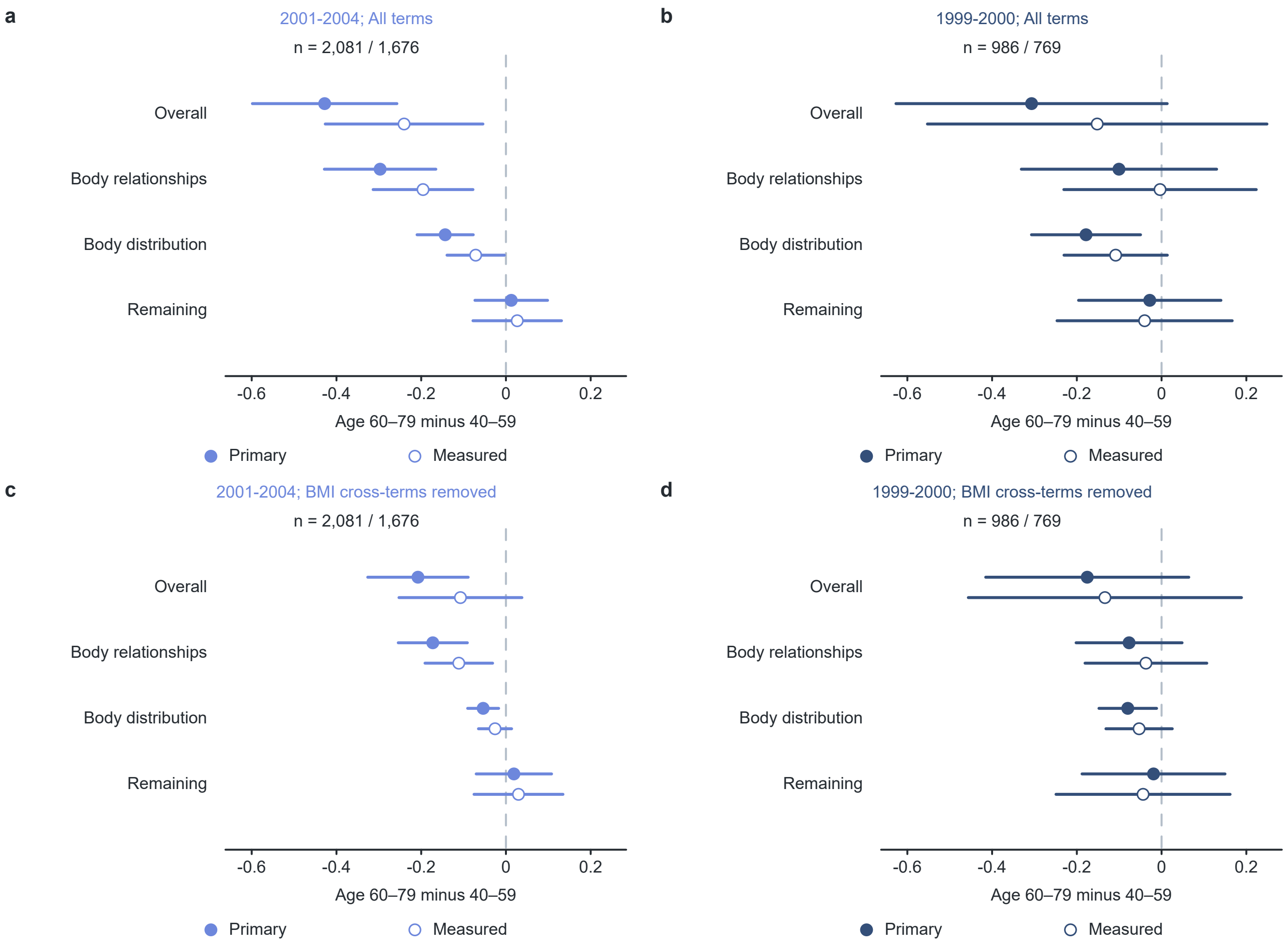


**a,b**, Age differences in cross-marker variation B in NHANES 2001–2004 and 1999–2000. **c,d**, Corresponding results after removing BMI cross-terms while retaining all other fixed weights. Filled circles represent the primary samples (n = 2,081 and 986); open circles represent the verified measured-body-composition subsets (n = 1,676 and 769). Every contrast compares ages 60–79 with 40–59.

The overall difference is partitioned into body-to-marker relationships, body-distribution differences and remaining covariance, using the definitions in Supplementary Note 8. Bars are pointwise 95% intervals with five official DXA imputations, 399 survey-design resamples per imputation, Rubin pooling and Barnard–Rubin degrees of freedom. Restricting to measured values also changes the population represented; Supplementary Tables S22–S23 describe those populations. Fig. 6f shows the corresponding primary decomposition. Source data.

Extended Data Figure 8 | DXA and routine body measurements in matched populations

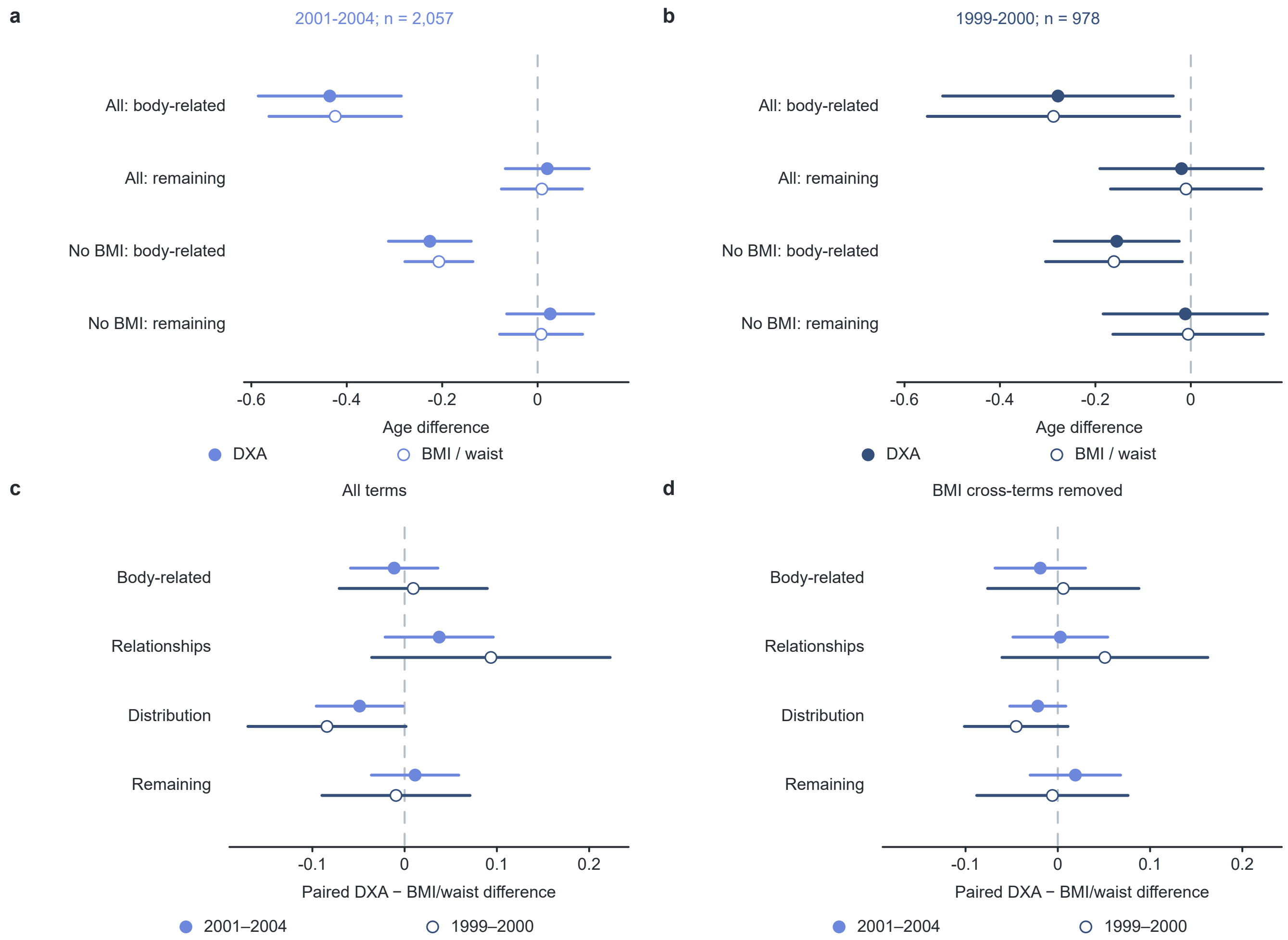


**a,b**, Body-related and remaining components of the older-minus-middle-aged B contrast using DXA (filled circles) or BMI/waist (open circles) in exactly the same people: n = 2,057 in 2001–2004 and n = 978 in 1999–2000. Each includes the full eight-marker definition and the version with BMI cross-terms removed. **c,d**, Direct paired DXA-minus-BMI/waist differences for the body-related, relationship, distribution and remaining components, with all terms and with BMI cross-terms removed, respectively. Blue filled circles represent 2001–2004 and dark-blue open circles represent 1999–2000.

All age contrasts compare ages 60–79 with 40–59. Within age, sex and cycle cells, identical adjustment for age and height precedes weighted unpenalised projection; matrices are averaged equally across sex and cycle cells. Bars are pointwise 95% intervals with five imputations and 399 survey-design resamples, Rubin pooling and Barnard–Rubin degrees of freedom. Direct paired contrasts preserve covariance between the two representations. Supplementary Tables S16, S18 provide their numerical values. Fig. 6i and Supplementary Table S19 separately address frozen cross-period transfer. Source data.

Extended Data Figure 9 | Cross-cohort and within-person age patterns in marker structure

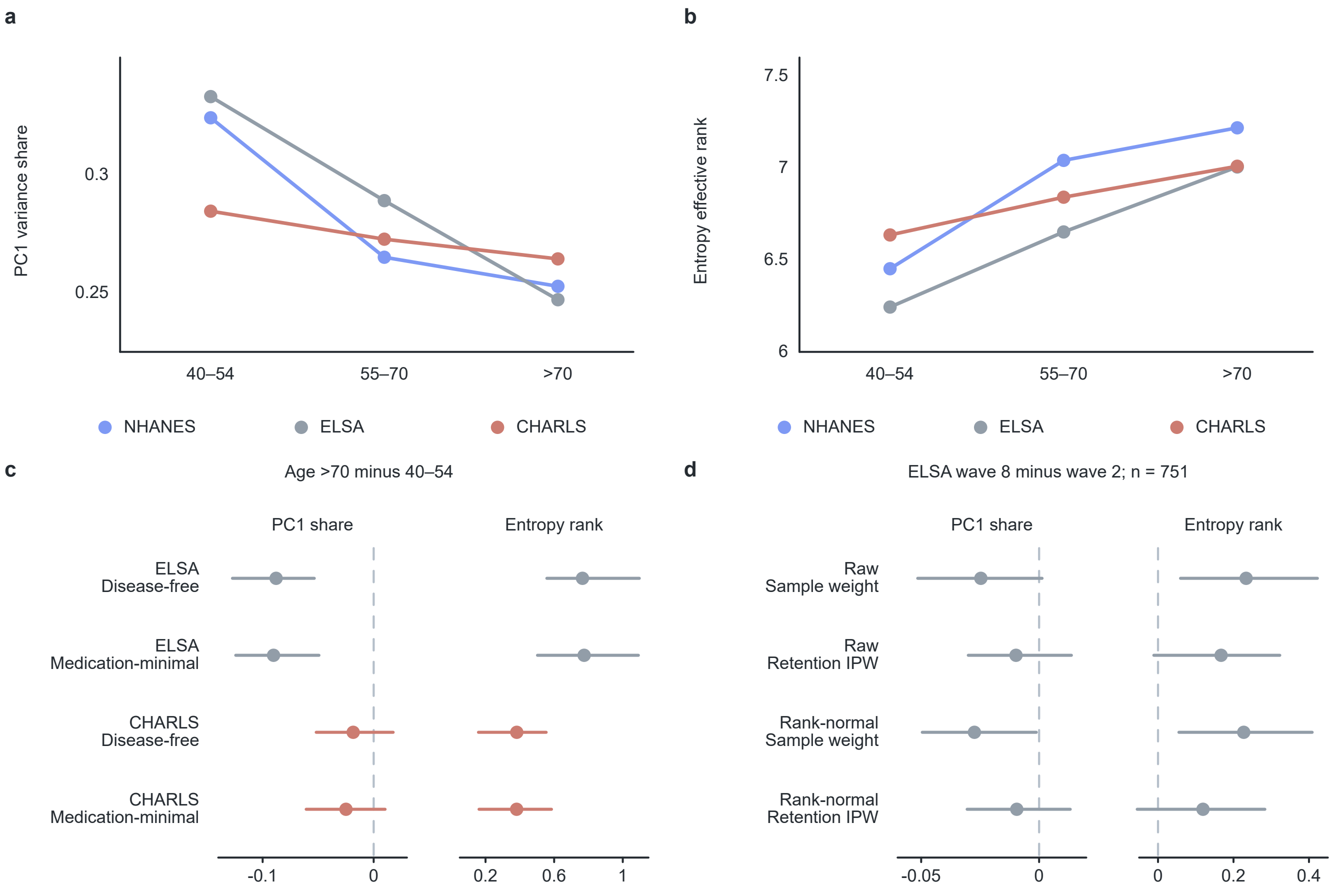


**a,b**, First principal component (PC1) variance share and entropy effective rank of the weighted eight-marker correlation matrix in NHANES, ELSA and CHARLS at ages 40–54, 55–70 and >70 years. Lines connect cross-sectional age strata; source-specific sample counts are supplied in Source Data. **c**, Older-minus-younger differences (>70 minus 40–54 years) in disease-free and medication-minimal ELSA and CHARLS populations. **d**, ELSA wave 8 minus wave 2 differences in the same 751 participants, comparing raw and rank-normalised measurements and sample weights with retention inverse-probability weights (IPW). Bars in c and d are 2.5th–97.5th percentile limits from 500 resamples. Panels a and b retain point estimates without interval summaries.

The correlation-spectrum analysis standardises markers within each analysis population. NHANES blood pressure follows the repeated-reading rule in Supplementary Note 1, with n = 1,017, 1,286 and 644 in ascending age strata. PC1 variance share describes concentration in the leading direction of that correlation matrix; entropy effective rank describes how its eigenvalue mass is distributed. Fig. 6 instead evaluates the frozen SAPA direction on the NHANES-reference scale. The paired comparison in d follows a selected surviving and remeasured population; the source-data table retains all weighting and transformation variants. Source data.

### Supplementary figures and legends

#### Supplementary Figure 1 | Additional clinical associations and overall prediction displays

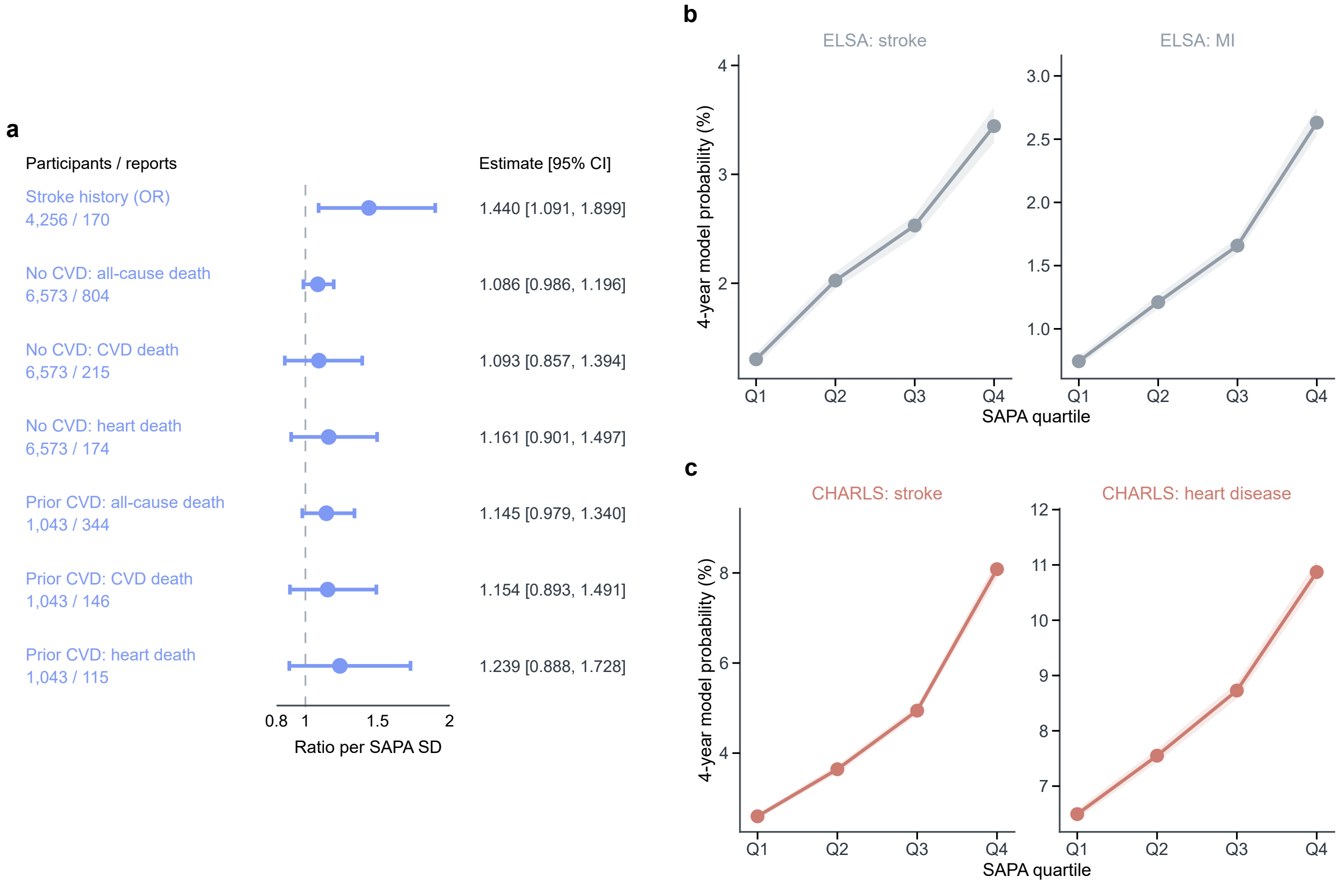


**a**, NHANES stroke-history odds ratio (OR) and mortality hazard ratios (HRs), per analysis-specific SAPA SD. Points and bars show estimates and 95% confidence intervals. Stroke history includes 4,256 participants/170 reports. Mortality analyses include 6,573 people without prior CVD and 1,043 with prior CVD; outcome-specific death counts are printed. The distinct OR and HR estimates are not pooled. Supplementary Table S4 gives model and population definitions.

**b,c**, Mean four-year model probabilities by cohort- and endpoint-specific SAPA quartile for ELSA (b) and CHARLS (c). ELSA stroke/MI include 3,956/3,941 participants; CHARLS stroke/broad heart disease include 6,628/5,455. Lines connect quartile means; bands are conditional 95% percentile intervals from 500 household/community resamples with fitted predictions and cutoffs fixed. CHARLS probabilities account for death before onset. These full-cohort interval-observation models differ from the lower-category Cox models in Fig. 2. Source data.

### Supplementary Figure 2 | Waist change and SAPA change

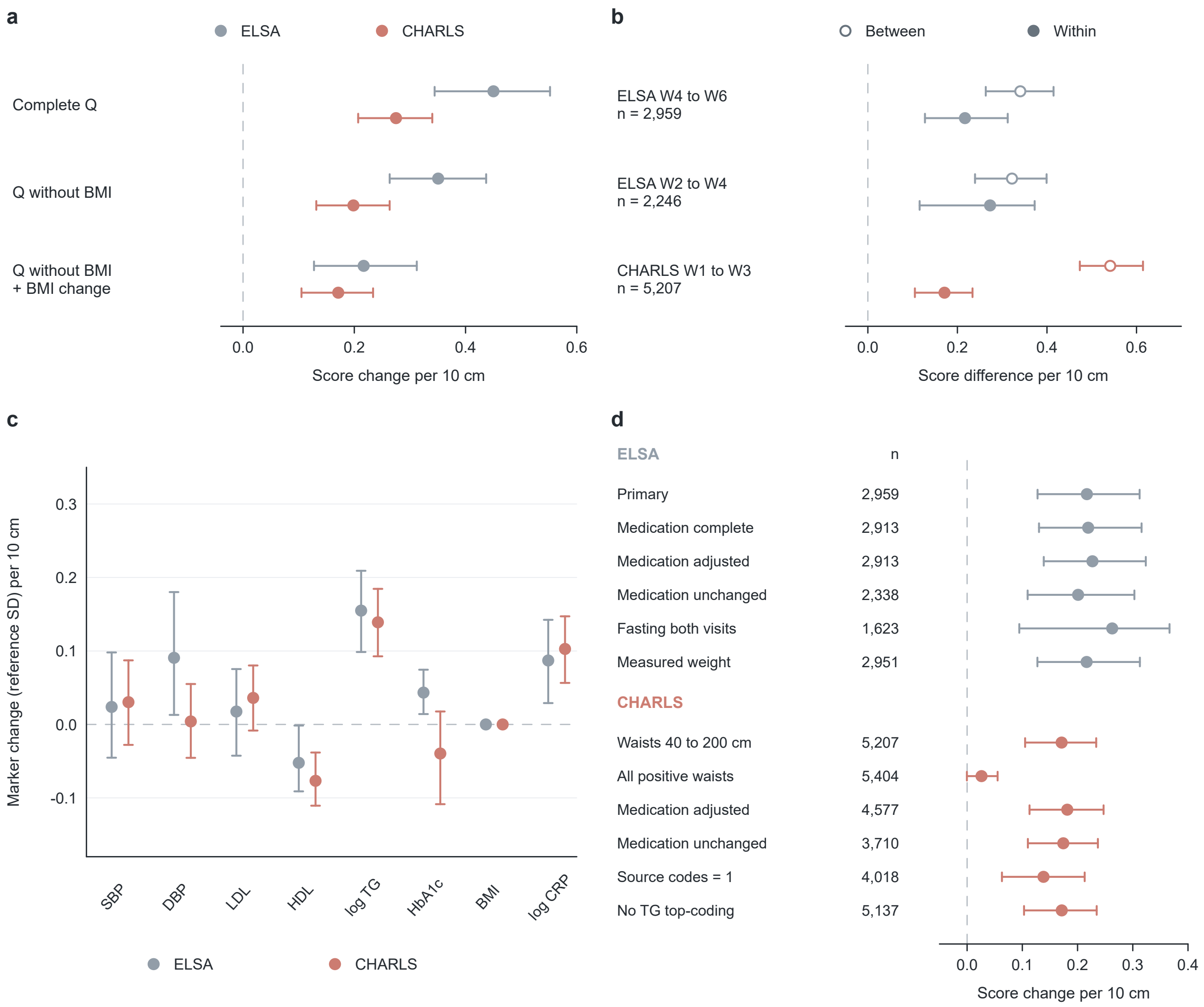


**a**, Associations per 10-cm waist increase for the clinical SAPA score Q, Q omitting BMI, and BMI-omitted Q conditioned on BMI change. The English Longitudinal Study of Ageing (ELSA), waves 4 to 6, included 2,959 participants; the China Health and Retirement Longitudinal Study (CHARLS), waves 1 to 3, included 5,207 with both waists within the prespecified 40–200 cm review range. **b**, Between-person (open circles) and within-person (filled circles) associations for BMI-omitted Q, conditioned on mean BMI or BMI change, respectively. The earlier ELSA window included 2,246 people and overlapped the primary window. **c**, BMI-conditioned within-person associations for all eight markers, in fixed NHANES-reference standard deviation (SD) per 10 cm. HDL retains its measured direction. BMI has a zero coefficient because BMI change is included as a predictor. **d**, BMI-conditioned associations for BMI-omitted Q in prespecified subsets; labels give participant counts. The CHARLS all-positive-waist comparison (5,404 people) includes 197 with at least one waist below 40 cm. The review range was fixed before fitting and is not an official quality standard. Supplementary Table S7 gives population definitions, medication-complete comparators and direct contrasts.

ELSA is grey-blue and CHARLS coral throughout. Points and bars show estimates and pointwise 95% percentile intervals from 999 household bootstrap resamples. All regressions include baseline age, sex, smoking and visit interval and retain follow-up blood-sampling weights. Related models share resamples. Q uses the frozen six-decimal weights without further score-SD scaling. ELSA uses its recorded fasting-eligibility indicators. The CHARLS source-code subset requires qc1_va003=1 and bl_fasting=1; the wave-1 code meaning remains unverified. Medication-adjusted models include recorded medication means or changes and leave marker inputs observed. ELSA timing follows nurse visits; individual blood-to-examination timing was unavailable in CHARLS. BP, blood pressure; BMI, body mass index; SBP, systolic BP; DBP, diastolic BP; LDL, low-density lipoprotein cholesterol; HDL, high-density lipoprotein cholesterol; TG, triglycerides; HbA1c, glycated haemoglobin; CRP, C-reactive protein. Source data.

### Supplementary Figure 3 | Transfer of the IL-1RA relationship from younger to older adults

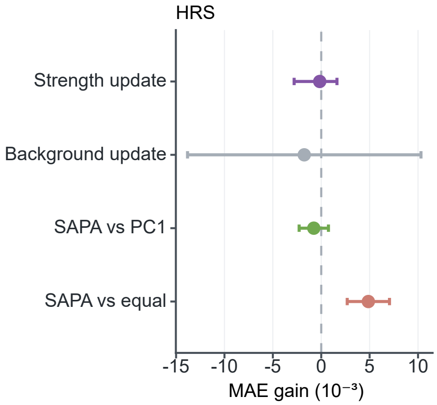


Paired mean absolute error (MAE) improvements in the age-transfer task: training at 55–69 (n = 1,757), evaluation at 70–84 (n = 1,287), with separate calibration/testing families and 200 family resamples with full retraining. The score-strength update uses the primary 95% basic interval; background update and weighting comparisons use three-comparison family intervals. Positive values favour the named update or SAPA. Units are $10^{-3}$ log-concentration. SAPA is purple, PC1 green, equal weights coral and background grey.

This analysis used the Health and Retirement Study (HRS). IL-1RA denotes interleukin-1 receptor antagonist; SAPA, shared adverse physiological axis; PC1, first principal component. Supplementary Table S8 and Supplementary Note 6 describe the corresponding models and population separation. Source data.
